# Genetic influences on neurodevelopmental disorders and their overlap with co-occurring conditions in childhood and adolescence: A meta-analysis

**DOI:** 10.1101/2022.02.17.22271089

**Authors:** Agnieszka Gidziela, Yasmin I. Ahmadzadeh, Giorgia Michelini, Andrea G. Allegrini, Jessica Agnew-Blais, Lok Yan Lau, Megan Duret, Francesca Procopio, Emily Daly, Angelica Ronald, Kaili Rimfeld, Margherita Malanchini

## Abstract

A systematic understanding of the aetiology of neurodevelopmental disorders (NDDs), their co-occurrence, and co-occurrence with other conditions during childhood and adolescence remains incomplete. This meta-analysis bridges gaps in our knowledge. First, we meta-analysed the literature on the relative contribution of genetic and environmental factors to NDDs. Second, we considered the literature on the overlap between different NDD categories. Lastly, we synthesized the literature on the co-occurrence between NDDs and disruptive, impulse control and conduct disorders (DICCs). We performed multilevel, random-effects meta-analyses on 296 independent studies, including over 4 million children and adolescents. We found all NDDs to be substantially heritable (family-based heritability (h^2^) = 0.66; SNP h^2^ = 0.19). Meta-analytic genetic correlations between NDDs, and between NDDs and DICCs were moderate to strong. However, given the paucity of available studies covering the co-occurrence of NDDs and DICCs, these could only be estimated for a few disorders. While our work provides direct evidence to inform and potentially guide clinical and educational diagnostic procedures and practice, it also highlights the imbalance in the research effort that has characterized developmental genetics research.

## Introduction

Neurodevelopmental disorders (NDDs) are a serious and complex health concern, starting from childhood^1^. NDDs affect around 15% of children and adolescents worldwide and lead to impaired cognition, communication, adaptive behavior, and psychomotor skills^2^. The fifth edition of the Diagnostic and Statistical Manual of Mental Disorders (DSM-5) categorizes the following seven disorders under NDDs: *intellectual disabilities, communication disorders, autism spectrum disorder (ASD), attention-deficit/hyperactivity disorder (ADHD), specific learning disorders, motor disorders and other neurodevelopmental disorders*^3^.NDDs often have lifelong trajectories: they can manifest as early as before the child reaches 12 months of age^4^ and can be identified and diagnosed before children enter primary education^5,3^. While some NDDs (e.g. ASD and ADHD) may persist throughout adolescence and adulthood^6,7^, others are more likely to alleviate as children get older (e.g., tic disorder^8^ and communication disorders^9^); nevertheless, all NDDs can lead to social and behavioural difficulties and reduced, or even a lack of, independence over the lifespan^6,7^.

A systematic understanding of the aetiology of NDDs remains incomplete. A disproportionate number of studies, reviews, and syntheses of extant literature have focused on ASD and ADHD. However, other neurodevelopmental conditions, despite showing similar prevalence rates and severity as ASD and ADHD, are less well understood and studied^10^. The focus on ASD and ADHD has resulted in several studies that have systematically synthesized the literature on the aetiology of these two NDDs, pointing to their substantial heritability– the extent to which observed individual differences are accounted for by underlying genetic differences.

A meta-analysis of 7 twin studies of clinically diagnosed ASD in childhood and adolescent samples (aged 2 to 23 years) yielded a grand heritability estimate of 0.74 (95% CIs= 0.70, 0.87)^11^. Similarly sizeable heritability estimates also emerged from a meta-analysis of 26 studies of ADHD in childhood and adolescence, which yielded a pooled estimate for additive genetic effects –independent effects of genetic variants– of 0.26 (95% CIs= 0.20, 0.32) and a pooled estimate for dominant genetic effects – interactive effects between genetic variants at one more loci– of 0.44 (95% CIs= 0.38, 0.51)^12^. A subsequent systematic review of 37 twin studies of ADHD, including studies that had adopted either categorical or dimensional measures, yielded a mean h^2^ of 0.74^13^. Heritability estimates were found to differ across the two major components of ADHD, with genetic factors playing a more substantial role in the aetiology of hyperactivity (h^2^= 0.71, 95% CIs= 0.63, 0.75; based on 9 studies), if compared inattention (h^2^= 0.56, 95% CIs= 0.48, 0.63; based on 13 studies)^14^.

In line with what observed for all complex traits, heritability estimates for ASD and ADHD obtained from DNA data are lower than those obtained from twin and family designs^15^, likely due, at least in part, to the additive models used to calculate heritability from single nucleotide polymorphism (SNP)^16^. SNP heritability can be calculated using large samples of individual-level genotype data^17^ or summary statistics from genome-wide association studies (GWAS)^18^, hypothesis-free studies aimed at discovering associations between genetic variation across the genome and individual differences in traits and disorders. The two largest studies to date that have estimated the SNP heritability of ASD and ADHD have applied linkage disequilibrium score (LDSC) regression to GWAS summary statistics^18^ and report estimates of 0.12 (SE= 0.01) for ASD^19^ and 0.22 (SE= 0.01) for ADHD^20^.

It is now well-established that NDDs often co-occur with one another, a phenomenon known as homotypic co-occurrence, and this points to a shared underlying liability between conditions^21,22^. Even in this instance, most genetic investigations have focused on examining the genetic correlations (i.e., the degree to which the same genetic variants contribute to the observed covariation between pairs of traits or disorders^23^) between ASD and ADHD, which were found to be strongly phenotypically correlated (0.54)^24^. Knowledge on the co-occurrence between ASD and ADHD has been synthesized by a meta-analysis of 11 twin studies that yielded a grand genetic correlation estimate of 0.59 (95% CIs= 0.49, 0.69)^25^. Aetiological sources of co-occurrence between the several remaining NDD categories have not been meta-analysed, nor has been the proportion of genetic variants shared between NDDs (SNP-based genetic correlation). The SNP-based genetic overlap between ASD and ADHD, based on summary-level data obtained from the largest GWAS meta-analyses to date has been estimated at 0.35 (SE = 0.01)^26^ in adult samples.

Individual studies point to a moderate to strong shared liability between ASD/ADHD and other NDDs. ASD and specific learning disorders were found to co-occur in 34% of children and their genetic correlation has been estimated at 0.70 (95% CIs= 0.63, 0.73)^27^. ADHD and specific learning disorders were found to co-occur in 16% of cases^27^ and their genetic correlation to range between 0.25 (95% CIs= -1, 1)^27^ to 0.60 (95% CIs= 0.52, 0.68)^28^. Genetic overlap has also been investigated between ASD/ADHD and motor disorders, with greater rates of co-occurrence for ADHD (50%)^29^ if compared to ASD (34%)^30^. For ADHD the genetic correlations ranged between 0.42 (95% CIs= 0.36, 0.48)^31^ with developmental coordination disorder to 0.99 (95% CIs= 0.81, 1) with tic disorder^27^. Genetic correlations were found to be strong between ASD and developmental coordination disorder (0.71, 95% CIs= 0.51, 0.91) and between ASD with tic disorder (0.60, 95% CIs= 0.42, 0.78)^27^. ASD was found to co-occur to a lesser extent with communication disorders, as indexed by weak phenotypic (a mean of -0.15 across language measures)^32^ and weak to moderate genetic correlations (ranging between -0. 18, 95% CIs= -0.32-0.04^32^ to 0.33, 95% CIs= 0.31, 0.35)^33^.

Another category of disorders that onset and progress through childhood and adolescence are Disruptive, Impulse Control and Conduct Disorders (DICCs), which the DSM-5 describes as disorders that share the underlying features of impulsive behavior, aggressiveness, and pathological rule breaking^3^. The DSM-5 identifies eight main DICC categories: *Oppositional Defiant Disorder, Intermittent Explosive Disorder, Conduct Disorder, Antisocial Personality Disorder, Pyromania, Kleptomania, Other Specified DICC Disorder, and Unspecified DICC Disorders*^3^ (**Figure 1**). The developmental nature of DICCs makes them an ideal primary target for the investigation of how NDDs co-occur with other disorders (i.e., heterotypic co-occurrence) during childhood and adolescence. However, the distinction between NDDs and DICCs in the published literature is often unclear, particularly for disorders that include clinical features that overlap with both NDD and DICC categories, such as ADHD. This lack of a clear-cut distinction is exemplified by the different classifications of ADHD across the iterations of the DSM, which transitioned from being included under the Attention-deficit and Disruptive Behaviour Disorders category (DSM-III-R)^34^, to Disorders Usually Diagnosed in Infancy, Childhood, and Adolescence (DSM-IV)^35^, to its current classification under the Neurodevelopmental Disorders category (DSM-5)^3^.

**Figure 1.**
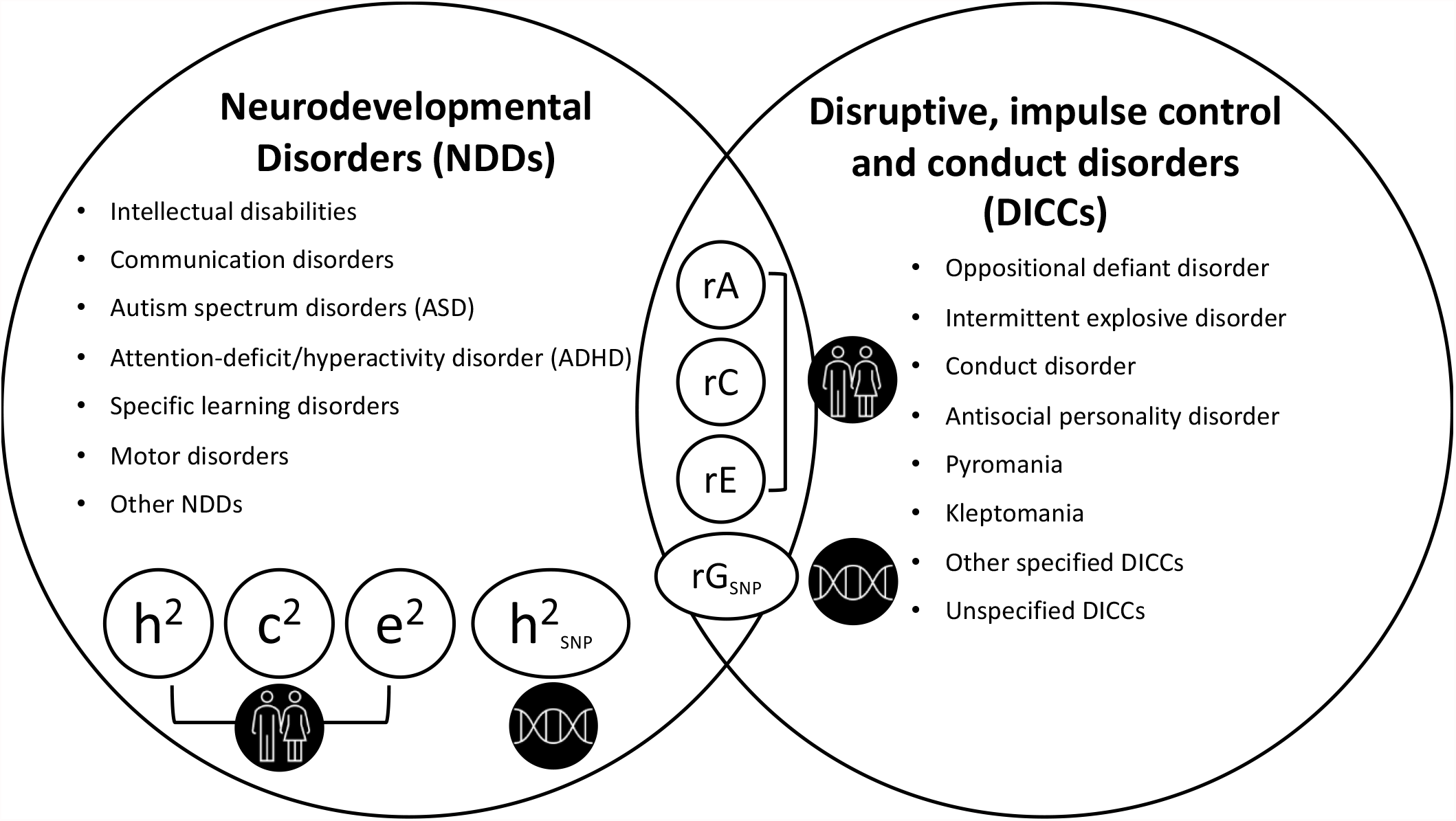
A visual summary of the three core aims of the current meta-analysis. Aim 1 (left): estimate family-based genetic (h^2^), shared environmental (c^2^) and nonshared environmental (e^2^) influences, as well as SNP heritability (h^2^) for all neurodevelopmental disorders (NDDs) identified by the DSM-5. Aim 2 (bottom left): Provide grand estimates of family-based genetic (rA), shared environmental (rC) and nonshared environmental (rE) correlations and SNP-based genetic correlations (rG_SNP_) between different NDDs. Aim 3 (right): Provide grand estimates of rA, rC, rE and rG_SNP_ between NDDs and disruptive, impulse control and conduct disorders (DICCs). Results for c^2^, e^2^, rC and rE are presented in **Supplementary Note 1**.

The most investigated example of symptom overlap between NDDs and DICCs involve ADHD and conduct disorder^36,37^, and ADHD and oppositional defiant disorder^38^. Studies reporting on the behavioural and cognitive profiles of ADHD and these disorders highlight how both disorders are characterised by disturbances in emotion regulation, attention problems, cognitive inflexibility, executive functioning, and impaired inhibition^37,39,40^. A shared symptomatology has also been observed between ASD and antisocial behaviour/personality disorder (that we refer to as conduct disorder in the current work due to antisocial personality disorder referring to adult diagnosis)^3,41,42^, with both disorders characterized by impairments in social reciprocity, expressing emotions and affective empathy^43,44^. Symptom resemblance that characterizes these disorder pairs is reflected in the phenotypic correlations between ADHD and conduct disorder (0.40)^38^ and between ADHD and oppositional defiant disorder (0.55)^38^, but not ASD and conduct disorder traits, for which the phenotypic correlation is modest (0.22 between social impairments and callous-unemotional traits and 0.21 between communication impairments and callous-unemotional traits)^45^.

Individual studies on the association between NDDs and DICCs are characterized by a great deal of heterogeneity and inconsistencies across co-occurring conditions. For example, one study found that, in a sample of 7-year-olds, genetic effects on ASD and callous-unemotional traits, one of the manifestations of conduct disorder in childhood and of antisocial personality disorder in adulthood, were largely independent, as indicated by small to moderate genetic correlations (0.23 (95% CIs= 0.16, 0.31) between the communication impairments domain of ASD and callous-unemotional traits, and 0.31 (95% CIs= 0.26, 0.36) between the social interaction impairments domain of ASD and callous-unemotional traits)^45^. Another study reported moderate genetic overlap of 0.43 (95% CIs= 0.34, 0.52) between ASD and psychopathic tendencies in 9-year-olds of 0.99 (95% CIs= 0.92, 1)^43^.

With three core aims (**Figure 1**), the current meta-analysis bridges major gaps in our knowledge of the aetiology of NDDs and of their co-occurrence with other developmental conditions. First, we meta-analysed studies on the relative contribution of genetic and environmental influences to all NDDs categories described in the DSM-5. Second, we meta-analysed estimates for the genetic and environmental comorbidities between different NDDs (homotypic co-occurrences). Third, given the historical associations between NDDs and DICCs, and their developmental onset and progression, we examined the aetiology of the co-occurrence between NDDs and DICCs (heterotypic co-occurrences).

In addition to addressing each disorder individually, we take a transdiagnostic approach by combining data across NDDs, their manifestations, and including categorical (i.e., presence or absence of a disorder) and quantitative (i.e., continuously measured symptoms) measures. Taking a transdiagnostic approach provides us with a holistic picture of the extent to which genetic and environmental factors contribute to NDDs, to their co-occurrence, and to their co-occurrence with other common developmental conditions. This will result in a clearer appreciation of the size, strength, pervasiveness, and developmental progression of the associations between different developmental disorders.

Clarifying the genetic and environmental aetiology of all NDDs and their homotypic and heterotypic co-occurrences will advance our knowledge of how developmental disorders cluster together, which could in turn inform educational and clinical practice^46^. Accurate and efficient evaluations and diagnoses, and related adjustments in the care and education of children with NDDs, will in turn have a positive impact on the children and their development, their families, and their educators^47^.

## Results

This *Results* section presents meta-analytic findings on genetic influences on NDDs and on their genetic overlap with other NDDs and DICCs. Meta-analytic estimates for shared and nonshared environmental factors and their overlap are presented in **Supplementary Note 1**. We report transdiagnostic grand estimates across all disorders and for broad NDD categories, comprising all studies that investigated the aetiology of a disorder either using diagnoses, categorical or quantitative measures. For example, the broad ADHD phenotype includes studies that have measured ADHD using diagnoses, clinical cut-offs, and continuous measures of ADHD traits, such as checklists and questionnaires. The only exception is intellectual disability (ID). We did not consider quantitative measures of general intelligence as indexing a continuum of ID given that ID, as described in the DSM-5 is a complex disorder, not only characterized by impairments in intellectual performance, but also in adaptive functioning and communication^3,44^. Finally, we considered specific manifestations of NDDs, for example, beyond ADHD, we also consider the hyperactive/impulsive and inattentive sub-types separately. Results for all sub-categories of NDDs and for their co-occurrence with other disorders are reported in **Supplementary Note 2, Supplementary Figures 2** and **3** and **Supplementary Tables 2, 4** and **6**.

### Searches and screening

Studies for this meta-analysis were selected during 3 screening stages including title and abstract screening, full text screening, and reference list screening (see ***Method*** for a detailed description). This selection process resulted in a total of 296 studies included in the current meta-analysis (**Figure 2**).

**Figure 2.**
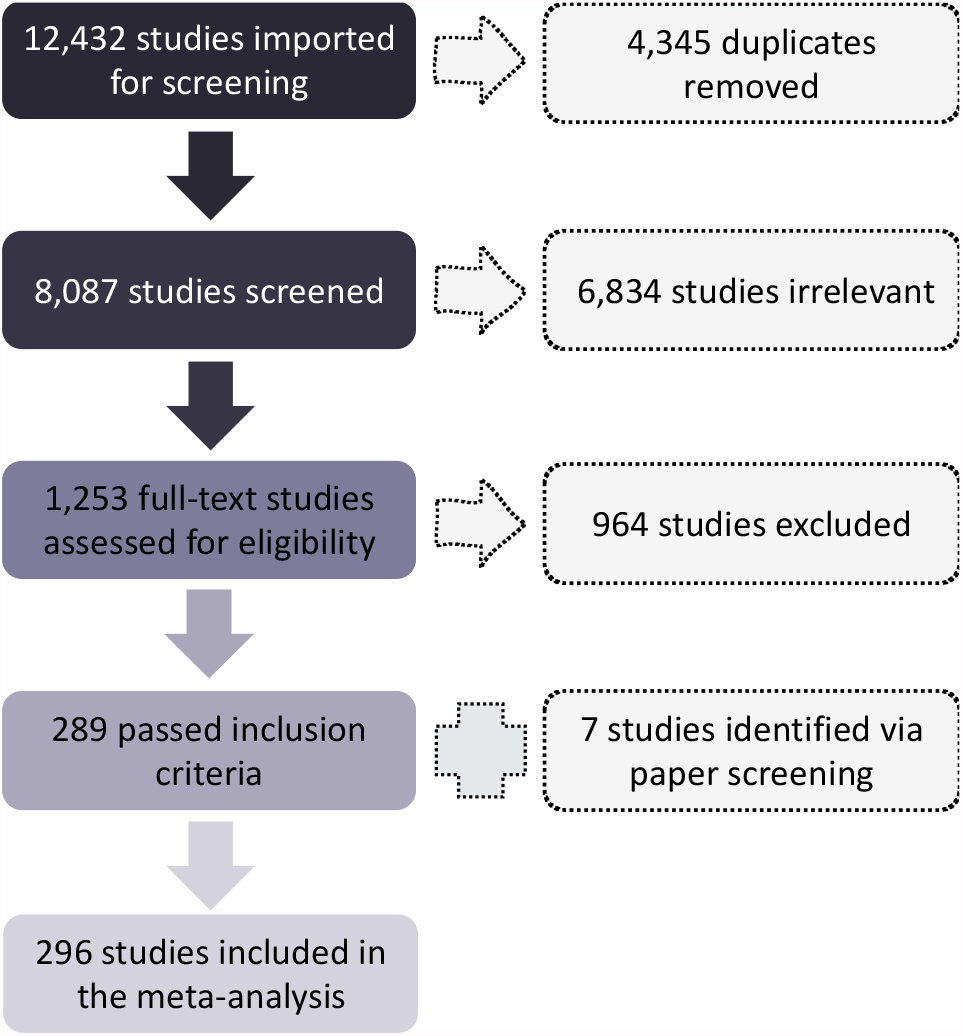
Diagram of searches and screening. This diagram ***p***resents an overview of the screening and selection process across primary and secondary searches.

### NDDs are substantially heritable

Our first aim was to synthesize the extant literature to obtain reliable estimates of the contribution of genetic factors to individual differences in all NDDs. We considered two broad categories of methods that allow for the estimation of *heritability*: family-based designs including related individuals (such as sibling comparisons and twin studies) and SNP heritability, the proportion of phenotypic variance explained by all measured single nucleotides polymorphisms (SNPs)^48^ (see ***Method***). Given the substantial differences in methodology and outcomes, findings across these two broad categories were meta-analyzed separately.

#### Family-based heritability (h^2^)

We identified a total of 237 family-based studies that investigated the proportion of variance in NDDs that is accounted for by genetic factors. Out of the total, 121 studies investigated ADHD, 89 studies specific learning disorders, 36 studies ASD, 23 studies communication disorders, 7 studies motor disorders and 2 studies intellectual disabilities. We applied multi-level random effect meta-analysis to obtain grand estimates for the heritability of NDDs. Across all NDDs and 237 studies, the grand *h*^*2*^ estimate was 0.66 (SE= 0.03). Grand *h*^*2*^ estimates differed, albeit not significantly, across NDD categories, ranging from 0.86 (SE= 0.44) for intellectual disabilities to 0.62 (SE= 0.04) for specific learning disorders (see **Figure 3** and **Supplementary Table 1**). Distributions of genetic influences across studies and NDDs are presented in **Supplementary Figure 1**.

**Figure 3.**
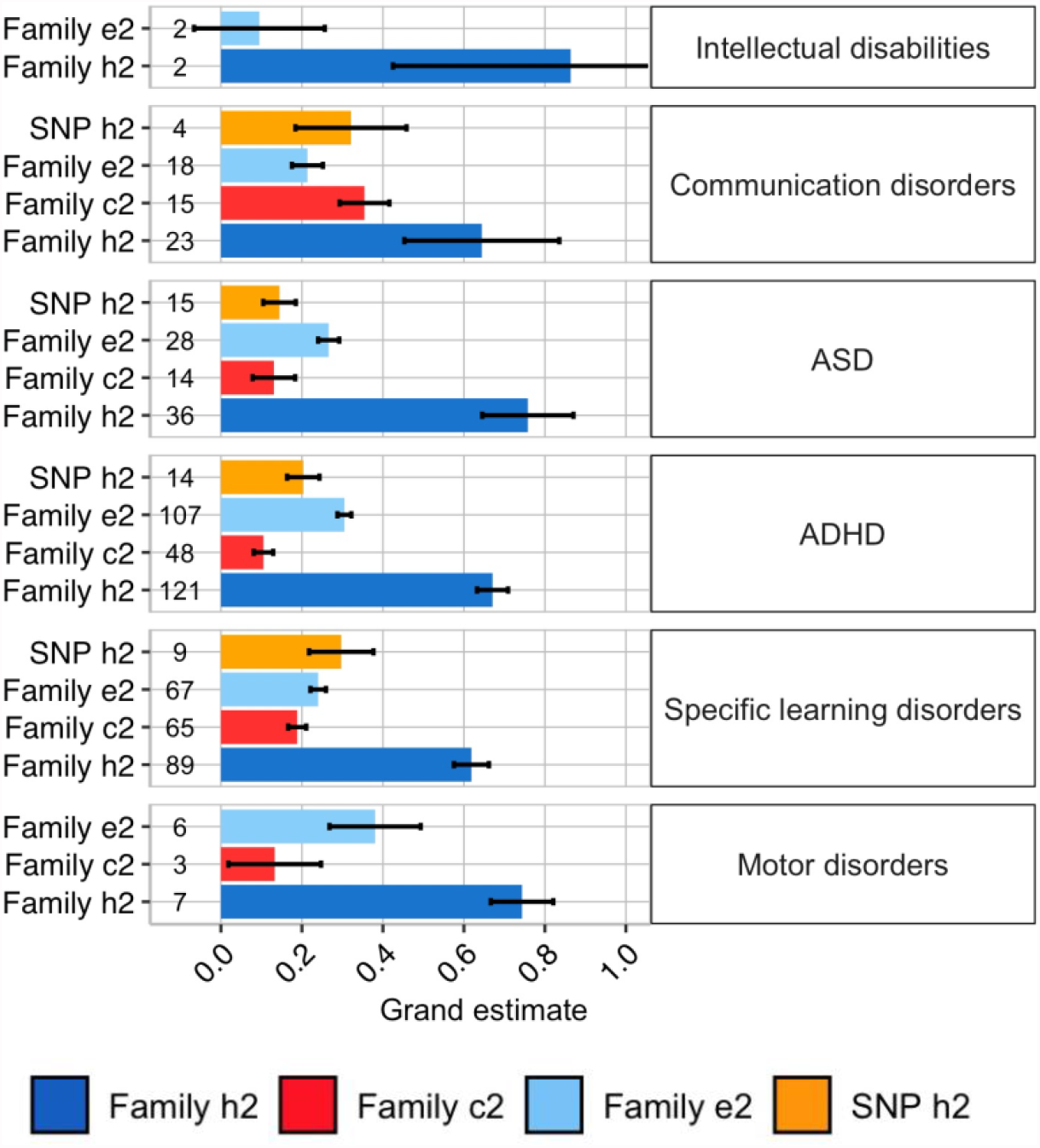
Meta-analytic family and SNP-based heritability (h^2^) shared environmental influences (c^2^) and nonshared environmental influences (e^2^) on variation in neurodevelopmental disorders (NDDs). Numbers preceding bars on the y-axis denote the number of studies identified that provided estimates for specific NDDs. Error bars signify standard errors of the grand estimates. The results for c^2^and e^2^ are discussed in **Supplementary Note 1** and reported in **Supplementary Table 1**.

#### SNP heritability (SNP h^2^)

Out of the total of 29 SNP-based studies that were identified, the only disorders that were addressed by at least two independent studies^49^, included ASD (15 studies), ADHD (14 studies), specific learning disorders (9 studies) and communication disorders (4 studies). SNP heritability across all NDDs was moderate (0.19, SE= 0.03) and ranged between 0.15 (SE= 0.04) for ASD to 0.30 (SE= 0.14) for communication disorders (**Figure 3** and **Supplementary Table 1**). SNP heritability estimates were not found to differ significantly across disorders, although the degree of precision in the estimates varied substantially depending on the sample size and number of individual studies included per disorder.

### Genetic factors underlie the widespread co-occurrence between NDDs during childhood and adolescence

When compared to the vast number of studies that had examined the aetiology of individual differences in each NDD, only a limited body of research (37 studies) had investigated the co-occurrence between NDDs in childhood and adolescence applying genetically informative methods. In fact, for some of the disorders, we were unable to find two independent statistics^49^, and therefore could not provide a meta-analytic estimate.

#### Family-based genetic correlations (rA)

When considering family-based designs (see ***Method*** and **Supplementary Note 3**), a sufficient number of studies to allow for meta-analysis was obtained for the following NDD pairs: ADHD & specific learning disorders (15 studies), ASD & ADHD (6 studies), ADHD & motor disorders (2 studies), communication disorders & motor disorders (2 studies), and communication disorders & specific learning disorders (2 studies). Only one study was identified for the following pairs of NDDs: ASD & communication disorders, ASD & specific learning disorders, ASD & motor disorders, and specific learning disorders & motor disorders, therefore these studies could only be included in the transdiagnostic meta-analysis, capturing the degree of genetic and environmental co-occurrence across all NDD pairs. In addition, 9 studies examined the co-occurrence between subtypes of specific learning disorders, such as dyslexia & dyscalculia, these studies have been included in the transdiagnostic meta-analysis and results of these finer-grained analyses are reported in **Supplementary Note 2**.

We first meta-analyzed genetic correlations across all NDD categories (transdiagnostic genetic co-occurrence), this yielded a moderate grand estimate of rA= 0.36 (SE= 0.12). When considering NDD categories separately, the strongest genetic overlaps were found between ADHD & motor disorders (rA= 0.90, SE= 0.82), and between ASD & ADHD (rA= 0.67, SE= 0.30), while the weakest genetic correlation was found for the association between ADHD & specific learning disorders (rA= 0.07, SE= 0.12; **Figure 4** and **Supplementary Table 3**).

**Figure 4.**
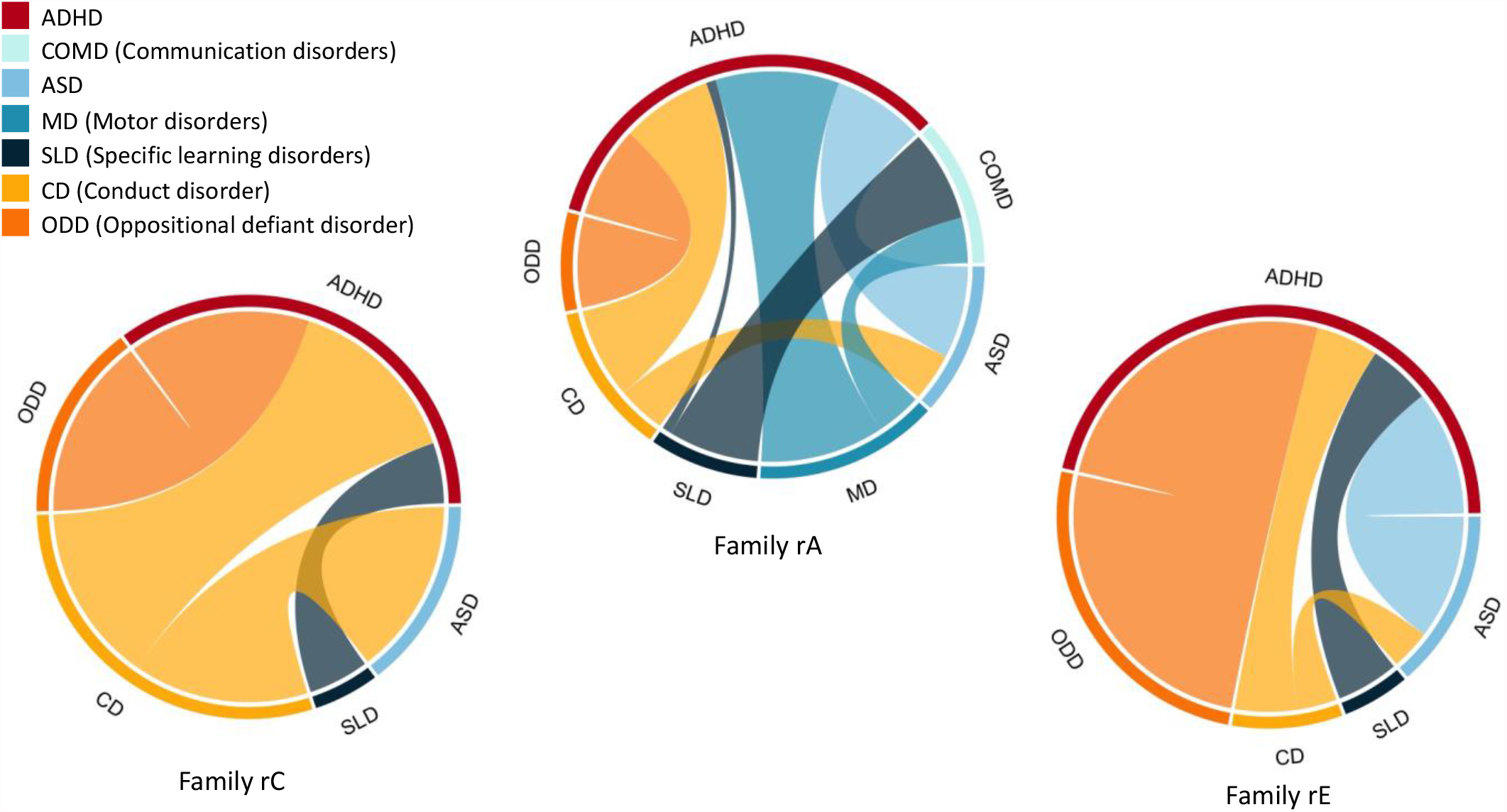
Strength of the meta-analytic genetic (rA), shared environmental (rC) and nonshared environmental (rE) correlations between neurodevelopmental disorders (NDDs) and their homotypic (other NDDs) and heterotypic (disruptive, impulse control and conduct disorders (DICCs)) co-occurrences. The outer layer of each circle shows all the different NDDs and DICCs for which meta-analytic correlation estimates could be computed. Each colored connector path indicates the strength of association between disorders, the thicker the connector path, the stronger the correlation between two disorders. The results for family rC and rE are presented in **Supplementary Note 1 and Supplementary Tables 3 and 5**.

#### SNP-based genetic correlations (rG_SNP_)

SNP-based designs in childhood and adolescent samples exclusively focused on the association between ASD & ADHD (5 studies) and subtypes of specific learning disorders (1 study). The transdiagnostic genetic correlation obtained meta-analyzing SNP-based designs was 0.39 (SE= 0.19) (**Supplementary Table 8**), in line with the estimate obtained from family-based designs. A grand genetic correlation of 0.20 (SE= 0.14) was found for the co-occurrence between ADHD and ASD. The one remaining study examined the co-occurrence between dyslexia & dyscalculia-related traits, specifically reading and mathematics abilities, which were strongly correlated (rG_SNP_= 0.74, SE= 0.17)^50^.

### NDDs share as much of their genetic underpinnings with their homotypic and heterotypic co-occurrences

Our third aim was to obtain meta-analytic estimates of the genetic associations between NDDs and DICCs. Our search yielded only 15 eligible family-based studies, and no SNP-based studies investigating the genetic overlap between NDDs and DICCs in childhood and adolescence. Meta-analytic genetic correlations could only be calculated for a few NDD and DICC pairs, namely ADHD & conduct disorder (6 studies), ADHD & oppositional defiant disorder (6 studies) and ASD & conduct disorder (3 studies). In addition, we identified 1 study that examined the co-occurrence between specific learning disorders & disruptive behaviour, finding a weak negative genetic correlation (rA= -0.14, SE= 0.06) ^51^.

#### Family-based genetic correlations (rA)

Across all co-occurrences between NDDs and DICCs (15 studies), the grand genetic correlation was 0.62 (SE= 0.20). A similarly strong genetic correlation was observed between ADHD & conduct disorder (6 studies) and ADHD & oppositional defiant disorder (6 studies): rA= 0.66 (SE= 0.36) and rA= 0.66 (SE= 0.18), respectively; a similar level of aetiological overlap to that observed between strongly genetically correlated NDDs such as for example ADHD & ASD (**Supplementary Table 5**). On the other hand, the genetic overlap between ASD & conduct disorder (3 studies) was much weaker, with a meta-analytic genetic correlation of 0.35 (SE= 0.10; **Figure 4)**.

### While the aetiology of NDDs is comparable for males and females, their co-occurrences differ by sex

NDDs do not affect males and females equally, males are four times more likely to be diagnosed with NDDs^52,53^. Studies have suggested that these differences in prevalence may be caused by quantitative genetic sex differences, differences in the degree to which genes influence variation in NDDs in males versus females^54^. To provide an overview of sex differences in NDDs, we conducted separate meta-analyses including all studies that had reported sex-specific estimates.

#### Sex differences the genetic aetiology of NDDs: Family-based heritability (h^2^)

We identified 68 family-based studies that investigated the genetic aetiology of individual differences in NDDs in male samples and 67 studies that reported estimates for female samples. Out of all studies involving sex-stratified samples, 38 studies focused on ADHD, 21 studies on ASD, 8 studies on specific learning disorders, 4 studies on communication disorders and 2 studies on motor disorders. Across all NDDs, family-based heritability was not significantly different between males and females (h^2^= 0.65, SE= 0.06 in males and 0.67, SE= 0.06 in females). Distributions of sex-specific family-based variance components for all NDDs, except for motor disorders for which a sufficient number of studies (>1) was not identified, are presented in **Figure 5** and **Supplementary Table 13**.

**Figure 5.**
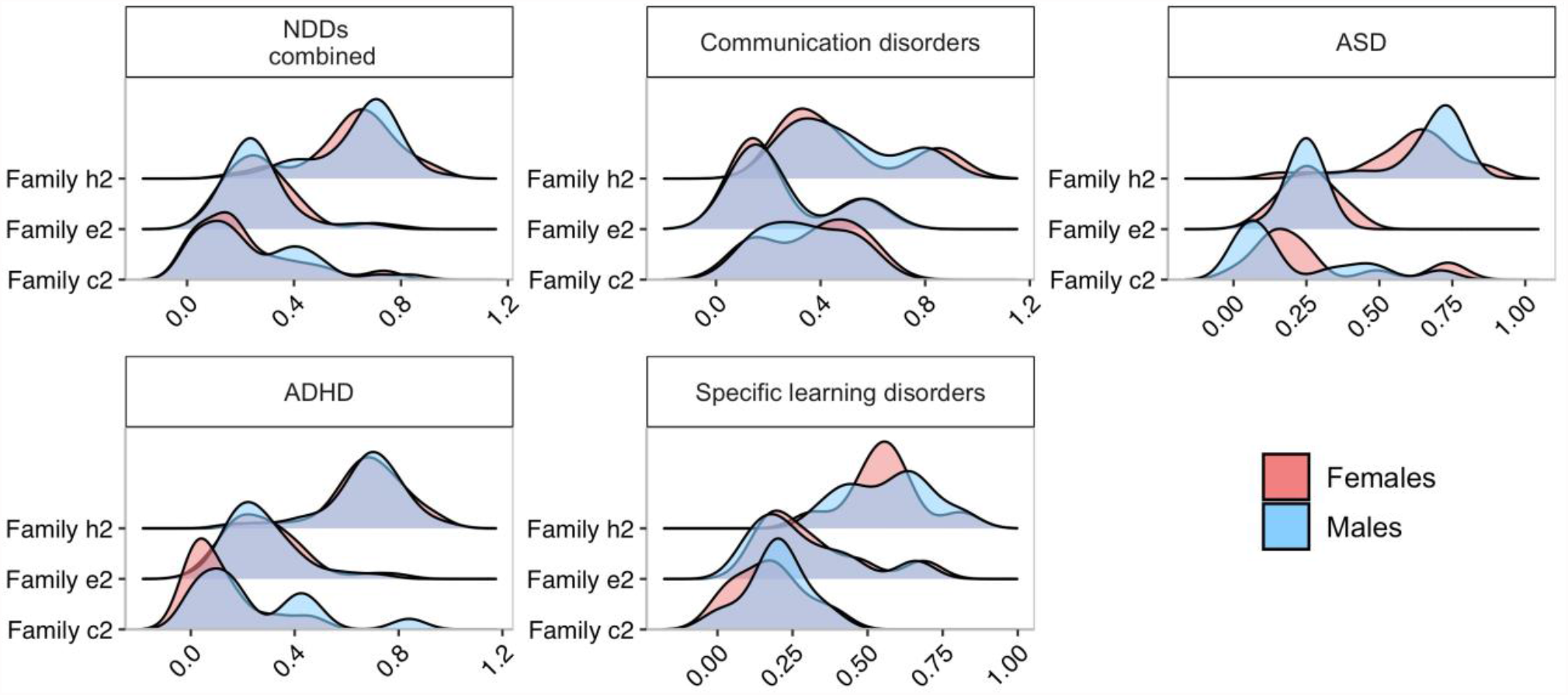
Distributions of the sex-specific meta-analytic estimates for the heritability (h^2^) and environmental contributions to neurodevelopmental disorders (NDDs). The top left panel shows the distributions of sex-specific estimates for the transdiagnostic meta-analysis, while the remaining panels the same estimates for specific NDDs for which a sufficient number of studies (>2) reporting sex-specific estimates was identified. The results for sex-specific c^2^, e^2^, rC and rE estimates are presented in **Supplementary Note 1 and Supplementary Table 13-15**.

#### Sex differences the genetic aetiology of NDDs: SNP heritability (SNP h^2^)

Marked differences in SNP heritability were observed between males and females across all NDDs (0.19, SE= 0.07 for males and 0.09, SE= 0.10 for females). However, these estimates were based on the only two studies to date that had calculated the SNP heritability of ASD and ADHD separately by sex (**Supplementary Table 13)**.

#### Sex differences in genetic overlap between NDDs

We identified only 4 family-based studies that had examined homotypic co-occurrences of NDDs in males and only 2 studies in females. Half of these studies considered the overlap between ASD & ADHD. The other half had considered the comorbidity between ASD & communication disorders (1 study in both male and female) and between developmental coordination disorder & tic disorder, two subtypes of motor disorder (1 study in males only). The grand family-based genetic correlation across all NDDs was estimated at 0.86 (SE= 0.58) for males and 0.25 (SE= 0.36) for females (**Supplementary Table 14)**.

Sex-specific grand estimates of family-based genetic correlations between specific disorders could not be calculated due to the limited number of available studies. The only exception was the co-occurrence between ASD & ADHD in males, where 2 studies were identified (rA= 0.79, SE= 0.42) (**Supplementary Table 14)**. SNP-based genetic correlations between NDDs could not be calculated for males and females separately due to a lack of studies that examined these associations separately by sex in samples of children and adolescents.

#### Sex differences in genetic overlap between NDDs and DICCs

Sources of co-occurrence between NDDs and DICCs could only be estimated between ADHD & conduct disorder and only in in females. In fact, one out of the only two studies that examined the sex-specific co-occurrence between ADHD and conduct disorders used a female-only sample. Hence, we could only meta-analyse the co-occurrence between ADHD & conduct disorder in females. We found a meta-analytic genetic correlation of 0.75 (SE= 0.58) (**Supplementary Table 15**).

### Genetic influences on NDDs are stable over development

We investigated developmental change and continuity in the relative contribution of genetic factors to NDDs by examining age-related differences in their aetiology and sources of their homotypic and heterotypic co-occurrences. We distinguished between the three following developmental stages: childhood (4-7 years), middle childhood (8-10 years) and adolescence (11-24 years). We grouped estimates in either of those three categories or across multiple categories, for example childhood & middle childhood (4-10 years), middle childhood & adolescence (8-24 years) and childhood & adolescence (4-24 years).

#### Age-related differences in the genetic aetiology of NDDs: Family-based heritability (h^2^)

Across all NDDs, 54 family-based studies reported estimates in childhood (4-7 years), 54 studies reported estimates in middle childhood (8-10 years) and 79 studies reported estimates in adolescence (11-24 years). The remaining studies involved populations whose age range spanned across categories, i.e., childhood & middle childhood (4-10 years; 14 studies), middle childhood & adolescence (8-24 years; 50 studies) and childhood & adolescence (4-24 years; 40 studies). We investigated age-related differences in heritability including all NDD categories (**Figure 6A**), with the exception of motor disorders for which we did not identify enough studies (>1) per age category. All estimates with standard errors, including those for age cross-categories are presented in **Supplementary Table 16**.

**Figure 6.**
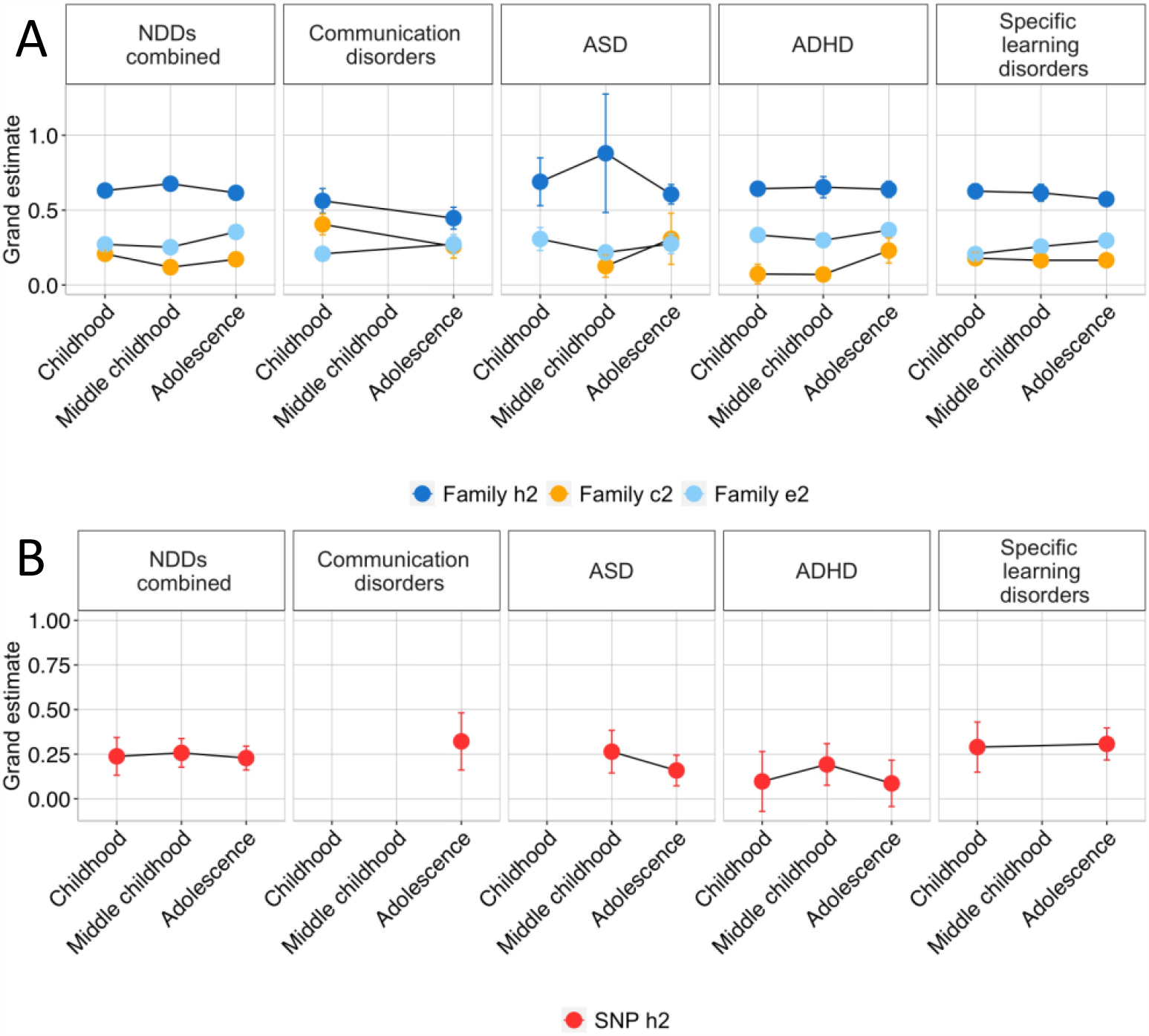
Age-related differences in family-based heritability (h^2^), shared (c^2^) and nonshared (e^2^) environmental influences on neurodevelopmental disorders (NDDs) (panel A) and SNP heritability (panel B). Developmental stages include childhood (4-7 years), middle childhood (8-10 years) and adolescence (11-24 years). Error bars represent standard errors of grand estimates. For intellectual disabilities and motor disorders we could not identify a sufficient number of studies (>1) reporting age-dependent estimates and we were consequently unable to derive meta-analytic estimates. The results for age-stratified c^2^, e^2^, rC and rE are reported in **Supplementary Note 1** and **Supplementary Tables 16-18**.

Across all NDDs, grand heritability remained relatively stable developmentally, with the estimate of 0.63 (SE= 0.03) in childhood, slight increase in middle childhood (0.68, SE= 0.04) and a subsequent drop back to 0.62 (SE= 0.08) in adolescence. This trend was consistent for some specific disorders (e.g., ASD and ADHD) but not for others (e.g., communication disorders and specific learning disorders) for which genetic influences decreased developmentally (**Figure 6A; Supplementary Table 16**).

#### Age-related differences in the genetic aetiology of NDDs: SNP heritability (SNP h^2^)

Out of a total of 29 SNP-based studies that were identified, 13 included adolescent samples, 7 samples in middle childhood and 6 samples in childhood, while 11 studies reported estimates across childhood & adolescence. SNP heritability was stable developmentally across NDDs, and the developmental trajectory mirrored that of family-based heritability (SNP h^2^= 0.24, SE= 0.11 in childhood; 0.26, SE= 0.08 in middle childhood and 0.23, SE= 0.07 in adolescence) (**Figure 6B; Supplementary Table 16**). For ASD, ADHD and specific learning disorders, the specific NDDs for which grand estimates could be calculated, the developmental trends were consistent with those observed for family-based heritability (**Figure 6B; Supplementary Table 16)**.

#### Age-related differences in genetic overlap between NDDs

Overall, we could not explore developmental trends in genetic correlations using either method due to a lack of available studies, the only exceptions were grand estimates for adolescence and across age categories (see **Supplementary Tables 17-18)**. Genetic correlations obtained for adolescent samples only were in line with those obtained for the total sample (for example, when considering the co-occurrence between ASD & ADHD the genetic correlation was 0.66 (0.49) in adolescent samples and 0.67 (0.30) across all age categories).

### Categorical versus continuous measurement of NDDs

Although we meta-analysed categorical (binary phenotypes, such as clinical diagnoses and cut-offs) and quantitative (sub-threshold symptom counts or test/questionnaire scores) measures together, we also report separate grand estimates for both measurement types across NDDs and their co-occurrences. Across all NDDs, categorical measures were observed to yield significantly higher family-based heritability estimates if compared to continuous phenotypes (0.77, SE= 0.07 vs. 0.64, SE= 0.03). However, the opposite was found for SNP-based heritability (0.17, SE= 0.03 for categorical measures vs. 0.25, SE= 0.06 for quantitative assessments). Differences in sources of variation in specific NDDs, as well specific homotypic and heterotypic co-occurrences are presented in **Supplementary Note 4, Supplementary Figure 22, and Supplementary Tables 25-27**.

### NDDs and co-occurring disorders are almost exclusively researched in Western countries

Research into the genetic aetiology of NDDs and of their homotypic and heterotypic co-occurrences is largely limited to Western countries, even though, according to the Global Burden of Disease study^55^, the prevalence of diagnosed NDDs is not uniform across the globe. The current meta-analysis provides an overview of how behaviour genetics research into NDDs is distributed across countries and continents.

#### Geographical differences in the genetic aetiology of NDDs: Family-based heritability (h^2^)

Out of the 237 studies investigating sources of individual differences in NDDs, 41% (96 studies) involved samples and cohorts based in the United Kingdom, 77 studies samples based in the United States, 24 studies Swedish samples, 19 studies Dutch samples, 11 studies Australian samples, 7 studies Canadian samples, 4 studies samples from China, and 2 studies samples from Norway. Other countries that contributed to the total grand estimate but did not have enough estimates for separate meta-analysis (i.e., only 1 study found from each country), included Finland, Japan, South Korea, and Italy. Estimates differed significantly across Countries. Considering all NDDs, the highest meta-analytic family-based heritability was estimated for Australian and Swedish samples (0.76, SE= 0.17 and 0.74 SE= 0.05, respectively), while the lowest was obtained for Canadian cohorts (0.43, SE= 0.09) (**Figure 7A; Supplementary Table 19**).

**Figure 7.**
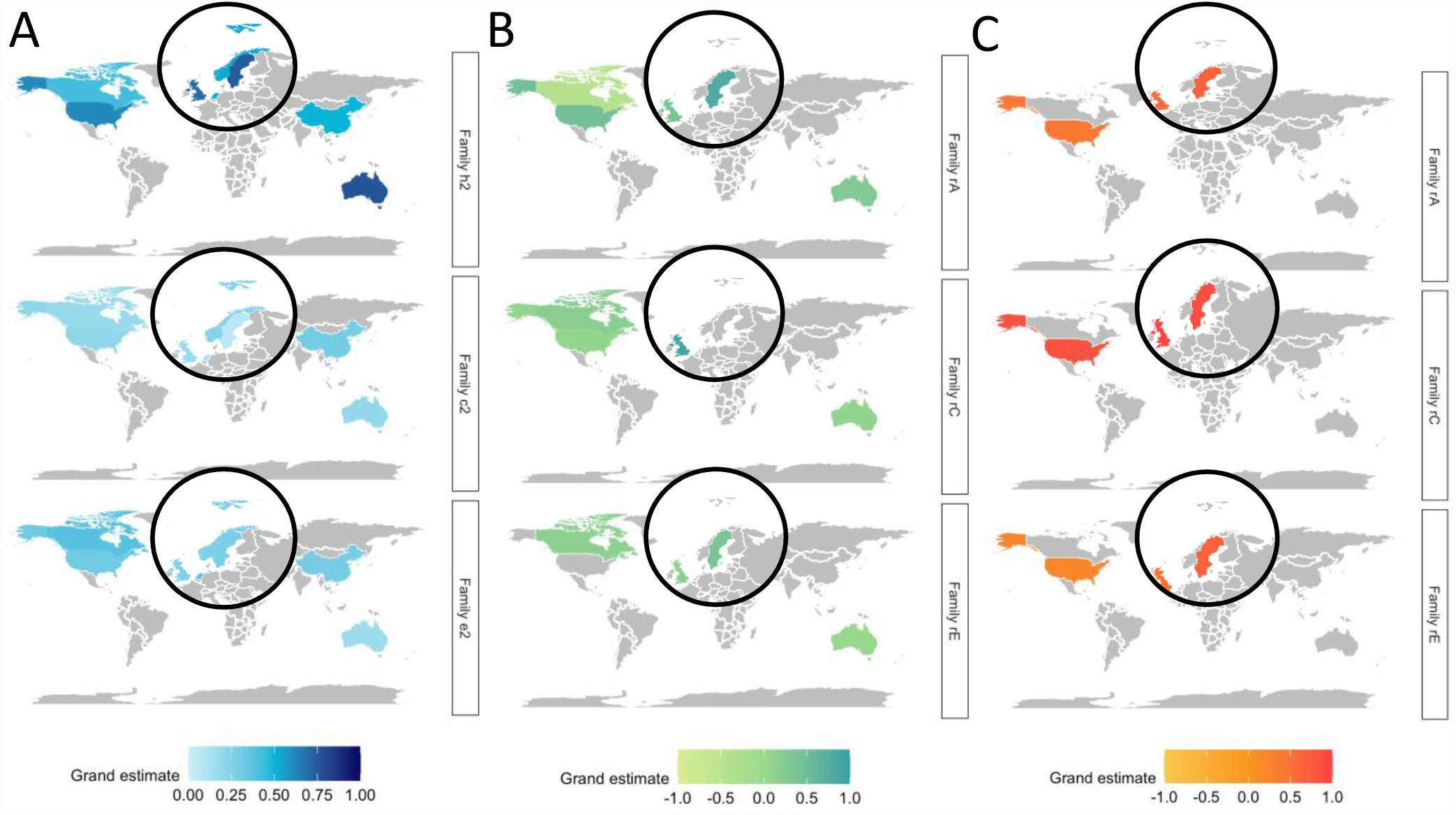
Geographical differences in the heritability of NDDs and their co-occurrences. *Panel A* illustrates differences in family-based heritability (h^2^) shared environmental (c^2^) and nonshared environmental (e^2^) influences across all neurodevelopmental disorders (NDDs). *Panel B* illustrates geographical differences in the genetic (rA), shared environmental (rC) and nonshared environmental (rE) overlap between NDDs. *Panel C* reports geographical differences in rA, rC and rE between NDDs and disruptive, impulse control and conduct disorders (DICCs). The areas shaded in grey are regions for which not enough relevant studies were identified (<2 studies). The circles show the zooming-in on Europe to allow for better visibility. The results for c^2^and e^2^as well as rC and rE are discussed in **Supplementary Note 1** and reported in **Supplementary Tables 19-21**.

When considering specific NDDs, these were investigated with different frequencies across countries: the aetiology of intellectual disabilities was exclusively investigated in Swedish cohorts (2 out of 2 studies), from where most studies addressing sources of variance in motor disorders also came from (4 out of a total of 7 studies). Communication disorders were mostly researched in the United Kingdom (17 out of a total of 23 studies), as were ASD (20 out of a total of 36 studies) and ADHD (42 out of a total of 121 studies). On the other hand, specific learning disorders were mostly investigated in the United States (47 out of a total of 89 studies).

#### Geographical differences in the genetic aetiology of NDDs: SNP heritability (SNP h^2^)

Studies exploring SNP heritability of NDDs focused entirely on European cohorts and were primarily conducted in the United Kingdom and the Netherlands (14 and 3 out of 29 SNP-based studies in total) (**Supplementary Table 19**).

#### *Geographical differences in genetic overlap between NDDs: Family-based genetic* correlations *(rA)*

Sources of homotypic co-occurrence with NDDs were investigated in 37 independent family-based studies, out of which the majority was conducted in the United Kingdom (49%) and United States (30%). The highest genetic correlation across all co-occurrences was estimated in Swedish cohorts (0.80, SE= 0.26 across 3 studies), while the lowest grand genetic overlap was estimated in Canadian samples (−0.44, SE= 0.24 across only 2 studies which investigated the association between ADHD and specific learning disorders; **Figure 7B; Supplementary Table 20**).

The genetic aetiology of the co-occurrence between ASD & ADHD during childhood and adolescence was exclusively researched in the United Kingdom and Sweden (3 out of a total of 6 studies each). The co-occurrence between ADHD & motor disorders was only explored by two studies, one conducted in Sweden and the other one in Australia. Most studies examining the genetic overlap between ADHD & specific learning disorders came from the United States (8 out of a total of 18 studies), whereas the overlap between communication disorders & motor disorders was only addressed by 2 studies conducted in the United Kingdom and Japan.

#### Geographical differences in genetic overlap between NDDs: SNP-based genetic correlations (rG_SNP_)

SNP-based studies (6 in total) addressing the co-occurrence between NDDs were exclusively conducted in combined samples from the United Kingdom and Denmark (**Supplementary Table 20**).

#### Geographical differences in genetic overlap between NDDs and DICCs: Family-based genetic correlations (rA)

A total of 15 family-based studies addressing the co-occurrence between NDDs and DICCs were identified, 40% of which were conducted in the United Kingdom, 20% in the United States and 20% in Sweden. Studies yielded consistently strong estimates of genetic correlations across the three regions: genetic correlations of 0.60 (SE= 0.29); 0.42 (SE= 0.15) and 0.68 (SE= 0.41), respectively (**Figure 7C** and **Supplementary Table 21**). The remaining 20 % of studies were conducted in Australia, Finland, and South Korea, but could not be meta-analysed separately as only one estimate was available for each country.

In terms of specific co-occurrences between NDDs and DICCs, half of the studies that explored genetic overlap between ADHD & conduct disorder, and ADHD & oppositional defiant disorder were conducted in the United States (3 studies each). Three out of 4 studies examining the association between ASD & conduct disorder were conducted in the United Kingdom and 1 study in Sweden.

### NDDs and co-occurring disorders are mostly researched in cohorts of European ancestry

Individuals of European ancestry represent 16% of the global population but 80% of participants in genomic (i.e., DNA-based) research^56^. This *Eurocentric bias*^57^ has created a major gap in our knowledge of the genetic aetiology of NDDs and their homotypic and heterotypic co-occurrences in non-White populations. In the following sections we highlight this major issue illustrating the disparity in the number of studies that have investigated ancestrally diverse versus ancestrally homogeneous samples, and how meta-analytic estimates differ at different levels of sample diversity.

#### Diversity-related differences in the genetic aetiology of NDDs: Family-based heritability (h^2^)

Given the general lack of diversity in participants’ ancestry, we could only examine this issue by calculating how samples differed between each other in terms of their percentage of participants of European ancestry. A related issue was also that less than half of the studies reported information on the ancestral composition of their sample (97 out of the 237 studies).

Across all NDDs, heritability was observed to increase with increasing percentage of participants of European ancestry, from 0.46 (SE= 0.07) when they constituted less than half of the sample to 0.66 (SE= 0.06) when 100% of the sample was of European ancestry (**Figure 8; Supplementary Table 22**). This trend was particularly observed for ADHD, where the heritability increased from 0.41 (SE= 0.12) in samples where European ancestry participants were the minority (less than 50%) to 0.67 (SE= 0.04) in samples where European ancestry participants were the totality. On the other hand, genetic influences on communication disorders and specific learning disorders remained stable across ancestral compositions: For communication disorders, heritability estimates ranged between 0.59 (SE= 0.27) in samples less than 75-99% of European ancestry to 0.56 (SE= 0.09) in samples 100% of European descent. For Specific learning disorders, heritability was 0.54 (SE= 0.16) in samples where European ancestry participants were in the minority vs. 0.61 (SE= 0.04) in samples 100% of European ancestry.

**Figure 8.**
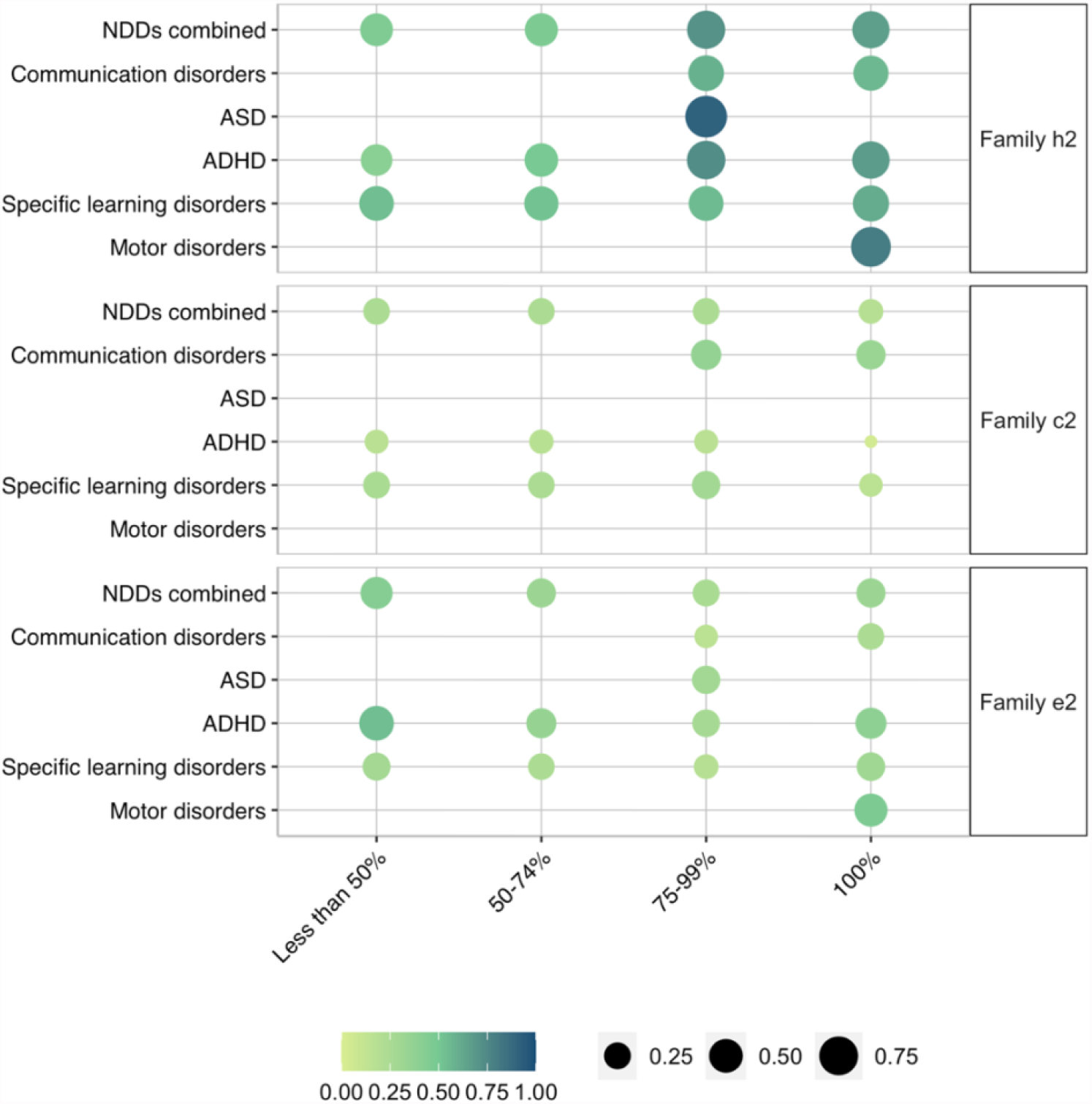
Changes in family-based heritability (h^2^), shared (c^2^) and nonshared (e^2^) environmental influences on neurodevelopmental disorders (NDDs), as a function of sample ancestral composition. Given the general lack of diversity, ancestral composition could only be quantified, and consequently meta-analysed, as percentage of the sample being of European ancestry, different categories based on these percentages are depicted on the x-axis. Grand estimates of h^2^, c^2^ and e^2^ are reflected in the size and colour intensity of each circle, the larger and darker the circle, the higher the grand estimate. Findings regarding c^2^ and e^2^ as well as environmental correlations are discussed in **Supplementary Note 1** and reported in **Supplementary Tables 22-24**.

#### Diversity-related differences in the genetic aetiology of NDDs: SNP heritability (SNP h^2^)

We did not identify SNP-based studies that used samples other than 100% European ancestry in populations of children and adolescents.

#### Ancestry-related differences in genetic overlap between NDDs: Family-based genetic correlations (rA)

Differences in sources of co-occurrence between NDDs could only be estimated for the genetic overlap between all NDDs, where a total of 6 studies were identified. Two studies (one focusing on the co-occurrence between ADHD & specific learning disorders, and the other on the co-occurrence between subtypes of specific learning disorders) reported estimates for sample where participants were between 75% and 99% of European ancestry, while 4 studies (2 on the co-occurrence between ADHD & specific learning disorders, and 2 on the co-occurrence between subtypes of specific learning disorders) included samples where 100% of the participants were of European descent. The meta-analytic genetic overlap between NDDs decreased, albeit not digniifcantly, from 0.63 (SE= 0.44) in samples where 75-99% of European ancestry to 0.54 (SE= 0.10) in samples entirely of European ancestry (**Supplementary Table 23)**.

#### *Ancestry-related differences in the heterotypic co-occurrences of NDDs: Family-based* genetic *correlations (rA)*

Estimating the sources of co-occurrence between NDDs and DICCs by percentage of sample diversity was similarly challenging as we could identify only 5 studies that included the relevant information. Out of the total number of studies, 3 involved samples of between 75% and 99% participants of European ancestry and focused on examining the genetic overlap between ADHD & conduct disorder and ADHD & oppositional defiant disorder, while 2 involved samples of 100% European descent and examined the genetic correlations between ADHD & oppositional defiant disorder and ADHD & disruptive behaviour. The meta-analytic genetic overlap between NDDs and DICCs increased, albeit not significantly, from 0.57 (SE= 0.25) in samples involving less than 100% of European ancestry participants to 0.71 (SE= 0.31) in 100% European ancestry samples (**Supplementary Table 24)**.

### Bias and heterogeneity assessment

We applied I^2^ statistics to assess heterogeneity in the estimates. The results of these analyses are reported in **Supplementary Note 10, Supplementary Tables 7-9** and **Supplementary Figure 6**. We applied Egger’s regression and inspected funnel plots to examine the impact of publication bias on our results, the outcomes of these analyses are reported in **Supplementary Note 11** and **Supplementary Tables 10-12** and Supplementary Figures **5-21**.

## Discussion

The findings of the current meta-analysis have important implications which have the potential to guide future research strategies, clinical and educational practice. First, by providing estimates of the relative contribution of genetic factors to all NDDs, our work responds to the need of moving beyond the nearly exclusive research focus on ASD and ADHD, and effectively synthesizes our knowledge across all NDDs. Second, by providing an account of the genetic overlap between NDDs, we highlight how genetic influences are implicated in the co-occurrence between multiple NDDs, identifying patterns of shared aetiological liability between neurodevelopmental conditions. Third, by providing a synthesis of the literature on the co-occurrence between NDDs and DICCs we highlight how these two separate groups of disorders identified by the DSM-5 share as much of their genetic aetiology as do disorders all classified as NDDs.

Our work provides meta-analytic evidence for the substantial heritability of all neurodevelopmental conditions, particularly when considering family-based studies, which indicated that around two thirds of the variation in NDDs is accounted for by genetic differences between children and adolescents. Heritability estimates differed, albeit not significantly, between NDDs, with the highest estimates obtained for intellectual disabilities and motor disorders, and the lowest estimates, despite still accounting for over 60% of the variation, obtained for specific learning disorders and communication disorders.

Although males are four times more likely to be diagnosed with NDDs than females^52,53^, we showed that, when meta-analysed, genetic effects on NDDs do not differ by sex. We also showed that genetic sources of variation in NDDs are remarkably stable across developmental stages, and this developmental stability was observed across all NDDs, irrespectively of the relative contribution of genetic factors to each disorder. Genetic effects were also mostly consistent when we separated studies that had considered diagnoses and clinical cut-offs from studies that had quantified NDDs as continuous traits.

Interestingly, we found that the genetic contributions to NDDs differed substantially as a function of geography. This highlights how estimates of genetic effects on disorders are sensitive to different environmental contexts. The high sensitivity to context of genetic effects has been observed for several other behavioural traits, including educational attainment, for which estimates of genetic effects not only differ as a function of geography, but also as a function of different historical times and political contexts^58,59^.

Our work on geographical differences on genetic effects on NDDs also highlighted the major gap in our knowledge of the aetiology of NDDs in non-Western countries, a gap that is only exceeded by the lack of ancestral diversity observed across all studies of NDDs. Importantly, the current study pointed to how genetic influences on NDDs were substantially reduced in more ancestrally diverse samples, again highlighting how heritability estimates are inextricably linked to our social context^60,61^, in a sense that increased ancestral homogeneity within the sample likely entails increased environmental homogeneity, reducing the environmental influences and inflating heritability in these populations.

The lack of diversity in genetic research remains its most striking limitation to date, particularly when considering DNA-based methods, limiting the extension of genetic findings to the entire population^62,63^. Limited research resources in under-represented populations are likely to have profound cascading effects for future advances in clinical practice, including pharmacological and behavioural treatment. Fortunately, there are major initiatives underway to re-balance these biases^64,65,66^.

Heritability estimates obtained from studies with SNP-based designs were overall modest, and did not differ significantly between disorders, although the degree of precision around these estimates varied substantially between conditions. DNA-based heritability estimates could not be obtained for motor disorders or intellectual disabilities. The gap in the heritability estimated from family studies and DNA-based studies is known as *missing heritability*^67^, and is observed for virtually all complex human traits, from height to cognitive ability, to NDDs. The current inability of GWA studies to tag interactive effects between genes (epistatic effect), and between genes and environments (GE interaction) may account for this missing heritability gap^68^. It is also possible that rare variants that are not tagged by SNP arrays commonly used in GWA studies play a substantial role^69^. Lastly, it has been proposed that family-based heritability estimates might be inflated^70^.

Our second aim was that of providing a clear account of how close NDDs are to one another aetiologically. We found that, while meta-analytic estimates indicated moderate genetic overlap, the degree of heterogeneity in these associations across disorders was large. We found a substantial genetic correlation between ASD and ADHD, ADHD and motor disorders, and communication disorders and specific learning disorders. On the other hand, genetic overlap was only moderate between communication disorders and motor disorders, and very weak between ADHD and specific learning disorders, which is consistent with the degree of symptom resemblance across these disorders.

Although we were able to explore general patterns of variation and co-occurrence, the aetiology of specific NDDs and of their associations could not be comprehensively characterised. The research gaps that we identified highlight an imbalance in focus across NDDs in developmental behavioural genetic research. When considering our first aim, we could only identify 2 family-based studies that investigated the genetic contributions to intellectual disabilities, if compared to 121 family-based and 14 SNP-based studies identified for ADHD, and 36 family-based and 15 SNP-based studies identified for ASD. This lack of research on intellectual disabilities, a neurodevelopmental disorder affecting 2.5% of children in the United Kingdom^71,72^ more than double the prevalence rate of ASD^73^ is reflected in, and likely partly due to, the lack of funding bodies devoted to researching NDDs other than ASD and ADHD, as well as a lack of publicly available data repositories and resources (e.g., ^74,75,76^).

We also identified very few studies that had examined the aetiology of motor disorders, another neurodevelopmental condition showing significant prevalence rates of 5-6% in school aged children^77^. This unbalanced research focus, that extends far beyond genetically informative research to touch developmental and therapeutic research^78,79,80,81^, has led to an uneven distribution of knowledge, which could lead to limited access to interventions for children with NDDs other than ASD, ADHD, and Dyslexia^82^.

Considering our second aim, no genetically informative studies were identified that had examined the co-occurrence between intellectual disabilities and all other NDDs, and between ADHD & communication disorders. Only one study was identified on the genetic overlap between ASD & communication disorders^33^, ASD & specific learning disorders^27^, ASD & motor disorders^27^ and between specific learning disorders & motor disorders^27^; therefore, preventing us from meta-analysing these NDD pairs separately.

The lack of equity in focus across NDDs was pronounced in analyses addressing our third aim. Sources of co-occurrence between NDDs and DICCs could only be investigated between ADHD & conduct disorder, ADHD & oppositional defiant disorder and between ASD & conduct disorder. Considering that in the DSM-5 the DICCs category comprises 8 distinct disruptive disorders, this highlights a major gap in our knowledge of the role that genetic and environmental factors play in the co-occurrence between all NDDs and DICCs. We hope that highlighting these major knowledge gaps will lead to future research efforts aimed at making developmental genetic research more diverse, consequently allowing for greater inclusion opportunities.

To conclude, this meta-analysis extends our knowledge of NDDs in several directions. First, it provides a holistic view of genetic and environmental contributions to all NDDs and commonly co-occurring disorders over development, revealing that NDDs are just as strongly genetically correlated with other NDDs, as they are with DICCs. Second, it identifies a lack of balance in the research efforts that have been invested across different disorders, which calls for future genetic research to focus on less investigated NDDs. Third, it provides knowledge about patterns of aetiological co-occurrence between NDDs, as well as between NDDs and DICCs, which we hope will inform clinical and educational diagnostics and practice, resulting for example in expanded diagnostic screening.

## Methods

The protocol for the current meta-analysis was registered with the international prospective register of systematic reviews (PROSPERO) and can be accessed at the following link: https://www.crd.york.ac.uk/prospero/display_record.php?RecordID=230158. This meta-analysis was conducted in line with the Preferred Reporting Items for Systematic Reviews and Meta-Analyses (PRISMA) guidelines^83^. Code is available at https://github.com/CoDEresearchlab/Meta_analysis_NDDs_DICCs.

### Identification of relevant studies

A total of 296 studies were included in the meta-analysis (**Figure 2**). Studies were identified during three searches: the primary search (**Supplementary Figure 23A**) conducted on the 20^th^ of January 2021, the secondary (confirmatory) search (**Supplementary Figure 23B**) conducted on the 15^th^ of April 2021 and the additional search of other relevant meta-analyses and reviews finalised on the 4^th^ of May 2021. Searches were conducted across three platforms: Web of Science, Ovid Medline, and Ovid Embase and the outputs managed with the aid of Covidence (https://www.covidence.org/). An in-depth description of indexes, timespans, search strategy and key words is included in **Supplementary Note 5**. All studies included in the meta-analysis are listed in **Supplementary Tables 28-33**.

### Screening and inclusion criteria

After the initial searches were conducted and duplicate studies removed, 8,087 studies met the criteria for the first stage of screening, which involved title and abstract scanning. All titles and abstracts were screened by two independent, blinded reviewers to ensure inter-rater agreement. Conflicts were resolved by a third independent reviewer. After this initial screening phase, 6,834 studies were excluded as deemed not relevant for the purpose of the current meta-analysis.

The title and abstract screening process resulted in a total of 1,253 potentially eligible studies. The full text of each study was screened by two independent, blinded reviewers. Reviewer discrepancies were identified and resolved by a third independent reviewer. This resulted in 289 eligible articles. In addition, during full text screening, relevant review articles, meta-analyses, editorials, and conference abstracts were flagged to aid the potential discovery of further relevant studies by either screening the References sections or contacting the authors of conference abstracts. Through this process 7 additional studies were identified, which resulted in a total of 296 studies included in the current meta-analysis (see **Figure 2**). Studies were considered relevant and selected to be included at the next screening stage based on the following criteria.

First, studies were only included if 75% or more of the sample consisted of children and/or adolescents. Based on guidelines from the World Health Organization (WHO; https://www.who.int/health-topics/adolescent-health#tab=tab_1), we defined the period from childhood to end of adolescence as ranging from age 4, the earliest age for compulsory schooling, to age 24, the end of adolescence. Second, we included studies that had measured NDDs and DICCs considering either formal clinical diagnoses, clinical cut-offs, and/or quantitative measures of symptoms. Third, studies were selected only if they featured data on at least one NDD (Aim 1), at least two NDDs (Aim 2), or at least one NDD and one DICC disorder (Aim 3).

Fourth, studies using family-based designs had to have reported at least one estimate of heritability (h^2^), shared environmental (c^2^) or nonshared environmental influence (e^2^), or genetic or environmental correlations. We included only single-generation family designs, that is studies that had used twin design^84^, sibling comparisons^85^, or extended twin designs^86^. We excluded multiple-generation family designs (e.g., children-of-twins^87^ and in-vitro fertilization^88^) due to the potential confounding in the genetic and environmental estimates that could have resulted from including parental traits in the models decomposing the covariance between family members^89^.

Fifth, studies using genomic designs were included only if they had reported at least one SNP-based heritability estimate and/or a genetic correlation (r_a_). Eligible SNP-based methods to quantify the proportion of phenotypic variance accounted for by common SNPs included genome-based restricted maximum likelihood (GREML)^16^, linkage-disequilibrium score regression (LDSC)^18^ and SbayesS, which is a Bayesian approach to analysis of GWA summary data^90^. Each method is described in greater detail in **Supplementary Note 6**. Sixth, studies that had selected participants based on other diagnoses not related to NDD or DICC categories or based extreme vulnerability environmental insult unrelated to NDDs or DICCs, such as alcohol abuse, were not included. Lastly, only studies published in English were included. Studies deemed eligible based on full text scanning were also scored in terms of their scientific quality by two reviewers (see details on the quality scoring checklist in **Supplementary Note 7**).

### Data extraction

Data extraction was conducted by the primary reviewer. Issues and uncertainties were resolved through discussion with co-authors. Missing data was requested from study authors via email or ResearchGate (for details, see **Supplementary Note 8**). Extracted data were compiled in a table, including information on study reference, project/cohort name, study design (e.g., classical twin study), model reported (e.g., full ACE model; when multiple models were reported, the best fitting model was selected for data synthesis), overall number of participants and number of participants in subgroups (e.g. number of monozygotic vs. dizygotic twins), average age and age range of the sample, cohort country(ies) of origin, participants ancestry (defined in terms on the percentage of participants of European ancestry in samples), broad types of NDD and DICC included (e.g., Specific Learning Disorder), sub-type of NDD and/or DICC included (e.g., dyslexia), specific phenotypes measured (e.g., reading fluency), measure statistics (e.g., binary (diagnosis) or continuous (symptoms continua)), measure (e.g., Conners rating scale for ADHD) and rater (e.g., parent reports), covariates included in the analyses (e.g., age and sex), statistics (e.g., family-based heritability, SNP-based genetic correlation etc.), and finally the estimated statistics and the provided index of measurement variance (e.g., standard error). For studies where neither standard deviation, standard errors nor 95% confidence intervals were reported, the 95% confidence intervals were calculated using the Cir function implemented in the R package *psychometric*^91,92^, based on the sample size of the study, and subsequently converted to standard errors via dividing the difference between upper and lower bound confidence intervals by 3.92^93^.

### Data synthesis

Heritability and environmental influences reported by selected studies were synthesised using a multilevel random-effects meta-analysis in metafor for R^49,92^. We used heritability/environmental influences and genetic/environmental correlation coefficients, along with standard errors as the measures of effect size^25^. However, to avoid the risk of Type I error introduced by the distribution characteristics of the correlation coefficient^94^, we transformed all estimates using Fisher’s z. Effect sizes were then weighted by their inverse variance weight so that larger samples were given more weighting and the standard error for the common effect size resulted as a function of the allocated weights. For results presentation, Fisher’s z was transformed back to variance components and correlation coefficients^95^. Multilevel random-effects models enabled varying true effect sizes across studies^25,96^. We introduced a 2-level structure to account for nested effects underlying heterogeneity and clustering across studies (Level 1: individual clustering; Level 2: cohort clustering). Given that males show an up to four times higher risk of NDDs^52,53^, we meta-analysed studies that provided sex-specific estimates in separate models to minimize sample heterogeneity across studies and report separate grand estimates for combined, male-only, and female-only samples.

### Aggregation of non-independent effects

Multilevel meta-analytic models allow to account for non-independence of estimates derived from partly or completely overlapping samples (i.e., estimates obtained from multiple studies that have used the same cohort of participants). To further account for the non-independence of sampling variance (i.e., when sampling errors correlate because data from partly the same individuals is used to estimate multiple effect sizes), we also aggregated multiple estimates within each individual study (e.g., estimates at multiple timepoints derived from the same study). Aggregation of dependent effects sizes was performed at the level of each study using the R package Meta-Analysis with Mean Differences^97,92^ (MAd), applying a default correlation between estimates of 0.5. We conducted several sensitivity analyses, comparing different aggregation methods, i.e., aggregating at the level of the study, cohort, and country, and varying the assumed correlation between dependent effect sizes (0.5, 0.3 and 0.9). Results of these additional checks are presented in **Supplementary Figure 24** and discussed in **Supplementary Note 9**. Since differences in aggregation strategy did not result in significant differences in meta-analytic effects, we report results obtained when the correlation between dependent effect sizes was set to 0.5.

### Bias and heterogeneity assessment

The potential for publication bias was explored using funnel plots and Egger’s linear regression^98^. The proportion of heterogeneity across estimates was estimated using the I^2^ statistics, which calculates the fraction of variance across studies that can be attributed to heterogeneity, rather than chance^99,100^. The I^2^ statistics was computed as a proportion of true variance of true effects to variance of the observed effects, in line with the following formula^101^:

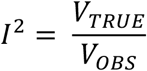

where V_TRUE_ is the variation of true effects and V_OBS_ is the variation due to sampling error. In other words, I^2^ can be interpreted as the dispersion of observed effects as compared to the dispersion that would be predicted just from sampling error^101^. The I^2^ statistics also provides insight into the degree to which confidence intervals from individual studies are independent^25,101^.

### Moderation analyses

We tested for the effect of several moderators. Selection of moderator terms was determined based on available data, considering completeness of reported moderator variables. We implemented a >50% rule of thumb, i.e., if 50% or more studies reported data on the moderating variable, we included this moderator in our analyses. For example, less than 50% of studies reported the percentage of participants of Asian ancestry in the sample, hence we did not include the percentage of Asian participants in moderation analyses. We considered the following 11 moderators: age group, design, type of model, rater, measurement, percentage of individuals who identified as White, number of covariates included in the analysis, measure adopted, country, and specific phenotype measured, each moderator is described in greater detail in **Supplementary Note 3**. Moderation analyses were conducted using a two-step procedure. First, only studies that reported data on the level of the moderator were selected (for example, only studies reporting estimates for adolescents). Second, analyses stratified by levels of the moderator were run using a multilevel random-effects meta-analysis in metafor for R^49,92^, i.e., a grand estimate was derived for adolescents and subsequently compared with estimates for other developmental stages (i.e., childhood and middle childhood) using the same procedure. We report unstratified estimates (**Supplementary Tables 1, 3 & 5**) and estimates stratified by specific phenotype measured (**Supplementary Tables 2, 4 & 6**), age category (**Supplementary Tables 16-18**), country (**Supplementary Tables 19-21**), and ancestry (**Supplementary Tables 22-24**) in the main text, whereas estimates stratified by all other moderators are reported in **Supplementary tables 34-47**.

## Data Availability

The code for all analyses is available at https://github.com/CoDEresearchlab/Meta_analysis_NDDs_DICCs.

## Contributions

Conceived and designed the study: AG, MM, YIA. Conducted literature search and study screening: AG, LYL., MD, FP and ED. Extracted and analysed the data: AG. Wrote the paper: AG., MM with helpful contributions from YIA, GM, AGA, JAB, FP, AR, and KR. All authors contributed to the interpretation of data, provided critical feedback on manuscript drafts, and approved the final draft.

## Acknowledgements

This meta-analysis was funded by a starting grant awarded to MM by the School of Biological and Behavioural Sciences at Queen Mary, University of London. AG is supported by a Queen Mary School of Biological and Behavioural Sciences PhD Fellowship awarded to MM.

GM was in receipt of a Klingenstein Third Generation Foundation fellowship (grant 20212999).

FP is supported by a National Institutes of Health [NIH] Subaward via The Regents of the University of California, Riverside [AG046938].

KR is supported by a Sir Henry Wellcome Postdoctoral Fellowship.

## References

1. Thapar, A. & Rutter, M. Neurodevelopmental disorders. Rutters Child Adolesc. Psychiatry 31–40 (2015).

2. Dietrich, K. N. et al. Principles and practices of neurodevelopmental assessment in children: lessons learned from the Centers for Children’s Environmental Health and Disease Prevention Research. Environ. Health Perspect. 113, 1437–1446 (2005).

3. Association, A. P. Neurodevelopmental disorders: DSM-5® selections. (American Psychiatric Pub, 2015).

4. Hyman, S. L., Levy, S. E. & Myers, S. M. Identification, evaluation, and management of children with autism spectrum disorder. Pediatrics 145, (2020).

5. Gillberg, C. The ESSENCE in child psychiatry: early symptomatic syndromes eliciting neurodevelopmental clinical examinations. Res. Dev. Disabil. 31, 1543–1551 (2010).

6. McGovern, C. W. & Sigman, M. Continuity and change from early childhood to adolescence in autism. J. Child Psychol. Psychiatry 46, 401–408 (2005).

7. Faraone, S. V., Biederman, J. & Mick, E. The age-dependent decline of attention deficit hyperactivity disorder: a meta-analysis of follow-up studies. Psychol. Med. 36, 159–165 (2006).

8. Kim, S. et al. Provisional Tic Disorder is not so transient. Sci. Rep. 9, 1–11 (2019).

9. Ellis, E. M. & Thal, D. J. Early language delay and risk for language impairment. Perspect. Lang. Learn. Educ. 15, 93–100 (2008).

10. Bishop, D. V. Which neurodevelopmental disorders get researched and why? PloS One 5, e15112 (2010).

11. Tick, B., Bolton, P., Happé, F., Rutter, M. & Rijsdijk, F. Heritability of autism spectrum disorders: a meta-analysis of twin studies. J. Child Psychol. Psychiatry 57, 585–595 (2016).

12. Burt, S. A. Rethinking environmental contributions to child and adolescent psychopathology: a meta-analysis of shared environmental influences. Psychol. Bull. 135, 608 (2009).

13. Faraone, S. V. & Larsson, H. Genetics of attention deficit hyperactivity disorder. Mol. Psychiatry 24, 562–575 (2019).

14. Nikolas, M. A. & Burt, S. A. Genetic and environmental influences on ADHD symptom dimensions of inattention and hyperactivity: a meta-analysis. J. Abnorm. Psychol. 119, 1 (2010).

15. Cheesman, R. et al. Childhood behaviour problems show the greatest gap between DNA-based and twin heritability. Transl. Psychiatry 7, 1–9 (2017).

16. Yang, J., Zeng, J., Goddard, M. E., Wray, N. R. & Visscher, P. M. Concepts, estimation and interpretation of SNP-based heritability. Nat. Genet. 49, 1304–1310 (2017).

17. Yang, J., Lee, S. H., Goddard, M. E. & Visscher, P. M. GCTA: a tool for genome-wide complex trait analysis. Am. J. Hum. Genet. 88, 76–82 (2011).

18. Bulik-Sullivan, B. K. et al. LD Score regression distinguishes confounding from polygenicity in genome-wide association studies. Nat. Genet. 47, 291–295 (2015).

19. Grove, J. et al. Identification of common genetic risk variants for autism spectrum disorder. Nat. Genet. 51, 431–444 (2019).

20. Demontis, D. et al. Discovery of the first genome-wide significant risk loci for attention deficit/hyperactivity disorder. Nat. Genet. 51, 63–75 (2019).

21. Pettersson, E., Anckarsäter, H., Gillberg, C. & Lichtenstein, P. Different neurodevelopmental symptoms have a common genetic etiology. J. Child Psychol. Psychiatry 54, 1356–1365 (2013).

22. Brimo, K. et al. The co-occurrence of neurodevelopmental problems in dyslexia. Dyslexia (2021).

23. Knopik, V. S., Neiderhiser, J. M., DeFries, J. C. & Plomin, R. Behavioral genetics. (Worth Publishers, Macmillan Learning, 2017).

24. Ronald, A., Simonoff, E., Kuntsi, J., Asherson, P. & Plomin, R. Evidence for overlapping genetic influences on autistic and ADHD behaviours in a community twin sample. J. Child Psychol. Psychiatry 49, 535–542 (2008).

25. Andersson, A. et al. Research Review: The strength of the genetic overlap between ADHD and other psychiatric symptoms–a systematic review and meta-analysis. J. Child Psychol. Psychiatry 61, 1173– 1183 (2020).

26. Yang, Z. et al. Cross-disorder GWAS meta-analysis for attention deficit/hyperactivity disorder, autism spectrum disorder, obsessive compulsive disorder, and Tourette syndrome. bioRxiv 770222 (2019).

27. Lichtenstein, P., Carlström, E., Råstam, M., Gillberg, C. & Anckarsäter, H. The genetics of autism spectrum disorders and related neuropsychiatric disorders in childhood. Am. J. Psychiatry 167, 1357– 1363 (2010).

28. Paloyelis, Y., Rijsdijk, F., Wood, A. C., Asherson, P. & Kuntsi, J. The genetic association between ADHD symptoms and reading difficulties: the role of inattentiveness and IQ. J. Abnorm. Child Psychol. 38, 1083–1095 (2010).

29. Pitcher, T. M., Piek, J. P. & Hay, D. A. Fine and gross motor ability in males with ADHD. Dev. Med. Child Neurol. 45, 525–535 (2003).

30. Licari, M. K. et al. Prevalence of motor difficulties in autism spectrum disorder: analysis of a population-based cohort. Autism Res. 13, 298–306 (2020).

31. Martin, N. C., Piek, J. P. & Hay, D. DCD and ADHD: a genetic study of their shared aetiology. Hum. Mov. Sci. 25, 110–124 (2006).

32. Taylor, M. J. et al. Language and traits of autism spectrum conditions: Evidence of limited phenotypic and etiological overlap. Am. J. Med. Genet. B Neuropsychiatr. Genet. 165, 587–595 (2014).

33. Dworzynski, K. et al. Developmental path between language and autistic-like impairments: A twin study. Infant Child Dev. Int. J. Res. Pract. 17, 121–136 (2008).

34. American Psychiatric Association, A. Diagnostic and statistical manual of mental disorders. vol. 3 (American Psychiatric Association Washington, DC, 1980).

35. Association, A. P. Diagnostic criteria from dsM-iV-tr. (2000).

36. Thorell, L. B. & Wåhlstedt, C. Executive functioning deficits in relation to symptoms of ADHD and/or ODD in preschool children. Infant Child Dev. 15, 503–518 (2006).

37. Bayard, F. et al. Distinct brain structure and behavior related to ADHD and conduct disorder traits. Mol. Psychiatry 25, 3020–3033 (2020).

38. Martin, N. C., Levy, F., Pieka, J. & Hay, D. A. A genetic study of attention deficit hyperactivity disorder, conduct disorder, oppositional defiant disorder and reading disability: Aetiological overlaps and implications. Int. J. Disabil. Dev. Educ. 53, 21–34 (2006).

39. Moffitt, T. E. The neuropsychology of conduct disorder. Dev. Psychopathol. 5, 135–151 (1993).

40. Rubia, K. “Cool” inferior frontostriatal dysfunction in attention-deficit/hyperactivity disorder versus “hot” ventromedial orbitofrontal-limbic dysfunction in conduct disorder: a review. Biol. Psychiatry 69, e69–e87 (2011).

41. Bronsard, G., Botbol, M. & Tordjman, S. Aggression in low functioning children and adolescents with autistic disorder. PloS One 5, e14358 (2010).

42. Moffitt, T. E., Caspi, A., Rutter, M. & Silva, P. A. Sex differences in antisocial behaviour: Conduct disorder, delinquency, and violence in the Dunedin Longitudinal Study. xvii, 278 (Cambridge University Press, 2001). doi:10.1017/CBO9780511490057.

43. Jones, A. P. et al. Phenotypic and aetiological associations between psychopathic tendencies, autistic traits, and emotion attribution. Crim. Justice Behav. 36, 1198–1212 (2009).

44. Rogers, J., Viding, E., Blair, R. J., Frith, U. & Happe, F. Autism spectrum disorder and psychopathy: shared cognitive underpinnings or double hit? Psychol. Med. 36, 1789–1798 (2006).

45. O’Nions, E. et al. Examining the genetic and environmental associations between autistic social and communication deficits and psychopathic callous-unemotional traits. PloS One 10, e0134331 (2015).

46. Skuse, D. H. Rethinking the nature of genetic vulnerability to autistic spectrum disorders. TRENDS Genet. 23, 387–395 (2007).

47. DesChamps, T. D., Ibañez, L. V., Edmunds, S. R., Dick, C. C. & Stone, W. L. Parenting stress in caregivers of young children with ASD concerns prior to a formal diagnosis. Autism Res. 13, 82–92 (2020).

48. Baselmans, B. M., Yengo, L.,van Rheenen, W. & Wray, N. R. Risk in relatives, heritability, SNP-based heritability, and genetic correlations in psychiatric disorders: A review. Biol. Psychiatry 89, 11–19 (2021).

49. Viechtbauer, W. & Viechtbauer, M. W. Package ‘metafor’. Compr. R Arch. Netw. Package ‘metafor’ Httpcran R-Proj. Orgwebpackagesmetaformetafor Pdf (2015).

50. Davis, O. S. et al. The correlation between reading and mathematics ability at age twelve has a substantial genetic component. Nat. Commun. 5, 1–6 (2014).

51. Newsome, J., Boisvert, D. & Wright, J. P. Genetic and environmental influences on the co-occurrence of early academic achievement and externalizing behavior. J. Crim. Justice 42, 45–53 (2014).

52. Christensen, D. L. et al. Prevalence and characteristics of autism spectrum disorder among children aged 8 years—autism and developmental disabilities monitoring network, 11 sites, United States, 2012. MMWR Surveill. Summ. 65, 1 (2018).

53. May, T., Sciberras, E., Brignell, A. & Williams, K. Autism spectrum disorder: updated prevalence and comparison of two birth cohorts in a nationally representative Australian sample. BMJ Open 7, e015549 (2017).

54. Martin, J. et al. A genetic investigation of sex bias in the prevalence of attention-deficit/hyperactivity disorder. Biol. Psychiatry 83, 1044–1053 (2018).

55. Metrics, I. for H. & Evaluation. Global Burden of Disease Collaborative Network. Global Burden of Disease Study 2016 (GBD 2016) Results. (2017).

56. Genetics for all. Nat. Genet. 51, 579–579 (2019).

57. Whose genomics? Nat. Hum. Behav. 3, 409–410 (2019).

58. Silventoinen, K. et al. Genetic and environmental variation in educational attainment: an individual-based analysis of 28 twin cohorts. Sci. Rep. 10, 12681 (2020).

59. Rimfeld, K. et al. Genetic influence on social outcomes during and after the Soviet era in Estonia. Nat. Hum. Behav. 2, 269–275 (2018).

60. Abdellaoui, A. et al. Genetic correlates of social stratification in Great Britain. Nat. Hum. Behav. 3, 1332–1342 (2019).

61. Belsky, D. W. et al. Genetics and the geography of health, behaviour and attainment. Nat. Hum. Behav. 3, 576–586 (2019).

62. Popejoy, A. B. & Fullerton, S. M. Genomics is failing on diversity. Nature 538, 161–164 (2016).

63. Martin, A. R. et al. Clinical use of current polygenic risk scores may exacerbate health disparities. Nat. Genet. 51, 584–591 (2019).

64. Finer, S. et al. Cohort Profile: East London Genes & Health (ELGH), a community-based population genomics and health study in British Bangladeshi and British Pakistani people. Int. J. Epidemiol. 49, 20– 21i (2020).

65. Wright, J. et al. Cohort profile: the Born in Bradford multi-ethnic family cohort study. Int. J. Epidemiol. 42, 978–991 (2013).

66. Ramsay, M., Sankoh, O., study, as members of the A.-G. & Consortium, the H. African partnerships through the H3Africa Consortium bring a genomic dimension to longitudinal population studies on the continent. International Journal of Epidemiology vol. 45 305–308 (2016).

67. Manolio, T. A. et al. Finding the missing heritability of complex diseases. Nat Reviews vol. 461 747– 753 (2009).

68. Aschard, H. et al. Inclusion of gene-gene and gene-environment interactions unlikely to dramatically improve risk prediction for complex diseases. Am. J. Hum. Genet. 90, 962–972 (2012).

69. Wainschtein, P. & et al. Recovery of trait heritability from whole genome sequence data Visscher 2019.pdf. bioRxiv 1–23 (2019).

70. Young, A. I. Solving the missing heritability problem. PLOS Genet. 15, e1008222 (2019).

71. Estimates of the population for the UK, England and Wales, Scotland and Northern Ireland -Office for National Statistics. https://www.ons.gov.uk/peoplepopulationandcommunity/populationandmigration/populationestimates/datasets/populationestimatesforukenglandandwalesscotlandandnorthernireland.

72. Hatton, C., Glover, G., Emerson, E. & Brown, I. People with learning disabilities in England 2015: Main report. Lond. Public Health Engl. Intellect. Disabil. 30, 1007–1021 (2016).

73. Mehlmann-Wicks, J. Autism spectrum disorder. The British Medical Association is the trade union and professional body for doctors in the UK. https://www.bma.org.uk/what-we-do/population-health/improving-the-health-of-specific-groups/autism-spectrum-disorder.

74. AGRE - Autism Genetic Resource Exchange. Autism Speaks https://www.autismspeaks.org/agre.

75. About iPSYCH. https://ipsych.dk/en/about-ipsych/.

76. Psychiatric Genomics Consortium. Psychiatric Genomics Consortium https://www.med.unc.edu/pgc/.

77. Zwicker, J. G., Missiuna, C., Harris, S. R. & Boyd, L. A. Developmental coordination disorder: a review and update. Eur. J. Paediatr. Neurol. 16, 573–581 (2012).

78. Valentine, A. Z. et al. A systematic review evaluating the implementation of technologies to assess, monitor and treat neurodevelopmental disorders: A map of the current evidence. Clin. Psychol. Rev. 80, 101870 (2020).

79. Valentine, A. Z. et al. Implementation of telehealth services to assess, monitor, and treat neurodevelopmental disorders: systematic review. J. Med. Internet Res. 23, e22619 (2021).

80. McGregor, K. K. How we fail children with developmental language disorder. Lang. Speech Hear. Serv. Sch. 51, 981–992 (2020).

81. Khan, K., Hall, C. L., Davies, E. B., Hollis, C. & Glazebrook, C. The effectiveness of web-based interventions delivered to children and young people with neurodevelopmental disorders: systematic review and meta-analysis. J. Med. Internet Res. 21, e13478 (2019).

82. Bigby, C. Social inclusion and people with intellectual disability and challenging behaviour: A systematic review. J. Intellect. Dev. Disabil. 37, 360–374 (2012).

83. Moher, D., Liberati, A., Tetzlaff, J., Altman, D. G. & Group, P. Preferred reporting items for systematic reviews and meta-analyses: the PRISMA statement. PLoS Med. 6, e1000097 (2009).

84. Rijsdijk, F. V. & Sham, P. C. Analytic approaches to twin data using structural equation models. Brief. Bioinform. 3, 119–133 (2002).

85. Kendler, K. S. et al. A novel sibling-based design to quantify genetic and shared environmental effects: application to drug abuse, alcohol use disorder and criminal behavior. Psychol. Med. 46, 1639–1650 (2016).

86. Posthuma, D. & Boomsma, D. I. A note on the statistical power in extended twin designs. Behav. Genet. 30, 147–158 (2000).

87. Eley, T. C. et al. The intergenerational transmission of anxiety: a children-of-twins study. Am. J. Psychiatry 172, 630–637 (2015).

88. Rice, F. et al. Disentangling prenatal and inherited influences in humans with an experimental design. Proc. Natl. Acad. Sci. 106, 2464–2467 (2009).

89. McAdams, T. A. et al. Revisiting the children-of-twins design: improving existing models for the exploration of intergenerational associations. Behav. Genet. 48, 397–412 (2018).

90. Zeng, J. et al. Signatures of negative selection in the genetic architecture of human complex traits. Nat. Genet. 50, 746–753 (2018).

91. Fletcher, T. D. & Fletcher, M. T. D. Package ‘psychometric’. Recuperado Httpcran Rproject Orgwebpackagespsychometricpsychometric Pdfel 4, (2013).

92. Team, R. C. 2020. R Lang. Environ. Stat. Comput. R Found. Stat. Comput. Vienna Austria Available Httpswww.R-Proj.OrgGoogleSch. (2019).

93. Cohen, J., Cohen, P., West, S. G. & Aiken, L. S. Applied multiple regression/correlation analysis for the behavioral sciences (Third; Routledge, Ed.). (2013).

94. Alexander, R. A., Scozzaro, M. J. & Borodkin, L. J. Statistical and empirical examination of the chi-square test for homogeneity of correlations in meta-analysis. Psychol. Bull. 106, 329 (1989).

95. Malanchini, M. et al. Aggressive behaviour in childhood and adolescence: the role of smoking during pregnancy, evidence from four twin cohorts in the EU-ACTION consortium. Psychol. Med. 49, 646–654 (2019).

96. Field, A. P. & Gillett, R. How to do a meta-analysis. Br. J. Math. Stat. Psychol. 63, 665–694 (2010).

97. Del Re, A. C., Hoyt, W. T. & Del Re, M. A. Package ‘MAd’. (2014).

98. Sterne, J. A. & Egger, M. Regression methods to detect publication and other bias in meta-analysis. Publ. Bias Meta-Anal. Prev. Assess. Adjust. 99, 110 (2005).

99. Higgins, J. P. & Thompson, S. G. Quantifying heterogeneity in a meta-analysis. Stat. Med. 21, 1539– 1558 (2002).

100. Higgins, J. P., Thompson, S. G., Deeks, J. J. & Altman, D. G. Measuring inconsistency in meta-analyses. Bmj 327, 557–560 (2003).

101. Borenstein, M., Higgins, J. P., Hedges, L. V. & Rothstein, H. R. Basics of meta-analysis: I2 is not an absolute measure of heterogeneity. Res. Synth. Methods 8, 5–18 (2017).

