## Supplementary material for "Genetic influences on neurodevelopmental disorders and their overlap with co-occurring conditions in childhood and adolescence: A meta-analysis": SI_META_NatGen_160222.docx

| Table of contents | |
| --- | --- |
| Supplementary Notes | Pages 2-21 |
| Supplementary Tables | Pages 22-110 |
| Supplementary Figures | Pages 111-136 |
| Supplementary References | Page 137 |

### Supplementary Notes

| Supplementary Notes: table of contents | |
| --- | --- |
| Supplementary Note 1 | Pages 3-7 |
| Supplementary Note 2 | Pages 8-9 |
| Supplementary Note 3 | Pages 10-11 |
| Supplementary Note 4 | Page 12 |
| Supplementary Note 5 | Pages 13-14 |
| Supplementary Note 6 | Page 15 |
| Supplementary Note 7 | Page 16 |
| Supplementary Note 8 | Page 17 |
| Supplementary Note 9 | Page 18 |
| Supplementary Note 10 | Pages 19-20 |
| Supplementary Note 11 | Page 21 |

**Supplementary Note 1**: Meta-analytic results for shared and nonshared environmental factors.

**Supplementary Note 2**: Meta-analytic results for NDDs phenotypic sub-categories.

**Supplementary Note 3**: Description of moderators.

**Supplementary Note 4**: Categorical versus continuous measurement of NDDs

**Supplementary Note 5**: Indexes, timespans, search strategy and key words.

**Supplementary Note 6**: Description of SNP-based methods targeted by the meta-analysis.

**Supplementary Note 7**: Quality scoring checklist.

**Supplementary Note 8**: Requesting missing data from study authors.

**Supplementary Note 9**: Aggregation sensitivity analyses.

**Supplementary Note 10**: Heterogeneity assessment.

**Supplementary Note 11**: Publication bias.

### Supplementary Note 1: Meta-analytic results for shared and nonshared environmental factors.

#### Aim 1: Environmental contributions to variation in NDDs are modest to moderate.

##### Shared environment (c^2^)

We identified 127 studies that reported information on shared environmental influences on NDDs, only a little over half (53.6%) of all studies that reported on h^2^ also reported on c^2^. Out of the total 127 studies, 65 studies focused on specific learning disorders, 48 on ADHD, 15 on communication disorders, 14 on ASD, 3 on motor disorders, and 0 studies included c^2^ *estimates* for intellectual disabilities, the only two studies that had examined the aetiology of intellectual disabilities had reported a model only including genetic and nonshared environmental factors (AE) as the best fitting model (see ***Methods*** and **Supplementary Note 3**). The contribution of shared environmental influences to all NDD categories was modest (c^2^ = 0.17, SE= 0.02), ranging from weak (c^2^ = 0.10, SE= 0.02) for ADHD to moderate (c^2^ = 0.36, SE= 0.06) for communication disorders (**Figure 3** in the main text and **Supplementary Table 1**).

##### Nonshared environment (e^2^)

We identified 195 family-based studies (82.2% of the total) that reported on the nonshared environmental contribution to NDDs, out of which 107 studies focused on ADHD, 67 on specific learning disorders, 28 on ASD, 18 on communication disorders, 6 on motor disorders and 2 studies on intellectual disabilities. Nonshared environmental influences on all NDDs were moderate (e^2^ = 0.29, SE= 0.02), but ranged from weak (e^2^ = 0.10, SE= 0.16) for intellectual disabilities to moderate (e^2^ = 0.38, SE= 0.11) for motor disorders. Nonshared environmental estimates did not differ significantly across all NDDs (**Figure 3** in the main text and **Supplementary Table 1**).

#### Aim 2: Shared environmental factors underlie the widespread co-occurrence between NDDs during childhood and adolescence

##### Shared environmental correlations (rC)

Since several studies only reported the most parsimonious, best-fitting, model (see **Supplementary Note 3**), meta-analytic estimates of rC could be derived from 16 studies (43.2% of the total number; **Supplementary Table 3**). A first meta-analysis of all NDD categories jointly, yielded a significant and substantial grand estimate for the shared environmental co-occurrence between different NDDs *(*rC= 0.63, SE= 0.32), although estimates varied substantially between studies, as indicated by the large meta-analytic standard error.

##### Nonshared environmental correlations (rE)

A total of 22 studies (59.5%) reported on the nonshared environmental co-occurrence between NDDs, this was largely due to the fact that different studies adopted different family-based designs, some of which do not provide nonshared environmental estimates^1^ (see **Supplementary** **Note 3).** The grand estimate for the transdiagnostic rE was 0.17, SE= 0.5. When we considered NDD categories separately, nonshared environmental correlations could only be estimated between ASD & ADHD (5 studies, rE = 0.22, SE= 0.13), and between ADHD & specific learning disorders (9 studies, rE = 0.11, SE= 0.05; **Figure 4** in the main text and **Supplementary Table 3**)

#### Aim 3: NDDs share as much of their environmental underpinnings with their homotypic (other NDDs) and heterotypic (DICCs) co-occurrences

##### Shared environmental correlations (rC)

Out of 15 studies that reported genetic correlations between NDDs and DICCs, 11 also reported shared environmental correlations (73.3%). These included 4 studies looking at the co-occurrence between ADHD & oppositional defiant disorder, 3 studies looking at the co-occurrence between ADHD & conduct disorder, and 3 studies looking at the co-occurrence between ASD & conduct disorder. A strong meta-analytic shared environmental correlation was found between all NDDs and DICCs (0.88, SE= 0.34). The grand shared environmental overlap was consistently estimated as very high for all co-occurring disorders for which we identified sufficient studies: rC= 0.96 (SE= 0.57) between ADHD & oppositional defiant disorder, rC= 0.94 (SE= 0.71) between ADHD & conduct disorder, and rC= 0.88 (SE= 0.57) between ASD & conduct disorder (**Figure 4** in the main text and **Supplementary Table 5**).

##### Nonshared environmental correlations (rE)

Thirteen out of 15 studies that reported on the genetic overlap between NDDs and DICCs also reported nonshared environmental correlations (86.7%). These 13 studies consisted of 5 studies targeting the co-occurrence between ADHD & conduct disorder, 5 studies that between ADHD & oppositional defiant disorder, and 3 studies the co-occurrence between ASD & conduct disorder. The nonshared environmental overlap across all NDD and DICC pairs was moderate (rE = 0.39, SE= 0.14), but differed between specific pairs of disorders. The strongest correlation (rE = 0.54, SE= 0.25) was found between ADHD & oppositional defiant disorder and was markedly higher if compared to the overlap between ADHD & conduct disorder (rE = 0.11, SE= 0.08) and between ASD & conduct disorder (0.07, SE= 0.08) (**Figure 4** in the main text and **Supplementary Table 4**).

#### Sex differences: While the aetiology of NDDs is comparable for males and females, their co-occurrences differ by sex

##### Sex differences in environmental aetiology of NDDs

Across all NDDs, family-based shared and nonshared environmental influences were not significantly different between males (c^2^= 0.35, SE= 0.09; e^2^= 0.31, SE= 0.05) and females (c^2^= 0.28, SE= 0.08; e^2^= 0.33, SE= 0.04). Distributions of sex-specific family-based variance components for all NDDs, except for motor disorders for which a sufficient number of studies (>1) was not identified, are presented in **Figure 5** in the main text and **Supplementary Table 13)**

##### Sex differences in environmental overlap between NDDs

Sex-specific shared environmental correlations could not be estimated, whereas nonshared environmental correlations were estimated at 0.09 (SE= 0.08) in males and 0.10 (SE= 0.11) in females (**Supplementary Table 14)**. Sex-specific grand estimates of environmental correlations between specific disorders are not reported because of the limited number of studies identified. The only exception was the co-occurrence between ASD & ADHD in males, where 2 studies were identified (rE = 0.20, SE= 0.14; **Supplementary Table 14)**. Due to the lack of available studies, the shared environmental overlap could not be calculated.

##### Sex differences in environmental overlap between NDDs and DICCs

We could only meta-analyse the co-occurrence between ADHD & conduct disorder in females. We found a meta-analytic nonshared environmental correlation of 0.06 (SE= 0.12; **Supplementary Table 15)**.

#### Developmental trends: Environmental influences on NDDs are stable from childhood to adolescence

##### Age-related differences in the environmental aetiology of NDDs

Across all NDDs, grand shared and nonshared environmental influences were observed to decrease from childhood (c^2^= 0.21, SE= 0.04; e^2^= 0.27, SE= 0.03) to middle childhood (c^2^= 0.12, SE= 0.03; e^2^= 0.25, SE= 0.02) followed by a later increase in adolescence (c^2^= 0.17, SE= 0.03; e^2^= 0.36, SE= 0.03). This trend was consistent across some specific NDDs, such as ASD and ADHD, but not for others. For example, for communication disorders and specific learning disorders genetic and shared environmental variance decreased while nonshared environmental variance increased developmentally (**Figure 6A** in the main text and **Supplementary Table 16**).

##### Age-related differences in environmental overlap between NDDs, as well as between NDDs and DICCs

Overall, we could not explore developmental trends in genetic and environmental correlations due to a lack of available studies, the only exceptions were grand estimates for adolescence (see **Supplementary Tables 17-18)**.

#### Categorical versus continuous measurement of NDDs

We found no significant differences in shared and nonshared environmental influences between measurement methods (**Supplementary Figure 22** and **Supplementary Table 25**). Furthermore, shared and nonshared environmental genetic overlap could not be compared across co-occurrences between NDDs, and between NDDs and DICCs, due to insufficient number of identified studies (**Supplementary Figure 22** and **Supplementary Tables 26 and 27**).

#### NDDs and co-occurring disorders are almost exclusively researched in Western countries

##### Geographical differences in environmental aetiology of NDDs

Grand shared environmental influences ranged between 0.30 (SE= 0.13) in Chinese cohorts and 0.07 (SE= 0.04) in Swedish cohorts (**Figure 7A** in the main text and **Supplementary Table 19**), whereas nonshared environmental influences were highest in Canada (0.38, SE= 0.07), if compared to the lowest grand estimate of nonshared environmental influence (0.17, SE= 0.05) obtained for Australian cohorts (**Figure 7A** in the main text and **Supplementary Table 19**).

##### Geographical differences in environmental overlap between NDDs

The highest meta-analytic estimate of shared environmental correlation was estimated in United Kingdom-based samples (0.91, SE= 0.29), while the lowest in United States-based studies (0.07, SE= 0.21; **Figure 7B** in the main text and **Supplementary Table 20**). The strongest grand estimate of nonshared environmental correlation was found in Swedish samples (0.36, SE= 0.12) while the lowest in Australian samples (0.03, SE= 0.09; **Figure 7B** in the main text and **Supplementary Table 20**).

##### Geographical differences in environmental overlap between NDDs and DICCs

Studies yielded consistently strong estimates of shared environmental correlation across the United Kingdom, United Stated and Sweden (0.97, SE= 0.57; 0.85, SE= 0.56; and 0.89, SE= 0.55; **Figure 7C** in the main text and **Supplementary Table 21**). Grand nonshared environmental correlations could only be calculated for United Kingdom and United States-based studies and were estimated at 0.49 (SE= 0.44) and 0.24 (SE= 0.09), respectively (**Figure 7C** in the main text and **Supplementary Table 21**).

#### NDDs and co-occurring disorders are mostly researched in cohorts of European ancestry

##### Ancestry-related differences in the environmental aetiology of NDDs

Meta-analytic shared environmental influences remained relatively stable across sample ancestral composition (mean of c^2^= 0.24) with only a slight drop observed when the sample included 100% of participants of European ancestry (c^2^= 0.19, SE= 0.04; **Figure 8** in the main text and **Supplementary Table 22**). However, estimates differed for specific disorders. The decrease in shared environmental influences in fully European descent samples was especially evident for ADHD, where the estimates dropped from a mean of 0.17 for more diverse categories to 0.04 (SE= 0.09) for 100% European ancestry samples. A similar pattern was observed for specific learning disorders, with estimates dropping from a mean of 0.26 to 0.16 (SE= 0.04) (**Figure 8** in the main text and **Supplementary Table 22)**.

All NDDs were subject to subtle changes in nonshared environmental influences depending on the ancestral composition of the samples, with the exception of motor disorders for which only studies using 100% European ancestry samples were found. Across all NDDs, the meta-analytic estimate for nonshared environmental influences decreased as the percentage of participants of European ancestry in the sample increased: from 0.44 (SE= 0.08) for samples where participants of European ancestry were in the minority, to 0.32 (SE= 0.13) for samples where they were between 50 and 74% to 0.25 (SE =0.03) for samples between 75 and 99% European ancestry) to 0.32 (SE= 0.05) for 100% European ancestry samples. This same trend was observed for ADHD (from 0.54, SE= 0.09 to 0.39, SE= 0.06) and specific learning disorders (0.28, SE= 0.06 to 0.19, SE= 0.06, although the estimate increased again for samples 100% of European descent, 0.30, SE= 0.07; **Figure 8** in the main text and **Supplementary Table 22)**. For communication disorders, e^2^ increased from 0.16 (SE= 0.11) for samples 75-99% European ancestry to 0.24 (SE= 0.06) for samples where all participants were of European ancestry.

##### Ancestry-related differences in environmental overlap between NDDs

Differences in sources of co-occurrence between NDDs could not be estimated for shared and nonshared environmental overlap. Estimates for samples comprising only individuals of European ancestry are presented in **Supplementary Table 23**.

##### Ancestry-related differences in environmental overlap between NDDs and DICCs

We were able to estimate the meta-analytic shared environmental overlap between NDDs and DICCs, as 4 out of 5 studies reporting on genetic correlations also reported on shared environmental correlations. The grand shared environmental overlap remained stable across samples ancestral composition (0.88, SE= 0.87 and 0.89, SE= 0.85, respectively; **Supplementary Table 24)**.

### Supplementary Note 2: Meta-analytic results for NDDs phenotypic sub-categories.

Where the number of studies identified was sufficiently large, we were able to stratify sources of variance and co-occurrence by specific phenotypic sub-categories to reflect within-category differences. **Supplementary Figure 2** presents family and SNP-based heritability, shared and nonshared environmental influences on sub-categories of NDDs, whereas **Supplementary Figure 3** shows family-based genetic, shared and nonshared environmental overlap between sub-categories of NDDs, as well as between sub-categories of NDDs and DICCs. All estimates with standard errors are presented in **Supplementary Tables 2-5.**

For example, within intellectual disabilities, we estimated heritability of learning disability (0.86, SE= 0.43), which constitutes one of the sub-categories. Within communication disorders, we distinguished 5 specific phenotypes, out of which specific language impairment had the highest meta-analytic heritability (0.87, SE= 0.60), whereas the lowest grand heritability estimate was estimated for stuttering (0.58, SE= 0.17). All ADHD-related specific phenotypes were highly heritable, ranging from 0.76 (SE= 0.07) for impulsivity to 0.65 (SE= 0.05) for inattention. For ASD, the highest grand heritability was found for restrictive and repetitive behaviours and interests (0.83, SE= 0.49), whereas the lowest was found for social impairments (0.67, SE= 0.05). Within motor disorders, we identified 4 specific sub-categories. The highest grand heritability estimate was found for motor coordination (0.82, SE= 0.08) and the weakest for tic disorders (0.56, SE= 0.17).

Specific learning disorders were divided into three primary sub-categories, i.e., dyslexia, dysgraphia, and dyscalculia-related phenotypes with heritabilities ranging from 0.62 (SE= 0.04) for dyslexia (and/or the continuously measured phenotype of reading ability) to 0.56 (SE= 0.18) for dysgraphia (and/or the continuously measured phenotype of writing ability), and 0.55 (SE= 0.04) for dyscalculia (and/or the continuously measured phenotype of mathematics ability). The three subcategories of dyslexia, dysgraphia, and dyscalculia were further divided into secondary sub-categories comprising specific reading, writing and mathematics-related phenotypes. Within the dyslexia sub-category, the highest meta-analytic heritability was estimated for decoding (0.69, SE= 0.14), while the lowest for vocabulary (0.25, SE= 0.14). Within the dysgraphia-related phenotype, writing ability had a grand heritability estimate of 0.56 (SE= 0.17). Within the Dyscalculia sub-category, we identified 4 further specific phenotypes, out of which broadly defined mathematics ability was most heritable, with a meta-analytic estimate of 0.57 (SE= 0.04), with the lowest grand heritability obtained for mathematics problem solving (0.36, SE= 0.18).

Stratified estimates for specific phenotypes could also be calculated for a few homotypic and heterotypic co-occurrent disorders. The co-occurrence between ASD & ADHD was divided into 4 sub-categories, out of which the highest meta-analytic genetic correlation was obtained between broadly defined ASD & ADHD (0.71, SE= 0.27), while the lowest was estimated between restrictive and repetitive behaviours and interests & inattention (0.16, SE= 0.11; see **Supplementary Table 4**).

We could only distinguish only one specific phenotype sub-category for the co-occurrence between ADHD & motor disorders, namely the association between ADHD & developmental coordination disorder for which grand genetic correlation of 0.91 (SE= 0.80) was found. The co-occurrence between ADHD & specific learning disorders was stratified into 6 phenotypic sub-categories, with the overlap ranging between 0.19 (SE= 0.22) for ADHD & reading ability and -0.32 (SE= 0.11) for inattention & mathematic ability. The co-occurrence between specific language impairment and dyslexia was the only specific phenotype sub-category identified for the co-occurrence between communication disorders & specific learning disorders and yielded grand genetic overlap of 0.66 (0.15), whereas the co-occurrence between subtypes of specific learning disorders was stratified into dyslexia and dyscalculia and quantitatively measured reading ability and mathematics ability, both of which yielded comparable meta-analytic genetic overlaps: 0.56 (SE= 0.07) and 0.55 (SE= 0.08), respectively.

When considering the genetic overlap between NDDs and DICCs, stratification was only possible for the co-occurrence between ADHD & oppositional defiant disorder, where the grand genetic overlap between hyperactivity & oppositional defiant disorder traits was stronger (0.80, SE=0.57) if compared to the genetic overlap between inattention & oppositional defiant disorder traits (0.52, SE= 0.10).

### Supplementary Note 3: Description of moderators.

#### Age

The age group moderator was created based on age range of the study, or the mean age when the age range was not reported, and consisted of six levels, three separate categories and three groups cutting across age categories: *childhood* (ages 4-7), *middle childhood* (ages 8-10), *adolescence* (ages 11-24), *childhood & middle childhood* (ages 4-10), *middle childhood & adolescence* (ages 8-24) and *childhood & adolescence* (ages 4-24). The same age categories were used across all methods.

#### Design

The design covariate consisted of different categories, depending on whether the study had employed family or SNP-based methods. For family-based studies, 8 types of designs were identified: *classical twin study, categorical threshold twin study, DFextremes twin study, classical twin and sibling study, categorical threshold twin and sibling study, DFextremes twin and sibling study, classical sibling study and categorical threshold sibling study*. We identified two types of designs for SNP-based studies: those using genome-wide (*GREML*) and summary-level data (*LDSC*).

#### Model

When meta-analysing family-based studies we also controlled for type of model, i.e., *full* *model* (twin or twin and sibling studies reporting A, C and E estimates), *DFextremes full model* (DFextremes studies reporting A, C and E estimates), *best model* (twin or twin and sibling studies reporting best-fitting parsimonious models, that is either AE, CE or E only models), *DFextremes best model* (DFextremes studies reporting best-fitting parsimonious models, that is either AE, CE or E only models), *A only model* (twin or twin and sibling studies reporting heritability estimates only, without providing estimates of C and E), *DFextremes A only model* (DFextremes studies reporting heritability estimates only, without providing estimates of C and E).

#### Rater

Eight types of raters were identified with the meta-analytic dataset, referring to both family and SNP-based studies. NDD and DICC symptoms were rated by either *parents, teachers, self-reports*, or *researchers*, with several studies reporting cross-rater measures assessed by *parents & teachers* and *parents & self-reports*. In addition, specific learning disorders and communication disorders symptoms were often assessed using reading, writing, mathematical and language ability tests, hence *test* was also included as an additional level of this covariate. A further level, *diagnosis*, was also incorporated to reflect clinical diagnosis of NDDs and DICCs.

#### Measurement scale

Measurement scale moderator involved two levels, *continuous* reflecting quantitatively measured symptoms and *categorical* reflecting binary diagnoses and clinical cut-offs.

#### Ancestry

From studies that reported on the ancestral composition of the sample used in analyses we recorded the percentage of participants of European ancestry. We created the *%European ancestry* and created a moderator with four levels: *less than 50%, more than 50% but less than 75%, more than 75% but less than 100%* and *100%.*

#### Number of covariates

Behaviour genetic studies often include covariates in the models or regress covariates out prior to analyses. It is a common procedure to control for age and sex in both family and SNP-based studies, and additionally controlling for batch effects and population stratification in molecular genetics studies^2,3^. To determine the impact of including covariates on estimate heterogeneity, we created a moderator by adding up the number of covariates used in each study. This resulted in a moderator including 5 levels: 0 to 4 covariates included.

#### Measure

Further heterogeneity between studies may arise from differences in the measurement instruments used to assess NDDs and DICCs. Diagnostic and assessment tools tend to be specific to the disorder being measured, therefore we created a moderator variable indexing the specific measurement instrument used to assess each NDD category, with levels varying within and between conditions.

#### Country

The last moderator involved the country where each cohort was based. We distinguished eight levels of this moderator: Australia, Canada, China, Netherlands, Norway, Sweden, United Kingdom, and United States.

### Supplementary Note 4: Categorical versus continuous measurement of NDDs.

#### Family-based studies

Categorical phenotypes were measured by 28 family-based studies, whereas 215 studies reported estimates for continuous phenotypes. Higher grand heritability was estimated for categorically measured NDDs (0.77, SE= 0.07), compared to NDDs measured on a continuum (0.64, SE= 0.03) (**Supplementary Figure 22; Supplementary Table 25**). No significant differences in shared and nonshared environmental influences were present between measurement methods.

Disparities in family-based genetic overlap was found across co-occurrences between NDDs, with grand genetic correlation of 0.56 (SE= 0.32) estimated from studies using categorical phenotypes and 0.31 (SE= 0.12) estimated from studies using quantitative measures (**Supplementary Figure 22** and **Supplementary Table 26**). Shared and nonshared environmental genetic overlap could not be compared across co-occurrences between NDDs due to insufficient number of identified studies. Similarly, sources of co-occurrence could not be compared between measurement scales for the co-occurrence between NDDs and DICCs as less than 2 studies investigated categorically defined phenotypes (**Supplementary Figure 22** and **Supplementary Table 27**).

#### SNP-based studies

Categorically and quantitatively defined NDDs were measured by 12 and 17 SNP-based studies, respectively. Just as family-based heritability, SNP heritability across NDDs differed between measures: categorical phenotypes yielded lower heritability (0.17, SE= 0.03) estimates if compared to quantitatively measured symptom scores (0.25, SE= 0.06; **Supplementary Figure 22** and **Supplementary Table 25**).

### Supplementary Note 5: Indexes, timespans, search strategy and key words.

Searches were conducted with the aid of Covidence software (https://www.covidence.org/) and using the following sources:

1) Web of Science.

Core Collection Indexes and timespans:

•Science Citation Index Expanded (SCI-Expanded) -- 1900-present

•Social Sciences Citation Index (SSCI) -- 1900-present

•Arts & Humanities Citation Index (A&HCI) -- 1975-present

•Emerging Sources Citation Index (ESCI) -- 2015-present

•Conference Proceedings Citation Index - Science (CPCI-S) -- 1990-present •Conference Proceedings Citation Index - Social Sciences & Humanities (CPCI SSH) -- 1990-present

2) Ovid platform.

Indexes and timespans:

• Embase (1974 - present)

• Ovid MEDLINE(R), including Epub Ahead of Print and In-Process & Other Non- Indexed Citations (1946 - present)

• LWW Health Library: Speech, Language & Hearing Collection

• Global Health (1973 - present)

• PsycINFO (1806 - present)

To identify studies focusing on the phenotypes of interest, we used the following key terms in the first (primary) search:

((heritab* OR genetic* OR twin* OR genom* OR sibling*) AND (Neurodevelopmental OR “Intellectual* Disabilit*” OR “Learning* Disabilit*” OR “Intellectual* Developmental*

Disorder*” OR “Global* Developmental* Delay” OR “Communication Disorder*” OR “Language Disorder*” OR “Speech* Sound* Disorder*” OR “Childhood-Onset* Fluency* Disorder*” OR Stutter* OR “Social Communication Disorder*” OR “Pragmatic Communication Disorder*” OR Autis* OR ASD OR “Attention-Deficit*” OR Hyperactiv* OR Hyperkinetic OR Inattent* OR ADHD OR “Specific Learning Disorder*” OR SLD OR Dyslex* OR Dysgraph* OR Dyscalcul* OR “Motor Disorder*” OR “Developmental Coordination Disorder*” OR Dysprax* OR “Stereotypic Movement Disorder*” OR “Tic* Disorder*” OR “Tourett* Disorder*” OR Disruptive OR “Impulse control” OR “Oppositional Defiant Disorder*” OR ODD OR “Intermittent* Explosive* Disorder*” OR “Conduct* disorder” OR Antisocial* OR APD OR Pyromani* OR Kleptomani* OR “behavio* problem*” OR Deliquen* OR Externalizing))

In the second (confirmatory) search, we decided to include an additional set of terms to capture studies focusing on Specific Learning Disorder and Communication Disorder measured on a continuum (i.e., reading, mathematics, writing, language) that had not been identified by the diagnosis-related search terms (i.e., dyslexia, dyscalculia, dysgraphia, language disorder). The following confirmatory search terms were used:

((heritab* OR genetic* OR twin* OR genom* OR sibling*) AND (Neurodevelopmental OR “Intellectual* Disabilit*” OR “Learning* Disabilit*” OR “Intellectual* Developmental*

Disorder*” OR “Global* Developmental* Delay” OR “Communication Disorder*” OR “Language Disorder*” OR “Speech* Sound* Disorder*” OR “Childhood-Onset* Fluency* Disorder*” OR Stutter* OR “Social Communication Disorder*” OR “Pragmatic Communication Disorder*” OR Autis* OR ASD OR “Attention-Deficit*” OR Hyperactiv* OR Hyperkinetic OR Inattent* OR ADHD OR “Specific Learning Disorder*” OR SLD OR Dyslex* OR Dysgraph* OR Dyscalcul* OR Reading OR Math* OR Writing OR Language OR “Motor Disorder*” OR “Developmental Coordination Disorder*” OR Dysprax* OR “Stereotypic Movement Disorder*” OR “Tic* Disorder*” OR “Tourett* Disorder*” OR Disruptive OR “Impulse control” OR “Oppositional Defiant Disorder*” OR ODD OR “Intermittent* Explosive* Disorder*” OR “Conduct* disorder” OR Antisocial* OR APD OR Pyromani* OR Kleptomani* OR “behavio* problem*” OR Deliquen* OR Externalizing))

### Supplementary Note 6: Description of SNP-based methods targeted by the meta-analysis.

#### Genome-wide complex trait analysis and restricted maximum likelihood (GCTA; REML)

Genome-wide complex trait analysis (GCTA) software employs restricted maximum likelihood method (REML) that allows for the estimation of the variance in a trait that is captured by single nucleotide polymorphisms (SNPs) assessed on SNP arrays commonly used in GWAS^4^. This method estimates SNP heritability from DNA using unrelated individuals. The first step is to calculate a genetic relatedness matrix by weighting genetic similarities between all possible pairs of individuals by the allele frequencies across all SNPs on the SNP array. The matrix of pair-by-pair genetic similarity is compared to the matrix of pair-by-pair phenotypic similarity using residual maximum likelihood estimation to obtain the proportion of phenotypic variation accounted for by genetic variation. GCTA can also be used to quantify the degree of shared genetic variance (genetic covariance) between two phenotypes, such as two disorders^4^.

#### Linkage disequilibrium score regression (LDSC)

LDSC quantifies the proportion of variance in a trait explained by common genetic variants (i.e., SNP heritability), as well as the proportion of shared genetic variance between traits (i.e., genetic covariance), using GWAS summary statistics^5^. LDSC applies regression to calculate the association between SNP test statistics obtained from GWAS results, and linkage disequilibrium (LD) scores, therefore allowing us to dissect the true polygenic signal (i.e., the contribution of multiple genetic variants of small effect to variability in a trait or disorder) from confounding signal, including for example false positive associations due to population stratification^5^.

#### Summary-data-based BayesS (SBayeS)

SBayeS is a Bayesian approach to estimating SNP heritability using GWA summary statistics^6^. SBayeS employs an array of linear mixed models using GWA data to estimate SNP heritability, as well as polygenicity and the relationship between variant effect sizes and minor allele frequencies^6^.

### Supplementary Note 7: Quality scoring checklist.

Quality scoring of studies included in the meta-analysis was conducted in line with a framework proposed by Kmet, Cook and Lee (2004)^7^.

We used the following checklist:

1. Question/objective sufficiently described?

2. Study design evident and appropriate?

3. Method of subject/comparison group selection or source of

information/input variables described and appropriate?

4. Subject (and comparison group, if applicable) characteristics sufficiently described?

5. Outcome and (if applicable) exposure measure(s) well defined and robust to

measurement / misclassification bias? Means of assessment reported?

6. Analytic methods described/justified and appropriate?

7. Some estimate of variance is reported for the main results?

8. Results reported in sufficient detail?

9. Conclusions supported by the results?

To estimate study quality, items were scored based on the scale developed by Kmet et. al. (2004)^7^, where: 0= NO, 1= PARTIAL and 2= YES. Quality scoring was conducted by the primary reviewer and checked by a secondary reviewer. Following completion of the checklist, we calculated the mean total score obtained by each reviewer to ensure inter-rater agreement. Reviewer discrepancies were identified and resolved through discussion.

### Supplementary Note 8: Requesting missing data from study authors.

### The first author of *Partitioning the heritability of Tourette syndrome and obsessive compulsive disorder reveals differences in genetic architecture*^8^ was contacted via e-mail about the age range of the sample. Response was received that the age range of the sample was not restricted and consisted of both children and adults. Therefore, the study was not included in the meta-analysis.

### We contacted authors of two other studies via ResearchGate, however we did not receive a response.

### Supplementary Note 9: Aggregation sensitivity analyses.

We explored multiple aggregation techniques, that is aggregating non-independent effect sizes by study, by cohort, as well as by country. Furthermore, we checked whether estimates differed when setting different correlation thresholds (r= 0.3, r= 0.5 and r= 0.9) for aggregating between effect sizes. Grand estimates across all NDDs and co-occurring disorders resulting from various aggregation methods are presented in **Supplementary Figure 24**. Grand estimates were not significantly different across aggregation methods and correlation thresholds, therefore we proceeded with aggregating by study and set a fixed correlation between related effect sizes of r= 0.5 for all downstream analyses.

### Supplementary Note 10: Heterogeneity assessment.

Across all NDDs we found that 74% of the total variance in family-based heritability was due to heterogeneity, out of which 53% could be attributed to between-cluster and 22% to within-cluster heterogeneity, where clusters refer to cohorts and individual studies (**Supplementary figure 6; Supplementary table 7**). The lowest I^2^ statistic was estimated for motor disorders (36%, with equal contribution of between and within-cluster heterogeneity of 18% each), while the highest one for ASD (86%, where 78% was attributed to between-cluster and 8% to within-cluster heterogeneity). When considering SNP heritability, the proportion of total variance accounted for by heterogeneity was very low across disorders (6-8%, most of which was represented by between-cluster heterogeneity). Total variance in shared environmental influences across NDDs was moderate (18%) and almost exclusively attributable to within-cluster heterogeneity. The highest proportion of variance in shared environmental influences accounted for by heterogeneity was found for ASD (41%) and was accounted for solely by within-cluster heterogeneity, while the lowest was found for specific learning disorders and motor disorders, for which variance explained by heterogeneity was less than 0.001%. A similar degree of heterogeneity was estimated for nonshared environmental factors, where the variance explained across NDDs was 38% (21% and 17% attributed to between and within-cluster heterogeneity, respectively) and ranged from 43% (accounted solely by within-cluster heterogeneity) for ADHD to less than 0.001% for intellectual disabilities.

Overall, genetic correlations between NDDs were estimated as 89%, with 34% attributed to between-cluster and 55% to within-cluster heterogeneity (**Supplementary figure 6; Supplementary table 8**). The largest proportion of total variance accounted for by heterogeneity was estimated for the co-occurrence between ADHD & motor disorders (99%, with equal contribution of between and within-cluster heterogeneity of 49%), whereas the lowest one was estimated for the co-occurrence between communication disorders & motor disorders and communication disorders & specific learning disorders (<0.001% each). Heterogeneity in SNP-based genetic overlap across co-occurrences between NDDs accounted for 49% of the total variance, with 33% attributed to between-cluster and 15% to within-cluster heterogeneity. Between ASD & ADHD, 24% of the total variance was explained by heterogeneity, all of which was accounted for by between-cluster heterogeneity.

Variance in shared environmental overlap across co-occurrences between NDDs accounted for by heterogeneity was estimated as 95%, with 36% attributed to between-cluster and 59% to within-cluster heterogeneity and for the only pair of NDDs where meta-analysis of shared environmental correlations was possible, i.e., ADHD & specific learning disorders, we found 53% of the total variance to be explained by heterogeneity with 6% attributed to between-cluster and 47% to within-cluster heterogeneity. Variance in nonshared environmental overlap across NDDs was modest (24%, all accounted for by between-cluster heterogeneity) and ranged from 62% (all accounted for by between-cluster heterogeneity) for the co-occurrence between ASD & ADHD to less than 0.001% for the co-occurrence between ADHD & specific learning disorders.

Finally, 93% of the total variance in genetic overlap across co-occurrences between NDDs and DICCs was accounted for by heterogeneity, with 55% attributed to between-cluster and 38% to within-cluster heterogeneity (**Supplementary Figure 6** and **Supplementary Table 9**). The variance explained by heterogeneity was high for co-occurrence between ADHD & conduct disorder (92%, with equal contribution of between and within-cluster heterogeneity, 46% each) and between ADHD & oppositional defiant disorder (84%, with equal contribution of between and within-cluster heterogeneity, 42% each), but much lower between ASD & conduct disorder (less than 0.001%). In case of shared environmental overlap between NDDs and DICCs, 95% of the variance was due to heterogeneity and was solely accounted for by within-cluster heterogeneity. The highest proportion of variance in shared environmental correlations explained by heterogeneity was estimated for co-occurrence between ADHD & conduct disorder (96%, with equal contribution of between and within-cluster heterogeneity, 48% each), whereas the lowest was estimated between ASD & conduct disorder (67%, all accounted for by within-cluster heterogeneity). Total variance in nonshared environmental overlap was high across all co-occurrences between NDDs and DICCs (91%, all accounted for by within-cluster heterogeneity), as well as between ADHD & oppositional defiant disorder (92%, equally accounted for by between and within-cluster heterogeneity, 46% each), whereas less than 0.001% of variance in nonshared environmental overlap between ADHD & conduct disorder and ASD & conduct disorder was explained by heterogeneity.

### Supplementary Note 11: Publication bias

Publication bias refers to the higher probability of studies reporting statistically significant findings being accepted for publication. In an unbiased scenario, we would expect to find as many studies reporting significant results, as those not rejecting the null hypothesis. The publication bias can be reflected by the linear relationship between the estimate and standard error^9^. **Supplementary Figures 5-11** include funnel plots of studies that reported estimates of heritability, shared and nonshared environmental influences on NDDs. **Supplementary Table 10** presents the results of Egger’s regressions for all NDDs, apart from intellectual disabilities where the number of parameters to be estimated was larger than the number of studies. A significant risk of publication bias (z= -3.95, beta= 0.73 (95% CIs: 0.69, .78), p< 0.001) for family-based heritability was found across all NDDs, largely driven by ADHD and specific learning disorders. The overall relationship between shared environmental influences and their standard errors was significant across all NDDs, suggesting the greater likelihood of reporting significant estimates in larger studies. This relationship was not significant for specific NDDs. Publication bias was also found for nonshared environmental influences across all NDDs, which was likely driven by nonshared environmental influences on ADHD. Risk of publication bias was not observed for SNP heritability.

**Supplementary Figures 12-17** include funnel plots of studies that reported estimates of genetic, shared and nonshared environmental overlap between NDDs. **Supplementary Table 11** presents the results of Egger’s regressions across all comorbidities between NDDs, as well as for comorbidities between ASD & ADHD and ADHD & specific learning disorders. For the remaining comorbidities between NDDs the number of parameters to be estimated was larger than the number of studies identified. Risk of publication bias was not significant for family-based genetic and environmental correlations nor for SNP-based genetic correlations.

**Supplementary Figures 18-21** include funnel plots of studies that reported estimates for the genetic, shared and nonshared environmental overlap between NDDs and DICCs. **Supplementary Table 12** presents the results of Egger’s regressions across all comorbidities between NDDs and DICCs, as well as for comorbidities between ADHD & conduct disorder and ADHD & oppositional defiant disorder and ASD & antisocial personality disorder. We found a significant relationship between environmental influences and standard errors, i.e., publication bias, for shared environmental correlation between all NDDs and all DICCs, and, when considering specific disorder categories, between ADHD & conduct disorder.

### Supplementary Tables

| Supplementary Tables: table of contents | |
| --- | --- |
| Supplementary Table 1 | Page 26 |
| Supplementary Table 2 | Pages 27-28 |
| Supplementary Table 3 | Page 29 |
| Supplementary Table 4 | Page 30 |
| Supplementary Table 5 | Page 31 |
| Supplementary Table 6 | Page 32 |
| Supplementary Table 7 | Page 33 |
| Supplementary Table 8 | Page 34 |
| Supplementary Table 9 | Page 35 |
| Supplementary Table 10 | Page 36 |
| Supplementary Table 11 | Page 37 |
| Supplementary Table 12 | Page 38 |
| Supplementary Table 13 | Page 39 |
| Supplementary Table 14 | Page 40 |
| Supplementary Table 15 | Page 41 |
| Supplementary Table 16 | Pages 42-43 |
| Supplementary Table 17 | Page 44 |
| Supplementary Table 18 | Page 45 |
| Supplementary Table 19 | Pages 46-47 |
| Supplementary Table 20 | Page 48 |
| Supplementary Table 21 | Page 49 |
| Supplementary Table 22 | Page 50 |
| Supplementary Table 23 | Page 51 |
| Supplementary Table 24 | Page 52 |
| Supplementary Table 25 | Page 53 |
| Supplementary Table 26 | Page 54 |
| Supplementary Table 27 | Page 55 |
| Supplementary Table 28 | Pages 56-75 |
| Supplementary Table 29 | Pages 76-80 |
| Supplementary Table 30 | Pages 81-84 |
| Supplementary Table 31 | Pages 85-87 |
| Supplementary Table 32 | Page 88 |
| Supplementary Table 33 | Page 89 |
| Supplementary Table 34 | Pages 90-91 |
| Supplementary Table 35 | Page 92 |
| Supplementary Table 36 | Page 93 |
| Supplementary Table 37 | Pages 94-95 |
| Supplementary Table 38 | Page 96 |
| Supplementary Table 39 | Page 97 |
| Supplementary Table 40 | Pages 98-99 |
| Supplementary Table 41 | Page 100 |
| Supplementary Table 42 | Page 101 |
| Supplementary Table 43 | Pages 102-103 |
| Supplementary Table 44 | Page 104 |
| Supplementary Table 45 | Page 105 |
| Supplementary Table 46 | Pages 106-108 |
| Supplementary Table 47 | Page 109 |

**Supplementary Table 1**: Heritability, shared and nonshared environmental influences on NDDs.

**Supplementary Table 2**: Heritability, shared and nonshared environmental influences on NDDs, stratified by specific phenotypic sub-categories.

**Supplementary Table 3**: Genetic, shared and nonshared environmental correlations between NDDs.

**Supplementary Table 4**: Genetic, shared and nonshared environmental correlations between NDDs, stratified by specific phenotypic sub-categories.

**Supplementary Table 5**: Genetic, shared and nonshared environmental correlations between NDDs and DICCs.

**Supplementary Table 6**: Genetic, shared and nonshared environmental correlations between NDDs and DICCs, stratified by specific phenotypic sub-categories.

**Supplementary Table 7**: Proportion of variance in heritability, shared and nonshared environmental influences on NDDs accounted for by heterogeneity.

**Supplementary Table 8**: Proportion of variance in genetic, shared and nonshared environmental correlations between NDDs accounted for by heterogeneity.

**Supplementary Table 9**: Proportion of variance in genetic, shared and nonshared environmental correlations between NDDs and DICCs accounted for by heterogeneity.

**Supplementary Table 10**: Results of Egger’s regression for studies addressing heritability and environmental influences on NDDs.

**Supplementary Table 11**: Results of Egger’s regression for studies addressing genetic and environmental overlap between NDDs.

**Supplementary Table 12**: Results of Egger’s regression for studies addressing genetic and environmental overlap between NDDs and DICCs.

**Supplementary Table 13**: Sex-specific heritability, shared and nonshared environmental influences on NDDs.

**Supplementary Table 14**: Sex-specific genetic, shared and nonshared environmental correlations between NDDs.

**Supplementary Table 15**: Sex-specific genetic, shared and nonshared environmental correlations between NDDs and DICCs.

**Supplementary Table 16**: Heritability, shared and nonshared environmental influences on NDDs, stratified by age categories.

**Supplementary Table 17**: Genetic, shared and nonshared environmental correlations between NDDs, stratified by age categories.

**Supplementary Table 18**: Genetic, shared and nonshared environmental correlations between NDDs and DICCs, stratified by age categories.

**Supplementary Table 19**: Heritability, shared and nonshared environmental influences on NDDs, stratified by countries.

**Supplementary Table 20**: Genetic, shared and nonshared environmental correlations between NDDs, stratified by countries.

**Supplementary Table 21**: Genetic, shared and nonshared environmental correlations between NDDs and DICCs, stratified by countries.

**Supplementary Table 22**: Heritability, shared and nonshared environmental influences on NDDs, stratified by the percentage of individuals of European ancestry.

**Supplementary Table 23**: Genetic, shared and nonshared environmental correlations between NDDs, stratified by the percentage of individuals of European ancestry.

**Supplementary Table 24**: Genetic, shared and nonshared environmental correlations between NDDs and DICCs, stratified by the percentage of individuals of European ancestry.

**Supplementary Table 25**: Heritability, shared and nonshared environmental influences on NDDs, stratified by measurement scales.

**Supplementary Table 26**: Genetic, shared and nonshared environmental correlations between NDDs, stratified by measurement scales.

**Supplementary Table 27**: Genetic, shared and nonshared environmental correlations between NDDs and DICCs, stratified by measurement scales.

**Supplementary Table 28**: Overview of family-based studies using samples of males and females combined. Co-occurrences between disorders annotated with an asterisk (*) indicate pairs of disorders for which meta-analysis could not be performed.

**Supplementary Table 29**: Overview of family-based studies using male samples. Co-occurrences between disorders annotated with an asterisk (*) indicate pairs of disorders for which meta-analysis could not be performed.

**Supplementary Table 30**: Overview of family-based studies using female samples. Co-occurrences between disorders annotated with an asterisk (*) indicate pairs of disorders for which meta-analysis could not be performed.

**Supplementary Table 31**: Overview of SNP-based studies using samples of males and females combined. Disorders annotated with an asterisk (*) indicate disorders for which meta-analysis could not be performed.

**Supplementary Table 32**: Overview of SNP-based studies using male samples. Disorders annotated with an asterisk (*) indicate disorders for which meta-analysis could not be performed.

**Supplementary Table 33**: Overview of SNP-based studies using female samples. Disorders annotated with an asterisk (*) indicate disorders for which meta-analysis could not be performed.

**Supplementary Table 34**: Heritability, shared and nonshared environmental influences on NDDs, stratified by designs.

**Supplementary Table 35**: Genetic, shared and nonshared environmental correlations between NDDs, stratified by designs.

**Supplementary Table 36**: Genetic, shared and nonshared environmental correlations between NDDs and DICCs, stratified by designs.

**Supplementary Table 37**: Heritability, shared and nonshared environmental influences on NDDs, stratified by models.

**Supplementary Table 38**: Genetic, shared and nonshared environmental correlations between NDDs, stratified by models.

**Supplementary Table 39**: Genetic, shared and nonshared environmental correlations between NDDs and DICCs, stratified by models.

**Supplementary Table 40**: Heritability, shared and nonshared environmental influences on NDDs, stratified by raters.

**Supplementary Table 41**: Genetic, shared and nonshared environmental correlations between NDDs, stratified by raters.

**Supplementary Table 42**: Genetic, shared and nonshared environmental correlations between NDDs and DICCs, stratified by raters.

**Supplementary Table 43**: Heritability, shared and nonshared environmental influences on NDDs, stratified by number of covariates included in analyses.

**Supplementary Table 44**: Genetic, shared and nonshared environmental correlations between NDDs, stratified by number of covariates included in analyses.

**Supplementary Table 45**: Genetic, shared and nonshared environmental correlations between NDDs and DICCs, stratified by number of covariates included in analyses.

**Supplementary Table 46**: Heritability, shared and nonshared environmental influences on NDDs, stratified by measurement instruments.

**Supplementary Table 47**: Genetic, shared and nonshared environmental correlations between NDDs, stratified by measurement instruments.

| **Supplementary Table 1**. Heritability, shared and nonshared environmental influences on NDDs. | | | | | | | | |
| --- | --- | --- | --- | --- | --- | --- | --- | --- |
| **NDDs** | **Family h^2^ (SE)** | **N** | **Family c^2^ (SE)** | **N** | **Family e^2^ (SE)** | **N** | **SNP h^2^ (SE)** | **N** |
| **NDDs combined** | 0.66 (0.03) | 237 | 0.17 (0.02) | 127 | 0.29 (0.02) | 195 | 0.19 (0.03) | 29 |
| **Intellectual disabilities** | 0.86 (0.44) | 2 | - | - | 0.1 (0.16) | 2 | - | - |
| **Communication disorders** | 0.64 (0.19) | 23 | 0.35 (0.06) | 15 | 0.21 (0.04) | 18 | 0.32 (0.14) | 4 |
| **ASD** | 0.76 (0.11) | 36 | 0.13 (0.05) | 14 | 0.27 (0.03) | 28 | 0.14 (0.04) | 15 |
| **ADHD** | 0.67 (0.04) | 121 | 0.11 (0.02) | 48 | 0.3 (0.02) | 107 | 0.20 (0.04) | 14 |
| **Specific learning disorders** | 0.62 (0.04) | 89 | 0.19 (0.02) | 65 | 0.24 (0.02) | 67 | 0.30 (0.08) | 9 |
| **Motor disorders** | 0.74 (0.08) | 7 | 0.13 (0.11) | 3 | 0.38 (0.11) | 6 | - | - |
| *Note.* H^2^= heritability; c^2^= shared environmental influences; e^2^= nonshared environmental influences; N= number of studies identified;  SE= standard error. | | | | | | | | |

| **Supplementary Table 2.** Heritability, shared and nonshared environmental influences on NDDs, stratified by specific phenotypic sub-categories. | | | | | | | | | |
| --- | --- | --- | --- | --- | --- | --- | --- | --- | --- |
| **Specific phenotypes from family-based studies** | | | | | | | **Specific phenotypes from SNP-based studies** | | |
| **NDDs** | **Family h2 (SE)** | **N** | **Family c2 (SE)** | **N** | **Family e2 (SE)** | **N** |  | **SNP h2 (SE)** | **N** |
| **Intellectual disabilities** | | | | | | | | | |
| Learning disability | 0.86 (0.44) | 2 | - | - | 0.1 (0.16) | 2 |  |  |  |
| **Communication disorders** | | | | | | | | | |
| Language ability | 0.65 (0.2) | 20 | 0.36 (0.07) | 13 | 0.21 (0.04) | 15 | Language ability | 0.32 (0.14) | 4 |
| Specific language impairment | 0.87 (0.6) | 2 | - | - | - | - |  |  |  |
| Speech | 0.8 (0.17) | 2 | - | - | 0.2 (0.15) | 2 |  |  |  |
| Stuttering | 0.58 (0.17) | 2 | - | - | 0.21 (0.12) | 2 |  |  |  |
| Syntax | 0.65 (0.37) | 2 | - | - | 0.49 (0.24) | 2 |  |  |  |
| **ASD** | | | | | | | | | |
| ASD | 0.79 (0.14) | 26 | 0.06 (0.04) | 12 | 0.26 (0.03) | 19 | ASD | 0.13 (0.04) | 10 |
| CIs | 0.76 (0.09) | 8 | - | - | 0.27 (0.06) | 5 | Sis | 0.2 (0.09) | 6 |
| RRBIs | 0.83 (0.49) | 10 | 0.24 (0.24) | 2 | 0.35 (0.09) | 6 |  |  |  |
| Sis | 0.67 (0.05) | 15 | 0.31 (0.22) | 3 | 0.3 (0.05) | 11 |  |  |  |
| Strict autism | 0.51 (0.28) | 2 | - | - | - | - |  |  |  |
| **ADHD** | | | | | | | | | |
| ADHD | 0.7 (0.05) | 54 | 0.12 (0.03) | 22 | 0.3 (0.03) | 47 | ADHD | 0.21 (0.04) | 11 |
| Hyperactivity | 0.66 (0.16) | 2 | - | - | 0.38 (0.11) | 2 | Hyperactivity/Impulsivity | 0.13 (0.11) | 5 |
| Impulsivity | 0.76 (0.07) | 2 | - | - | 0.24 (0.08) | 2 | Inattention | 0.27 (0.17) | 4 |
| Hyperactivity/Impulsivity | 0.69 (0.06) | 63 | 0.16 (0.06) | 24 | 0.27 (0.03) | 56 |  |  |  |
| Inattention | 0.65 (0.05) | 65 | 0.08 (0.03) | 26 | 0.28 (0.02) | 58 |  |  |  |
| **Specific learning disorders** | | | | | | | | | |
| Dyslexia | 0.62 (0.04) | 76 | 0.19 (0.02) | 55 | 0.23 (0.02) | 55 |  |  |  |
| Dysgraphia | 0.56 (0.18) | 3 | 0.08 (0.08) | 3 | 0.38 (0.12) | 3 |  |  |  |
| Dyscalculia | 0.55 (0.04) | 30 | 0.19 (0.04) | 24 | 0.27 (0.02) | 25 |  |  |  |
| Decoding | 0.69 (0.14) | 7 | 0.17 (0.1) | 6 | 0.15 (0.06) | 6 |  |  |  |
| Grammar | 0.55 (0.1) | 2 | 0.3 (0.24) | 2 | 0.26 (0.1) | 2 |  |  |  |
| Nonword reading | 0.67 (0.13) | 3 | - | - | - | - |  |  |  |
| Orthographic skills | 0.49 (0.15) | 4 | 0.46 (0.18) | 2 | - | - |  |  |  |
| Phonological skills | 0.59 (0.09) | 13 | 0.2 (0.08) | 11 | 0.23 (0.06) | 10 |  |  |  |
| Rapid naming | 0.6 (0.12) | 7 | 0.17 (0.13) | 5 | 0.25 (0.08) | 5 |  |  |  |
| Reading ability | 0.62 (0.04) | 51 | 0.19 (0.03) | 33 | 0.23 (0.03) | 34 |  |  |  |
| Reading comprehension | 0.56 (0.07) | 11 | 0.19 (0.07) | 10 | 0.26 (0.05) | 10 |  |  |  |
| Reading fluency | 0.64 (0.13) | 5 | 0.16 (0.09) | 4 | 0.25 (0.06) | 4 |  |  |  |
| Spelling | 0.62 (0.11) | 8 | 0.14 (0.08) | 6 | 0.23 (0.06) | 6 |  |  |  |
| Vocabulary | 0.25 (0.14) | 4 | 0.57 (0.15) | 4 | 0.18 (0.07) | 4 |  |  |  |
| Word reading | 0.65 (0.08) | 16 | 0.22 (0.06) | 13 | 0.12 (0.04) | 13 |  |  |  |
| Writing ability | 0.56 (0.18) | 3 | 0.08 (0.08) | 3 | 0.38 (0.12) | 3 |  |  |  |
| Calculations | 0.39 (0.13) | 3 | - | - | 0.55 (0.23) | 2 |  |  |  |
| Mathematic ability | 0.57 (0.04) | 27 | 0.19 (0.04) | 22 | 0.25 (0.02) | 22 |  |  |  |
| Mathematic fluency | 0.52 (0.14) | 5 | 0.21 (0.14) | 4 | 0.27 (0.09) | 4 |  |  |  |
| Mathematic problems solving | 0.36 (0.19) | 2 | 0.28 (0.19) | 2 | 0.36 (0.13) | 2 |  |  |  |
| **Motor disorders** | | | | | | | | | |
| Coordination | 0.82 (0.07) | 2 | - | - | 0.38 (0.26) | 2 |  |  |  |
| DCD | 0.69 (0.13) | 2 | 0.12 (0.15) | 2 | 0.43 (0.2) | 3 |  |  |  |
| Motor control | 0.68 (0.12) | 2 | - | - | 0.41 (0.33) | 2 |  |  |  |
| Tics | 0.56 (0.17) | 2 | - | - | 0.44 (0.16) | 2 |  |  |  |
| *Note.* H^2^= heritability; c^2^= shared environmental influences; e^2^= nonshared environmental influences; N= number of studies identified;  SE= standard error; Sis= social impairments; CIs= communication impairments; RRBIs= restrictive, repetitive behaviours and interests; DCD= developmental coordination disorder. | | | | | | | | | |

| **Supplementary Table 3**. Genetic, shared and nonshared environmental correlations between NDDs. | | | | | | | | |
| --- | --- | --- | --- | --- | --- | --- | --- | --- |
| **NDDs** | **Family rA (SE)** | **N** | **Family rC (SE)** | **N** | **Family rE (SE)** | **N** | **SNP rG (SE)** | **N** |
| **NDDs combined** | 0.36 (0.12) | 37 | 0.63 (0.33) | 16 | 0.17 (0.05) | 22 | 0.39 (0.19) | 6 |
| **ASD & ADHD** | 0.67 (0.3) | 6 | - | - | 0.22 (0.13) | 5 | 0.26 (0.14) | 5 |
| **ADHD & motor disorders** | 0.9 (0.82) | 2 | - | - | - | - | - | - |
| **ADHD & specific learning disorders** | 0.07 (0.12) | 18 | 0.32 (0.14) | 7 | 0.11 (0.04) | 9 | - | - |
| **Communication disorders & motor disorders** | 0.33 (0.16) | 2 | - | - | - | - | - | - |
| **Communication disorders & specific learning disorders** | 0.66 (0.15) | 2 | - | - | - | - | - | - |
| *Note.* rA= genetic correlation; rC= shared environmental correlation; rE= nonshared environmental correlation; N= number of studies identified; SE= standard error. | | | | | | | | |

| **Supplementary Table 4.** Genetic, shared and nonshared environmental correlations between NDDs, stratified by specific phenotypic sub-categories. | | | | | | |
| --- | --- | --- | --- | --- | --- | --- |
| **NDDs** | **Family rA (SE)** | **N** | **Family rC (SE)** | **N** | **Family rE (SE)** | **N** |
| **ASD & ADHD** | | | | | | |
| ASD & ADHD | 0.71 (0.27) | 4 | - | - | 0.27 (0.11) | 3 |
| Hyperactivity & Sis | 0.22 (0.19) | 2 | - | - | 0.02 (0.08) | 2 |
| Inattention & RRBIs | 0.16 (0.11) | 2 | - | - | 0.09 (0.11) | 2 |
| Inattention & Sis | 0.27 (0.24) | 2 | - | - | 0.03 (0.08) | 2 |
| **ADHD & motor disorders** | | | | | | |
| ADHD & DCD | 0.91 (0.8) | 2 | - | - | - | - |
| **ADHD & specific learning disorders** | | | | | | |
| ADHD & Dyslexia | 0.07 (0.12) | 17 | 0.32 (0.15) | 7 | 0.11 (0.04) | 9 |
| ADHD & Dyscalculia | -0.29 (0.11) | 2 | - | - | 0.09 (0.1) | 2 |
| ADHD & Reading ability | 0.19 (0.22) | 6 | 0.12 (0.11) | 3 | 0.1 (0.08) | 3 |
| Hyperactivity & Reading ability | 0.11 (0.08) | 11 | 0.66 (0.19) | 4 | 0.03 (0.05) | 6 |
| Inattention & Reading ability | 0.07 (0.16) | 13 | 0.43 (0.26) | 5 | 0.16 (0.06) | 7 |
| inattention & Maths ability | -0.32 (0.11) | 2 | - | - | 0.15 (0.1) | 2 |
| **Communication disorders & specific learning disorders** | | | | | | |
| Specific language disorder & dyslexia | 0.66 (0.15) | 2 | - | - | - | - |
| *Note.* rA= genetic correlation; rC= shared environmental correlation; rE= nonshared environmental correlation; N= number of studies identified; SE= standard error; Sis= social impairments; RRBIs= restrictive, repetitive behaviours and interests; DCD= developmental coordination disorder. | | | | | | |

| **Supplementary Table 5**. Genetic, shared and nonshared environmental correlations between NDDs and DICCs. | | | | | | |
| --- | --- | --- | --- | --- | --- | --- |
| **NDDs and DICCs** | **Family rA (SE)** | **N** | **Family rC (SE)** | **N** | **Family rE (SE)** | **N** |
| **NDDs and DICCs combined** | 0.62 (0.19) | 15 | 0.88 (0.34) | 11 | 0.38 (0.14) | 13 |
| **ADHD & conduct disorder** | 0.66 (0.36) | 6 | 0.94 (0.71) | 3 | 0.11 (0.08) | 5 |
| **ADHD & oppositional defiant disorder** | 0.66 (0.18) | 6 | 0.96 (0.57) | 4 | 0.54 (0.25) | 5 |
| **ASD & conduct disorder** | 0.35 (0.10) | 3 | 0.88 (0.57) | 3 | 0.07 (0.08) | 3 |
| *Note.* rA= genetic correlation; rC= shared environmental correlation; rE= nonshared environmental correlation; N= number of studies identified; SE= standard error. | | | | | | |

| **Supplementary Table 6.** Genetic, shared and nonshared environmental correlations between NDDs and DICCs, stratified by specific phenotypic sub-categories. | | | | | | |
| --- | --- | --- | --- | --- | --- | --- |
| **NDDs and DICCs** | **Family rA (SE)** | **N** | **Family rC (SE)** | **N** | **Family rE (SE)** | **N** |
| **ADHD & oppositional defiant disorder** | | | | | | |
| ADHD & oppositional defiant disorder | 0.58 (0.2) | 5 | 0.95 (0.68) | 3 | 0.29 (0.1) | 4 |
| Hyperactivity & oppositional defiant disorder | 0.8 (0.57) | 2 | 0.87 (0.86) | 2 | 0.87 (0.74) | 2 |
| Inattention & oppositional defiant disorder | 0.52 (0.1) | 2 | - | - | 0.49 (0.11) | 2 |
| *Note.* rA= genetic correlation; rC= shared environmental correlation; rE= nonshared environmental correlation; N= number of studies identified; SE= standard error. | | | | | | |

| **Supplementary Table 7.** Proportion of variance in heritability, shared and nonshared environmental influences on NDDs accounted for by heterogeneity. | | | | | | | | | | | | |
| --- | --- | --- | --- | --- | --- | --- | --- | --- | --- | --- | --- | --- |
|  | **Family h2** | | | **Family c2** | | | **Family e2** | | | **SNP h2** | | |
| **NDDs** | **I^2^_t_** | **I^2^_b_** | **I^2^_w_** | **I^2^_t_** | **I^2^_b_** | **I^2^_w_** | **I^2^_t_** | **I^2^_b_** | **I^2^_w_** | **I^2^_t_** | **I^2^_b_** | **I^2^_w_** |
| **NDDs combined** | 0.75 | 0.53 | 0.21 | 0.18 | <0.001 | 0.18 | 0.38 | 0.21 | 0.17 | <0.001 | <0.001 | <0.001 |
| **Intellectual disabilities** | 0.84 | 0.42 | 0.42 | - | - | - | <0.001 | <0.001 | <0.001 | - | - | - |
| **Communication disorders** | 0.82 | 0.74 | 0.09 | 0.21 | <0.001 | 0.21 | 0.09 | <0.001 | 0.9 | <0.001 | <0.001 | <0.001 |
| **ASD** | 0.86 | 0.78 | 0.07 | 0.41 | <0.001 | 0.41 | 0.11 | <0.001 | 0.11 | <0.001 | <0.001 | <0.001 |
| **ADHD** | 0.78 | 0.54 | 0.24 | 0.03 | 0.03 | <0.001 | 0.43 | <0.001 | 0.43 | <0.001 | <0.001 | <0.001 |
| **Specific learning disorders** | 0.47 | 0.33 | 0.14 | <0.001 | <0.001 | <0.001 | 0.05 | 0.05 | <0.001 | <0.001 | <0.001 | <0.001 |
| **Motor disorders** | 0.36 | 0.18 | 0.18 | <0.001 | <0.001 | <0.001 | 0.37 | 0.18 | 0.18 | - | - | - |
| *Note.* H^2^= heritability; c^2^= shared environmental influences; e^2^= nonshared environmental influences; N= number of studies identified;  SE= standard error; I^2^_t_= total variance accounted for by heterogeneity; I^2^_b_= between-cluster heterogeneity; I^2^_w_= within-cluster heterogeneity. | | | | | | | | | | | | |

| **Supplementary Table 8.** Proportion of variance in genetic, shared and nonshared environmental correlations between NDDs accounted for by heterogeneity. | | | | | | | | | | | | |
| --- | --- | --- | --- | --- | --- | --- | --- | --- | --- | --- | --- | --- |
|  | **Family rA** | | | **Family rC** | | | **Family rE** | | | **SNP rG** | | |
| **NDDs** | **I^2^_t_** | **I^2^_b_** | **I^2^_w_** | **I^2^_t_** | **I^2^_b_** | **I^2^_w_** | **I^2^_t_** | **I^2^_b_** | **I^2^_w_** | **I^2^_t_** | **I^2^_b_** | **I^2^_w_** |
| **NDDs combined** | 0.89 | 0.34 | 0.55 | 0.95 | 0.36 | 0.59 | 0.24 | 0.24 | <0.001 | 0.49 | 0.33 | 0.16 |
| **ASD & ADHD** | 0.94 | 0.65 | 0.29 | - | - | - | 0.62 | 0.62 | <0.001 | 0.24 | <0.001 | 0.24 |
| **ADHD & motor disorders** | 0.99 | 0.49 | 0.49 | - | - | - | - | - | - | - | - | - |
| **ADHD & specific learning disorders** | 0.79 | 0.17 | 0.62 | 0.53 | 0.06 | 0.47 | <0.001 | <0.001 | <0.001 | - | - | - |
| **Communication disorders & motor disorders** | <0.001 | <0.001 | <0.001 | - | - | - | - | - | - | - | - | - |
| **Communication disorders & specific learning disorders** | <0.001 | <0.001 | <0.001 | - | - | - | - | - | - | - | - | - |
| *Note.* rA= genetic correlation; rC= shared environmental correlation; rE= nonshared environmental correlation; N= number of studies identified; SE= standard error; I^2^_t_= total variance accounted for by heterogeneity; I^2^_b_= between-cluster heterogeneity; I^2^_w_= within-cluster heterogeneity. | | | | | | | | | | | | |

| **Supplementary Table 9.** Proportion of variance in genetic, shared and nonshared environmental correlations between NDDs and DICCs accounted for by heterogeneity. | | | | | | | | | |
| --- | --- | --- | --- | --- | --- | --- | --- | --- | --- |
|  | **Family rA** | | | **Family rC** | | | **Family rE** | | |
| **NDDs and DICCs** | **I^2^_t_** | **I^2^_b_** | **I^2^_w_** | **I^2^_t_** | **I^2^_b_** | **I^2^_w_** | **I^2^_t_** | **I^2^_b_** | **I^2^_w_** |
| **NDDs and DICCs combined** | 0.93 | 0.55 | 0.38 | 95 | 0 | 95 | 91 | 0 | 91 |
| **ADHD & conduct disorder** | 0.93 | 0.46 | 0.46 | 96 | 48 | 48 | <0.001 | <0.001 | <0.001 |
| **ADHD & oppositional defiant disorder** | 0.83 | 0.42 | 0.42 | 94 | 47 | 47 | 93 | 46 | 46 |
| **ASD & conduct disorder** | <0.001 | <0.001 | <0.001 | 67 | <0.001 | 67 | <0.001 | <0.001 | <0.001 |
| *Note.* rA= genetic correlation; rC= shared environmental correlation; rE= nonshared environmental correlation; N= number of studies identified; SE= standard error; I^2^_t_= total variance accounted for by heterogeneity; I^2^_b_= between-cluster heterogeneity; I^2^_w_= within-cluster heterogeneity. | | | | | | | | | |

| **Supplementary Table 10**. Results of Egger’s regression for studies addressing heritability and environmental influences on NDDs. | | | | | | | | | | | | |
| --- | --- | --- | --- | --- | --- | --- | --- | --- | --- | --- | --- | --- |
|  | **Family h2** | | | **Family c2** | | | **Family e2** | | | **SNP h2** | | |
|  | **Z** | **P** | **Estimate (95% CIs)** | **Z** | **P** | **Estimate (95% CIs)** | **Z** | **P** | **Estimate (95% CIs)** | **Z** | **P** | **Estimate (95% CIs)** |
| **NDDs combined** | 0.0 | <0.001 | 0.73 (0.69-0.78) | 3.82 | <0.001 | 0.03 (-0.04-0.1) | 3.76 | <0.001 | 0.17 (0.11-0.22) | 1.59 | 0.11 | 0.09 (-0.05-0.22) |
| **Communication disorders** | 0.71 | 0.48 | 0.43 (0.23-0.63) | -1.8 | 0.07 | 0.6 (0.33-0.88) | 1.62 | 0.1 | 0.05 (-0.14-0.25) | 1.62 | 0.1 | 0.05 (-0.14-0.25) |
| **ASD** | 0.14 | 0.89 | 0.68 (0.57-0.79) | 1.65 | 0.1 | -0.01 (-0.15-0.14) | 0.65 | 0.52 | 0.23 (0.13-0.33) | 1.49 | 0.14 | 0.01 (-0.18-0.2) |
| **ADHD** | -2.58 | 0.01 | 0.75 (0.69-0.81) | 1.83 | 0.07 | 0.01 (-0.09-0.11) | 3.43 | <0.001 | 0.17 (0.09-0.24) | -0.17 | 0.87 | 0.22 (0.01-0.42) |
| **Specific learning disorders** | -5.03 | <0.001 | 0.75 (0.69-0.81) | 1.52 | 0.13 | 0.08 (-0.06-0.22) | 1.62 | 0.1 | 0.16 (0.06-0.27) | -0.25 | 0.81 | 0.38 (-0.34-1.11) |
| **Motor disorders** | -1.19 | 0.23 | 0.83 (0.71-0.95) | 0.27 | 0.78 | 0.04 (-0.62-0.71) | 0.81 | 0.42 | 0.09 (-0.56-0.74) | - | - | - |
| *Note.* H^2^= heritability; c^2^= shared environmental influences; e^2^= nonshared environmental influences; N= number of studies identified;  CIs= confidence intervals; Estimate= the limit estimate; -= number of parameters to be estimated was larger than the number of observations; Z= z-value of the test statistic; P= p-value. | | | | | | | | | | | | |

| **Supplementary Table 11**. Results of Egger’s regression for studies addressing genetic and environmental overlap between NDDs. | | | | | | | | | | | | |
| --- | --- | --- | --- | --- | --- | --- | --- | --- | --- | --- | --- | --- |
|  | **Family rA** | | | **Family rC** | | | **Family rE** | | | **SNP rG** | | |
|  | **Z** | **P** | **Estimate (95% CIs)** | **Z** | **P** | **Estimate (95% CIs)** | **Z** | **P** | **Estimate (95% CIs)** | **Z** | **P** | **Estimate (95% CIs)** |
| **NDDs combined** | -0.97 | 0.33 | 0.42 (0.16-0.68) | 1.84 | 0.07 | 0.09 (-0.36-0.54) | 1.65 | 0.1 | <0.001 (-0.2-0.2) | 1.07 | 0.28 | -0.38 (-1.61-0.85) |
| **ASD & ADHD** | -0.49 | 0.62 | 0.68 (-0.03-1.39) | - | - | - | 0.73 | 0.47 | 0.01 (-0.5-0.52) | 0.47 | 0.64 | -0.14 (-1.71-1.44) |
| **ADHD & specific learning disorders** | -0.02 | 0.99 | 0.08 (-0.31-0.47) | 1.17 | 0.24 | -0.02 (-0.46-0.42) | 1.15 | 0.25 | -0.04 (-0.32-0.23) | - | - | - |
| *Note.* rA= genetic correlation; rC= shared environmental correlation; rE= nonshared environmental correlation; N= number of studies identified; CIs= confidence intervals; Estimate= the limit estimate; -= number of parameters to be estimated was larger than the number of observations; Z= z-value of the test statistic; P= p-value. | | | | | | | | | | | | |

| **Supplementary Table 12**. Results of Egger’s regression for studies addressing genetic and environmental overlap between NDDs and DICCs. | | | | | | | | | |
| --- | --- | --- | --- | --- | --- | --- | --- | --- | --- |
|  | **Family rA** | | | **Family rC** | | | **Family rE** | | |
|  | **Z** | **P** | **Estimate (95% CIs)** | **Z** | **P** | **Estimate (95% CIs)** | **Z** | **P** | **Estimate (95% CIs)** |
| **NDDs and DICCs combined** | -0.79 | 0.43 | 0.63 (0.26, 1) | 3.62 | <0.001 | -0.17 (-0.42, 0.07) | 0.78 | 0.44 | 0.12 (-0.11, 0.35) |
| **ADHD & conduct disorder** | 0.32 | 0.75 | 0.38 (-0.28, 1.04) | 2.88 | <0.001 | -0.43 (-0.95, 0.09) | 1.1 | 0.27 | -0.15 (-0.64, 0.34) |
| **ADHD & oppositional defiant disorder** | -0.66 | 0.51 | 0.73 (0.32, 1.14) | 1.46 | 0.14 | 0.06 (-0.78, 0.89) | -0.79 | 0.43 | 0.63 (0.14, 1.12) |
| **ASD & conduct disorder** | 0.52 | 0.60 | -0.06 (-1.61, 1.49) | 0.45 | 0.65 | -0.24 (-4.32, 3.84) | 0.85 | 0.40 | -0.16 (-0.71, 0.38) |
| *Note.* rA= genetic correlation; rC= shared environmental correlation; rE= nonshared environmental correlation; N= number of studies identified; CIs= confidence intervals; Estimate= the limit estimate; Z= z-value of the test statistic; P= p-value. | | | | | | | | | |

| **Supplementary Table 13**. Sex-specific heritability, shared and nonshared environmental influences on NDDs. | | | | | | | | | | | | | | | | |
| --- | --- | --- | --- | --- | --- | --- | --- | --- | --- | --- | --- | --- | --- | --- | --- | --- |
|  | **Males** | | **Females** | | **Males** | | **Females** | | **Males** | | **Females** | | **Males** | | **Females** | |
|  | **Family h2 (SE)** | **N** | **Family h2 (SE)** | **N** | **Family c2 (SE)** | **N** | **Family c2 (SE)** | **N** | **Family e2 (SE)** | **N** | **Family e2 (SE)** | **N** | **SNP h2 (SE)** | **N** | **SNP h2 (SE)** | **N** |
| **NDDs combined** | 0.65 (0.06) | 68 | 0.67 (0.06) | 67 | 0.35 (0.08) | 36 | 0.28 (0.08) | 34 | 0.31 (0.04) | 63 | 0.33 (0.04) | 61 | 0.19 (0.07) | 2 | 0.09 (0.10) | 2 |
| **Intellectual disabilities** | - | - | - | - | - | - | - | - | - | - | - | - | - | - | - | - |
| **Communication disorders** | 0.64 (0.33) | 4 | 0.67 (0.42) | 4 | 0.35 (0.14) | 3 | 0.35 (0.16) | 3 | 0.28 (0.14) | 4 | 0.29 (0.14) | 4 | - | - | - | - |
| **ASD** | 0.64 (0.16) | 21 | 0.68 (0.09) | 23 | 0.46 (0.20) | 12 | 0.30 (0.14) | 12 | 0.28 (0.06) | 19 | 0.24 (0.02) | 21 | - | - | - | - |
| **ADHD** | 0.68 (0.08) | 38 | 0.71 (0.08) | 38 | 0.38 (0.17) | 14 | 0.13 (0.07) | 12 | 0.32 (0.06) | 36 | 0.34 (0.06) | 35 | 0.20 (0.08) | 2 | 0.13 (0.11) | 2 |
| **Specific learning disorders** | 0.61 (0.08) | 9 | 0.61 (0.09) | 9 | 0.21 (0.07) | 8 | 0.18 (0.06) | 8 | 0.30 (0.07) | 8 | 0.34 (0.08) | 8 | - | - | - | - |
| **Motor disorders** | 0.59 (0.36) | 2 | 0.58 (0.34) | 2 | -. | - | -. | - | 0.24 (0.09) | 2 | 0.27 (0.08) | 2 | - | - | - | - |
| *Note.* H^2^= heritability; c^2^= shared environmental influences; e^2^= nonshared environmental influences; N= number of studies identified;  SE= standard error. | | | | | | | | | | | | | | | | |

| **Supplementary Table 14**. Sex-specific genetic, shared and nonshared environmental correlations between NDDs. | | | | | | | | | | | | |
| --- | --- | --- | --- | --- | --- | --- | --- | --- | --- | --- | --- | --- |
|  | **Males** | | **Females** | | **Males** | | **Females** | | **Males** | | **Females** | |
| **NDDs** | **Family rA (SE)** | **N** | **Family rA (SE)** | **N** | **Family rC (SE)** | **N** | **Family rC (SE)** | **N** | **Family rE (SE)** | **N** | **Family rE (SE)** | **N** |
| **NDDs combined** | 0.86 (0.58) | 4 | 0.25 (0.36) | 2 | - | - | - | - | 0.09 (0.08) | 3 | 0.10 (0.11) | 2 |
| **ASD & ADHD** | 0.79 (0.42) | 2 | - | - | - | - | - | - | 0.20 (0.14) | 2 | - | - |
| *Note.* rA= genetic correlation; rC= shared environmental correlation; rE= nonshared environmental correlation; N= number of studies identified; SE= standard error. | | | | | | | | | | | | |

| **Supplementary Table 15**. Sex-specific genetic, shared and nonshared environmental correlations between NDDs and DICCs. | | | | | | | | | | | | |
| --- | --- | --- | --- | --- | --- | --- | --- | --- | --- | --- | --- | --- |
|  | **Males** | | **Females** | | **Males** | | **Females** | | **Males** | | **Females** | |
| **NDDs and DICCs** | **Family rA (SE)** | **N** | **Family rA (SE)** | **N** | **Family rC (SE)** | **N** | **Family rC (SE)** | **N** | **Family rE (SE)** | **N** | **Family rE (SE)** | **N** |
| **NDDs and DICCs combined** | - | - | 0.75 (0.58) | 2 | - | - | - | - | - | - | 0.06 (0.12) | 2 |
| **ADHD & conduct disorder** | - | - | 0.75 (0.58) | 2 | - | - | - | - | - | - | 0.06 (0.12) | 2 |
| *Note.* rA= genetic correlation; rC= shared environmental correlation; rE= nonshared environmental correlation; N= number of studies identified; SE= standard error. | | | | | | | | | | | | |

| **Supplementary Table 16.** Heritability, shared and nonshared environmental influences on NDDs, stratified by age categories. | | | | | | | | |
| --- | --- | --- | --- | --- | --- | --- | --- | --- |
| **NDDs** | **Family h2 (SE)** | **N** | **Family c2 (SE)** | **N** | **Family e2 (SE)** | **N** | **SNP h2 (SE)** | **N** |
| **NDDs combined** | | | | | | | | |
| Childhood (4-7y) | 0.63 (0.03) | 54 | 0.21 (0.04) | 36 | 0.27 (0.03) | 51 | 0.24 (0.11) | 6 |
| Middle childhood (8-10y) | 0.68 (0.04) | 54 | 0.12 (0.03) | 33 | 0.25 (0.02) | 51 | 0.26 (0.08) | 7 |
| Adolescence (11-24y) | 0.62 (0.04) | 79 | 0.17 (0.03) | 47 | 0.35 (0.03) | 72 | 0.23 (0.07) | 13 |
| Childhood & middle childhood (4-10y) | 0.67 (0.06) | 14 | 0.33 (0.08) | 7 | 0.21 (0.05) | 11 | - | - |
| Childhood & adolescence (4-24y) | 0.72 (0.07) | 40 | 0.20 (0.05) | 19 | 0.20 (0.03) | 31 | 0.17 (0.03) | 11 |
| Middle childhood & adolescence (8-24y) | 0.69 (0.04) | 50 | 0.14 (0.04) | 19 | 0.28 (0.03) | 31 | - | - |
| **Communication disorders** | | | | | | | | |
| Childhood (4-7y) | 0.56 (0.08) | 15 | 0.41 (0.07) | 12 | 0.21 (0.05) | 14 | - | - |
| Adolescence (11-24y) | 0.45 (0.07) | 7 | 0.26 (0.08) | 5 | 0.27 (0.06) | 5 | 0.32 (0.16) | 3 |
| Childhood & middle childhood (4-10y) | 0.92 (0.75) | 2 | - | - | - | - | - | - |
| **ASD** | | | | | | | | |
| Childhood (4-7y) | 0.69 (0.16) | 3 | - | - | 0.31 (0.08) | 3 | - | - |
| Middle childhood (8-10y) | 0.88 (0.40) | 11 | 0.13 (0.07) | 5 | 0.22 (0.05) | 9 | 0.26 (0.12) | 4 |
| Adolescence (11-24y) | 0.61 (0.07) | 9 | 0.31 (0.17) | 4 | 0.28 (0.07) | 7 | 0.16 (0.09) | 7 |
| Childhood & adolescence (4-24y) | 0.79 (0.17) | 5 | 0.02 (0.05) | 3 | 0.21 (0.13) | 4 | 0.13 (0.05) | 7 |
| Middle childhood & adolescence (8-24y) | 0.75 (0.07) | 10 | 0.13 (0.08) | 3 | 0.29 (0.04) | 8 | - | - |
| **ADHD** | | | | | | | | |
| Childhood (4-7y) | 0.64 (0.05) | 21 | 0.07 (0.06) | 7 | 0.33 (0.04) | 19 | 0.10 (0.17) | 2 |
| Middle childhood (8-10y) | 0.65 (0.07) | 28 | 0.07 (0.04) | 12 | 0.30 (0.04) | 28 | 0.19 (0.12) | 3 |
| Adolescence (11-24y) | 0.64 (0.05) | 44 | 0.23 (0.08) | 17 | 0.37 (0.03) | 39 | 0.09 (0.13) | 3 |
| Childhood & middle childhood (4-10y) | 0.68 (0.10) | 7 | 0.39 (0.13) | 2 | 0.27 (0.07) | 6 | - | - |
| Childhood & adolescence (4-24y) | 0.73 (0.08) | 24 | 0.19 (0.06) | 10 | 0.20 (0.04) | 20 | 0.21 (0.05) | 7 |
| Middle childhood & adolescence (8-24y) | 0.73 (0.06) | 19 | 0.04 (0.07) | 4 | 0.30 (0.04) | 15 | - | - |
| **Specific learning disorders** | | | | | | | | |
| Childhood (4-7y) | 0.63 (0.05) | 18 | 0.18 (0.04) | 18 | 0.21 (0.03) | 18 | 0.29 (0.14) | 3 |
| Middle childhood (8-10y) | 0.62 (0.06) | 20 | 0.17 (0.04) | 18 | 0.26 (0.03) | 19 | - | - |
| Adolescence (11-24y) | 0.57 (0.03) | 33 | 0.17 (0.03) | 27 | 0.30 (0.03) | 29 | 0.31 (0.09) | 8 |
| Childhood & middle childhood (4-10y) | 0.59 (0.10) | 6 | 0.24 (0.13) | 5 | 0.24 (0.07) | 6 | - | - |
| Childhood & adolescence (4-24y) | 0.61 (0.10) | 11 | 0.22 (0.06) | 8 | 0.20 (0.05) | 8 | - | - |
| Middle childhood & adolescence (8-24y) | 0.65 (0.06) | 26 | 0.22 (0.06) | 13 | 0.18 (0.04) | 12 | - | - |
| **Motor disorders** | | | | | | | | |
| Childhood & adolescence (4-24y) | 0.73 (0.09) | 4 | 0.21 (0.15) | 2 | 0.20 (0.12) | 3 | - | - |
| *Note.* H^2^= heritability; c^2^= shared environmental influences; e^2^= nonshared environmental influences; N= number of studies identified;  SE= standard error. | | | | | | | | |

| **Supplementary Table 17.** Genetic, shared and nonshared environmental correlations between NDDs, stratified by age categories. | | | | | | | | |
| --- | --- | --- | --- | --- | --- | --- | --- | --- |
| **NDDs** | **Family rA (SE)** | **N** | **Family rC (SE)** | **N** | **Family rE (SE)** | **N** | **SNP rG (SE)** | **N** |
| **NDDs combined** | | | | | | | | |
| Adolescence (11-24y) | 0.40 (0.23) | 11 | 0.80 (0.37) | 8 | 0.18 (0.05) | 10 | 0.73 (0.29) | 2 |
| Childhood & middle childhood (4-10y) | -0.17 (0.30) | 4 | - | - | 0.12 (0.10) | 3 | - | - |
| Childhood & adolescence (4-24y) | 0.16 (0.13) | 8 | - | 3 | 0.04 (0.07) | 4 | - | - |
| **ASD & ADHD** | | | | | | | | |
| Adolescence (11-24y) | 0.66 (0.49) | 3 | 0.15 (0.07) | 3 | 0.15 (0.07) | 3 | - | - |
| **ADHD & specific learning disorders** | | | | | | | | |
| Adolescence (11-24y) | -0.12 (0.16) | 5 | 0.26 (0.11) | 4 | 0.12 (0.06) | 4 | - | - |
| Childhood & middle childhood (4-10y) | -0.12 (0.36) | 3 | - | - | 0.12 (0.10) | 3 | - | - |
| Childhood & adolescence (4-24y) | -0.07 (0.20) | 3 | - | - | 0.05 (0.09) | 2 | - | - |
| **Communication disorders & motor disorders** | | | | | | | | |
| Childhood & adolescence (4-24y) | 0.33 (0.16) | 2 | - | - | - | - | - | - |
| *Note.* rA= genetic correlation; rC= shared environmental correlation; rE= nonshared environmental correlation; N= number of studies identified; SE= standard error. | | | | | | | | |

| **Supplementary Table 18.** Genetic, shared and nonshared environmental correlations between NDDs and DICCs, stratified by age categories. | | | | | | |
| --- | --- | --- | --- | --- | --- | --- |
| **NDDs and DICCs** | **Family rA (SE)** | **N** | **Family rC (SE)** | **N** | **Family rE (SE)** | **N** |
| **NDDs and DICCs combined** | | | | | | |
| Adolescence (11-24y) | 0.73 (.29) | 3 | 0.70 (0.63) | 2 | 0.82 (0.64) | 2 |
| Childhood & adolescence (4-24y) | 0.83 (0.61) | 3 | 0.09 (0.56) | 2 | 0.27 (0.08) | 3 |
| **ADHD & conduct disorder** | | | | | | |
| Childhood & adolescence (4-24y) | 0.90 (0.81) | 2 | - | - | 0.15 (0.18) | 2 |
| *Note.* rA= genetic correlation; rC= shared environmental correlation; rE= nonshared environmental correlation; N= number of studies identified; SE= standard error. | | | | | | |

| **Supplementary Table 19.** Heritability, shared and nonshared environmental influences on NDDs, stratified by countries. | | | | | | | | |
| --- | --- | --- | --- | --- | --- | --- | --- | --- |
| **NDDs** | **Family h2 (SE)** | **N** | **Family c2 (SE)** | **N** | **Family e2 (SE)** | **N** | **SNP h2 (SE)** | **N** |
| **NDDs combined** | | | | | | | | |
| Australia | 0.76 (0.17) | 11 | 0.21 (0.07) | 9 | 0.17 (0.05) | 8 | - | - |
| Australia & United States & Norway & Sweden | 0.74 (0.13) | 2 | 0.05 (0.11) | 2 | 0.24 (0.09) | 2 | - | - |
| Canada | 0.43 (0.09) | 7 | 0.18 (0.09) | 6 | 0.38 (0.07) | 6 | - | - |
| China | 0.5 (0.15) | 4 | 0.3 (0.13) | 3 | 0.29 (0.12) | 4 | - | - |
| Netherlands | 0.52 (0.26) | 19 | 0.12 (0.12) | 5 | 0.37 (0.13) | 17 | 0.47 (0.22) | 3 |
| Norway | 0.53 (0.09) | 2 | 0.25 (0.23) | 2 | 0.28 (0.14) | 2 | - | - |
| Sweden | 0.74 (0.05) | 24 | 0.07 (0.04) | 9 | 0.28 (0.03) | 22 | - | - |
| United Kingdom | 0.7 (0.06) | 96 | 0.18 (0.02) | 53 | 0.27 (0.02) | 85 | 0.22 (0.06) | 14 |
| United States | 0.61 (0.04) | 77 | 0.22 (0.03) | 44 | 0.32 (0.04) | 53 | - | - |
| **Intellectual disabilities** | | | | | | | | |
| Sweden | 0.86 (0.44) | 2 | - | - | 0.1 (0.16) | 2 | - | - |
| **Communication disorders** | | | | | | | | |
| Canada | 0.32 (0.2) | 2 | 0.38 (0.18) | 2 | 0.35 (0.12) | 2 | - | - |
| Netherlands | 0.45 (0.19) | 2 | - | - | 0.3 (0.18) | 2 | - | - |
| United Kingdom | 0.77 (0.41) | 17 | 0.35 (0.07) | 11 | 0.2 (0.04) | 13 | 0.32 (0.14) | 4 |
| United States | 0.71 (0.38) | 2 | - | - | - | - | - | - |
| **ASD** | | | | | | | | |
| Netherlands | 0.5 (0.17) | 2 | - | - | 0.52 (0.16) | 2 | - | - |
| Sweden | 0.74 (0.05) | 10 | 0.09 (0.06) | 5 | 0.28 (0.04) | 9 | - | - |
| United Kingdom | 0.8 (0.24) | 20 | 0.19 (0.08) | 8 | 0.24 (0.04) | 15 | 0.18 (0.08) | 7 |
| United States | 0.8 (0.5) | 3 | - | - | - | - | - | - |
| **ADHD** | | | | | | | | |
| Australia | 0.83 (0.31) | 7 | 0.26 (0.11) | 6 | 0.11 (0.05) | 5 | - | - |
| Australia & United States & Norway & Sweden | 0.73 (0.14) | 2 | 0.03 (0.12) | 2 | 0.26 (0.1) | 2 | - | - |
| Canada | 0.45 (0.16) | 3 | - | - | 0.38 (0.19) | 2 | - | - |
| China | 0.49 (0.33) | 2 | 0.26 (0.17) | 2 | 0.31 (0.24) | 2 | - | - |
| Netherlands | 0.52 (0.27) | 15 | 0.05 (0.08) | 4 | 0.28 (0.03) | 12 | 0.42 (0.24) | 2 |
| Sweden | 0.75 (0.07) | 18 | 0.04 (0.06) | 6 | 0.27 (0.04) | 17 | - | - |
| United Kingdom | 0.71 (0.03) | 42 | 0.2 (0.11) | 14 | 0.29 (0.02) | 39 | 0.08 (0.11) | 4 |
| United States | 0.62 (0.06) | 30 | 0.12 (0.06) | 12 | 0.38 (0.05) | 25 | - | - |
| **Specific learning disorders** | | | | | | | | |
| Australia | 0.72 (0.11) | 5 | 0.09 (0.07) | 4 | 0.23 (0.06) | 4 | - | - |
| Canada | 0.53 (0.13) | 4 | 0.1 (0.11) | 4 | 0.39 (0.09) | 4 | - | - |
| Netherlands | 0.59 (0.19) | 2 | - | - | 0.33 (0.13) | 2 | - | - |
| United Kingdom | 0.59 (0.03) | 33 | 0.17 (0.03) | 26 | 0.29 (0.02) | 29 | 0.31 (0.08) | 8 |
| United States | 0.57 (0.05) | 47 | 0.24 (0.04) | 33 | 0.21 (0.03) | 30 | - | - |
| **Motor disorders** | | | | | | | | |
| Sweden | 0.69 (0.12) | 4 | 0.06 (0.17) | 2 | 0.36 (0.12) | 4 | - | - |
| *Note.* H^2^= heritability; c^2^= shared environmental influences; e^2^= nonshared environmental influences; N= number of studies identified;  SE= standard error. | | | | | | | | |

| **Supplementary Table 20.** Genetic, shared and nonshared environmental correlations between NDDs, stratified by countries. | | | | | | | | |
| --- | --- | --- | --- | --- | --- | --- | --- | --- |
| **NDDs** | **Family rA (SE)** | **N** | **Family rC (SE)** | **N** | **Family rE (SE)** | **N** | **SNP rG (SE)** | **N** |
| **NDDs combined** | | | | | | | | |
| Australia | 0.27 (0.08) | 2 | 0.1 (0.09) | 2 | 0.02 (0.08) | 2 | - | - |
| Canada | -0.44 (0.24) | 2 | 0.19 (0.2) | 2 | 0.16 (0.15) | 2 | - | - |
| Sweden | 0.8 (0.26) | 3 | - | - | 0.36 (0.12) | 2 | - | - |
| United Kingdom | 0.37 (0.1) | 18 | 0.91 (0.29) | 10 | 0.16 (0.04) | 14 | 0.74 (0.28) | 2 |
| United States | 0.44 (0.07) | 11 | 0.07 (0.2) | 2 | - | - | - | - |
| **ASD & ADHD** | | | | | | | | |
| Sweden | 0.8 (0.25) | 3 | - | - | 0.36 (0.12) | 2 | - | - |
| United Kingdom | 0.28 (0.09) | 3 | - | - | 0.1 (0.07) | 3 | - | - |
| **ADHD & specific learning disorders** | | | | | | | | |
| Canada | -0.44 (0.24) | 2 | 0.19 (0.2) | 2 | 0.16 (0.15) | 2 | - | - |
| United Kingdom | 0.06 (0.16) | 6 | 0.48 (0.2) | 3 | 0.13 (0.05) | 5 | - | - |
| United States | 0.39 (0.09) | 8 | - | - | - | - | - | - |
| **Communication disorders & specific learning disorders** | | | | | | | | |
| United Kingdom | 0.66 (0.15) | 2 | - | - | - | - | - | - |
| *Note.* rA= genetic correlation; rC= shared environmental correlation; rE= nonshared environmental correlation; N= number of studies identified; SE= standard error. | | | | | | | | |

| **Supplementary Table 21.** Genetic, shared and nonshared environmental correlations between NDDs and DICCs, stratified by countries. | | | | | | |
| --- | --- | --- | --- | --- | --- | --- |
| **NDDs and DICCs** | **Family rA (SE)** | **N** | **Family rC (SE)** | **N** | **Family rE (SE)** | **N** |
| **NDDs and DICCs combined** | | | | | | |
| Sweden | 0.68 (0.41) | 3 | 0.89 (0.55) | 2 | 0.68 (0.64) | 3 |
| United Kingdom | 0.58 (0.29) | 3 | 0.97 (0.57) | 3 | 0.49 (0.44) | 3 |
| United States | 0.42 (0.15) | 6 | 0.85 (0.55) | 5 | 0.24 (0.09) | 4 |
| **ADHD & conduct disorder** | | | | | | |
| United States | 0.41 (0.17) | 3 | 0.99 (0.28) | 2 | 0.12 (0.14) | 2 |
| **ADHD & oppositional defiant disorder** | | | | | | |
| United States | 0.59 (0.32) | 3 | 0.99 (0.57) | 2 | 0.25 (0.14) | 2 |
| **ASD & conduct disorder** | | | | | | |
| United Kingdom | 0.33 (0.13) | 2 | 0.93 (0.77) | 2 | 0.04 (0.08) | 2 |
| *Note.* rA= genetic correlation; rC= shared environmental correlation; rE= nonshared environmental correlation; N= number of studies identified; SE= standard error. | | | | | | |

| **Supplementary Table 22.** Heritability, shared and nonshared environmental influences on NDDs, stratified by the percentage of individuals of European ancestry. | | | | | | | | |
| --- | --- | --- | --- | --- | --- | --- | --- | --- |
| **NDDs** | **Family h2 (SE)** | **N** | **Family c2 (SE)** | **N** | **Family e2 (SE)** | **N** | **SNP h2 (SE)** | **N** |
| **NDDs combined** | | | | | | | | |
| Less than 50% | 0.46 (0.07) | 7 | 0.24 (0.08) | 6 | 0.43 (0.08) | 7 | - | - |
| 50-74% | 0.47 (0.08) | 12 | 0.24 (0.08) | 9 | 0.32 (0.13) | 9 | - | - |
| 75-99% | 0.71 (0.07) | 37 | 0.24 (0.06) | 15 | 0.25 (0.03) | 32 | - | - |
| 100% | 0.66 (0.06) | 41 | 0.19 (0.04) | 29 | 0.32 (0.05) | 40 | 0.19 (0.03) | 29 |
| **Communication disorders** | | | | | | | | |
| 75-99% | 0.59 (0.27) | 3 | 0.36 (0.15) | 3 | 0.16 (0.11) | 3 | - | - |
| 100% | 0.56 (0.09) | 11 | 0.33 (0.1) | 8 | 0.24 (0.06) | 10 | 0.32 (0.14) | 4 |
| **ASD** | | | | | | | | |
| 75-99% | 0.91 (0.57) | 9 | - | - | 0.29 (0.06) | 6 | - | - |
| **ADHD** | | | | | | | | |
| Less than 50% | 0.41 (0.12) | 3 | 0.17 (0.15) | 2 | 0.54 (0.09) | 3 | - | - |
| 50-74% | 0.49 (0.11) | 5 | 0.18 (0.13) | 3 | 0.35 (0.19) | 4 | - | - |
| 75-99% | 0.73 (0.06) | 20 | 0.17 (0.07) | 6 | 0.27 (0.04) | 19 | - | - |
| 100% | 0.67 (0.04) | 11 | 0.04 (0.09) | 3 | 0.39 (0.05) | 10 | 0.2 (0.04) | 14 |
| **Specific learning disorders** | | | | | | | | |
| Less than 50% | 0.54 (0.16) | 5 | 0.25 (0.09) | 5 | 0.28 (0.06) | 5 | - | - |
| 50-74% | 0.52 (0.1) | 7 | 0.24 (0.1) | 6 | 0.24 (0.06) | 6 | - | - |
| 75-99% | 0.55 (0.09) | 7 | 0.29 (0.12) | 6 | 0.19 (0.06) | 6 | - | - |
| 100% | 0.61 (0.04) | 22 | 0.16 (0.04) | 19 | 0.3 (0.07) | 21 | 0.3 (0.08) | 9 |
| **Motor disorders** | | | | | | | | |
| 100% | 0.8 (0.05) | 2 | - | - | 0.47 (0.27) | 2 | - | - |
| *Note.* H^2^= heritability; c^2^= shared environmental influences; e^2^= nonshared environmental influences; N= number of studies identified; SE= standard error. | | | | | | | | |

| **Supplementary Table 23.** Genetic, shared and nonshared environmental correlations between NDDs, stratified by the percentage of individuals of European ancestry. | | | | | | | | |
| --- | --- | --- | --- | --- | --- | --- | --- | --- |
| **NDDs** | **Family rA (SE)** | **N** | **Family rC (SE)** | **N** | **Family rE (SE)** | **N** | **SNP rG (SE)** | **N** |
| **NDDs combined** | | | | | | | | |
| 75-99% | 0.63 (0.44) | 2 | - | - | - | - | - | - |
| 100% | 0.54 (0.1) | 4 | 0.93 (0.18) | 2 | 0.24 (0.09) | 4 | 0.39 (0.19) | 6 |
| **ASD & ADHD** | | | | | | | | |
| 100% | - | - | - | - | - | - | 0.26 (0.14) | 5 |
| **ADHD & specific learning disorders** | | | | | | | | |
| 100% | 0.48 (0.13) | 2 | - | - | 0.26 (0.15) | 2 | - | - |
| *Note.* rA= genetic correlation; rC= shared environmental correlation; rE= nonshared environmental correlation; N= number of studies identified; SE= standard error. | | | | | | | | |

| **Supplementary Table 24.** Genetic, shared and nonshared environmental correlations between NDDs and DICCs, stratified by the percentage of individuals of European ancestry. | | | | | | |
| --- | --- | --- | --- | --- | --- | --- |
| **NDDs and DICCs** | **Family rA (SE)** | **N** | **Family rC (SE)** | **N** | **Family rE (SE)** | **N** |
| **NDDs and DICCs combined** | | | | | | |
| 75-99% | 0.57 (0.25) | 3 | 0.88 (0.87) | 2 | - | - |
| 100% | 0.71 (0.31) | 2 | 0.89 (0.85) | 2 | 0.74 (0.49) | 2 |
| **ADHD & conduct disorder** | | | | | | |
| 75-99% | 0.41 (0.22) | 2 | - | - | - | - |
| **ADHD & oppositional defiant disorder** | | | | | | |
| 75-99% | 0.61 (0.48) | 2 | - | - | - | - |
| *Note.* rA= genetic correlation; rC= shared environmental correlation; rE= nonshared environmental correlation; N= number of studies identified; SE= standard error. | | | | | | |

| **Supplementary Table 25.** Heritability, shared and nonshared environmental influences on NDDs, stratified by measurement scales. | | | | | | | | |
| --- | --- | --- | --- | --- | --- | --- | --- | --- |
| **NDDs** | **Family h2 (SE)** | **N** | **Family c2 (SE)** | **N** | **Family e2 (SE)** | **N** | **SNP h2 (SE)** | **N** |
| **NDDs combined** | | | | | | | | |
| Categorical | 0.77 (0.07) | 28 | 0.19 (0.08) | 12 | 0.28 (0.06) | 25 | 0.17 (0.03) | 12 |
| Continuous | 0.64 (0.03) | 215 | 0.16 (0.02) | 116 | 0.28 (0.01) | 175 | 0.25 (0.06) | 17 |
| **Intellectual disabilities** | | | | | | | | |
| Categorical | 0.86 (0.44) | 2 | - | - | 0.1 (0.16) | 2 | - | - |
| **Communication disorders** | | | | | | | | |
| Categorical | 0.67 (0.24) | 6 | 0.47 (0.12) | 4 | 0.13 (0.06) | 5 | - | - |
| Continuous | 0.65 (0.2) | 19 | 0.3 (0.06) | 12 | 0.25 (0.05) | 14 | 0.32 (0.14) | 4 |
| **ASD** | | | | | | | | |
| Categorical | 0.83 (0.08) | 11 | 0.03 (0.08) | 5 | 0.18 (0.06) | 9 | 0.13 (0.05) | 7 |
| Continuous | 0.72 (0.15) | 29 | 0.18 (0.07) | 9 | 0.27 (0.03) | 23 | 0.2 (0.08) | 8 |
| **ADHD** | | | | | | | | |
| Categorical | 0.79 (0.1) | 13 | 0.05 (0.08) | 5 | 0.26 (0.07) | 12 | 0.21 (0.04) | 8 |
| Continuous | 0.66 (0.04) | 109 | 0.11 (0.03) | 43 | 0.31 (0.02) | 96 | 0.16 (0.1) | 6 |
| **Specific learning disorders** | | | | | | | | |
| Continuous | 0.62 (0.04) | 89 | 0.19 (0.02) | 65 | 0.24 (0.02) | 67 | 0.31 (0.08) | 8 |
| **Motor disorders** | | | | | | | | |
| Categorical | 0.72 (0.08) | 5 | 0.13 (0.11) | 3 | 0.38 (0.12) | 6 | - | - |
| Continuous | 0.69 (0.2) | 3 | - | - | - | - | - | - |
| *Note.* H^2^= heritability; c^2^= shared environmental influences; e^2^= nonshared environmental influences; N= number of studies identified;  SE= standard error. | | | | | | | | |

| **Supplementary Table 26.** Genetic, shared and nonshared environmental correlations between NDDs, stratified by measurement scales. | | | | | | | | |
| --- | --- | --- | --- | --- | --- | --- | --- | --- |
| **NDDs co-occurrences** | **Family rA (SE)** | **N** | **Family rC (SE)** | **N** | **Family rE (SE)** | **N** | **SNP rG (SE)** | **N** |
| **NDDs combined** | | | | | | | | |
| Categorical | 0.56 (0.32) | 3 | - | - | - | - | - | - |
| Continuous | 0.31 (0.12) | 34 | 0.67 (0.33) | 15 | 0.18 (0.05) | 21 | 0.74 (0.28) | 2 |
| **ASD & ADHD** | | | | | | | | |
| Continuous | 0.56 (0.34) | 5 | - | - | 0.22 (0.13) | 5 | - | - |
| **ADHD & motor disorders** | | | | | | | | |
| Categorical | 0.9 (0.82) | 2 | - | - | - | - | - | - |
| **ADHD & specific learning disorders** | | | | | | | | |
| Continuous | 0.06 (0.12) | 17 | 0.32 (0.14) | 7 | 0.11 (0.04) | 9 | - | - |
| **Communication disorders & specific learning disorders** | | | | | | | | |
| Continuous | 0.66 (0.15) | 2 | - | - | - | - | - | - |
| *Note.* rA= genetic correlation; rC= shared environmental correlation; rE= nonshared environmental correlation; N= number of studies identified; SE= standard error. | | | | | | | | |

| **Supplementary Table 27.** Genetic, shared and nonshared environmental correlations between NDDs and DICCs, stratified by measurement scales. | | | | | | |
| --- | --- | --- | --- | --- | --- | --- |
| **Co-occurences between NDDs and DICCs** | **Family rA (SE)** | **N** | **Family rC (SE)** | **N** | **Family rE (SE)** | **N** |
| **NDDs and DICCs combined** | | | | | | |
| Continuous | 0.62 (0.19) | 15 | 0.88 (0.34) | 11 | 0.38 (0.14) | 13 |
| **ADHD & conduct disorder** | | | | | | |
| Continuous | 0.66 (0.36) | 6 | 0.94 (0.71) | 3 | 0.11 (0.08) | 5 |
| **ADHD & oppositional defiant disorder** | | | | | | |
| Continuous | 0.66 (0.18) | 6 | 0.96 (0.57) | 4 | 0.54 (0.25) | 5 |
| **ASD & conduct disorder** | | | | | | |
| Continuous | 0.35 (0.10) | 3 | 0.88 (0.57) | 3 | 0.07 (0.08) | 3 |
| *Note.* rA= genetic correlation; rC= shared environmental correlation; rE= nonshared environmental correlation; N= number of studies identified; SE= standard error. | | | | | | |

| **Supplementary Table 28.** Overview of family-based studies using samples of males and females combined. Co-occurrences between disorders annotated with an asterisk (*) indicate pairs of disorders for which meta-analysis could not be performed. | | | |
| --- | --- | --- | --- |
| **Reference** | **Cohort** | **Age category** | **Country** |
| **Heritability and environmental influences on intellectual disabilities** | | | |
| Du Rietz et. al. (2021)^10^ | Medical Birth Register, Multi-Generation Register | Childhood & Adolescence | Sweden |
| Taylor et. al. (2019)^11^ | The Child and Adolescent Twin Study in Sweden (CATSS) | Middle Childhood & Adolescence | Sweden |
| **Heritability and environmental influences on communication disorders** | | | |
| Bishop & Hayiou-Thomas (2008)^12^ | Twins Early Development Study (TEDS) | Childhood | United Kingdom |
| Cheesman et. al. (2017)^13^ | Twins Early Development Study (TEDS) | Adolescence | United Kingdom |
| DeThorne et. al. (2006)^14^ | Western reserve twin project (WRTP) | Childhood | United States |
| Hayiou-Thomas, Dale & Plomin (2012)^15^ | Twins Early Development Study (TEDS) | Childhood & Middle Childhood | United Kingdom |
| Hayiou-Thomas, Dale & Plomin (2014)^16^ | Twins Early Development Study (TEDS) | Childhood | United Kingdom |
| Hohnen & Stevenson (1999)^17^ | Twin study in London | Childhood | United Kingdom |
| Tomblin & Buckwalter (1998)^18^ | Twin study in Iowa | Childhood | United States |
| Trzaskowski, Dale & Plomin (2013)^19^ | Twins Early Development Study (TEDS) | Adolescence | United Kingdom |
| van Beijsterveldt, Felsenfeld & Boomsma (2010)^20^ | Netherlands twin register (NTR) | Childhood | Netherlands |
| Bishop (2002)^21^ | Twin study in the United Kingdom | Childhood & Adolescence | United Kingdom |
| Bishop (2005)^22^ | Twins Early Development Study (TEDS) | Childhood | United Kingdom |
| Bishop, Adams & Norbury (2006)^23^ | Twins Early Development Study (TEDS) | Childhood | United Kingdom |
| Bishop, Laws, Adams & Norbury (2006)^24^ | Twins Early Development Study (TEDS) | Childhood | United Kingdom |
| Bishop, North & Donlan (1996)^25^ | Twin study in the United Kingdom | Childhood & Middle Childhood | United Kingdom |
| Dale, Rice, Rimfeld & Hayiou-Thomas (2018)^26^ | Twins Early Development Study (TEDS) | Adolescence | United Kingdom |
| Dionne et. al. (2011)^27^ | The Quebec Newborn Twin Study (QNTS) | Childhood | Canada |
| Dworzynski, Remington, Rijsdijk, Howell & Plomin (2007)^28^ | Twins Early Development Study (TEDS) | Childhood | United Kingdom |
| Hoekstra, Bartels, Van Leeuwen & Boomsma (2009)^29^ | Netherlands twin register (NTR) | Middle Childhood & Adolescence | Netherlands |
| Mimeau et. al. (2018)^30^ | The Quebec Newborn Twin Study (QNTS) | Childhood | Canada |
| Price, Dale & Plomin (2004)^31^ | Twins Early Development Study (TEDS) | Childhood | United Kingdom |
| Tosto et. al. (2017)^32^ | Twins Early Development Study (TEDS) | Childhood | United Kingdom |
| Trzaskowski et. al. (2013)^33^ | Twins Early Development Study (TEDS) | Adolescence | United Kingdom |
| Viding et. al. (2004)^34^ | Twins Early Development Study (TEDS) | Childhood | United Kingdom |
| **Heritability and environmental influences on ASD** | | | |
| Bailey et. al. (1995)^35^ | The twin study of Folstein & Rutter | Childhood & Adolescence | United Kingdom |
| Cheesman et. al. (2017)^13^ | Twins Early Development Study (TEDS) | Adolescence | United Kingdom |
| Deng et. al. (2015)^36^ | Twin study in China | Childhood & Adolescence | China |
| Du Rietz et. al. (2021)^10^ | Medical Birth Register, Multi-Generation Register | Childhood & Adolescence | Sweden |
| Dworzynski et. al. (2008)^37^ | Twins Early Development Study (TEDS) | Childhood & Middle Childhood | United Kingdom |
| Dworzynski, Happe, Bolton & Ronald (2009)^38^ | Twins Early Development Study (TEDS) | Middle Childhood & Adolescence | United Kingdom |
| Frazier et. al. (2014)^39^ | Interactive Autism Network (IAN) | Middle Childhood | United States |
| Hallet, Ronald & Happe (2009)^40^ | Twins Early Development Study (TEDS) | Middle Childhood | United Kingdom |
| Hoekstra, Bartels, Verweij & Boomsma (2007)^41^ | Netherlands twin register (NTR) | Adolescence | Netherlands |
| Jones et. al. (2009)^42^ | Twins Early Development Study (TEDS) | Middle Childhood | United Kingdom |
| Lichtenstein, Carlstrom, Rastam, Gillberg & Anckarsater (2010)^1^ | Swedish Twin Register | Middle Childhood | Sweden |
| Lundstrom et. al. (2012)^43^ | The Child and Adolescent Twin Study in Sweden (CATSS) | Middle Childhood & Adolescence | Sweden |
| Pinto, Rijsdijk, Ronald, Asherson & Kuntsi (2016)^44^ | Twins Early Development Study (TEDS) | Middle Childhood | United Kingdom |
| Polderman, Posthuma, De Sonnerville, Verlhulst & Boomsma (2006)^45^ | Netherlands twin register (NTR) | Childhood | Netherlands |
| Robinson et. al. (2011)^46^ | Twins Early Development Study (TEDS) | Adolescence | United Kingdom |
| Robinson et. al. (2012)^47^ | Twins Early Development Study (TEDS) | Middle Childhood & Adolescence | United Kingdom |
| Ronald et. al. (2006)^48^ | Twins Early Development Study (TEDS) | Middle Childhood | United Kingdom |
| Ronald, Happe, Price, Baron-Cohen & Plomin (2006)^49^ | Twins Early Development Study (TEDS) | Middle Childhood | United Kingdom |
| Ronald, Larsson, Anckarsater & Lichtenstein (2014)^50^ | The Child and Adolescent Twin Study in Sweden (CATSS) | Middle Childhood & Adolescence | Sweden |
| Ronald, Simonoff, Kuntsi, Asherson & Plomin (2008)^51^ | Twins Early Development Study (TEDS) | Middle Childhood | United Kingdom |
| Scherff et. al. (2014)^52^ | Twins Early Development Study (TEDS) | Adolescence | United Kingdom |
| Scourfield, Martin, Eley & McGuffin (2004)^53^ | The Cardiff Study of All Wales and Northwest of England Twins (CaStANET) | Childhood & Adolescence | United Kingdom |
| Taylor et. al. (2018)^54^ | The Child and Adolescent Twin Study in Sweden (CATSS) | Middle Childhood & Adolescence | Sweden |
| Taylor et. al. (2019)^11^ | The Child and Adolescent Twin Study in Sweden (CATSS) | Middle Childhood & Adolescence | Sweden |
| Taylor et. al. (2020)^55^ | The Child and Adolescent Twin Study in Sweden (CATSS) | Middle Childhood & Adolescence | Sweden |
| Taylor, Charman & Ronald (2015)^56^ | Twins Early Development Study (TEDS) | Adolescence | United Kingdom |
| Tick et. al. (2016)^57^ | Twins Early Development Study (TEDS) | Middle Childhood | United Kingdom |
| Towers et. al. (2000)^58^ | The Nonshared Environment in Adolescent Development (NEAD) | Adolescence | United States |
| Trzaskowski, Dale & Plomin (2013)^19^ | Twins Early Development Study (TEDS) | Adolescence | United Kingdom |
| Yip et. al. (2018)^59^ | Swedish Medical Register, Multi-Generation Register | Childhood | Sweden |
| Hallmayer et. al. (2011)^60^ | California Autism Twins Study | Adolescence | United States |
| Lundstrom et. al. (2011)^61^ | The Child and Adolescent Twin Study in Sweden (CATSS) | Middle Childhood & Adolescence | Sweden |
| Taniai et. al. (2008)^62^ | Nagoya North District Care Center for Disabled Children, Nagoya Child Welfare Center, and Nagoya West District Care Center for Disabled Children | Childhood & Adolescence | Japan |
| Lundstrom et. al. (2010)^63^ | The Child and Adolescent Twin Study in Sweden (CATSS) | Middle Childhood & Adolescence | Sweden |
| Colvert et. al. (2015)^64^ | Twins Early Development Study (TEDS) | Middle Childhood | United Kingdom |
| Ronald, Happe & Plomin (2005)^65^ | Twins Early Development Study (TEDS) | Childhood | United Kingdom |
| **Heritability and environmental influences on ADHD** | | | |
| Boomsma, Van Beijsterveldt, Odinstova, Neale & Dolan (2020)^66^ | The Young Netherlands Twin Register (YNTR) | Middle Childhood | Netherlands |
| Brikell et. al. (2016)^67^ | Twin Study of Child and Adolescent Development (TCHAD) | Middle Childhood | Sweden |
| Brooker et. al. (2020)^68^ | Wisconsin Twin Panel | Adolescence | United States |
| Burt, Krueger, McGue & Iacono (2001)^69^ | The Minnesota Twin Family Study (MTFS) | Middle Childhood & Adolescence | United States |
| Burt, Larsson, Lichtenstein & Klump (2012)^70^ | The Michigan State University Twin Registry | Childhood & Middle Childhood | United States |
| Chang, Lichtenstein & Larsson (2012)^71^ | Twin Study of Child and Adolescent Development (TCHAD) | Middle Childhood & Adolescence | Sweden |
| Chang, Lichtenstein, Asherson & Larsson (2013)^72^ | Twin Study of Child and Adolescent Development (TCHAD) | Middle Childhood | Sweden |
| Cheesman et. al. (2017)^13^ | Twins Early Development Study (TEDS) | Adolescence | United Kingdom |
| Chen et. al. (2016)^73^ | Chinese Child and Adolescent Twin Register | Childhood & Adolescence | China |
| Cheung, Fazier-Wood, Asherson, Rijsdijk & Kuntsi (2014)^74^ | Twins Early Development Study (TEDS) | Childhood & Middle Childhood | United Kingdom |
| Coolidge, Thede & Toung (2000)^75^ | Twin study in Colorado | Middle Childhood | United States |
| Curran et. al. (2003)^76^ | The Childhood Hyperactivity and Inattention Project (CHIP) | Childhood & Adolescence | United Kingdom |
| de Zeuw, van Beijsterveldt, Lubke, Glasner & Boomsma (2015)^77^ | Netherlands twin register (NTR) | Childhood | Netherlands |
| Derks et. al. (2008)^78^ | Netherlands twin register (NTR) | Childhood | Netherlands |
| Derks, Dolan, Hudziak, Neale & Boomsma (2007)^79^ | Netherlands twin register (NTR) | Childhood | Netherlands |
| Derks, Hudziak, van Beijsterveldts, Dolan & Boomsma (2006)^80^ | Netherlands twin register (NTR) | Childhood | Netherlands |
| Dick, Viken, Kaprio, Pulkkinen & Rose (2005)^81^ | The Finnish Twin Cohort Study | Adolescence | Finland |
| Dolan, De Zeeuw, Zayats, Van Beijsterveldt & Boomsma (2020)^82^ | Netherlands twin register (NTR) | Adolescence | Netherlands |
| Du Rietz et. al. (2021)^10^ | Medical Birth Register, Multi-Generation Register | Childhood & Adolescence | Sweden |
| Ebejer et. al. (2010)^83^ | Australian Twin Register, Colorado Birth Registry, and Medical Birth Registries in Norway and Sweden | Childhood | Australia, United States, Norway, Sweden |
| Ebejer et. al. (2015)^84^ | The Brisbane Longitudinal Twin Study | Middle Childhood & Adolescence | Australia |
| Edelbrock, Rende, Plomin & Thompson (1995)^85^ | Western reserve twin project (WRTP) | Childhood & Adolescence | United States |
| Gould, Coventry, Olson & Byrne (2018)^86^ | National Assessment Program in Numeracy and Literacy (NAPLAN) | Childhood & Adolescence | Australia |
| Greven, Asherson, Rijsdijk & Plomin (2011)^87^ | Twins Early Development Study (TEDS) | Middle Childhood | United Kingdom |
| Greven, Harlaar, Dale & Plomin (2011)^88^ | Twins Early Development Study (TEDS) | Adolescence | United Kingdom |
| Greven, Kovas, Willcutt, Petrill & Plomin (2014)^89^ | Twins Early Development Study (TEDS) | Adolescence | United Kingdom |
| Greven, Rijsdijk, Asherson & Plomin (2012)^90^ | Twins Early Development Study (TEDS) | Middle Childhood | United Kingdom |
| Greven, Rijsdijk, Plomin (2011)^91^ | Twins Early Development Study (TEDS) | Middle Childhood | United Kingdom |
| Hay, Bennett, Levy, Sergeant & Swanson (2007)^92^ | The Australian Twin ADHD Project (ATAP) | Childhood & Middle Childhood | Australia |
| Heutink, Verhuls & Boomsma (2006)^93^ | Netherlands twin register (NTR) | Childhood | Netherlands |
| Hudziak, Derks, Althoff, Rettew & Boomsma (2005)^94^ | Netherlands twin register (NTR) | Childhood | Netherlands |
| Hur (2014)^95^ | The South Korean Twin Registry (SKTR) | Childhood | South Korea |
| Jaffee, Hanscombe, Haworth, Davis & Plomin (2012)^96^ | Twins Early Development Study (TEDS) | Middle Childhood | United Kingdom |
| Johnson, McGue & Iacono (2005)^97^ | The Minnesota Twin Family Study (MTFS) | Adolescence | United States |
| Kan et. al. (2013)^98^ | Netherlands twin register (NTR) | Childhood & Middle Childhood | Netherlands |
| Kan, van Beijsterveldt, Bartels & Boomsma (2014)^99^ | Netherlands twin register (NTR) | Adolescence | Netherlands |
| Kuja-Halkola, Lichtenstein, D'Onforio & Larsson (2015)^100^ | Twin Study of Child and Adolescent Development (TCHAD) | Middle Childhood | Sweden |
| Kuntsi & Stevenson (2001)^101^ | Twin study in Southern England | Childhood & Adolescence | United Kingdom |
| Kuntsi et. al. (2014)^102^ | Twins Early Development Study (TEDS) | Middle Childhood | United Kingdom |
| Kuntsi, Gayan & Stevenson (2000)^103^ | Twins Early Development Study (TEDS) | Childhood & Adolescence | United Kingdom |
| Kuntsi, Rijsdijk, Ronald, Asherson & Plomin (2005)^104^ | Twins Early Development Study (TEDS) | Childhood | United Kingdom |
| Larsson, Anckarsater, Rastam, Chang & Lichtenstein (2012)^105^ | Swedish Twin Register | Middle Childhood & Adolescence | Sweden |
| Larsson, Dilshad, Lichtenstein & Barker (2011)^106^ | Twin Study of Child and Adolescent Development (TCHAD) | Middle Childhood & Adolescence | Sweden |
| Lemery-Chalfant, Doelger & Goldsmith (2008)^107^ | Wisconsin Twin Panel | Middle Childhood | United States |
| Levy, Hay, McStephen, Wood & Waldman (1997)^108^ | The Australian Twin ADHD Project (ATAP) | Childhood & Adolescence | Australia |
| Lewis & Plomin (2015)^109^ | Twins Early Development Study (TEDS) | Childhood | United Kingdom |
| Lewis, Haworth & Plomin (2014)^110^ | Twins Early Development Study (TEDS) | Adolescence | United Kingdom |
| Lichtenstein, Carlstrom, Rastam, Gillberg & Anckarsater (2010)^1^ | Swedish Twin Register | Middle Childhood | Sweden |
| Lifford, Harold & Thapar (2009)^111^ | The Cardiff Study of All Wales and Northwest of England Twins (CaStANET), South Wales Family Study (SWFS) | Adolescence | United Kingdom |
| Little, Hart, Schatschneider & Taylor (2016)^112^ | Florida Twin Project on Behavior and Environment (FTP-BE) | Adolescence | United States |
| LoParo & Waldman (2014)^113^ | Twin study in Georgia | Middle Childhood | United States |
| Martin, Piek & Hay (2006)^114^ | The Australian Twin ADHD Project (ATAP) | Childhood & Adolescence | Australia |
| McLoughlin, Ronald, Kuntsi, Asherson & Plomin (2007)^115^ | Twins Early Development Study (TEDS) | Childhood & Middle Childhood | United Kingdom |
| Merwood et. al. (2013)^116^ | Twins Early Development Study (TEDS) | Adolescence | United Kingdom |
| Michelini, Eley, Gregory & McAdams (2015)^117^ | The Genesis 12-19 (G1219) Study | Adolescence | United Kingdom |
| Mikolajewski, Allan, Hart, Lonigan & Taylor (2013)^118^ | The Florida Twin Project on Reading (FTP-R) | Childhood & Adolescence | United States |
| Molenaar, Midderldorp, van Beijsterveldt & Boomsma (2015)^119^ | Netherlands twin register (NTR) | Childhood | Netherlands |
| Moruzzi, Rijsdijk & Battaglia (2014)^120^ | Twin study in Italy | Middle Childhood & Adolescence | Italy |
| Nikolas, Klump & Burt (2015)^121^ | The Michigan State University Twin Registry (MSUTR) | Childhood & Adolescence | United States |
| Niv, Tuvblad, Raine, Wang & Baker (2012)^122^ | Southern California Twin Project | Adolescence | United States |
| Paloyelis, Rijsdijk, Wood, Asherson & Kuntsi (2010)^123^ | Twins Early Development Study (TEDS) | Middle Childhood | United Kingdom |
| Peng et. al. (2016)^124^ | Missouri Twin Study | Adolescence | United States |
| Pingault et. al. (2015)^125^ | Twins Early Development Study (TEDS) | Middle Childhood | United Kingdom |
| Pinto, Rijsdijk, Ronald, Asherson & Kuntsi (2016)^44^ | Twins Early Development Study (TEDS) | Middle Childhood | United Kingdom |
| Plourde et. al. (2015)^126^ | The Quebec Newborn Twin Study (QNTS) | Childhood & Middle Childhood | Canada |
| Plourde, Boivin, Brendgen, Vitaro & Dionne (2017)^127^ | The Quebec Newborn Twin Study (QNTS) | Adolescence | Canada |
| Polderman et. al. (2011)^128^ | Netherlands twin register (NTR) | Childhood | Netherlands |
| Polderman, Posthuma, De Sonnerville, Verlhulst & Boomsma (2006)^45^ | Netherlands twin register (NTR) | Childhood | Netherlands |
| Polderman, van Dongen & Boomsma (2011)^129^ | Netherlands twin register (NTR) | Adolescence | Netherlands |
| Price et. al. (2005)^130^ | Twins Early Development Study (TEDS) | Childhood | United Kingdom |
| Quinn et. al. (2016)^131^ | The Child and Adolescent Twin Study in Sweden (CATSS) | Middle Childhood & Adolescence | Sweden |
| Ronald, Larsson, Anckarsater & Lichtenstein (2014)^50^ | The Child and Adolescent Twin Study in Sweden (CATSS) | Middle Childhood & Adolescence | Sweden |
| Ronald, Simonoff, Kuntsi, Asherson & Plomin (2008)^51^ | Twins Early Development Study (TEDS) | Middle Childhood | United Kingdom |
| Rosenberg, Pennington, Willcut & Olson (2012)^132^ | Colorado Learning Disabilities Research Center | Middle Childhood & Adolescence | United States |
| Rydell, Taylor & Larsson (2017)^133^ | Preschool Twin Study in Sweden (PETSS) | Childhood | Sweden |
| Saudino & Plomin (2007)^134^ | Twins Early Development Study (TEDS) | Childhood | United Kingdom |
| Saunders et. al. (2019)^135^ | The Child and Adolescent Twin Study in Sweden (CATSS) | Middle Childhood & Adolescence | Sweden |
| Siebelink et. al. (2019)^136^ | Twins Early Development Study (TEDS) | Adolescence | United Kingdom |
| Simonoff et. al. (1998)^137^ | Virginia twin study of adolescent behavioral development (VTSABD) | Adolescence | United States |
| Stern et. al. (2020)^138^ | E-RISK | Childhood | United Kingdom |
| Stevenson (1992)^139^ | Twin study in London | Adolescence | United Kingdom |
| Stevenson, Pennington, Gilger, DeFries & Gillis (1993)^140^ | Twin study in London | Adolescence | United Kingdom |
| Taylor et. al. (2019)^11^ | The Child and Adolescent Twin Study in Sweden (CATSS) | Middle Childhood & Adolescence | Sweden |
| Taylor, Allan, Mikolajewski & Hart (2013)^141^ | The Florida Twin Project on Reading (FTP-R) | Childhood & Adolescence | United States |
| Taylor, Charman & Ronald (2015)^56^ | Twins Early Development Study (TEDS) | Adolescence | United Kingdom |
| Thapar, Hervas & McGuffin (1995)^142^ | The Cardiff Births Survey (CBS) | Middle Childhood & Adolescence | United Kingdom |
| Towers et. al. (2000)^58^ | The Nonshared Environment in Adolescent Development (NEAD) | Adolescence | United States |
| Trzaskowski, Dale & Plomin (2013)^33^ | Twins Early Development Study (TEDS) | Adolescence | United Kingdom |
| Tuvblad, Zheng, Raine & Baker (2009)^143^ | UoC Twin Study of Risk Factors for Antisocial Behavior | Middle Childhood | United States |
| Tye et. al. (2012)^144^ | Twins Early Development Study (TEDS), The Neurophysiological Study of Activity and Attention in Twins (NEAAT) | Middle Childhood & Adolescence | United Kingdom |
| Vendlinski et. al. (2014)^145^ | Wisconsin Twin Panel | Childhood | United States |
| Waszczuk, Zavos & Eley (2020)^146^ | Twins Early Development Study (TEDS) | Adolescence | United Kingdom |
| Willcutt et. al. (2007)^147^ | Colorado Twin Register, Autstralian Twin Register, Medical Birth Register | Childhood | Australia, United States, Norway, Sweden |
| Willcutt et. al. (2010)^148^ | Colorado Learning Disabilities Research Center | Middle Childhood & Adolescence | United States |
| Wood, Rijsdijk, Asherson & Kuntsi (2009)^149^ | Twins Early Development Study (TEDS) | Middle Childhood | United Kingdom |
| Wood, Rijsdijk, Asherson & Kuntsi (2011)^150^ | Twins Early Development Study (TEDS) | Middle Childhood | United Kingdom |
| Wood, Rijsdijk, Saudino, Asherson & Kuntsi (2008)^151^ | Twins Early Development Study (TEDS) | Middle Childhood | United Kingdom |
| Zheng, Pingault, Unger & Rijsdijk (2020)^152^ | Qingdao Twin Registry (QTR) | Adolescence | China |
| Zumberge, Baker & Manis (2007)^153^ | The Southern California Twin register | Middle Childhood | United States |
| Burt, McGue, Krueger & Iacono (2005)^154^ | The Minnesota Twin Family Study (MTFS) | Middle Childhood & Adolescence | United States |
| Chen et. al. (2017)^155^ | Medical Birth Register, The Swedish Twin Register, The Multi-Generation Register | Childhood & Adolescence | Sweden |
| Crosbie et. al. (2013)^156^ | Ontario Science Centre (OSC) | Childhood | Canada |
| Eilertsen et. al. (2018)^157^ | The Norwegian mother and child cohort study (MoBa) | Childhood | Norway |
| Fedko et. al. (2017)^158^ | Netherlands twin register (NTR) | Middle Childhood | Netherlands |
| Haberstick et. al. (2008)^159^ | National Longitudinal Study of Adolescent Health | Childhood & Adolescence | United States |
| Lundstrom et. al. (2011)^61^ | The Child and Adolescent Twin Study in Sweden (CATSS) | Middle Childhood & Adolescence | Sweden |
| Martin, Levy, Pieka & Hay (2006)^160^ | The Australian Twin ADHD Project (ATAP) | Childhood & Adolescence | Australia |
| Merwood, Asherson & Larsson (2013)^161^ | Twin Study of Child and Adolescent Development (TCHAD) | Adolescence | Sweden |
| Mogensen, Larsson, Lundholm & Almqvist (2011)^162^ | Twin Study of Child and Adolescent Development (TCHAD) | Adolescence | Sweden |
| Nadder, Silberg, Eaves, Maes & Meyer (1998)^163^ | Virginia twin study of adolescent behavioral development (VTSABD) | Childhood & Adolescence | United States |
| Rhee, Waldman, Hay & Levy (1999)^164^ | Australian Twin Register | Childhood & Adolescence | Australia |
| Rimfeld et. al. (2021)^165^ | Twins Early Development Study (TEDS) | Adolescence | United Kingdom |
| Singh & Waldman (2010)^166^ | Georgia Twin Register | Childhood & Adolescence | United States |
| Willcutt, Pennington & DeFries (2000)^167^ | Colorado Learning Disabilities Research Center | Middle Childhood & Adolescence | United States |
| Willcutt, Pennington, Olson & DeFries (2007)^168^ | Colorado Learning Disabilities Research Center | Middle Childhood & Adolescence | United States |
| Merwood et. al. (2014)^169^ | The Cardiff Study of All Wales and Northwest of England Twins (CaStANET) | Childhood & Adolescence | United Kingdom |
| Thapar, Harringnton, Ross & McGuffin (2000)^170^ | The Greater Manchester Twin Register | Childhood & Adolescence | United Kingdom |
| Ehringer, Rhee, Young, Corley & Hewitt (2006)^171^ | Colorado Twin Register | Adolescence | United States |
| Smith et. al. (2011)^172^ | Center for AntisocialDrug Dependence (CADD) | Adolescence | United States |
| Thapar, Harringnton & McGuffin (2001)^173^ | The Greater Manchester Twin Register | Childhood & Adolescence | United Kingdom |
| Martin, Scourfield & McGuffin (2002)^174^ | Twin study in South Wales | Childhood & Adolescence | United Kingdom |
| **Heritability and environmental influences on specific learning disorders** | | | |
| Alarcon, DeFries, Light & Pennington (1997)^175^ | Colorado Learning Disabilities Research Center | Middle Childhood & Adolescence | United States |
| Bishop (2001)^176^ | Local United Kingdom sample | Childhood & Adolescence | United Kingdom |
| Cheesman et. al. (2017)^13^ | Twins Early Development Study (TEDS) | Adolescence | United Kingdom |
| Cheung, Fazier-Wood, Asherson, Rijsdijk & Kuntsi (2014)^74^ | Twins Early Development Study (TEDS) | Childhood & Middle Childhood | United Kingdom |
| Davis et. al. (2001)^177^ | Colorado Twin Study of Reading Disability | Middle Childhood & Adolescence | United States |
| Davis et. al. (2008)^178^ | Twins Early Development Study (TEDS) | Middle Childhood | United Kingdom |
| Davis et. al. (2014)^179^ | Twins Early Development Study (TEDS), Avon Longitudinal Study of Parents and Children (ALSPAC) | Adolescence | United Kingdom |
| DeFries & Alarcon (1996)^180^ | Colorado Learning Disabilities Research Center | Middle Childhood & Adolescence | United States |
| DeFries, Knopik & Wadsworth (1999)^181^ | Colorado Learning Disabilities Research Center | Middle Childhood & Adolescence | United States |
| Ebejer et. al. (2010)^83^ | Australian Twin Register, Colorado Birth Registry, Medical Birth Registries in Norway and Sweden | Middle Childhood | Australia, United States, Norway, Sweden |
| Erbeli, Hart, Wagner & Taylor (2018)^182^ | The Florida Twin Project on Reading (FTP-R) | Childhood & Adolescence | United States |
| Erbeli, Hart & Taylor (2019)^183^ | Florida Twin Project on Behavior and Environment (FTP-BE) | Middle Childhood | United States |
| Gayan & Olson (2001)^184^ | Colorado Learning Disabilities Research Center | Middle Childhood & Adolescence | United States |
| Greven, Harlaar, Dale & Plomin (2011)^88^ | Twins Early Development Study (TEDS) | Adolescence | United Kingdom |
| Greven, Kovas, Willcutt, Petrill & Plomin (2014)^89^ | Twins Early Development Study (TEDS) | Adolescence | United Kingdom |
| Greven, Rijsdijk, Asherson & Plomin (2012)^90^ | Twins Early Development Study (TEDS) | Middle Childhood | United Kingdom |
| Harlaar, Kovas, Dale, Petrill & Plomin (2012)^185^ | Twins Early Development Study (TEDS) | Adolescence | United Kingdom |
| Harlaar, Trzaskowski, Dale & Plomin (2014)^186^ | Twins Early Development Study (TEDS) | Childhood | United Kingdom |
| Hart, Petrill, Thompson & Plomin (2009)^187^ | Western reserve twin project (WRTP) | Childhood & Middle Childhood | United States |
| Hensler, Schatschneider, Taylor & Wagner (2010)^188^ | The Florida Twin Project on Reading (FTP-R) | Childhood | United States |
| Hohnen & Stevenson (1999)^17^ | Twin study in London | Childhood | United Kingdom |
| Kovas et. al. (2007)^189^ | Twins Early Development Study (TEDS) | Childhood | United Kingdom |
| Little, Hart, Schatschneider & Taylor (2016)^112^ | Florida Twin Project on Behavior and Environment (FTP-BE) | Adolescence | United States |
| Marlow et. al. (2001)^190^ | Twin study in Reading | Childhood & Adolescence | United Kingdom |
| Newsome, Boisvert & Wright (2014)^191^ | The Early Childhood Longitudinal Study (ECLS) | Childhood | United States |
| Olson, Gillis, Rack, DeFries & Fulker (1991)^192^ | Colorado Reading Project | Childhood & Adolescence | United States |
| Paloyelis, Rijsdijk, Wood, Asherson & Kuntsi (2010)^123^ | Twins Early Development Study (TEDS) | Middle Childhood | United Kingdom |
| Petrill et. al. (2007)^193^ | Western reserve twin project (WRTP) | Childhood & Middle Childhood | United States |
| Plourde et. al. (2015)^126^ | The Quebec Newborn Twin Study (QNTS) | Childhood & Middle Childhood | Canada |
| Plourde, Boivin, Brendgen, Vitaro & Dionne (2017)^127^ | The Quebec Newborn Twin Study (QNTS) | Adolescence | Canada |
| Polderman et. al. (2011)^128^ | Netherlands twin register (NTR) | Middle Childhood | Netherlands |
| Rosenberg, Pennington, Willcut & Olson (2012)^132^ | Colorado Learning Disabilities Research Center | Middle Childhood & Adolescence | United States |
| Samuelsson et. al. (2007)^194^ | Colorado Twin Register | Childhood | Australia |
| Taylor et. al. (2019)^11^ | The Child and Adolescent Twin Study in Sweden (CATSS) | Middle Childhood & Adolescence | Sweden |
| Tosto et. al. (2014)^195^ | Twins Early Development Study (TEDS) | Adolescence | United Kingdom |
| Trzaskowski, Dale & Plomin (2013)^19^ | Twins Early Development Study (TEDS) | Adolescence | United Kingdom |
| Wadsworth, DeFries, Willcutt, Pennington & Olson (2015)^196^ | Colorado Learning Disabilities Research Center | Childhood & Adolescence | United States |
| Wadsworth, DeFries, Willcutt, Pennington & Olson (2016)^197^ | Longitudinal Twin Study of Early Reading Development | Middle Childhood & Adolescence | United States |
| Wadsworth, Olson & DeFries (2010)^198^ | Colorado Reading Project, Colorado Learning Disabilities Research Center | Middle Childhood & Adolescence | United States |
| Wadsworth, Olson, Penningtom & DeFries (2000)^199^ | Colorado Reading Project, Colorado Learning Disabilities Research Center | Middle Childhood & Adolescence | United States |
| Willcutt et. al. (2010)^148^ | Colorado Learning Disabilities Research Center | Middle Childhood & Adolescence | United States |
| Willcutt et. al. (2019)^200^ | Colorado Learning Disabilities Research Center | Middle Childhood & Adolescence | United States |
| Willcutt, Pennington & DeFries (2000)^201^ | Colorado Learning Disabilities Research Center | Middle Childhood & Adolescence | United States |
| Zumberge, Baker & Manis (2007) | The Southern California Twin register | Middle Childhood | United States |
| Astrom, Wadsworth, Olson, Willcutt & DeFries (2011)^202^ | Colorado Learning Disabilities Research Center | Middle Childhood | United States |
| Betjemann et. al. (2010)^203^ | Colorado Learning Disabilities Research Center | Middle Childhood & Adolescence | United States |
| Bishop, Adams & Norbury (2004)^204^ | Twins Early Development Study (TEDS) | Childhood | United Kingdom |
| Castles, Datta, Gayan & Olson (1999) | Colorado Learning Disabilities Research Center | Middle Childhood & Adolescence | United States |
| Christopher et. al. (2013)^206^ | International Longitudinal Twin Study (ILTS) | Childhood | United States |
| Daucourt, Haughbrook, Van Bergen & Hart (2020)^207^ | Florida Twin Project on Behavior and Environment (FTP-BE) | Childhood & Adolescence | United States |
| DeFries, Fulker & LaBuda (1987)^208^ | Colorado Reading Project | Adolescence | United States |
| Erbeli, Hart & Taylor (2018)^209^ | The Florida Twin Project on Reading (FTP-R) | Childhood | United States |
| Friend et. al. (2009)^210^ | Colorado Twin Register | Childhood | United States |
| Friend, DeFries, Wadsworth & Olson (2007)^211^ | Colorado Learning Disabilities Research Center | Middle Childhood | United States |
| Garon-Carrier et. al. (2017)^212^ | The Quebec Newborn Twin Study (QNTS) | Childhood | Canada |
| Gayan & Olson (2003)^213^ | Colorado Learning Disabilities Research Center | Middle Childhood & Adolescence | United States |
| Gillis, DeFries & Fulker (1992)^214^ | Colorado Reading Project | Childhood & Adolescence | United States |
| Grasby & Coventry (2016)^215^ | Australian Twin Register | Middle Childhood | Australia |
| Harlaar, Dale & Plomin (2007)^216^ | Twins Early Development Study (TEDS) | Childhood | United Kingdom |
| Hart et. al. (2013)^217^ | The Florida Twin Project on Reading (FTP-R) | Childhood | United States |
| Hawke, Stallings, Wadsworth & DeFries (2008)^218^ | Colorado Reading Project, Colorado Learning Disabilities Research Center | Middle Childhood & Adolescence | United States |
| Knopik et. al. (2002)^219^ | Colorado Learning Disabilities Research Center | Middle Childhood & Adolescence | United States |
| Knopik, Alarcon & DeFries (1997)^220^ | Colorado Learning Disabilities Research Center | Middle Childhood & Adolescence | United States |
| Kovas et. al. (2013)^221^ | Twins Early Development Study (TEDS) | Childhood | United Kingdom |
| Kovas, Haworth, Harlaar, Petrill, Dale & Plomin (2007)^222^ | Twins Early Development Study (TEDS) | Middle Childhood | United Kingdom |
| Lazaroo et. al. (2019)^223^ | Brisbane Longitudinal Twin Study | Adolescence | Australia |
| Logan et. al. (2013)^224^ | Western reserve twin project (WRTP) | Childhood & Adolescence | United States |
| Malanchini et. al. (2017)^225^ | Twins Early Development Study (TEDS) | Middle Childhood | United Kingdom |
| Malanchini et. al. (2020)^226^ | Twins Early Development Study (TEDS) | Adolescence | United Kingdom |
| Malanchini, Engelhardt, Grotzinger, Harden & Tucker-Drob (2019)^227^ | Texas Twin Project | Middle Childhood & Adolescence | United States |
| Martin, Levy, Pieka & Hay (2006)^160^ | The Australian Twin ADHD Project (ATAP) | Childhood & Adolescence | Australia |
| Oliver, Dale & Plomin (2007)^228^ | Twins Early Development Study (TEDS) | Childhood & Middle Childhood | United Kingdom |
| Petrill et. al. (2010)^229^ | Western reserve twin project (WRTP) | Childhood & Middle Childhood | United States |
| Rimfeld et. al. (2018)^230^ | Twins Early Development Study (TEDS) | Childhood | United Kingdom |
| Rimfeld et. al. (2019)^231^ | Twins Early Development Study (TEDS) | Childhood | United Kingdom |
| Rimfeld, Ayorech, Dale, Kovas & Plomin (2016)^232^ | Twins Early Development Study (TEDS) | Adolescence | United Kingdom |
| Rimfeld, Kovas, Dale & Plomin (2015)^233^ | Twins Early Development Study (TEDS) | Adolescence | United Kingdom |
| Shakeshaft et. al. (2013)^234^ | Twins Early Development Study (TEDS) | Adolescence | United Kingdom |
| Swagerman et. al. (2017)^235^ | Netherlands twin register (NTR) | Middle Childhood | Netherlands |
| Taylor & Schatschneider (2010)^236^ | The Florida Twin Project on Reading (FTP-R) | Childhood | United States |
| Taylor, Erbeli, Hart & Johnson (2020)^237^ | The Florida Twin Project on Reading (FTP-R) | Adolescence | United States |
| Tosto et. al. (2017)^32^ | Twins Early Development Study (TEDS) | Childhood | United Kingdom |
| Tosto et. al. (2019)^238^ | Western reserve twin project (WRTP) | Middle Childhood & Adolescence | United States |
| Tosto, Malykh, Voronin, Plomin & Kovas (2013)^239^ | Twins Early Development Study (TEDS) | Adolescence | United Kingdom |
| Trzaskowski et. al. (2013)^19^ | Twins Early Development Study (TEDS) | Adolescence | United Kingdom |
| Wadsworth, Olson, Willcutt & DeFries (2012)^240^ | Colorado Reading Project | Middle Childhood & Adolescence | United States |
| Willcutt, Pennington, Olson & DeFries (2007)^168^ | Colorado Learning Disabilities Research Center | Middle Childhood & Adolescence | United States |
| Wong, Chow, Ho, Waye & Bishop (2014)^241^ | Chinese Twin Study of Reading Development | Childhood & Adolescence | China |
| Keenan et. al. (2006)^242^ | Colorado Learning Disabilities Research Center | Middle Childhood & Adolescence | United States |
| **Heritability and environmental influences on motor disorders** | | | |
| Du Rietz et. al. (2021)^10^ | Medical Birth Register, Multi-Generation Register | Childhood & Adolescence | Sweden |
| Lichtenstein, Carlstrom, Rastam, Gillberg & Anckarsater (2010)^1^ | Swedish Twin Register | Middle Childhood | Sweden |
| Martin, Piek & Hay (2006)^114^ | The Australian Twin ADHD Project (ATAP) | Childhood & Adolescence | Australia |
| Molenaar, Midderldorp, van Beijsterveldt & Boomsma (2015)^119^ | Netherlands twin register (NTR) | Childhood | Netherlands |
| Taylor et. al. (2019)^11^ | The Child and Adolescent Twin Study in Sweden (CATSS) | Middle Childhood & Adolescence | Sweden |
| Bishop (2002)^21^ | Twin study in the United Kingdom | Childhood & Adolescence | United Kingdom |
| Mataix-Cols et. al. (2015)^243^ | Multi-Generation Register, National Patient Register | Childhood & Adolescence | Sweden |
| Fliers et. al. (2009)^244^ | International Multicenter ADHD Genetics Study | Adolescence | Netherlands |
| **Genetic and environmental overlap between ASD & ADHD** | | | |
| Lichtenstein, Carlstrom, Rastam, Gillberg & Anckarsater (2010)^1^ | Swedish Twin Register | Middle childhood & Adolescence | Sweden |
| Lundstrom et. al. (2011)^61^ | The Child and Adolescent Twin Study in Sweden (CATSS) | Middle childhood & Adolescence | Sweden |
| Pinto, Rijsdijk, Ronald, Asherson & Kuntsi (2016)^44^ | Twins Early Development Study (TEDS) | Middle childhood | United Kingdom |
| Ronald, Larsson, Anckarsater & Lichtenstein (2014)^50^ | The Child and Adolescent Twin Study in Sweden (CATSS) | Middle childhood | Sweden |
| Taylor et. al. (2013)^245^ | Twins Early Development Study (TEDS) | Middle childhood | United Kingdom |
| Taylor, Charman & Ronald (2015)^56^ | Twins Early Development Study (TEDS) | Adolescence | United Kingdom |
| **Genetic and environmental overlap between ADHD & motor disorders** | | | |
| Lichtenstein, Carlstrom, Rastam, Gillberg & Anckarsater (2010)^1^ | Swedish Twin Register | Middle childhood & Adolescence | Sweden |
| Martin, Piek & Hay (2006)^114^ | The Australian Twin ADHD Project (ATAP) | Childhood & Adolescence | Australia |
| **Genetic and environmental overlap between ADHD & specific learning disorders** | | | |
| Cheung, Fazier-Wood, Asherson, Rijsdijk & Kuntsi (2014)^74^ | Twins Early Development Study (TEDS) | Childhood & Middle Childhood | United Kingdom |
| Greven, Harlaar, Dale & Plomin (2011)^88^ | Twins Early Development Study (TEDS) | Adolescence | United Kingdom |
| Greven, Kovas, Willcutt, Petrill & Plomin (2014)^89^ | Twins Early Development Study (TEDS) | Adolescence | United Kingdom |
| Greven, Rijsdijk, Asherson & Plomin (2012)^90^ | Twins Early Development Study (TEDS) | Middle childhood | United Kingdom |
| Lichtenstein, Carlstrom, Rastam, Gillberg & Anckarsater (2010)^1^ | Swedish Twin Register | Middle childhood & Adolescence | Sweden |
| Light, Pennington, Gilger & DeFries (1995)^246^ | Colorado Reading Project | Middle childhood & Adolescence | United States |
| Paloyelis, Rijsdijk, Wood, Asherson & Kuntsi (2010)^123^ | Twins Early Development Study (TEDS) | Middle childhood | United Kingdom |
| Plourde et. al. (2015)^126^ | The Quebec Newborn Twin Study (QNTS) | Childhood & Middle Childhood | Canada |
| Plourde, Boivin, Brendgen, Vitaro & Dionne (2017)^127^ | The Quebec Newborn Twin Study (QNTS) | Adolescence | Canada |
| Polderman et. al. (2011)^128^ | Netherlands twin register (NTR) | Childhood & Middle Childhood | Netherlands |
| Rosenberg, Pennington, Willcut & Olson (2012)^132^ | Colorado Learning Disabilities Research Center | Middle childhood & Adolescence | United States |
| Stevenson, Pennington, Gilger, DeFries & Gillis (1993)^140^ | Twin study in London | Adolescence | United Kingdom |
| Wadsworth, DeFries, Willcutt, Pennington & Olson (2015)^196^ | Colorado Learning Disabilities Research Center | Childhood & Adolescence | United States |
| Wadsworth, DeFries, Willcutt, Pennington & Olson (2016)^197^ | Longitudinal Twin Study of Early Reading Development | Middle childhood & Adolescence | United States |
| Willcutt et. al. (2010)^148^ | Colorado Learning Disabilities Research Center | Middle childhood & Adolescence | United States |
| Willcutt, Pennington & DeFries (2000)^201^ | Colorado Learning Disabilities Research Center | Middle childhood & Adolescence | United States |
| Willcutt, Pennington, Olson & DeFries (2007)^168^ | Colorado Learning Disabilities Research Center | Middle childhood & Adolescence | United States |
| Martin, Levy, Pieka & Hay (2006)^160^ | The Australian Twin ADHD Project (ATAP) | Childhood & Adolescence | Australia |
| **Genetic and environmental overlap between ASD & communication disorders*** | | | |
| Dworzynski et. al. (2008)^37^ | Twins Early Development Study (TEDS) | Childhood & Middle Childhood | United Kingdom |
| **Genetic and environmental overlap between ASD & motor disorders*** | | | |
| Lichtenstein, Carlstrom, Rastam, Gillberg & Anckarsater (2010)^1^ | Swedish Twin Register | Middle childhood & Adolescence | Sweden |
| **Genetic and environmental overlap between ASD & specific learning disorders*** | | | |
| Lichtenstein, Carlstrom, Rastam, Gillberg & Anckarsater (2010)^1^ | Swedish Twin Register | Middle childhood & Adolescence | Sweden |
| **Genetic and environmental overlap between motor disorders & specific learning disorders*** | | | |
| Lichtenstein, Carlstrom, Rastam, Gillberg & Anckarsater (2010)^1^ | Swedish Twin Register | Middle childhood & Adolescence | Sweden |
| **Genetic and environmental overlap between communication disorders & motor disorders** | | | |
| Bishop (2002)^21^ | Twin study in the United Kingdom | Childhood & Adolescence | United Kingdom |
| Ooki (2005)^247^ | Twin study in Japan | Childhood & Adolescence | Japan |
| **Genetic and environmental overlap between communication disorders & specific learning disorders** | | | |
| Bishop (2001)^176^ | Local United Kingdom sample | Childhood & Adolescence | United Kingdom |
| Tosto et. al. (2017)^32^ | Twins Early Development Study (TEDS) | Childhood | United Kingdom |
| **Genetic and environmental overlap between subtypes of specific learning disorders** | | | |
| Davis et. al. (2008)^178^ | Twins Early Development Study (TEDS) | Middle childhood | United Kingdom |
| Davis et. al. (2014)^179^ | Twins Early Development Study (TEDS), Avon Longitudinal Study of Parents and Children (ALSPAC) | Adolescence | United Kingdom |
| Greven, Kovas, Willcutt, Petrill & Plomin (2014)^89^ | Twins Early Development Study (TEDS) | Adolescence | United Kingdom |
| Harlaar, Kovas, Dale, Petrill & Plomin (2012)^185^ | Twins Early Development Study (TEDS) | Adolescence | United Kingdom |
| Willcutt et. al. (2019)^200^ | Colorado Learning Disabilities Research Center | Middle childhood & Adolescence | United States |
| Gillis, DeFries & Fulker (1992)^214^ | Colorado Reading Project | Childhood & Adolescence | United States |
| Knopik, Alarcon & DeFries (1997)^220^ | Colorado Learning Disabilities Research Center | Middle childhood & Adolescence | United States |
| Kovas, Haworth, Harlaar, Petrill, Dale & Plomin (2007)^222^ | Twins Early Development Study (TEDS) | Middle childhood | United Kingdom |
| Oliver, Dale & Plomin (2007)^228^ | Twins Early Development Study (TEDS) | Childhood & Middle childhood | United Kingdom |
| **Genetic and environmental overlap between ADHD & conduct disorder** | | | |
| Burt, Krueger, McGue & Iacono (2001)^69^ | The Minnesota Twin Family Study (MTFS) | Middle childhood & Adolescence | United States |
| Dick, Viken, Kaprio, Pulkkinen & Rose (2005)^81^ | The Finnish Twin Cohort Study | Adolescence | Finland |
| Tuvblad, Zheng, Raine & Baker (2009)^143^ | The Southern California Twin register | Middle childhood | United States |
| Hur (2015)^248^ | The South Korean Twin Registry (SKTR) | Childhood & Adolescence | South Korea |
| Martin, Levy, Pieka & Hay (2006)^160^ | The Australian Twin ADHD Project (ATAP) | Childhood & Adolescence | Australia |
| Coolidge, Thede & Toung (2000)^75^ | Twin study in Colorado | Middle childhood | United States |
| **Genetic and environmental overlap between ADHD & oppositional defiant disorder** | | | |
| Burt, Krueger, McGue & Iacono (2001)^69^ | The Minnesota Twin Family Study (MTFS) | Middle childhood & Adolescence | United States |
| Dick, Viken, Kaprio, Pulkkinen & Rose (2005)^81^ | The Finnish Twin Cohort Study | Adolescence | Finland |
| Tuvblad, Zheng, Raine & Baker (2009)^143^ | The Southern California Twin register | Middle childhood | United States |
| Wood, Rijsdijk, Asherson & Kuntsi (2009)^149^ | Twins Early Development Study (TEDS) | Middle childhood | United Kingdom |
| Martin, Levy, Pieka & Hay (2006)^160^ | The Australian Twin ADHD Project (ATAP) | Childhood & Adolescence | Australia |
| Coolidge, Thede & Toung (2000)^75^ | Twin study in Colorado | Middle childhood | United States |
| **Genetic and environmental overlap between ASD & conduct disorder** | | | |
| Jones et. al. (2009)^42^ | Twins Early Development Study (TEDS) | Middle childhood | United Kingdom |
| O'Nions et. al. (2015)^249^ | Twins Early Development Study (TEDS) | Middle childhood | United Kingdom |
| **Genetic and environmental overlap between ASD & conduct disorder*** | | | |
| Lundstrom et. al. (2011)^61^ | The Child and Adolescent Twin Study in Sweden (CATSS) | Middle childhood & Adolescence | Sweden |
| **Genetic and environmental overlap between specific learning disorders & disruptive behaviour*** | | | |
| Newsome, Boisvert & Wright (2014)^191^ | The Early Childhood Longitudinal Study (ECLS) | Childhood | United States |

| **Supplementary Table 29.** Overview of family-based studies using male samples. Co-occurrences between disorders annotated with an asterisk (*) indicate pairs of disorders for which meta-analysis could not be performed. | | | |
| --- | --- | --- | --- |
| **Reference** | **Cohort** | **Age category** | **Country** |
| **Heritability and environmental influences on communication disorders** | | | |
| Spinath, Price, Dale & Plomin (2004)^250^ | Twins Early Development Study (TEDS) | Childhood | United Kingdom |
| Taylor et. al. (2014)^251^ | Twins Early Development Study (TEDS) | Adolescence | United Kingdom |
| Ooki (2005)^247^ | Twin study in Japan | Childhood & Adolescence | Japan |
| Viding et. al. (2004)^34^ | Twins Early Development Study (TEDS) | Childhood | United Kingdom |
| **Heritability and environmental influences on ASD** | | | |
| Cheesman et. al. (2017)^13^ | Twins Early Development Study (TEDS) | Adolescence | United Kingdom |
| Constantino & Todd (2003)^252^ | Missouri Twin Study | Adolescence | United States |
| Frazier et. al. (2014)^39^ | Interactive Autism Network (IAN) | Middle Childhood | United States |
| Hallett, Ronald, Rijsdijk & Happe (2012)^253^ | Twins Early Development Study (TEDS) | Childhood & Middle Childhood | United Kingdom |
| Hoekstra, Happe, Baron-Cohen & Ronald (2010)^254^ | Twins Early Development Study (TEDS) | Middle Childhood | United Kingdom |
| Holmboe et. al. (2014)^255^ | Twins Early Development Study (TEDS) | Middle Childhood | United Kingdom |
| Robinson et. al. (2011)^46^ | Twins Early Development Study (TEDS) | Adolescence | United Kingdom |
| Robinson et. al. (2012)^47^ | Twins Early Development Study (TEDS) | Middle Childhood & Adolescence | United Kingdom |
| Ronald et. al. (2006)^48^ | Twins Early Development Study (TEDS) | Middle Childhood | United Kingdom |
| Ronald, Larsson, Anckarsater & Lichtenstein (2014)^50^ | The Child and Adolescent Twin Study in Sweden (CATSS) | Middle Childhood | Sweden |
| Ronald, Simonoff, Kuntsi, Asherson & Plomin (2008)^51^ | Twins Early Development Study (TEDS) | Middle Childhood | United Kingdom |
| Scherff et. al. (2014)^52^ | Twins Early Development Study (TEDS) | Adolescence | United Kingdom |
| Taylor et. al. (2013)^245^ | Twins Early Development Study (TEDS) | Middle Childhood | United Kingdom |
| Taylor et. al. (2014)^251^ | Twins Early Development Study (TEDS) | Adolescence | United Kingdom |
| Taylor et. al. (2018)^54^ | The Child and Adolescent Twin Study in Sweden (CATSS) | Middle Childhood & Adolescence | Sweden |
| Taylor et. al. (2020)^55^ | The Child and Adolescent Twin Study in Sweden (CATSS) | Middle Childhood & Adolescence | Sweden |
| Taylor, Gillberg, Lichtenstein & Lundstrom (2017)^256^ | The Child and Adolescent Twin Study in Sweden (CATSS) | Middle Childhood & Adolescence | Sweden |
| Hallmayer et. al. (2011)^60^ | California Autism Twins Study | Adolescence | United States |
| Mazefsky et. al. (2008)^257^ | Autism Genetic Resource Exchange (AGRE) | Childhood & Adolescence | United States |
| Taniai et. al. (2008)^62^ | Nagoya North District Care Center for Disabled Children, Nagoya Child Welfare Center, and Nagoya West District Care Center for Disabled Children | Childhood & Adolescence | Japan |
| Ronald, Happe & Plomin (2005)^65^ | Twins Early Development Study (TEDS) | Childhood | United Kingdom |
| **Heritability and environmental influences on ADHD** | | | |
| Cheesman et. al. (2017)^13^ | Twins Early Development Study (TEDS) | Adolescence | United Kingdom |
| Cole, Ball, Martin, Scourfield & McGuffin (2009)^258^ | Cardiff Study of All Wales and North England Twins | Childhood & Adolescence | United Kingdom |
| Constantino, Hudziak & Todd (2003)^259^ | Missouri Twin Study | Childhood & Adolescence | United States |
| de Zeuw, van Beijsterveldt, Lubke, Glasner & Boomsma (2015)^77^ | Netherlands twin register (NTR) | Childhood | Netherlands |
| Dick, Viken, Kaprio, Pulkkinen & Rose (2005)^81^ | The Finnish Twin Cohort Study | Adolescence | Finland |
| Eaves et. al. (1997)^260^ | Virginia twin study of adolescent behavioral development (VTSABD) | Middle Childhood & Adolescence | United States |
| Eaves et. al. (2000)^261^ | Virginia twin study of adolescent behavioral development (VTSABD) | Middle Childhood & Adolescence | United States |
| Gregory, Eley, O'Connor & Plomin (2004)^262^ | Twins Early Development Study (TEDS) | Childhood | United Kingdom |
| Greven, Rijsdijk, Plomin (2011)^91^ | Twins Early Development Study (TEDS) | Middle Childhood | United Kingdom |
| Hudziak, Rudiger, Neale, Heath & Todd (2000)^263^ | Missouri Twin Study | Middle Childhood & Adolescence | United States |
| Jaffee, Hanscombe, Haworth, Davis & Plomin (2012)^96^ | Twins Early Development Study (TEDS) | Middle Childhood | United Kingdom |
| Kuntsi, Rijsdijk, Ronald, Asherson & Plomin (2005)^104^ | Twins Early Development Study (TEDS) | Middle Childhood | United Kingdom |
| Kuo, Lin, Yang, Soong & Chen (2004)^264^ | Twin study in Taipei City | Adolescence | Taiwan |
| Larsson, Lichtenstein & Larsson (2006)^265^ | Twin Study of Child and Adolescent Development (TCHAD) | Middle Childhood | Sweden |
| Lifford, Harold & Thapar (2009)^111^ | The Cardiff Study of All Wales and Northwest of England Twins (CaStANET), South Wales Family Study (SWFS) | Adolescence | United Kingdom |
| Ronald, Larsson, Anckarsater & Lichtenstein (2014)^50^ | The Child and Adolescent Twin Study in Sweden (CATSS) | Middle Childhood | Sweden |
| Ronald, Simonoff, Kuntsi, Asherson & Plomin (2008)^51^ | Twins Early Development Study (TEDS) | Middle Childhood | United Kingdom |
| Rydell, Taylor & Larsson (2017)^133^ | Preschool Twin Study in Sweden (PETSS) | Childhood | Sweden |
| Saudino & Plomin (2007)^134^ | Twins Early Development Study (TEDS) | Childhood | United Kingdom |
| Taylor et. al. (2013)^245^ | Twins Early Development Study (TEDS) | Middle Childhood | United Kingdom |
| van Beijsterveldt, Verhulst, Molenaar & Boomsma (2004)^266^ | Netherlands twin register (NTR) | Childhood | Netherlands |
| Vierikko, Pulkkinen, Kaprio & Rose (2004)^267^ | The Finnish Twin Cohort Study | Adolescence | Finland |
| Burt, McGue, Krueger & Iacono (2005)^154^ | The Minnesota Twin Family Study (MTFS) | Middle Childhood & Adolescence | United States |
| de Zeeuw, van Beijsterveldt, Ehli, de Geus & Boomsma (2017)^268^ | Netherlands twin register (NTR) | Childhood & Adolescence | Netherlands |
| Do et. al. (2019)^269^ | Add Health | Childhood & Adolescence | United States |
| Larsson, Larsson & Lichtenstein (2004)^270^ | Young Twins Study | Adolescence | Sweden |
| Nadder, Rutter, Silberg, Maes & Eaves (2002)^271^ | Virginia twin study of adolescent behavioral development (VTSABD) | Middle Childhood & Adolescence | United States |
| Nadder, Silberg, Eaves, Maes & Meyer (1998)^163^ | Virginia twin study of adolescent behavioral development (VTSABD) | Childhood & Adolescence | United States |
| Rietveld, Hudziak, Bartels, Van Beijsterveldt & Boomsma (2004)^272^ | Netherlands twin register (NTR) | Childhood | Netherlands |
| Saudino, Ronald & Plomin (2005)^273^ | Twins Early Development Study (TEDS) | Childhood | United Kingdom |
| Silberg et. al. (1996)^274^ | Virginia twin study of adolescent behavioral development (VTSABD) | Middle Childhood & Adolescence | United States |
| Sherman, Iacono & McGue (1997)^275^ | The Minnesota Twin Family Study (MTFS) | Adolescence | United States |
| Smith et. al. (2011)^172^ | Center for Antisocial Drug Dependence (CADD) | Adolescence | United States |
| **Heritability and environmental influences on specific learning disorders** | | | |
| Alarcon, DeFries & Fulker (1995)^276^ | Colorado Learning Disabilities Research Center | Middle Childhood & Adolescence | United States |
| Bates et. al. (2004)^277^ | Study of melanocytic naevi (moles) | Adolescence | Australia |
| Eaves et. al. (1997)^260^ | Virginia twin study of adolescent behavioral development (VTSABD) | Middle Childhood & Adolescence | United States |
| Harlaar, Spinath, Dale & Plomin (2005)^278^ | Twins Early Development Study (TEDS) | Childhood | United Kingdom |
| Reynolds et. al. (1996)^279^ | Virginia twin study of adolescent behavioral development (VTSABD) | Middle Childhood | United States |
| Tosto et. al. (2014)^195^ | Twins Early Development Study (TEDS) | Adolescence | United Kingdom |
| Grasby & Coventry (2016)^215^ | Australian Twin Register | Middle Childhood | Australia |
| Shakeshaft et. al. (2013)^234^ | Twins Early Development Study (TEDS) | Adolescence | United Kingdom |
| Tosto et. al. (2019)^238^ | Twins Early Development Study (TEDS) | Adolescence | United Kingdom |
| **Heritability and environmental influences on motor disorders** | | | |
| van Beijsterveldt, Verhulst, Molenaar & Boomsma (2004)^266^ | Netherlands twin register (NTR) | Childhood | Netherlands |
| Ooki (2005)^247^ | Twin study in Japan | Childhood & Adolescence | Japan |
| **Genetic and environmental overlap between ASD & ADHD** | | | |
| Constantino, Hudziak & Todd (2003)^259^ | Missouri Twin Study | United States | BEST |
| Ronald, Simonoff, Kuntsi, Asherson & Plomin (2008)^51^ | Twins Early Development Study (TEDS) | United Kingdom | BEST |
| **Genetic and environmental overlap between ADHD & conduct disorder*** | | | |
| Silberg et. al. (1996)^274^ | Virginia twin study of adolescent behavioral development (VTSABD) | Twin study | United States |

| **Supplementary Table 30.** Overview of family-based studies using female samples. Co-occurrences between disorders annotated with an asterisk (*) indicate pairs of disorders for which meta-analysis could not be performed. | | | |
| --- | --- | --- | --- |
| **Reference** | **Cohort** | **Age category** | **Country** |
| **Heritability and environmental influences on communication disorders** | | | |
| Spinath, Price, Dale & Plomin (2004)^250^ | Twins Early Development Study (TEDS) | Childhood | United Kingdom |
| Taylor et. al. (2014)^251^ | Twins Early Development Study (TEDS) | Adolescence | United Kingdom |
| Ooki (2005)^247^ | Twin study in Japan | Childhood & Adolescence | Japan |
| Viding et. al. (2004)^34^ | Twins Early Development Study (TEDS) | Childhood | United Kingdom |
| **Heritability and environmental influences on ASD** | | | |
| Cheesman et. al. (2017)^13^ | Twins Early Development Study (TEDS) | Adolescence | United Kingdom |
| Constantino & Todd (2003)^252^ | Missouri Twin Study | Adolescence | United States |
| Constantino, Hudziak & Todd (2003)^259^ | Missouri Twin Study | Childhood & Adolescence | United States |
| Frazier et. al. (2014)^39^ | Interactive Autism Network (IAN) | Middle Childhood | United States |
| Hallett, Ronald, Rijsdijk & Happe (2012)^253^ | Twins Early Development Study (TEDS) | Childhood & Middle Childhood | United Kingdom |
| Hoekstra, Happe, Baron-Cohen & Ronald (2010)^254^ | Twins Early Development Study (TEDS) | Middle Childhood | United Kingdom |
| Holmboe et. al. (2014)^255^ | Twins Early Development Study (TEDS) | Middle Childhood | United Kingdom |
| Lundstrom et. al. (2012)^43^ | The Child and Adolescent Twin Study in Sweden (CATSS) | Middle Childhood & Adolescence | Sweden |
| Robinson et. al. (2011)^46^ | Twins Early Development Study (TEDS) | Adolescence | United Kingdom |
| Robinson et. al. (2012)^47^ | Twins Early Development Study (TEDS) | Middle Childhood & Adolescence | United Kingdom |
| Ronald et. al. (2006)^48^ | Twins Early Development Study (TEDS) | Middle Childhood | United Kingdom |
| Ronald, Larsson, Anckarsater & Lichtenstein (2014)^50^ | The Child and Adolescent Twin Study in Sweden (CATSS) | Middle Childhood | Sweden |
| Ronald, Simonoff, Kuntsi, Asherson & Plomin (2008)^51^ | Twins Early Development Study (TEDS) | Middle Childhood | United Kingdom |
| Scherff et. al. (2014)^52^ | Twins Early Development Study (TEDS) | Adolescence | United Kingdom |
| Taylor et. al. (2013)^245^ | Twins Early Development Study (TEDS) | Middle Childhood | United Kingdom |
| Taylor et. al. (2014)^251^ | Twins Early Development Study (TEDS) | Adolescence | United Kingdom |
| Taylor et. al. (2018)^54^ | The Child and Adolescent Twin Study in Sweden (CATSS) | Middle Childhood & Adolescence | Sweden |
| Taylor et. al. (2020)^55^ | The Child and Adolescent Twin Study in Sweden (CATSS) | Middle Childhood & Adolescence | Sweden |
| Taylor, Gillberg, Lichtenstein & Lundstrom (2017)^256^ | The Child and Adolescent Twin Study in Sweden (CATSS) | Middle Childhood & Adolescence | Sweden |
| Hallmayer et. al. (2011)^60^ | California Autism Twins Study | Adolescence | United States |
| Mazefsky et. al. (2008)^257^ | Autism Genetic Resource Exchange (AGRE) | Childhood & Adolescence | United States |
| Taniai et. al. (2008)^62^ | Nagoya North District Care Center for Disabled Children, Nagoya Child Welfare Center, and Nagoya West District Care Center for Disabled Children | Childhood & Adolescence | Japan |
| Ronald, Happe & Plomin (2005)^65^ | Twins Early Development Study (TEDS) | Childhood | United Kingdom |
| **Heritability and environmental influences on ADHD** | | | |
| Cheesman et. al. (2017)^13^ | Twins Early Development Study (TEDS) | Adolescence | United Kingdom |
| Cole, Ball, Martin, Scourfield & McGuffin (2009)^258^ | Cardiff Study of All Wales and North England Twins | Childhood & Adolescence | United Kingdom |
| de Zeuw, van Beijsterveldt, Lubke, Glasner & Boomsma (2015)^77^ | Netherlands twin register (NTR) | Childhood | Netherlands |
| Dick, Viken, Kaprio, Pulkkinen & Rose (2005)^81^ | The Finnish Twin Cohort Study | Adolescence | Finland |
| Eaves et. al. (1997)^260^ | Virginia twin study of adolescent behavioral development (VTSABD) | Middle Childhood & Adolescence | United States |
| Eaves et. al. (2000)^261^ | Virginia twin study of adolescent behavioral development (VTSABD) | Middle Childhood & Adolescence | United States |
| Gregory, Eley, O'Connor & Plomin (2004)^262^ | Twins Early Development Study (TEDS) | Childhood | United Kingdom |
| Greven, Rijsdijk, Plomin (2011)^91^ | Twins Early Development Study (TEDS) | Middle Childhood | United Kingdom |
| Hudziak, Rudiger, Neale, Heath & Todd (2000)^263^ | Missouri Twin Study | Middle Childhood & Adolescence | United States |
| Jaffee, Hanscombe, Haworth, Davis & Plomin (2012)^96^ | Twins Early Development Study (TEDS) | Middle Childhood | United Kingdom |
| Kuntsi, Rijsdijk, Ronald, Asherson & Plomin (2005)^104^ | Twins Early Development Study (TEDS) | Middle Childhood | United Kingdom |
| Kuo, Lin, Yang, Soong & Chen (2004)^264^ | Twin study in Taipei City | Adolescence | Taiwan |
| Larsson, Lichtenstein & Larsson (2006)^265^ | Twin Study of Child and Adolescent Development (TCHAD) | Middle Childhood | Sweden |
| Lifford, Harold & Thapar (2009)^111^ | The Cardiff Study of All Wales and Northwest of England Twins (CaStANET), South Wales Family Study (SWFS) | Adolescence | United Kingdom |
| Ronald, Larsson, Anckarsater & Lichtenstein (2014)^50^ | The Child and Adolescent Twin Study in Sweden (CATSS) | Middle Childhood | Sweden |
| Ronald, Simonoff, Kuntsi, Asherson & Plomin (2008)^51^ | Twins Early Development Study (TEDS) | Middle Childhood | United Kingdom |
| Rydell, Taylor & Larsson (2017)^133^ | Preschool Twin Study in Sweden (PETSS) | Childhood | Sweden |
| Saudino & Plomin (2007)^134^ | Twins Early Development Study (TEDS) | Childhood | United Kingdom |
| Taylor et. al. (2013)^245^ | Twins Early Development Study (TEDS) | Middle Childhood | United Kingdom |
| van Beijsterveldt, Verhulst, Molenaar & Boomsma (2004)^266^ | Netherlands twin register (NTR) | Childhood | Netherlands |
| Vierikko, Pulkkinen, Kaprio & Rose (2004)^267^ | The Finnish Twin Cohort Study | Adolescence | Finland |
| Burt, McGue, Krueger & Iacono (2005)^154^ | The Minnesota Twin Family Study (MTFS) | Middle Childhood & Adolescence | United States |
| de Zeeuw, van Beijsterveldt, Ehli, de Geus & Boomsma (2017)^268^ | Netherlands twin register (NTR) | Childhood & Adolescence | Netherlands |
| Do et. al. (2019)^269^ | Add Health | Childhood & Adolescence | United States |
| Knopik, Heath, Bucholz, Madden & Waldron (2009) | Missouri Adolescent Female Twin Study cohort | Adolescence | United States |
| Larsson, Larsson & Lichtenstein (2004)^270^ | Young Twins Study | Adolescence | Sweden |
| Nadder, Rutter, Silberg, Maes & Eaves (2002)^271^ | Virginia twin study of adolescent behavioral development (VTSABD) | Middle Childhood & Adolescence | United States |
| Nadder, Silberg, Eaves, Maes & Meyer (1998)^163^ | Virginia twin study of adolescent behavioral development (VTSABD) | Childhood & Adolescence | United States |
| Neuman et. al. (2001)^281^ | Missouri Twin Study | Adolescence | United States |
| Rietveld, Hudziak, Bartels, Van Beijsterveldt & Boomsma (2004)^272^ | Netherlands twin register (NTR) | Childhood | Netherlands |
| Saudino, Ronald & Plomin (2005)^273^ | Twins Early Development Study (TEDS) | Childhood | United Kingdom |
| Silberg et. al. (1996)^274^ | Virginia twin study of adolescent behavioral development (VTSABD) | Middle Childhood & Adolescence | United States |
| Smith et. al. (2011)^172^ | Center for Antisocial Drug Dependence (CADD) | Adolescence | United States |
| **Heritability and environmental influences on specific learning disorders** | | | |
| Alarcon, DeFries & Fulker (1995)^276^ | Colorado Learning Disabilities Research Center | Middle Childhood & Adolescence | United States |
| Bates et. al. (2004)^277^ | Study of melanocytic naevi (moles) | Adolescence | Australia |
| Eaves et. al. (1997)^260^ | Virginia twin study of adolescent behavioral development (VTSABD) | Middle Childhood & Adolescence | United States |
| Harlaar, Spinath, Dale & Plomin (2005)^278^ | Twins Early Development Study (TEDS) | Childhood | United Kingdom |
| Reynolds et. al. (1996)^279^ | Virginia twin study of adolescent behavioral development (VTSABD) | Middle Childhood | United States |
| Tosto et. al. (2014)^195^ | Twins Early Development Study (TEDS) | Adolescence | United Kingdom |
| Grasby & Coventry (2016)^215^ | Australian Twin Register | Middle Childhood | Australia |
| Shakeshaft et. al. (2013)^234^ | Twins Early Development Study (TEDS) | Adolescence | United Kingdom |
| Tosto et. al. (2019)^238^ | Twins Early Development Study (TEDS) | Adolescence | United Kingdom |
| **Heritability and environmental influences on motor disorders** | | | |
| van Beijsterveldt, Verhulst, Molenaar & Boomsma (2004)^266^ | Netherlands twin register (NTR) | Childhood | Netherlands |
| Ooki (2005)^247^ | Twin study in Japan | Childhood & Adolescence | Japan |
| **Genetic and environmental overlap between ASD & ADHD*** | | | |
| Ronald, Simonoff, Kuntsi, Asherson & Plomin (2008)^51^ | Twins Early Development Study (TEDS) | Middle childhood | United Kingdom |
| **Genetic and environmental overlap between ADHD & conduct disorder** | | | |
| Silberg et. al. (1996)^274^ | Virginia twin study of adolescent behavioral development (VTSABD) | Middle childhood & Adolescence | United States |
| Knopik, Heath, Bucholz, Madden & Waldron (2009)^280^ | Missouri Adolescent Female Twin Study cohort | Adolescence | United States |

| **Supplementary Table 31.** Overview of SNP-based studies using samples of males and females combined. Disorders annotated with an asterisk (*) indicate disorders for which meta-analysis could not be performed. | | | |
| --- | --- | --- | --- |
| **Reference** | **Cohort** | **Age category** | **Country** |
| **Heritability and environmental influences on communication disorders** | | | |
| Cheesman et. al. (2017)^13^ | Twins Early Development Study (TEDS) | Adolescence | United Kingdom |
| Trzaskowski, Dale & Plomin (2013)^19^ | Twins Early Development Study (TEDS) | Adolescence | United Kingdom |
| Trzaskowski et. al. (2013)^33^ | Twins Early Development Study (TEDS) | Adolescence | United Kingdom |
| Verhoef, Shapland, Fisher, Dale & St Pourcain (2020)^282^ | Avon Longitudinal Study of Parents and Children (ALSPAC) | Middle Childhood | United Kingdom |
| **Heritability and environmental influences on ASD** | | | |
| Cheesman et. al. (2017)^13^ | Twins Early Development Study (TEDS) | Adolescence | United Kingdom |
| Gandal et. al. (2018)^283^ | Psychiatric Genomics Consortium (PGC), iPSYCH | Childhood & Adolescence | United Kingdom, Denmark |
| Grove et. al. (2019)^284^ | Psychiatric Genomics Consortium (PGC), iPSYCH | Childhood & Adolescence | United Kingdom, Denmark |
| Hill et. al. (2016)^285^ | Psychiatric Genomics Consortium (PGC) | Childhood & Adolescence | United Kingdom |
| Lee et. al. (2013)^286^ | Psychiatric Genomics Consortium (PGC) | Childhood & Adolescence | United Kingdom |
| Serdarevic et. al. (2020)^287^ | Generation R | Childhood | Netherlands |
| Solberg et. al. (2019)^288^ | Psychiatric Genomics Consortium (PGC), iPSYCH | Childhood & Adolescence | United Kingdom, Denmark |
| St Pourcain et. al. (2014)^289^ | Avon Longitudinal Study of Parents and Children (ALSPAC) | Middle Childhood | United Kingdom |
| St Pourcain et. al. (2018)^290^ | Avon Longitudinal Study of Parents and Children (ALSPAC) | Middle Childhood | United Kingdom |
| St Pourcain et. al. (2018) ^291^ | Avon Longitudinal Study of Parents and Children (ALSPAC) | Middle Childhood | United Kingdom |
| Stergiakouli et. al. (2017)^292^ | Avon Longitudinal Study of Parents and Children (ALSPAC) | Middle Childhood | United Kingdom |
| Trzaskowski, Dale & Plomin (2013)^19^ | Avon Longitudinal Study of Parents and Children (ALSPAC) | Adolescence | United Kingdom |
| Warrier & Baron-Cohen (2018)^293^ | Avon Longitudinal Study of Parents and Children (ALSPAC) | Adolescence | United Kingdom |
| The Autism Spectrum Disorders Working Group of The Psychiatric Genomics Consortium (2017)^294^ | Psychiatric Genomics Consortium (PGC) | Childhood & Adolescence | United Kingdom |
| Pettersson et. al. (2019)^295^ | Psychiatric Genomics Consortium (PGC), iPSYCH | Childhood & Adolescence | United Kingdom, Denmark |
| **Heritability and environmental influences on ADHD** | | | |
| Artigas et. al. (2020)^296^ | Psychiatric Genomics Consortium (PGC), iPSYCH | Childhood & Adolescence | United Kingdom, Denmark |
| Cheesman et. al. (2017)^13^ | Twins Early Development Study (TEDS) | Adolescence | United Kingdom |
| Demontis et. al. (2019)^297^ | Psychiatric Genomics Consortium (PGC), iPSYCH | Childhood & Adolescence | United Kingdom, Denmark |
| Hill et. al. (2016)^285^ | Psychiatric Genomics Consortium (PGC) | Childhood & Adolescence | United Kingdom |
| Lee et. al. (2013)^286^ | Psychiatric Genomics Consortium (PGC) | Childhood & Adolescence | United Kingdom |
| Martin et. al. (2018)^298^ | Psychiatric Genomics Consortium (PGC), iPSYCH | Childhood & Adolescence | United Kingdom, Denmark |
| Micalizzi et. al. (2021)^299^ | Philadelphia Neurodevelopmental Cohort | Middle Childhood & Adolescence | United States |
| Middeldorp et. al. (2016)^300^ | Avon Longitudinal Study of Parents and Children (ALSPAC) | Childhood | United Kingdom |
| Pappa et. al. (2015)^301^ | Generation R, Netherlands twin register (NTR) | Childhood & Middle Childhood | Netherlands |
| Rovira et. al. (2020)^302^ | Psychiatric Genomics Consortium (PGC), iPSYCH, IMpACT | Middle Childhood | United Kingdom, Denmark, United States |
| Solberg et. al. (2019)^288^ | Psychiatric Genomics Consortium (PGC), iPSYCH | Childhood & Adolescence | United Kingdom, Denmark |
| Stergiakouli et. al. (2017)^292^ | Avon Longitudinal Study of Parents and Children (ALSPAC) | Childhood | United Kingdom |
| Trzaskowski, Dale & Plomin (2013)^19^ | Twins Early Development Study (TEDS) | Adolescence | United Kingdom |
| Pettersson et. al. (2019)^295^ | Psychiatric Genomics Consortium (PGC), iPSYCH | Childhood & Adolescence | United Kingdom, Denmark |
| **Heritability and environmental influences on specific learning disorders** | | | |
| Cheesman et. al. (2017)^13^ | Twins Early Development Study (TEDS) | Adolescence | United Kingdom |
| Davis et. al. (2014)^179^ | Twins Early Development Study (TEDS), Avon Longitudinal Study of Parents and Children (ALSPAC) | Adolescence | United Kingdom |
| Gialluisi et. al. (2020)^303^ | Study-specific multi-site cohort | Childhood & Adolescence | Multiple sites |
| Harlaar, Trzaskowski, Dale & Plomin (2014)^186^ | Twins Early Development Study (TEDS) | Childhood | United Kingdom |
| Trzaskowski, Dale & Plomin (2013)^19^ | Twins Early Development Study (TEDS) | Adolescence | United Kingdom |
| Rimfeld et. al. (2018)^230^ | Twins Early Development Study (TEDS) | Childhood | United Kingdom |
| Rimfeld, Kovas, Dale & Plomin (2015)^233^ | Twins Early Development Study (TEDS) | Adolescence | United Kingdom |
| Trzaskowski et. al. (2013)^33^ | Twins Early Development Study (TEDS) | Adolescence | United Kingdom |
| Verhoef, Shapland, Fisher, Dale & St Pourcain (2020)^282^ | Avon Longitudinal Study of Parents and Children (ALSPAC) | Childhood | United Kingdom |
| **Genetic and environmental overlap between ASD & ADHD** | | | |
| Demontis et. al. (2019)^297^ | Psychiatric Genomics Consortium (PGC), iPSYCH | Childhood & Adolescence | United Kingdom, Denmark |
| Grove et. al. (2019)^284^ | Psychiatric Genomics Consortium (PGC), iPSYCH | Childhood & Adolescence | United Kingdom, Denmark |
| Solberg et. al. (2019)^288^ | Psychiatric Genomics Consortium (PGC) | Childhood & Adolescence | United Kingdom |
| Stergiakouli et. al. (2017)^292^ | Avon Longitudinal Study of Parents and Children (ALSPAC) | Childhood & Middle Childhood | United Kingdom |
| Lee et. al. (2013)^286^ | Psychiatric Genomics Consortium (PGC) | Childhood & Adolescence | United Kingdom |

| **Supplementary Table 32.** Overview of SNP-based studies using male samples. Disorders annotated with an asterisk (*) indicate disorders for which meta-analysis could not be performed. | | | |
| --- | --- | --- | --- |
| **Reference** | **Cohort** | **Age category** | **Country** |
| **Heritability and environmental influences on ASD*** | | | |
| Martin et. al. (2021)^304^ | Psychiatric Genomics Consortium (PGC), iPSYCH | Childhood & Adolescence | United Kingdom, Denmark |
| **Heritability and environmental influences on ADHD** | | | |
| Martin et. al. (2018)^298^ | Psychiatric Genomics Consortium (PGC), iPSYCH | Childhood & Adolescence | United Kingdom, Denmark |
| Martin et. al. (2021)^304^ | Psychiatric Genomics Consortium (PGC), iPSYCH | Childhood & Adolescence | United Kingdom, Denmark |

| **Supplementary Table 33.** Overview of SNP-based studies using female samples. Disorders annotated with an asterisk (*) indicate disorders for which meta-analysis could not be performed. | | | |
| --- | --- | --- | --- |
| **Reference** | **Cohort** | **Age category** | **Country** |
| **Heritability and environmental influences on ASD*** | | | |
| Martin et. al. (2021)^304^ | Psychiatric Genomics Consortium (PGC), iPSYCH | Childhood & Adolescence | United Kingdom, Denmark |
| **Heritability and environmental influences on ADHD** | | | |
| Martin et. al. (2018)^298^ | Psychiatric Genomics Consortium (PGC), iPSYCH | Childhood & Adolescence | United Kingdom, Denmark |
| Martin et. al. (2021)^304^ | Psychiatric Genomics Consortium (PGC), iPSYCH | Childhood & Adolescence | United Kingdom, Denmark |

| **Supplementary Table 34.** Heritability, shared and nonshared environmental influences on NDDs, stratified by designs. | | | | | | | | | |
| --- | --- | --- | --- | --- | --- | --- | --- | --- | --- |
| **Family-based designs** | | | | | | | **SNP-based designs** | | |
| **NDDs** | **Family h2 (SE)** | **N** | **Family c2 (SE)** | **N** | **Family e2 (SE)** | **N** |  | **SNP h2 (SE)** | **N** |
| **NDDs combined** | | | | | | | | | |
| Categorical threshold sibling study | 0.67 (0.24) | 3 | - | - | 0.2 (0.11) | 2 | GCTA (REML) | 0.21 (0.05) | 19 |
| Categorical threshold twin and sibling study | 0.85 (0.19) | 2 | - | - | 0.37 (0.21) | 3 | LDSC | 0.17 (0.04) | 13 |
| DF extremes twin and sibling study | 0.83 (0.38) | 4 | 0.17 (0.13) | 4 | - | - |  |  |  |
| Classical twin and sibling study | 0.57 (0.09) | 8 | 0.08 (0.09) | 2 | 0.45 (0.08) | 8 |  |  |  |
| Categorical threshold twin study | 0.74 (0.07) | 23 | 0.25 (0.08) | 11 | 0.27 (0.07) | 21 |  |  |  |
| DF extremes twin study | 0.7 (0.11) | 57 | 0.19 (0.05) | 22 | 0.27 (0.05) | 20 |  |  |  |
| Classical twin study | 0.65 (0.03) | 157 | 0.15 (0.02) | 95 | 0.27 (0.01) | 151 |  |  |  |
| **Communication disorders** | | | | | | | | | |
| Categorical threshold twin study | 0.47 (0.1) | 5 | 0.47 (0.12) | 4 | 0.13 (0.06) | 5 | GCTA (REML) | 0.32 (0.14) | 4 |
| DF extremes twin study | 0.78 (0.41) | 8 | 0.31 (0.12) | 5 | 0.22 (0.09) | 5 | LDSC | - | - |
| Classical twin study | 0.56 (0.09) | 11 | 0.29 (0.07) | 7 | 0.25 (0.06) | 8 |  |  |  |
| **ASD** | | | | | | | | | |
| Categorical threshold twin study | 0.87 (0.11) | 7 | 0.09 (0.15) | 3 | 0.16 (0.07) | 6 | GCTA (REML) | 0.17 (0.07) | 9 |
| DF extremes twin study | 0.78 (0.36) | 11 | - | - | 0.33 (0.07) | 5 | LDSC | 0.13 (0.05) | 8 |
| Classical twin study | 0.68 (0.04) | 20 | 0.16 (0.07) | 8 | 0.26 (0.03) | 19 |  |  |  |
| **ADHD** | | | | | | | | | |
| Categorical threshold twin and sibling study | 0.84 (0.21) | 2 | - | - | 0.13 (0.09) | 2 | GCTA (REML) | 0.17 (0.06) | 8 |
| DF extremes twin and sibling study | 0.94 (0.46) | 2 | 0.06 (0.25) | 2 | - | - | LDSC | 0.22 (0.05) | 7 |
| Classical twin and sibling study | 0.56 (0.1) | 7 | 0.08 (0.09) | 2 | 0.45 (0.08) | 7 |  |  |  |
| Categorical threshold twin study | 0.76 (0.1) | 13 | 0.14 (0.09) | 5 | 0.28 (0.08) | 12 |  |  |  |
| DF extremes twin study | 0.75 (0.18) | 11 | 0.04 (0.08) | 3 | 0.36 (0.14) | 2 |  |  |  |
| Classical twin study | 0.67 (0.03) | 91 | 0.1 (0.03) | 38 | 0.29 (0.02) | 87 |  |  |  |
| **Specific learning disorders** | | | | | | | | | |
| DF extremes twin and sibling study | 0.5 (0.13) | 2 | 0.2 (0.15) | 2 | - | - | GCTA (REML) | 0.31 (0.08) | 8 |
| DF extremes twin study | 0.62 (0.06) | 30 | 0.21 (0.06) | 14 | 0.25 (0.06) | 9 | LDSC | - | - |
| Classical twin study | 0.62 (0.05) | 63 | 0.18 (0.02) | 55 | 0.25 (0.02) | 60 |  |  |  |
| **Motor disorders** | | | | | | | | | |
| Categorical threshold twin and sibling study | - | - | - | - | 0.64 (0.18) | 2 |  |  |  |
| Categorical threshold twin study | 0.71 (0.1) | 3 | 0.12 (0.12) | 2 | 0.25 (0.12) | 3 |  |  |  |
| Classical twin study | 0.71 (0.23) | 2 | - | - | - | - |  |  |  |
| *Note.* H^2^= heritability; c^2^= shared environmental influences; e^2^= nonshared environmental influences; N= number of studies identified;  SE= standard error; GCTA= genome-wide complex trait analysis; REML= restricted maximum likelihood; LDSC= linkage disequilibrium score regression. | | | | | | | | | |

| **Supplementary Table 35.** Genetic, shared and nonshared environmental correlations between NDDs, stratified by designs. | | | | | | | | | |
| --- | --- | --- | --- | --- | --- | --- | --- | --- | --- |
| **Family-based designs** | | | | | | | **SNP-based designs** | | |
| **NDDs** | **Family rA (SE)** | **N** | **Family rC (SE)** | **N** | **Family rE (SE)** | **N** |  | **SNP rG (SE)** | **N** |
| **NDDs combined** | | | | | | | | | |
| Categorical threshold twin study | 0.67 (0.49) | 2 | - | - | - | - | GCTA (REML) | 0.5 (0.36) | 3 |
| DF extremes twin study | 0.38 (0.08) | 15 | - | - | 0.13 (0.12) | 2 | LDSC | 0.26 (0.14) | 3 |
| Classical twin study | 0.31 (0.17) | 21 | 0.69 (0.37) | 15 | 0.18 (0.05) | 20 |  |  |  |
| **ASD & ADHD** | | | | | | | | | |
| Classical twin study | 0.56 (0.34) | 5 | - | - | 0.22 (0.13) | 5 | GCTA (REML) | 0.36 (0.49) | 2 |
|  |  |  |  |  |  |  | LDSC | 0.26 (0.14) | 3 |
| **ADHD & motor disorders** | | | | | | | | | |
| Categorical threshold twin study | 0.9 (0.82) | 2 | - | - | - | - |  | - | - |
| **ADHD & specific learning disorders** | | | | | | | | | |
| DF extremes twin study | 0.41 (0.09) | 9 | - | - | - | - |  | - | - |
| Classical twin study | -0.09 (0.12) | 9 | 0.32 (0.14) | 7 | 0.10 (0.05) | 8 |  | - | - |
| *Note.* rA= genetic correlation; rC= shared environmental correlation; rE= nonshared environmental correlation; N= number of studies identified; SE= standard error; GCTA= genome-wide complex trait analysis; REML= restricted maximum likelihood; LDSC= linkage disequilibrium score regression. | | | | | | | | | |

| **Supplementary Table 36.** Genetic, shared and nonshared environmental correlations between NDDs and DICCs, stratified by designs. | | | | | | |
| --- | --- | --- | --- | --- | --- | --- |
| **NDDs and DICCs** | **Family rA (SE)** | **N** | **Family rC (SE)** | **N** | **Family rE (SE)** | **N** |
| **NDDs and DICCs combined** | | | | | | |
| Classical twin study | 0.62 (0.19) | 15 | 0.88 (0.34) | 11 | 0.38 (0.14) | 13 |
| **ADHD & conduct disorder** | | | | | | |
| Classical twin study | 0.66 (0.36) | 6 | 0.94 (0.71) | 3 | 0.11 (0.08) | 5 |
| **ADHD & oppositional defiant disorder** | | | | | | |
| Classical twin study | 0.66 (0.18) | 6 | 0.96 (0.57) | 4 | 0.54 (0.25) | 5 |
| **ASD & conduct disorder** | | | | | | |
| Classical twin study | 0.35 (0.10) | 3 | 0.88 (0.57) | 3 | 0.07 (0.08) | 3 |
| *Note.* rA= genetic correlation; rC= shared environmental correlation; rE= nonshared environmental correlation; N= number of studies identified; SE= standard error; GCTA= genome-wide complex trait analysis; REML= restricted maximum likelihood; LDSC= linkage disequilibrium score regression. | | | | | | |

| **Supplementary Table 37.** Heritability, shared and nonshared environmental influences on NDDs, stratified by models. | | | | | | |
| --- | --- | --- | --- | --- | --- | --- |
| **NDDs** | **Family h2 (SE)** | **N** | **Family c2 (SE)** | **N** | **Family e2 (SE)** | **N** |
| **NDDs combined** | | | | | | |
| A only | 0.74 (0.16) | 11 | - | - | - | - |
| Best fitting | 0.7 (0.05) | 82 | - | - | 0.34 (0.02) | 81 |
| Full ACE | 0.61 (0.03) | 104 | 0.16 (0.02) | 104 | 0.22 (0.01) | 104 |
| DF extremes A only | 0.77 (0.16) | 31 | - | - | - | - |
| DF extremes best fitting | 0.72 (0.16) | 18 | 0.24 (0.07) | 12 | 0.33 (0.06) | 7 |
| DF extremes full ACE | 0.6 (0.07) | 15 | 0.17 (0.05) | 15 | 0.24 (0.05) | 15 |
| Twin correlations | 0.67 (0.07) | 14 | 0.17 (0.07) | 4 | 0.37 (0.06) | 13 |
| **Communication disorders** | | | | | | |
| A only | 0.55 (0.2) | 3 | - | - | - | - |
| Best fitting | 0.57 (0.22) | 4 | - | - | 0.51 (0.19) | 4 |
| Full ACE | 0.47 (0.06) | 11 | 0.37 (0.07) | 11 | 0.19 (0.04) | 11 |
| DF extremes A only | 0.94 (0.56) | 4 | - | - | - | - |
| DF extremes best fitting | 0.55 (0.2) | 3 | 0.45 (0.26) | 2 | - | - |
| DF extremes full ACE | 0.47 (0.13) | 5 | 0.3 (0.11) | 5 | 0.23 (0.09) | 5 |
| **ASD** | | | | | | |
| A only | 0.83 (0.38) | 4 | - | - | - | - |
| Best fitting | 0.71 (0.09) | 13 | - | - | 0.28 (0.04) | 13 |
| Full ACE | 0.72 (0.1) | 11 | 0.11 (0.06) | 11 | 0.21 (0.05) | 11 |
| DF extremes A only | 0.86 (0.45) | 3 | - | - | - | - |
| DF extremes best fitting | 0.67 (0.07) | 6 | - | - | 0.33 (0.07) | 5 |
| Twin correlations | 0.7 (0.07) | 4 | 0.16 (0.08) | 2 | 0.25 (0.11) | 4 |
| **ADHD** | | | | | | |
| A only | 0.7 (0.21) | 4 | - | - | - | - |
| Best fitting | 0.7 (0.05) | 61 | - | - | 0.33 (0.02) | 59 |
| Full ACE | 0.65 (0.04) | 43 | 0.1 (0.02) | 43 | 0.24 (0.02) | 43 |
| DF extremes A only | 0.79 (0.28) | 9 | - | - | - | - |
| DF extremes best fitting | 0.88 (0.24) | 4 | 0.08 (0.2) | 3 | - | - |
| Twin correlations | 0.67 (0.1) | 11 | 0.2 (0.13) | 2 | 0.38 (0.07) | 10 |
| **Specific learning disorders** | | | | | | |
| A only | 0.58 (0.09) | 4 | - | - | - | - |
| Best fitting | 0.73 (0.17) | 9 | - | - | 0.34 (0.11) | 9 |
| Full ACE | 0.6 (0.05) | 54 | 0.18 (0.02) | 54 | 0.24 (0.02) | 54 |
| DF extremes A only | 0.64 (0.09) | 16 | - | - | - | - |
| DF extremes best fitting | 0.55 (0.08) | 7 | 0.22 (0.08) | 7 | - | - |
| DF extremes full ACE | 0.63 (0.08) | 9 | 0.18 (0.07) | 9 | 0.24 (0.06) | 9 |
| **Motor disorders** | | | | | | |
| Best fitting | 0.77 (0.18) | 3 | - | - | 0.39 (0.14) | 4 |
| Full ACE | 0.69 (0.1) | 3 | 0.13 (0.11) | 3 | 0.24 (0.13) | 3 |
| *Note.* H^2^= heritability; c^2^= shared environmental influences; e^2^= nonshared environmental influences; N= number of studies identified;  SE= standard error. | | | | | | |

| **Supplementary Table 38.** Genetic, shared and nonshared environmental correlations between NDDs, stratified by models. | | | | | | |
| --- | --- | --- | --- | --- | --- | --- |
| **NDDs** | **Family rA (SE)** | **N** | **Family rC (SE)** | **N** | **Family rE (SE)** | **N** |
| **NDDs combined** | | | | | | |
| A only | 0.68 (0.48) | 2 | - | - | - | - |
| Best fitting | 0.31 (0.24) | 8 | - | - | 0.14 (0.05) | 7 |
| Full ACE | 0.31 (0.13) | 16 | 0.67 (0.39) | 15 | 0.18 (0.06) | 16 |
| DF extremes A only | 0.37 (0.09) | 13 | - | - | - | - |
| **ASD & ADHD** | | | | | | |
| Best fitting | 0.68 (0.49) | 3 | - | - | 0.18 (0.09) | 3 |
| Full ACE | 0.42 (0.17) | 2 | - | - | 0.31 (0.21) | 2 |
| **ADHD & specific learning disorders** | | | | | | |
| Best fitting | 0.14 (0.16) | 5 | - | - | 0.11 (0.08) | 4 |
| Full ACE | -0.18 (0.21) | 6 | 0.31 (0.15) | 6 | 0.1 (0.05) | 6 |
| DF extremes A only | 0.38 (0.11) | 8 | - | - | - | - |
| *Note.* rA= genetic correlation; rC= shared environmental correlation; rE= nonshared environmental correlation; N= number of studies identified; SE= standard error. | | | | | | |

| **Supplementary Table 39.** Genetic, shared and nonshared environmental correlations between NDDs and DICCs, stratified by models. | | | | | | |
| --- | --- | --- | --- | --- | --- | --- |
| **NDDs and DICCs** | **Family rA (SE)** | **N** | **Family rC (SE)** | **N** | **Family rE (SE)** | **N** |
| **NDDs and DICCs combined** | | | | | | |
| Best fitting | 0.69 (0.3) | 7 | - | - | 0.15 (0.07) | 5 |
| Full ACE | 0.48 (0.14) | 10 | 0.9 (0.35) | 10 | 0.42 (0.18) | 10 |
| **ADHD & conduct disorder** | | | | | | |
| Best fitting | 0.78 (0.5) | 4 | - | - | 0.14 (0.13) | 3 |
| Full ACE | 0.33 (0.12) | 3 | 0.94 (0.71) | 3 | 0.07 (0.1) | 3 |
| **ADHD & oppositional defiant disorder** | | | | | | |
| Best fitting | 0.69 (0.24) | 3 | - | - | 0.42 (0.13) | 2 |
| Full ACE | 0.56 (0.24) | 4 | 0.96 (0.57) | 4 | 0.54 (0.3) | 4 |
| **ASD & conduct disorder** | | | | | | |
| Full ACE | 0.35 (0.11) | 3 | 0.88 (0.57) | 3 | 0.06 (0.08) | 3 |
| *Note.* rA= genetic correlation; rC= shared environmental correlation; rE= nonshared environmental correlation; N= number of studies identified; SE= standard error. | | | | | | |

| **Supplementary Table 40.** Heritability, shared and nonshared environmental influences on NDDs, stratified by raters. | | | | | | | | |
| --- | --- | --- | --- | --- | --- | --- | --- | --- |
| **NDDs** | **Family h2 (SE)** | **N** | **Family c2 (SE)** | **N** | **Family e2 (SE)** | **N** | **SNP h2 (SE)** | **N** |
| **NDDs combined** | | | | | | | | |
| Diagnosis | 0.81 (0.15) | 7 | 0.02 (0.09) | 2 | 0.3 (0.11) | 6 | 0.17 (0.04) | 11 |
| Parent | 0.7 (0.04) | 110 | 0.15 (0.03) | 48 | 0.25 (0.02) | 93 | 0.19 (0.07) | 10 |
| Parent & Self | 0.72 (0.1) | 8 | 0.09 (0.15) | 2 | 0.31 (0.06) | 8 | - | - |
| Parent & Teacher | 0.72 (0.06) | 17 | 0.04 (0.08) | 5 | 0.3 (0.04) | 14 | - | - |
| Researcher | 0.71 (0.18) | 2 | 0.02 (0.05) | 2 | 0.18 (0.16) | 2 | - | - |
| Self-report | 0.5 (0.07) | 19 | 0.12 (0.11) | 5 | 0.55 (0.05) | 17 | 0.05 (0.18) | 2 |
| Teacher | 0.65 (0.03) | 29 | 0.18 (0.07) | 12 | 0.34 (0.05) | 28 | 0.3 (0.19) | 5 |
| Cognitive test | 0.6 (0.04) | 98 | 0.21 (0.02) | 71 | 0.25 (0.02) | 73 | 0.29 (0.07) | 10 |
| **Intellectual disabilities** | | | | | | | | |
| Diagnosis | 0.86 (0.44) | 2 | - | - | 0.1 (0.16) | 2 | - | - |
| **Communication disorders** | | | | | | | | |
| Parent | 0.76 (0.22) | 7 | 0.43 (0.14) | 4 | 0.14 (0.06) | 6 | - | - |
| Teacher | 0.62 (0.11) | 2 | - | - | 0.17 (0.08) | 2 | - | - |
| Cognitive test | 0.6 (0.21) | 18 | 0.31 (0.06) | 12 | 0.25 (0.05) | 13 | 0.32 (0.14) | 4 |
| **ASD** | | | | | | | | |
| Diagnosis | 0.85 (0.15) | 4 | 0.01 (0.1) | 2 | 0.19 (0.11) | 3 | 0.12 (0.05) | 6 |
| Parent | 0.78 (0.21) | 27 | 0.19 (0.07) | 11 | 0.24 (0.03) | 20 | 0.2 (0.07) | 8 |
| Parent & Teacher | 0.63 (0.11) | 3 | - | - | 0.41 (0.12) | 3 | - | - |
| Self-report | 0.52 (0.12) | 2 | - | - | - | - | - | - |
| Teacher | 0.58 (0.07) | 6 | 0.04 (0.1) | 2 | 0.42 (0.07) | 5 | 0 (0.21) | 2 |
| **ADHD** | | | | | | | | |
| Diagnosis | 0.79 (0.24) | 4 | - | - | 0.29 (0.16) | 4 | 0.21 (0.05) | 7 |
| Parent | 0.7 (0.04) | 83 | 0.09 (0.03) | 34 | 0.23 (0.02) | 72 | 0.13 (0.1) | 5 |
| Parent & Self | 0.72 (0.1) | 8 | 0.09 (0.15) | 2 | 0.31 (0.06) | 8 | - | - |
| Parent & Teacher | 0.71 (0.05) | 15 | 0.04 (0.08) | 5 | 0.29 (0.05) | 12 | - | - |
| Self-report | 0.5 (0.08) | 18 | 0.12 (0.11) | 5 | 0.56 (0.05) | 16 | 0.02 (0.18) | 2 |
| Teacher | 0.65 (0.05) | 18 | 0.16 (0.11) | 5 | 0.37 (0.04) | 17 | 0.38 (0.23) | 3 |
| **Specific learning disorders** | | | | | | | | |
| Parent | 0.72 (0.25) | 2 | - | - | 0.23 (0.08) | 2 | - | - |
| Teacher | 0.67 (0.05) | 5 | 0.16 (0.06) | 4 | 0.22 (0.04) | 5 | - | - |
| Cognitive test | 0.6 (0.04) | 85 | 0.19 (0.02) | 62 | 0.24 (0.02) | 63 | 0.32 (0.09) | 8 |
| **Motor disorders** | | | | | | | | |
| Diagnosis | 0.73 (0.15) | 3 | - | - | 0.32 (0.16) | 3 | - | - |
| Parent | 0.71 (0.11) | 4 | 0.12 (0.12) | 2 | 0.39 (0.12) | 4 | - | - |
| *Note.* H^2^= heritability; c^2^= shared environmental influences; e^2^= nonshared environmental influences; N= number of studies identified;  SE= standard error. | | | | | | | | |

| **Supplementary Table 41.** Genetic, shared and nonshared environmental correlations between NDDs, stratified by raters. | | | | | | | | |
| --- | --- | --- | --- | --- | --- | --- | --- | --- |
| **NDDs** | **Family rA (SE)** | **N** | **Family rC (SE)** | **N** | **Family rE (SE)** | **N** | **SNP rG (SE)** | **N** |
| **NDDs combined** | | | | | | | | |
| Parent | 0.34 (0.16) | 15 | 0.64 (0.45) | 5 | 0.17 (0.07) | 9 | - | - |
| Parent & Teacher | 0.41 (0.07) | 8 | - | - | 0.18 (0.1) | 3 | - | - |
| Teacher | 0.08 (0.52) | 3 | 0.88 (0.57) | 3 | 0.18 (0.1) | 3 | - | - |
| Cognitive test | 0.5 (0.09) | 11 | 0.69 (0.42) | 7 | 0.17 (0.07) | 7 | 0.25 (0.14) | 5 |
| **ASD & ADHD** | | | | | | | | |
| Parent | 0.67 (0.3) | 5 | - | - | 0.22 (0.12) | 4 | - | - |
| **ADHD & motor disorders** | | | | | | | | |
| Parent | 0.9 (0.82) | 2 | - | - | - | - | - | - |
| **ADHD & specific learning disorders** | | | | | | | | |
| Parent | -0.03 (0.13) | 8 | 0.25 (0.12) | 3 | 0.11 (0.06) | 4 | - | - |
| Parent & Teacher | 0.43 (0.08) | 7 | - | - | 0.26 (0.15) | 2 | - | - |
| Teacher | -0.4 (0.23) | 2 | 0.69 (0.2) | 2 | 0.1 (0.08) | 2 | - | - |
| **Communication disorders & specific learning disorders** | | | | | | | | |
| Cognitive test | 0.66 (0.15) | 2 | - | - | - | - | - | - |
| *Note.* rA= genetic correlation; rC= shared environmental correlation; rE= nonshared environmental correlation; N= number of studies identified; SE= standard error. | | | | | | | | |

| **Supplementary Table 42.** Genetic, shared and nonshared environmental correlations between NDDs and DICCs, stratified by raters. | | | | | | |
| --- | --- | --- | --- | --- | --- | --- |
| **NDDs and DICCs** | **Family rA (SE)** | **N** | **Family rC (SE)** | **N** | **Family rE (SE)** | **N** |
| **NDDs and DICCs combined** | | | | | | |
| Parent | 0.72 (0.34) | 6 | 0.93 (0.57) | 4 | 0.2 (0.09) | 5 |
| Parent & Self | 0.63 (0.5) | 2 | 0.97 (0.53) | 2 | 0.7 (0.61) | 2 |
| Parent & Teacher | 0.6 (0.28) | 3 | 0.82 (0.68) | 3 | 0.66 (0.6) | 2 |
| Self-report | 0.51 (0.25) | 2 | - | - | 0.11 (0.14) | 2 |
| **ADHD & conduct disorder** | | | | | | |
| Parent | 0.85 (0.61) | 3 | - | - | 0.22 (0.15) | 2 |
| **ADHD & oppositional defiant disorder** | | | | | | |
| Parent | 0.73 (0.32) | 2 | - | - | - | - |
| *Note.* rA= genetic correlation; rC= shared environmental correlation; rE= nonshared environmental correlation; N= number of studies identified; SE= standard error. | | | | | | |

| **Supplementary Table 43.** Heritability, shared and nonshared environmental influences on NDDs, stratified by number of covariates included in analyses. | | | | | | | | |
| --- | --- | --- | --- | --- | --- | --- | --- | --- |
| **NDDs** | **Family h2 (SE)** | **N** | **Family c2 (SE)** | **N** | **Family e2 (SE)** | **N** | **SNP h2 (SE)** | **N** |
| **NDDs combined** | | | | | | | | |
| 0 | 0.67 (0.04) | 56 | 0.22 (0.05) | 25 | 0.31 (0.03) | 39 | - | - |
| 1 | 0.68 (0.06) | 56 | 0.16 (0.04) | 25 | 0.27 (0.03) | 40 | 0.16 (0.07) | 2 |
| 2 | 0.64 (0.03) | 113 | 0.15 (0.02) | 69 | 0.3 (0.03) | 104 | 0.17 (0.16) | 3 |
| 3 | 0.61 (0.11) | 9 | 0.18 (0.07) | 5 | 0.31 (0.08) | 9 | 0.26 (0.06) | 14 |
| 4 | 0.73 (0.18) | 5 | 0.17 (0.08) | 3 | 0.23 (0.07) | 4 | - | - |
| **Intellectual disabilities** | | | | | | | | |
| 1 | 0.86 (0.44) | 2 | - | - | 0.1 (0.16) | 2 | - | - |
| **Communication disorders** | | | | | | | | |
| 0 | 0.47 (0.1) | 5 | 0.52 (0.11) | 3 | 0.15 (0.07) | 4 | - | - |
| 1 | 0.77 (0.24) | 7 | 0.29 (0.15) | 3 | 0.21 (0.1) | 5 | - | - |
| 2 | 0.5 (0.06) | 10 | 0.28 (0.07) | 8 | 0.26 (0.09) | 8 | - | - |
| **ASD** | | | | | | | | |
| 0 | 0.8 (0.19) | 11 | 0.03 (0.05) | 4 | 0.3 (0.1) | 6 | - | - |
| 1 | 0.76 (0.09) | 3 | - | - | 0.25 (0.09) | 3 | - | - |
| 2 | 0.68 (0.04) | 20 | 0.17 (0.08) | 8 | 0.26 (0.03) | 17 | - | - |
| **ADHD** | | | | | | | | |
| 0 | 0.68 (0.05) | 31 | 0.17 (0.07) | 12 | 0.36 (0.04) | 26 | - | - |
| 1 | 0.71 (0.09) | 25 | 0.08 (0.05) | 10 | 0.29 (0.05) | 21 | 0.17 (0.07) | 2 |
| 2 | 0.65 (0.04) | 58 | 0.09 (0.03) | 24 | 0.33 (0.04) | 54 | - | - |
| 3 | 0.66 (0.22) | 4 | - | - | 0.34 (0.17) | 4 | 0.15 (0.11) | 5 |
| 4 | 0.83 (0.16) | 3 | - | - | 0.11 (0.09) | 2 | - | - |
| **Specific learning disorders** | | | | | | | | |
| 0 | 0.58 (0.06) | 13 | 0.22 (0.07) | 8 | 0.21 (0.06) | 6 | - | - |
| 1 | 0.66 (0.07) | 26 | 0.21 (0.06) | 14 | 0.18 (0.03) | 15 | - | - |
| 2 | 0.59 (0.03) | 46 | 0.18 (0.03) | 39 | 0.26 (0.02) | 41 | - | - |
| 3 | 0.56 (0.06) | 6 | 0.17 (0.08) | 4 | 0.32 (0.06) | 6 | 0.31 (0.09) | 7 |
| **Motor disorders** | | | | | | | | |
| 1 | 0.7 (0.09) | 4 | 0.21 (0.15) | 2 | 0.43 (0.17) | 4 | - | - |
| 2 | 0.8 (0.05) | 2 | - | - | - | - | - | - |
| *Note.* H^2^= heritability; c^2^= shared environmental influences; e^2^= nonshared environmental influences; N= number of studies identified;  SE= standard error. | | | | | | | | |

| **Supplementary Table 44.** Genetic, shared and nonshared environmental correlations between NDDs, stratified by number of covariates included in analyses. | | | | | | |
| --- | --- | --- | --- | --- | --- | --- |
| **NDDs** | **Family rA (SE)** | **N** | **Family rC (SE)** | **N** | **Family rE (SE)** | **N** |
| **NDDs combined** | | | | | | |
| 0 | 0.35 (0.08) | 8 | - | - | - | - |
| 1 | 0.51 (0.22) | 7 | 0.1 (0.09) | 2 | 0.02 (0.08) | 2 |
| 2 | 0.3 (0.22) | 20 | 0.8 (0.35) | 13 | 0.17 (0.03) | 17 |
| 3 | 0.53 (0.11) | 2 | - | - | 0.44 (0.14) | 2 |
| **ASD & ADHD** | | | | | | |
| 2 | 0.68 (0.49) | 4 | - | - | 0.18 (0.09) | 4 |
| **ADHD & motor disorders** | | | | | | |
| 1 | 0.9 (0.82) | 2 | - | - | - | - |
| **ADHD & specific learning disorders** | | | | | | |
| 0 | 0.36 (0.13) | 4 | - | - | - | - |
| 1 | 0.28 (0.1) | 4 | - | - | - | - |
| 2 | -0.13 (0.13) | 9 | 0.4 (0.14) | 6 | 0.12 (0.05) | 7 |
| **Communication disorders & specific learning disorders** | | | | | | |
| 2 | 0.66 (0.15) | 2 | - | - | - | - |
| *Note.* rA= genetic correlation; rC= shared environmental correlation; rE= nonshared environmental correlation; N= number of studies identified; SE= standard error. | | | | | | |

| **Supplementary Table 45.** Genetic, shared and nonshared environmental correlations between NDDs and DICCs, stratified by number of covariates included in analyses. | | | | | | |
| --- | --- | --- | --- | --- | --- | --- |
| **NDDs and DICCs** | **Family rA (SE)** | **N** | **Family rC (SE)** | **N** | **Family rE (SE)** | **N** |
| **NDDs and DICCs combined** | | | | | | |
| 0 | 0.39 (0.13) | 6 | 0.71 (0.6) | 4 | 0.2 (0.09) | 5 |
| 1 | 0.58 (0.19) | 5 | 0.94 (0.55) | 4 | 0.44 (0.34) | 4 |
| 2 | 0.93 (0.74) | 3 | 0.93 (0.77) | 2 | 0.58 (0.41) | 3 |
| **ADHD & conduct disorder** | | | | | | |
| 0 | 0.43 (0.24) | 2 | - | - | 0.12 (0.16) | 2 |
| 1 | 0.37 (0.1) | 3 | 0.87 (0.86) | 2 | 0.05 (0.1) | 2 |
| **ADHD & oppositional defiant disorder** | | | | | | |
| 0 | 0.62 (0.25) | 2 | - | - | 0.35 (0.17) | 2 |
| 1 | 0.56 (0.29) | 3 | 0.87 (0.86) | 2 | 0.32 (0.1) | 2 |
| *Note.* rA= genetic correlation; rC= shared environmental correlation; rE= nonshared environmental correlation; N= number of studies identified; SE= standard error. | | | | | | |

| **Supplementary Table 46.** Heritability, shared and nonshared environmental influences on NDDs, stratified by measurement instruments. | | | | | | | | | |
| --- | --- | --- | --- | --- | --- | --- | --- | --- | --- |
| **Measures from family-based studies** | | | | | | | **Measures from SNP-based studies** | | |
| **NDDs** | **Family h2 (SE)** | **N** | **Family c2 (SE)** | **N** | **Family e2 (SE)** | **N** |  | **SNP h2 (SE)** | **N** |
| **Intellectual disabilities** | | | | | | | | | |
| ICD-9/ICD-10 | 0.86 (0.44) | 2 | - | - | 0.1 (0.16) | 2 |  |  |  |
| **Communication disorders** | | | | | | | | | |
| Clinical evaluation | 0.75 (0.13) | 3 | - | - | 0.27 (0.15) | 2 | TOAL | 0.32 (0.16) | 3 |
| Goldman-Fristoe Test of Articulation | 0.58 (0.2) | 2 | 0.27 (0.2) | 2 | 0.16 (0.12) | 2 |  | - | - |
| MCDI | 0.46 (0.13) | 3 | 0.53 (0.11) | 3 | 0.05 (0.05) | 3 |  | - | - |
| TEGI | 0.74 (0.32) | 2 | 0.12 (0.2) | 2 | 0.19 (0.2) | 2 |  | - | - |
| **ASD** | | | | | | | | | |
| A-TAC | 0.73 (0.06) | 8 | 0.14 (0.08) | 3 | 0.29 (0.04) | 7 | AQ | - | - |
| ADI-R & ADOS | 0.81 (0.62) | 2 | 0.28 (0.3) | 2 | - | - | CAST | 0.03 (0.18) | 2 |
| AQ | 0.51 (0.1) | 3 | - | - | 0.2 (0.17) | 2 | ICD-9/ICD-10 | 0.12 (0.05) | 7 |
| ADI-R | 0.81 (0.45) | 3 | 0.3 (0.22) | 2 | 0.14 (0.22) | 2 | SCDC | 0.24 (0.1) | 4 |
| CAST | 0.7 (0.04) | 14 | 0.09 (0.06) | 4 | 0.27 (0.03) | 11 |  | - | - |
| DAWBA | 0.75 (0.15) | 3 | - | - | 0.22 (0.17) | 2 |  | - | - |
| DSM-4/DSM-5 | 0.69 (0.08) | 2 | - | - | 0.31 (0.09) | 2 |  | - | - |
| ICD-9/ICD-10 | 0.8 (0.12) | 3 | 0.01 (0.1) | 2 | 0.19 (0.11) | 3 |  | - | - |
| **ADHD** | | | | | | | | | |
| A-TAC | 0.78 (0.1) | 5 | 0.03 (0.07) | 2 | 0.25 (0.05) | 5 | CBRS | 0.13 (0.13) | 3 |
| ATBRS | 0.82 (0.07) | 3 | 0.23 (0.14) | 3 | 0.12 (0.08) | 3 | ICD-9/ICD-10 | 0.21 (0.21) | 7 |
| CBCL | 0.61 (0.09) | 14 | 0.05 (0.06) | 5 | 0.25 (0.04) | 11 | SDQ | 0.09 (0.09) | 4 |
| CBCL & YSR | 0.78 (0.06) | 3 | - | - | 0.25 (0.09) | 3 | TRF | 0.53 (0.53) | 2 |
| CBRS | 0.72 (0.03) | 29 | 0.18 (0.16) | 11 | 0.24 (0.03) | 28 |  | - | - |
| DBD | 0.69 (0.25) | 3 | 0.16 (0.13) | 3 | 0.19 (0.07) | 3 |  | - | - |
| DBRS | 0.76 (0.11) | 4 | 0.03 (0.1) | 3 | 0.25 (0.08) | 3 |  | - | - |
| DCB | 0.67 (0.07) | 2 | - | - | - | - |  | - | - |
| DICA | 0.69 (0.21) | 3 | - | - | - | - |  | - | - |
| DISC | 0.51 (0.1) | 5 | 0.03 (0.16) | 2 | 0.54 (0.11) | 4 |  | - | - |
| DSM-4/DSM-5 | 0.77 (0.29) | 9 | 0.11 (0.11) | 4 | 0.35 (0.08) | 7 |  | - | - |
| DuPaul ADHD Rating Scale | 0.75 (0.05) | 4 | 0.29 (0.12) | 2 | 0.25 (0.07) | 4 |  | - | - |
| ECRS | 0.77 (0.23) | 2 | - | - | 0.28 (0.1) | 2 |  | - | - |
| ICD-9/ICD-10 | 0.87 (0.11) | 3 | - | - | 0.12 (0.05) | 3 |  | - | - |
| Rutter Scales | 0.75 (0.15) | 4 | - | - | 0.26 (0.13) | 2 |  | - | - |
| SBQ | 0.61 (0.26) | 2 | - | - | 0.38 (0.19) | 2 |  | - | - |
| SDQ | 0.65 (0.1) | 15 | 0.07 (0.12) | 4 | 0.43 (0.07) | 14 |  | - | - |
| SWAN | 0.73 (0.16) | 8 | 0.35 (0.09) | 5 | 0.14 (0.05) | 7 |  | - | - |
| TRF | 0.6 (0.12) | 4 | - | - | 0.46 (0.08) | 3 |  | - | - |
| **Specific learning disorders** | | | | | | | | | |
| Comprehensive Test  of Phonological Processing | 0.55 (0.17) | 3 | 0.22 (0.16) | 3 | 0.27 (0.1) | 3 | GCSE | 0.34 (0.2) | 2 |
| FCAT | 0.46 (0.13) | 4 | 0.31 (0.14) | 4 | 0.23 (0.07) | 4 | NFER | 0.31 (0.16) | 3 |
| GCSE | 0.61 (0.07) | 5 | 0.22 (0.07) | 5 | 0.18 (0.04) | 5 | PIAT | 0.24 (0.22) | 2 |
| National Curriculum | 0.64 (0.08) | 7 | 0.15 (0.05) | 7 | 0.23 (0.03) | 7 | TOWRE | 0.36 (0.2) | 2 |
| NFER | 0.49 (0.06) | 9 | 0.17 (0.07) | 7 | 0.33 (0.05) | 7 | National Curriculum | 0.33 (0.18) | 2 |
| PIAT | 0.56 (0.07) | 21 | 0.22 (0.06) | 14 | 0.25 (0.07) | 13 |  | - | - |
| PIAT & GOAL | 0.59 (0.09) | 5 | 0.21 (0.07) | 4 | 0.35 (0.1) | 4 |  | - | - |
| PIAT & TOWRE | 0.66 (0.15) | 2 | - | - | - | - |  | - | - |
| PIAT & WISC | 0.59 (0.15) | 5 | 0.23 (0.19) | 3 | 0.11 (0.18) | 2 |  | - | - |
| PIAT & WRAT | 0.51 (0.2) | 3 | - | - | - | - |  | - | - |
| TOWRE | 0.7 (0.07) | 8 | 0.13 (0.06) | 8 | 0.17 (0.04) | 8 |  | - | - |
| WISC | 0.41 (0.27) | 2 | - | - | - | - |  | - | - |
| The Woodcock–Johnson Tests  of Cognitive Abilities | 0.57 (0.11) | 8 | 0.19 (0.1) | 7 | 0.24 (0.06) | 7 |  | - | - |
| TOWRE & The Woodcock–Johnson  Tests of Cognitive Abilities | 0.77 (0.16) | 2 | - | - | - | - |  | - | - |
| WRAT | 0.48 (0.19) | 2 | 0.33 (0.18) | 2 | 0.2 (0.12) | 2 |  | - | - |
| **Motor disorders** | | | | | | | | | |
| A-TAC | 0.58 (0.12) | 2 | - | - | 0.42 (0.12) | 2 | - | - | - |
| *Note.* H^2^= heritability; c^2^= shared environmental influences; e^2^= nonshared environmental influences; N= number of studies identified;  SE= standard error; TOAL= Test of Adolescent and Adult Language; MCDI= MacArthur-Bates Communicative Development Inventories; TEGI= Test of Early Grammatical Impairment; A-TAC= Autism-Tics, AD/HD, and other Comorbidities Inventory; ADI-R= The Autism Diagnostic Interview-Revised; ADOS= Autism Diagnostic Observation Schedule; AQ= Autism Spectrum Quotient; CAST= Childhood Autism Spectrum Test; SCDC= Social and Communication Disorders Checklist; DAWBA= Developmental and Well-Being Assessment; DSM= Diagnostic Statistical Manual; ICD= International Classification of Diseases; ATBRS= Australian Twin Behaviour Rating Scale; CBRS= Conners Comprehensive Behaviour Rating Scale; CBCL= Child Behavior Checklist; YSR= Youth Self-Report; DBD= Disruptive Behavior Disorder Rating Scale; DBRS= The Disruptive Behavior Rating Scale; DCB= Devereux Child Behavior Rating Scale; DICA= Diagnostic Interview for Children and Adolescents; DISC= Diagnostic Interview Schedule for Children; ECRS= Emory Combined Rating Scale; SBQ= Social  Behavior Questionnaire; SDQ= Strengths and Difficulties Questionnaire; SWAN= Strengths and Weaknesses of Attention-Deficit/Hyperactivity-symptoms and Normal-behaviors; TRF= Teacher Report Form; FCAT= The Florida Comprehensive Assessment Test; GCSE= General Certificate of Secondary Education; NFER= National Foundation for Educational Research; PIAT= The Peabody Individual Achievement Test; GOAL= Greater Opportunities for Adult Learning Success; TOWRE= Test of Word Reading Efficiency; WISC= Wechsler Intelligence Scale for Children; WRAT= Wide Range Achievement Test. | | | | | | | | | |

| **Supplementary Table 47.** Genetic, shared and nonshared environmental correlations between NDDs, stratified by measurement instruments. | | | | | | |
| --- | --- | --- | --- | --- | --- | --- |
| **NDDs** | **Family rA (SE)** | **N** | **Family rC (SE)** | **N** | **Family rE (SE)** | **N** |
| **ASD & ADHD** | | | | | | |
| A-TAC | 0.8 (0.25) | 3 | - | - | 0.36 (0.12) | 2 |
| CAST & CBRS | 0.26 (0.1) | 2 | - | - | 0.1 (0.08) | 2 |
| **ADHD & specific learning disorders** | | | | | | |
| CBRS & PIAT | -0.29 (0.1) | 2 | 0.23 (0.13) | 2 | 0.1 (0.08) | 2 |
| CBRS & RDQ | 0.48 (0.13) | 2 | - | - | 0.26 (0.15) | 2 |
| DBRS & PIAT | 0.33 (0.25) | 3 | - | - | - | - |
| DICA & PIAT | 0.35 (0.18) | 2 | - | - | - | - |
| *Note.* rA= genetic correlation; rC= shared environmental correlation; rE= nonshared environmental correlation; N= number of studies identified; SE= standard error; A-TAC= Autism-Tics, AD/HD, and other Comorbidities Inventory; CAST= Childhood Autism Spectrum Test; CBRS= Conners Comprehensive Behaviour Rating Scale; PIAT= The Peabody Individual Achievement Test; DBRS= The Disruptive Behavior Rating Scale; DICA= Diagnostic Interview for Children and Adolescents; RDQ= Reading Difficulties Questionnaire. | | | | | | |

### Supplementary Figures

| Supplementary Figures: table of contents | |
| --- | --- |
| Supplementary Figure 1 | Page 113 |
| Supplementary Figure 2 | Page 114 |
| Supplementary Figure 3 | Page 115 |
| Supplementary Figure 4 | Page 116 |
| Supplementary Figure 5 | Page 117 |
| Supplementary Figure 6 | Page 118 |
| Supplementary Figure 7 | Page 119 |
| Supplementary Figure 8 | Page 120 |
| Supplementary Figure 9 | Page 121 |
| Supplementary Figure 10 | Page 122 |
| Supplementary Figure 11 | Page 123 |
| Supplementary Figure 12 | Page 124 |
| Supplementary Figure 13 | Page 125 |
| Supplementary Figure 14 | Page 126 |
| Supplementary Figure 15 | Page 127 |
| Supplementary Figure 16 | Page 128 |
| Supplementary Figure 17 | Page 129 |
| Supplementary Figure 18 | Page 130 |
| Supplementary Figure 19 | Page 131 |
| Supplementary Figure 20 | Page 132 |
| Supplementary Figure 21 | Page 133 |
| Supplementary Figure 22 | Page 134 |
| Supplementary Figure 23 | Page 135 |
| Supplementary Figure 24 | Page 136 |

**Supplementary Figure 1**: Distribution of sources of variation in NDDs, their overlap and overlap with DICCs.

**Supplementary Figure 2**: Heritability and environmental influences on specific phenotypes within NDDs categories.

**Supplementary Figure 3**: Genetic and environmental overlap between specific phenotypes within the NDDs category and between specific phenotypes within the NDDs and DICCs category.

**Supplementary Figure 4**: Variance in heritability and environmental influences on NDDs, variance in genetic and environmental correlations between NDDs and variance in genetic and environmental correlations between NDDs and DICCs that can be attributed to heterogeneity (the I^2^ statistic).

**Supplementary Figure 5**: Funnel plots involving all studies addressing heritability and environmental influences on NDDs.

**Supplementary Figure 6**: Funnel plots involving all studies addressing heritability and environmental influences on intellectual disabilities.

**Supplementary Figure 7**: Funnel plots involving all studies addressing heritability and environmental influences on communication disorders.

**Supplementary Figure 8**: Funnel plots involving all studies addressing heritability and environmental influences on ASD.

**Supplementary Figure 9**: Funnel plots involving all studies addressing heritability and environmental influences on ADHD.

**Supplementary Figure 10**: Funnel plots involving all studies addressing heritability and environmental influences on specific learning disorders.

**Supplementary Figure 11**: Funnel plots involving all studies addressing heritability and environmental influences on motor disorders.

**Supplementary Figure 12**: Funnel plots involving all studies addressing genetic and environmental overlap between NDDs.

**Supplementary Figure 13**: Funnel plots involving all studies addressing genetic and environmental overlap between ASD & ADHD.

**Supplementary Figure 14**: Funnel plots involving all studies addressing genetic and environmental overlap between ADHD & motor disorders.

**Supplementary Figure 15**: Funnel plots involving all studies addressing genetic and environmental overlap between ADHD & specific learning disorders.

**Supplementary Figure 16**: Funnel plots involving all studies addressing genetic and environmental overlap between communication disorders & motor disorders.

**Supplementary Figure 17**: Funnel plots involving all studies addressing genetic and environmental overlap between communication disorders & specific learning disorders.

**Supplementary Figure 18**: Funnel plots involving all studies addressing genetic and environmental overlap between neurodevelopmental disorders (NDDs) and disruptive, impulse control and conduct disorders (DICCs).

**Supplementary Figure 19**: Funnel plots involving all studies addressing genetic and environmental overlap between ADHD & conduct disorder.

**Supplementary Figure 20**: Funnel plots involving all studies addressing genetic and environmental overlap between ADHD & oppositional defiant disorder.

**Supplementary Figure 21**: Funnel plots involving all studies addressing genetic and environmental overlap between ASD & conduct disorder.

**Supplementary Figure 22**: Sources of variation in NDDs, genetic and environmental overlap between NDDs and genetic and environmental overlap between NDDs and DICCs, stratified by measurement scales, i.e., categorical versus continuous measurement.

**Supplementary Figure 23**: Diagram of searches and screening.

**Supplementary Figure 24**: Grand heritability and environmental influences on NDDs, grand genetic and environmental correlations between NDDs and grand genetic and environmental correlations between NDDs and DICCs obtained using different aggregation techniques, i.e., aggregating by study, cohort, and country, using correlation thresholds of r= 0.3, r= 0.5 and r= 0.9.

**Supplementary Figure 1.** Panel **A** presents distribution of heritability and environmental influences on neurodevelopmental disorders (NDDs (top panel), as well as genetic and environmental correlations between NDDs (right bottom panel) and between NDDs and disruptive, impulse control and conduct disorders (DICCs) (left bottom panel). Panel **B** presents density plot of heritability and environmental influences on NDDs (top panel), as well as genetic and environmental correlations between NDDs (middle panel) and between NDDs and DICCs (bottom panel).

**A**

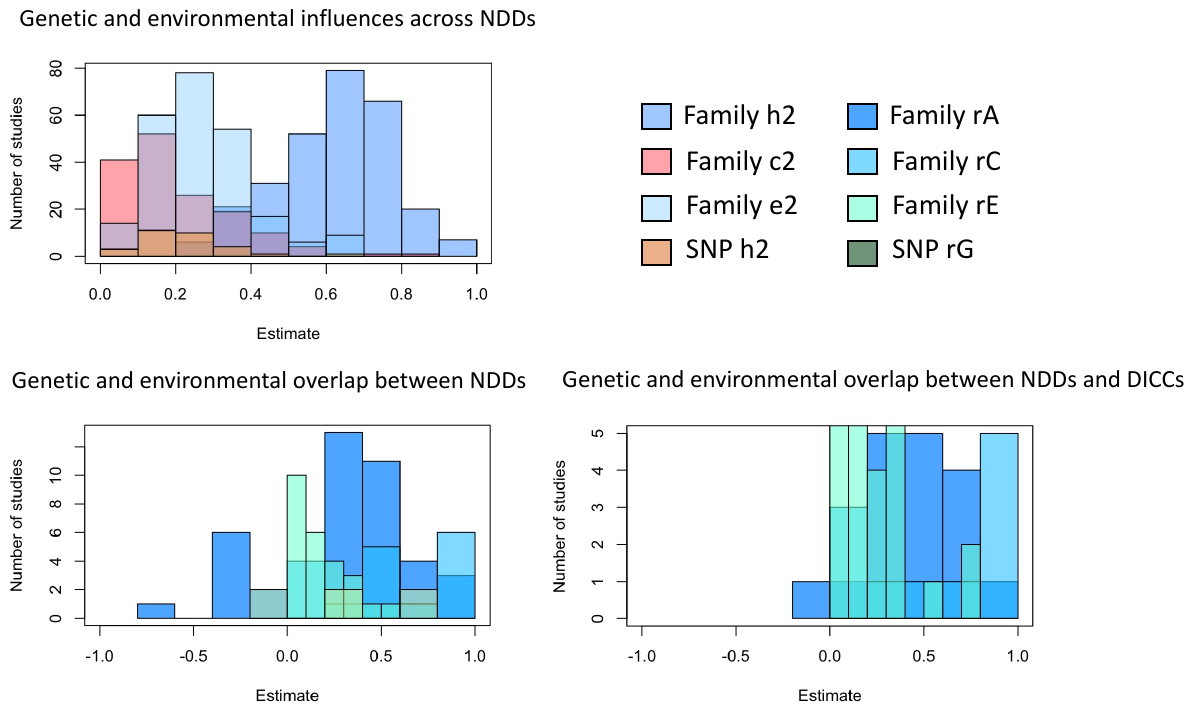

**B**

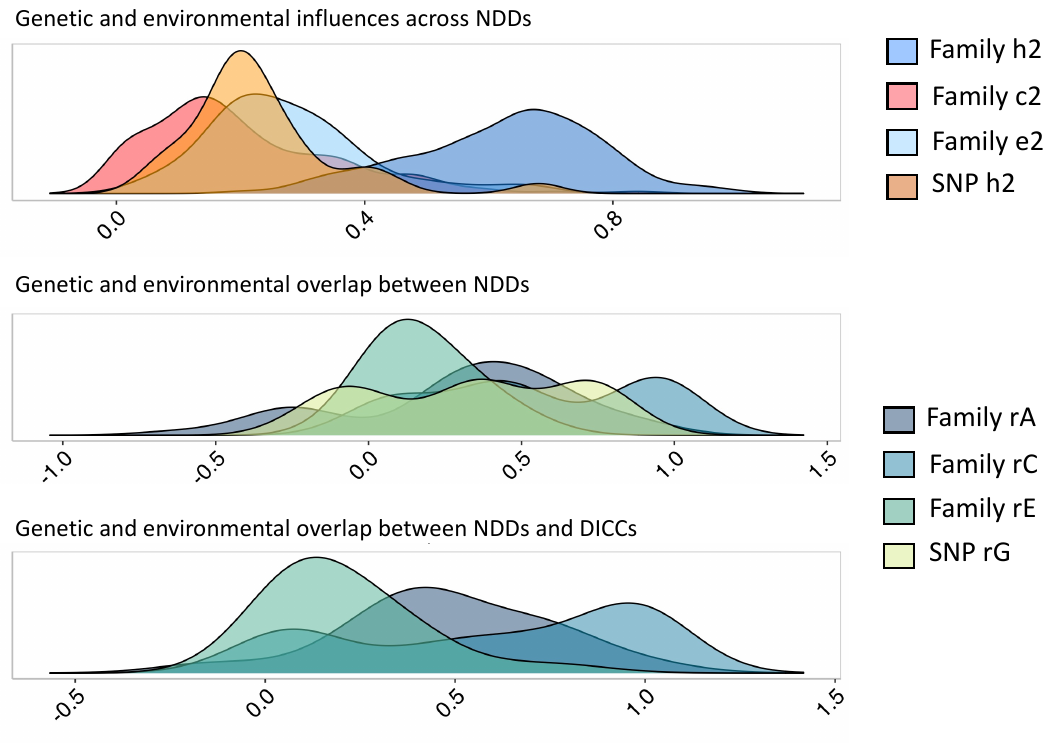

**Supplementary Figure 2**. Heritability and environmental influences on specific phenotypes within neurodevelopmental disorders (NDDs) categories.

**Supplementary Figure 3.** Genetic and environmental overlap between specific phenotypes within the neurodevelopmental disorders (NDDs) category and between specific phenotypes within the NDDs and disruptive, impulse control and conduct disorders (DICCs) category.

**Supplementary Figure 4**. Variance in heritability and environmental influences on neurodevelopmental disorders (NDDs) (top panel), variance in genetic and environmental correlations between NDDs (middle panel) and variance in genetic and environmental correlations between NDDs and disruptive, impulse control and conduct disorders (DICCs) that can be attributed to heterogeneity (the I^2^ statistic).

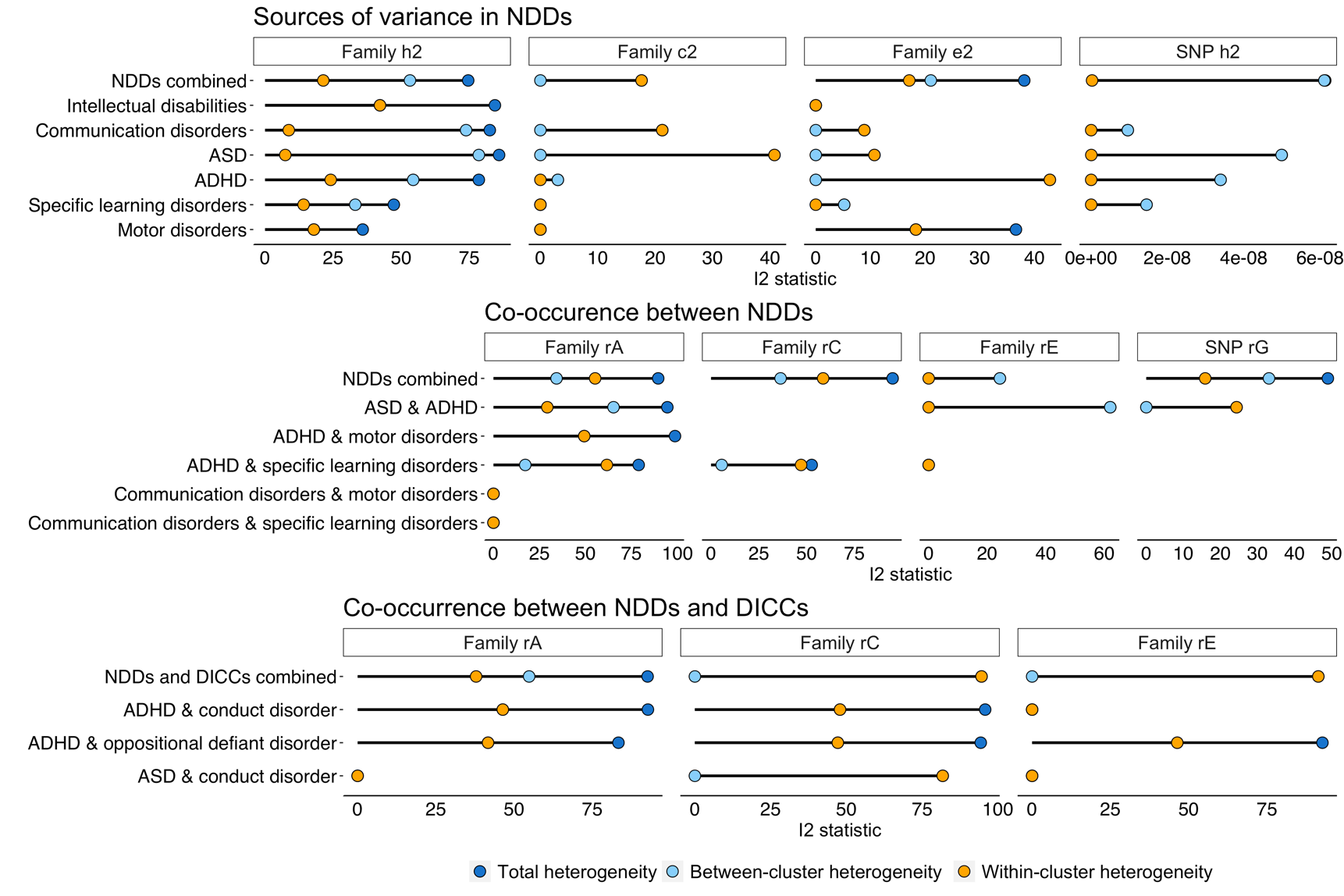

| **Supplementary Figure 5.** Funnel plots involving all studies addressing heritability and environmental influences on neurodevelopmental disorders (NDDs). | |
| --- | --- |
| 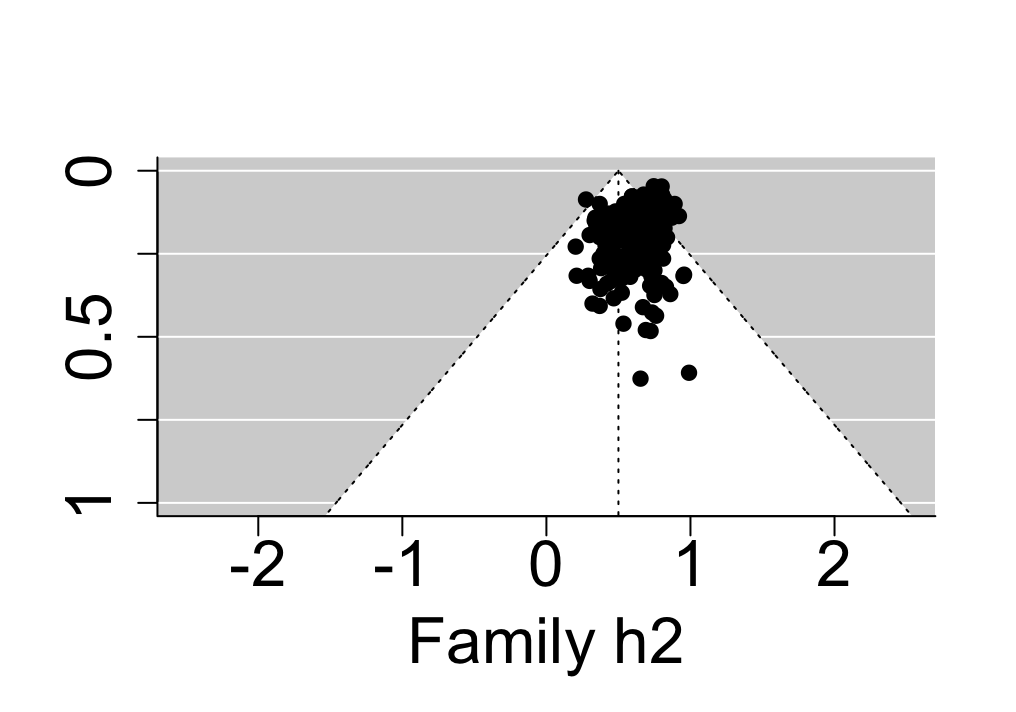 | 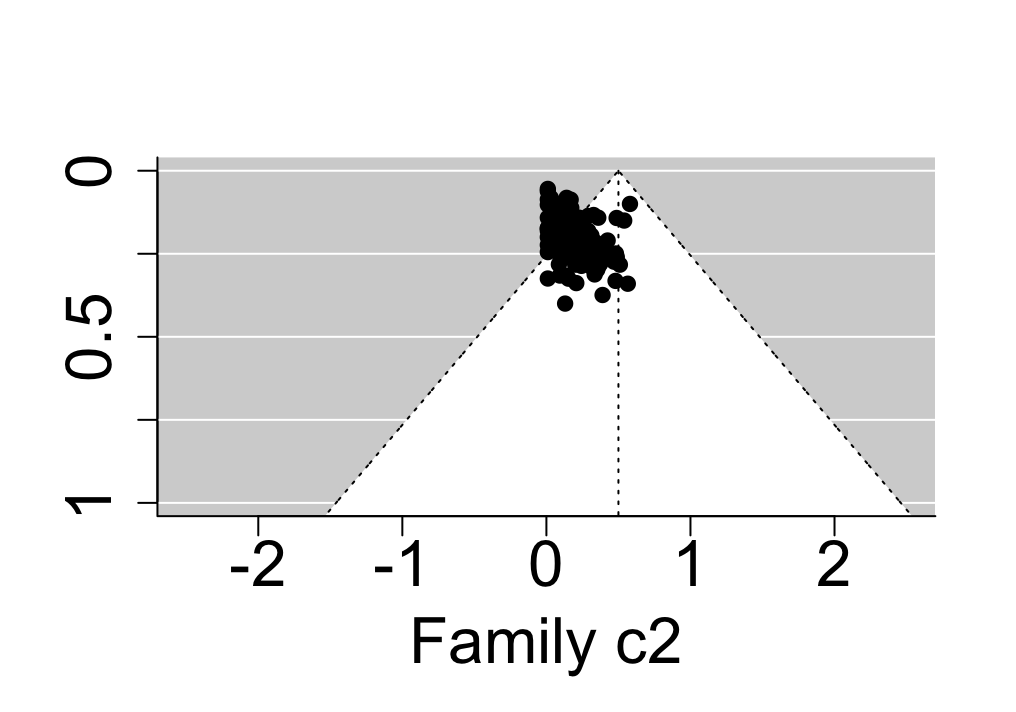 |
| 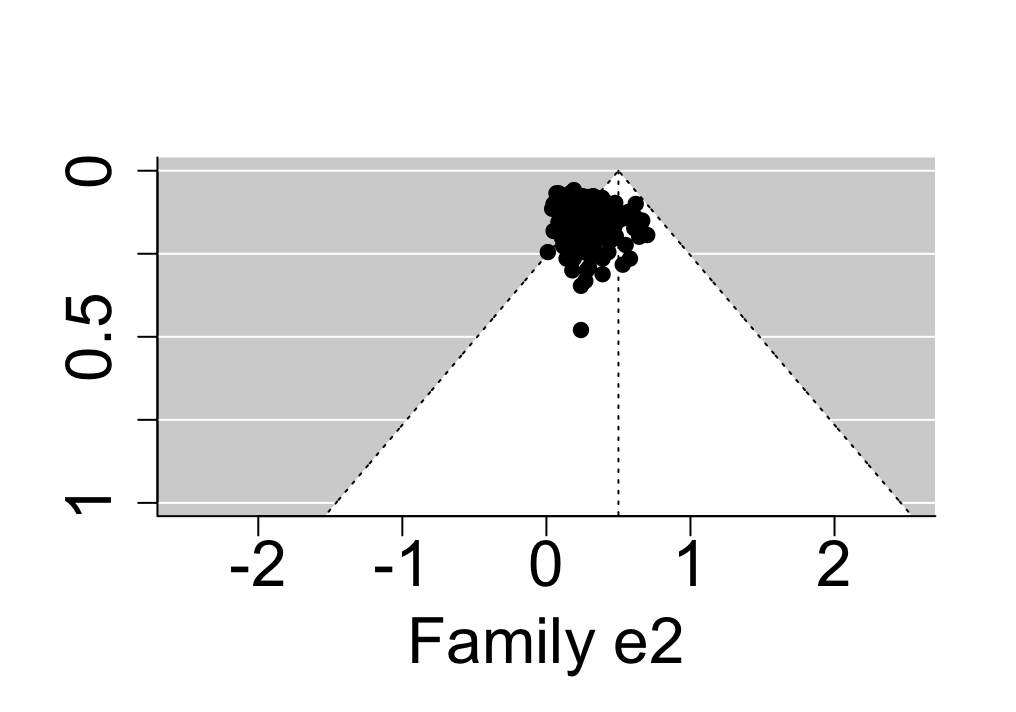 | 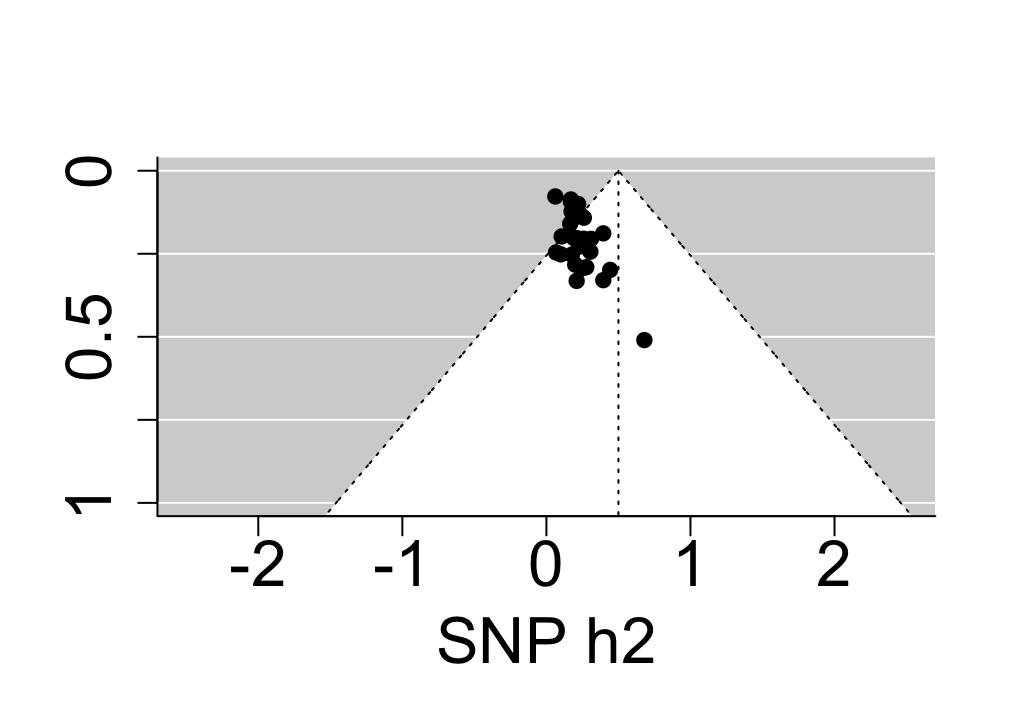 |

| **Supplementary Figure 6.** Funnel plots involving all studies addressing heritability and environmental influences on intellectual disabilities. | |
| --- | --- |
| 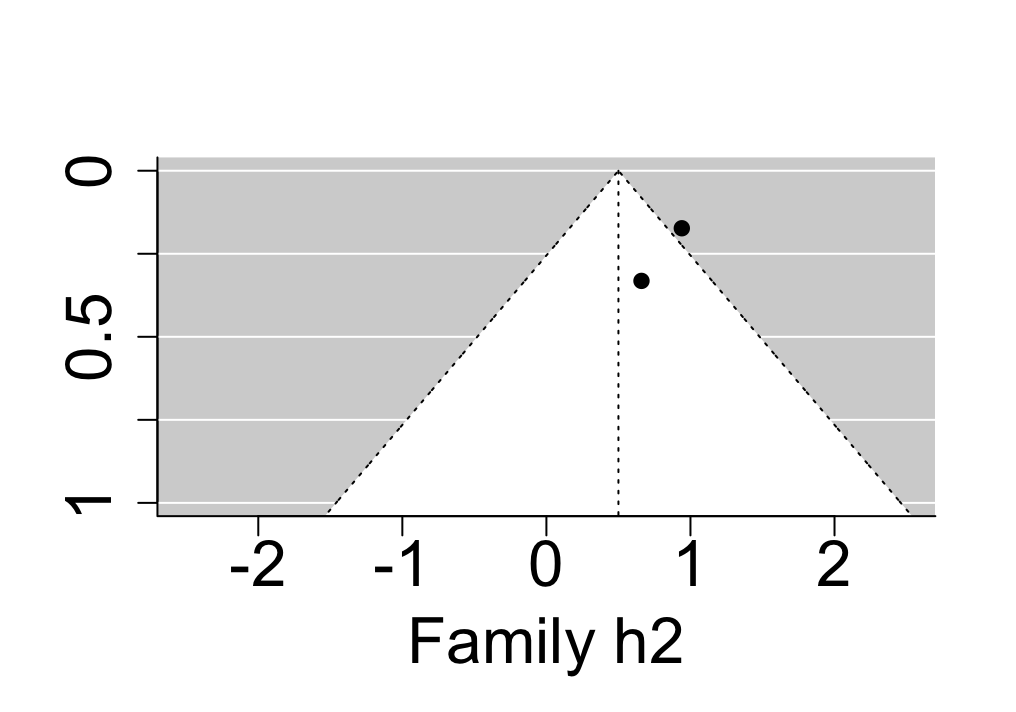 | 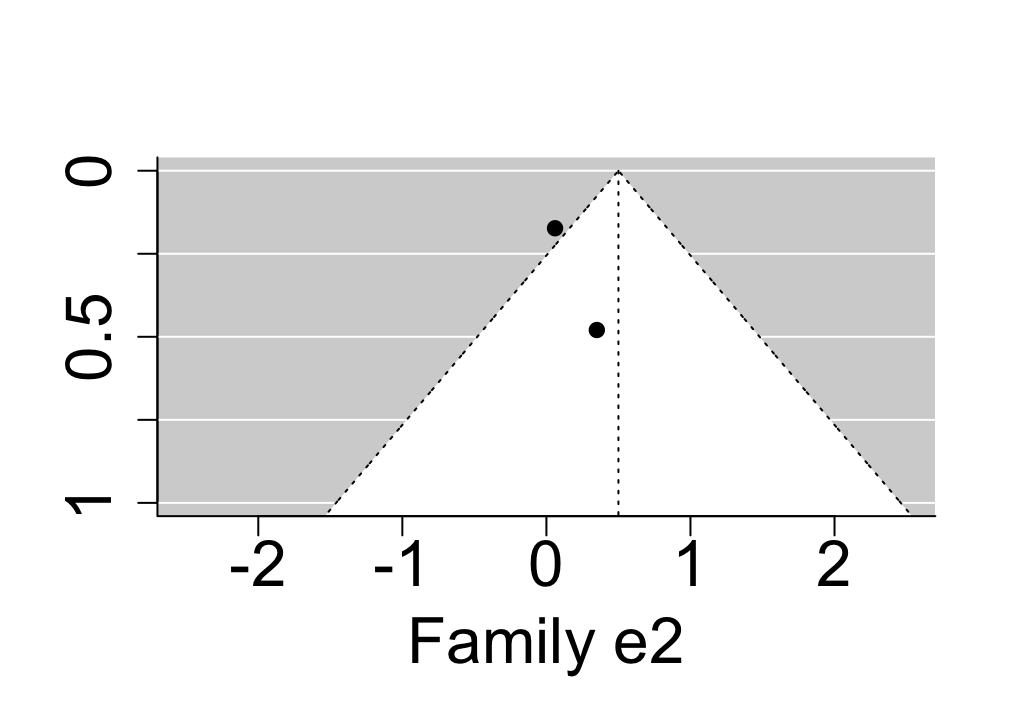 |

| **Supplementary Figure 7.** Funnel plots involving all studies addressing heritability and environmental influences on communication disorders. | |
| --- | --- |
| 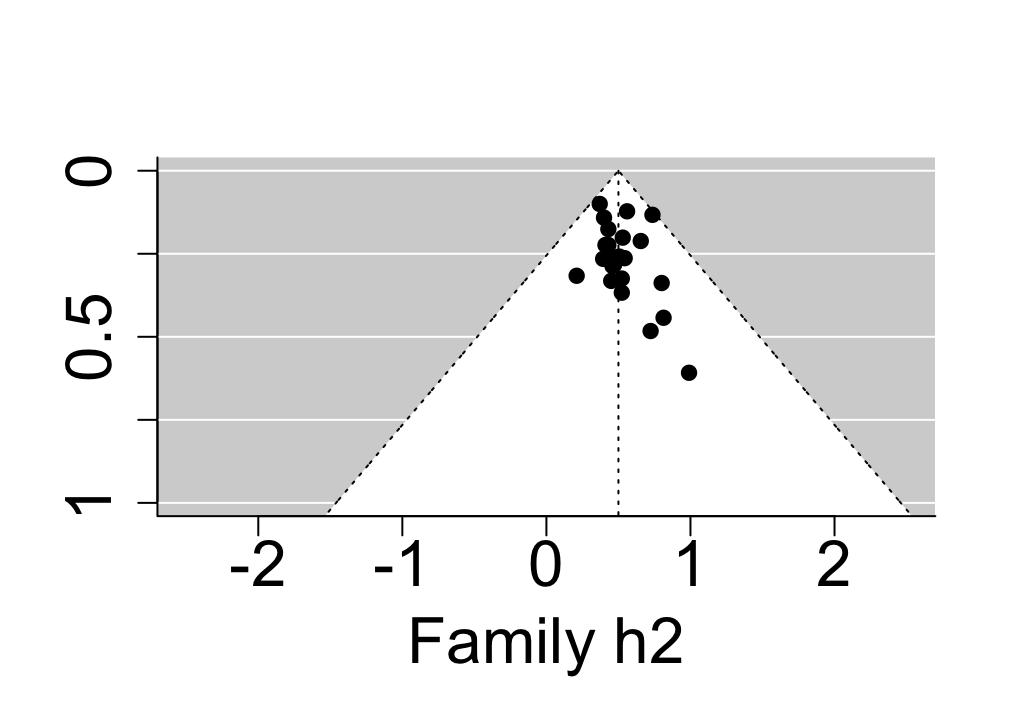 | 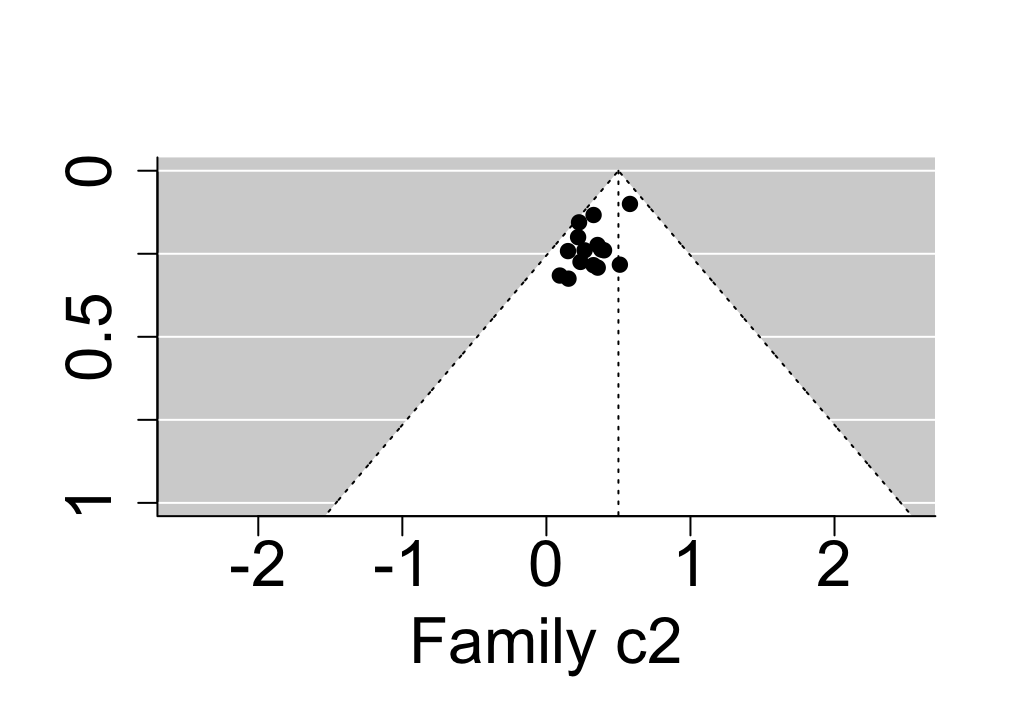 |
| 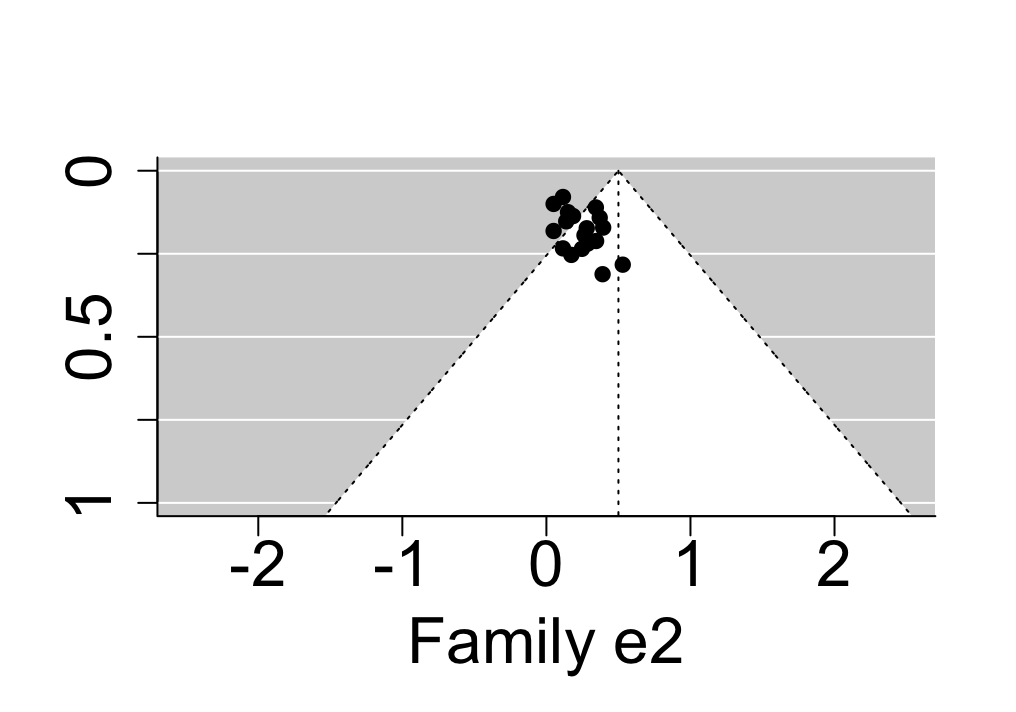 | 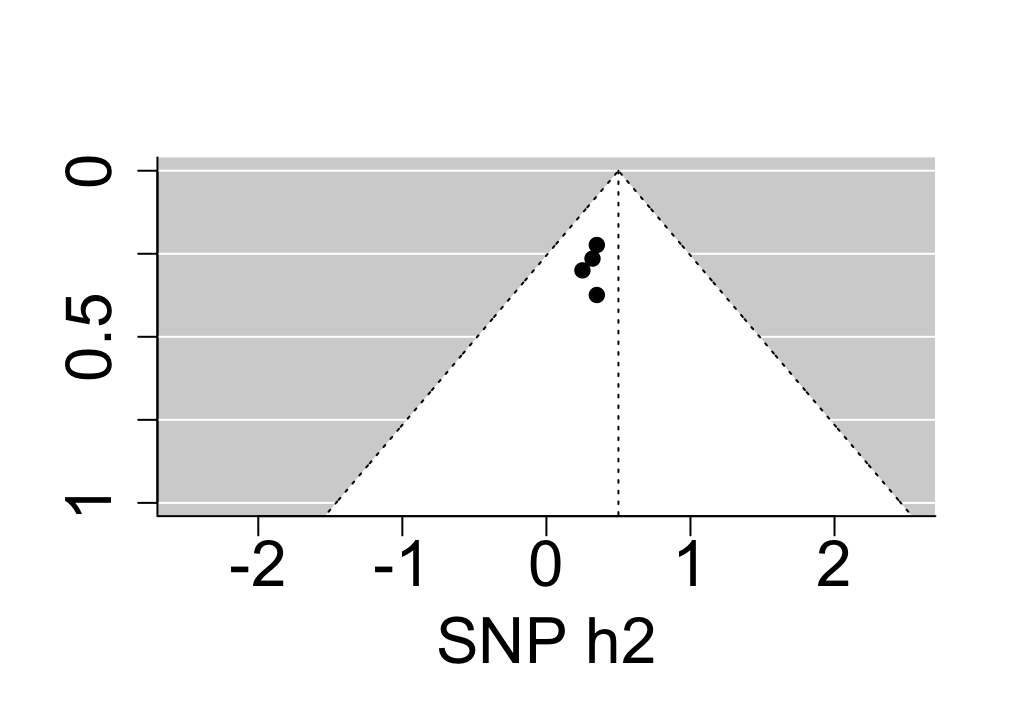 |

| **Supplementary Figure 8.** Funnel plots involving all studies addressing heritability and environmental influences on ASD. | |
| --- | --- |
| 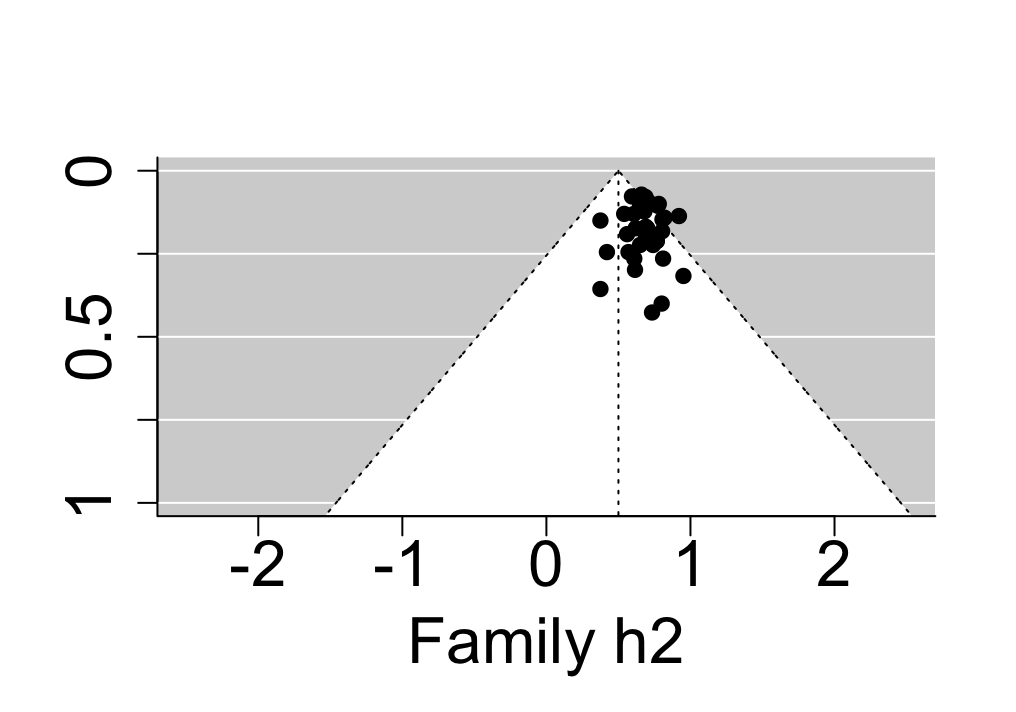 | 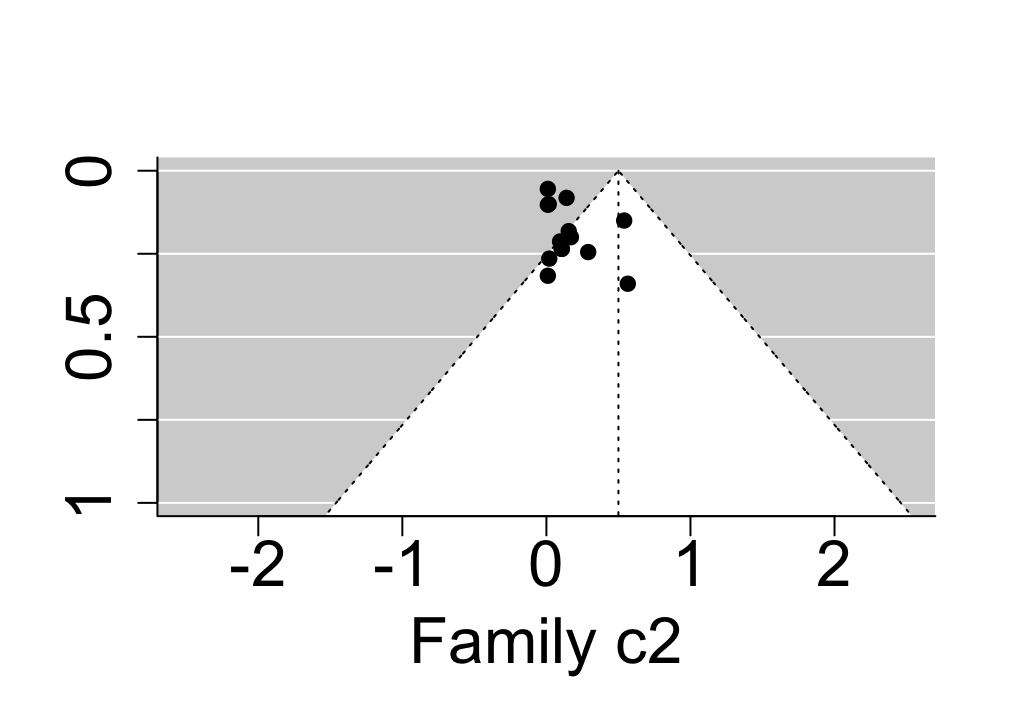 |
| 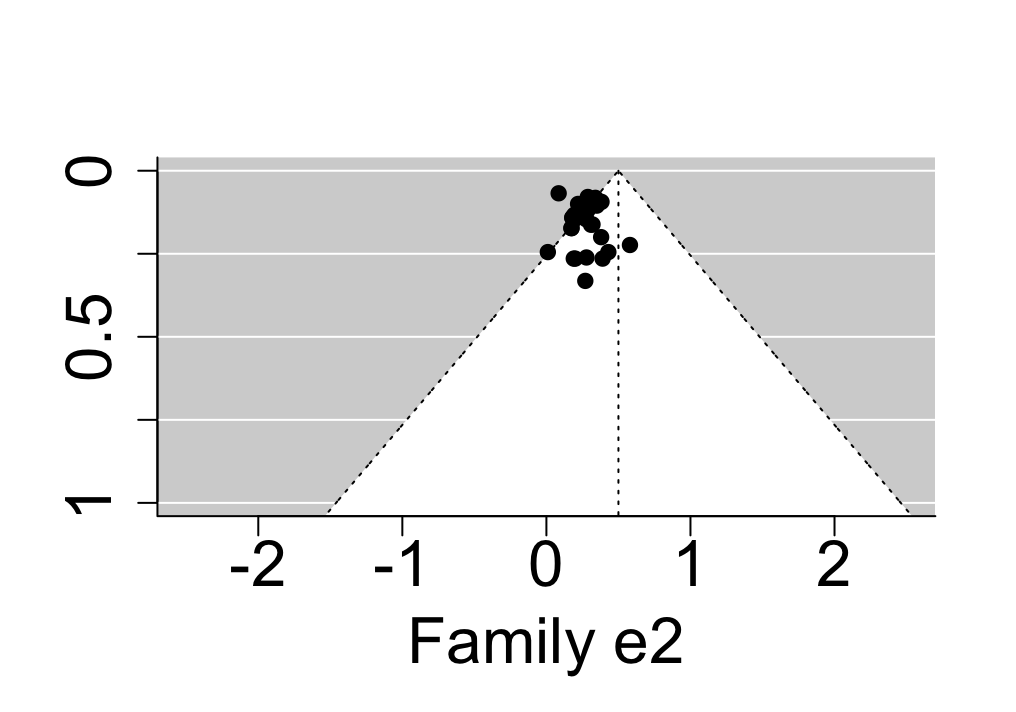 | 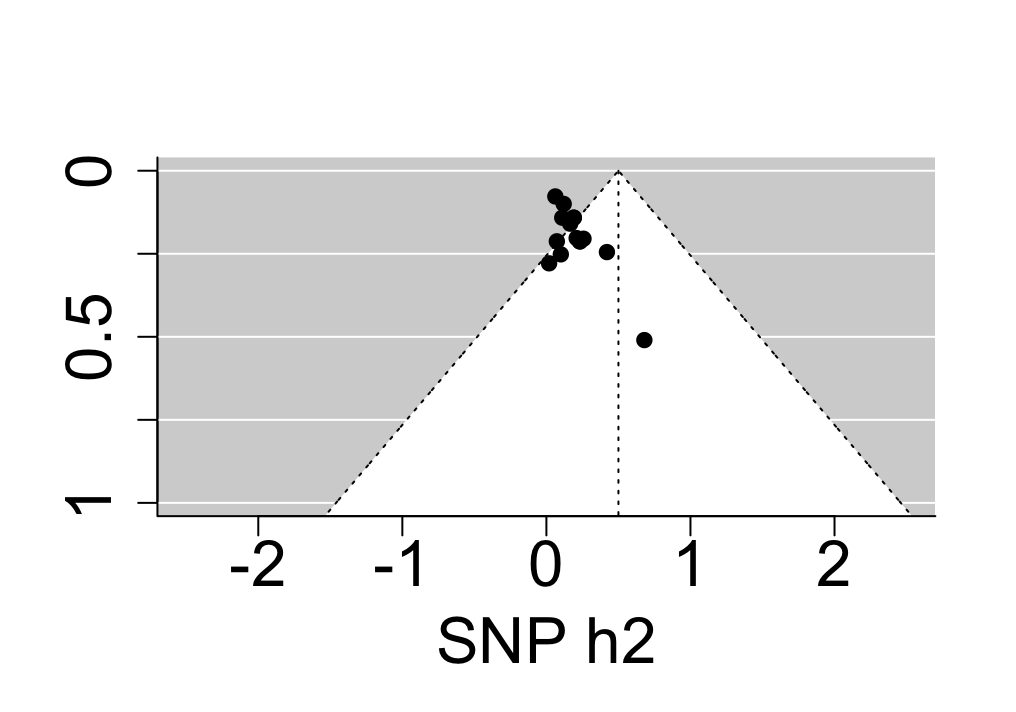 |

| **Supplementary Figure 9.** Funnel plots involving all studies addressing heritability and environmental influences on ADHD. | |
| --- | --- |
| 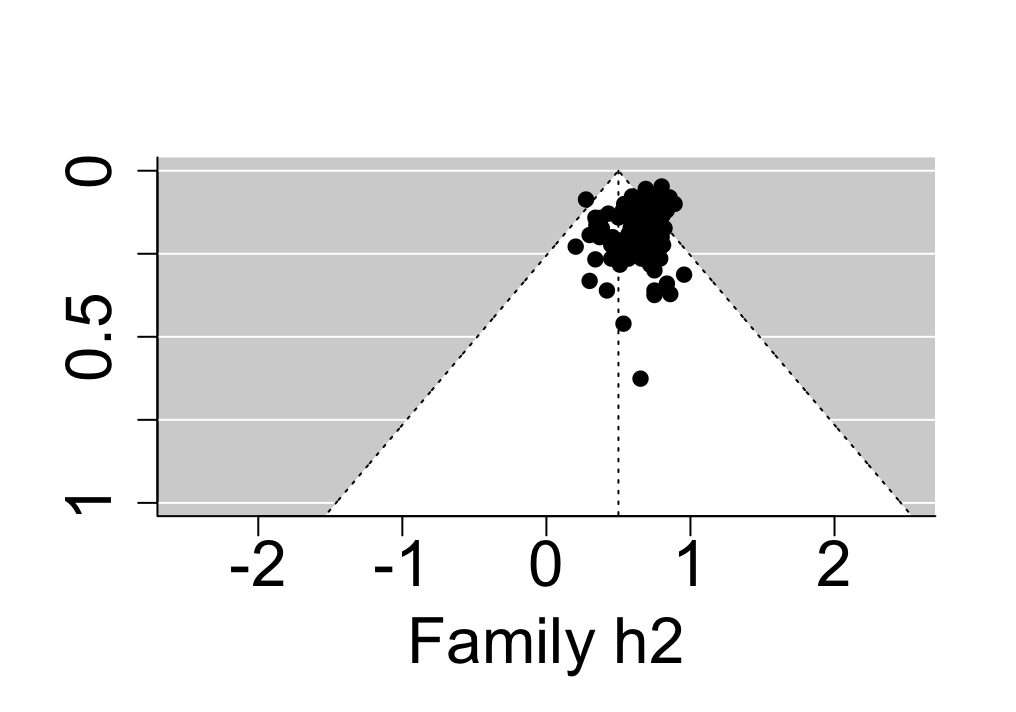 | 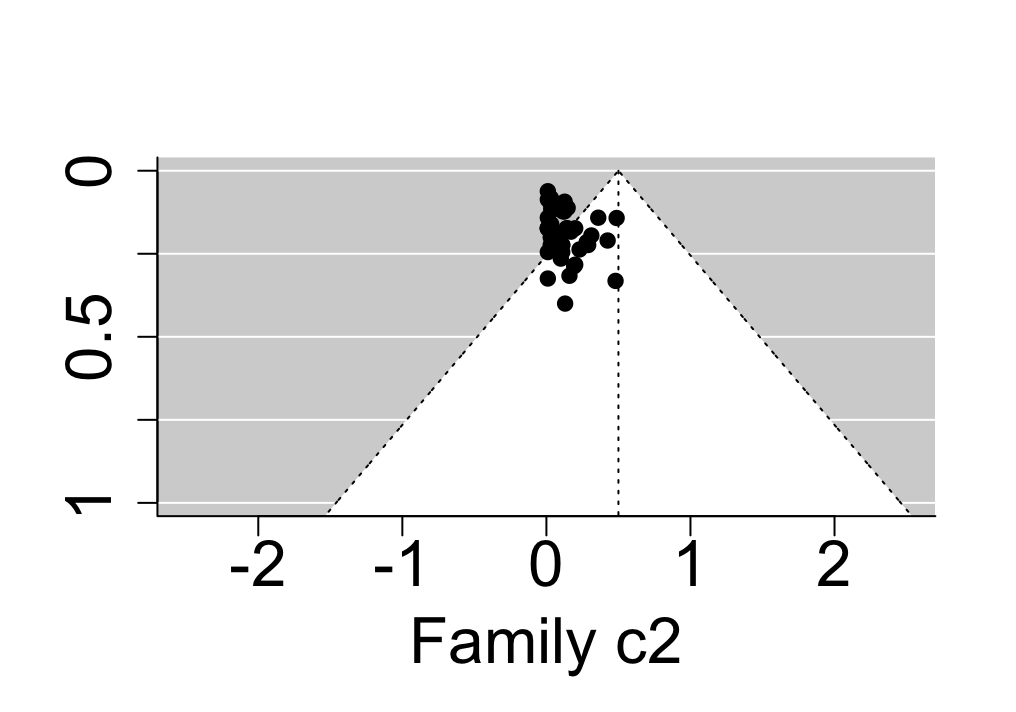 |
| 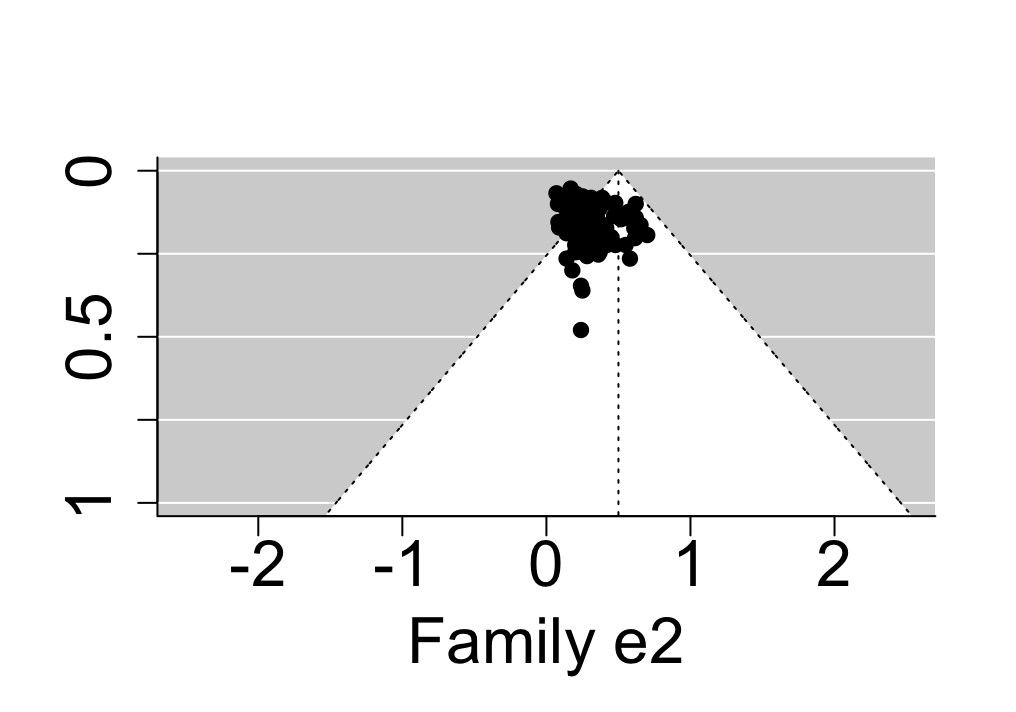 | 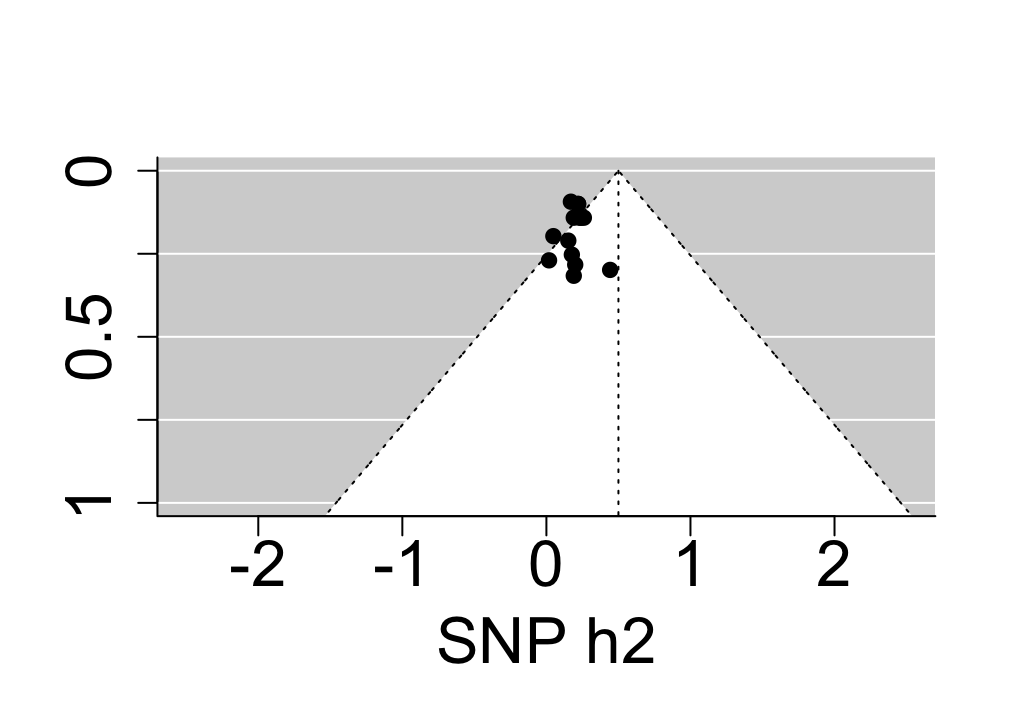 |

| **Supplementary Figure 10.** Funnel plots involving all studies addressing heritability and environmental influences on specific learning disorders. | |
| --- | --- |
| 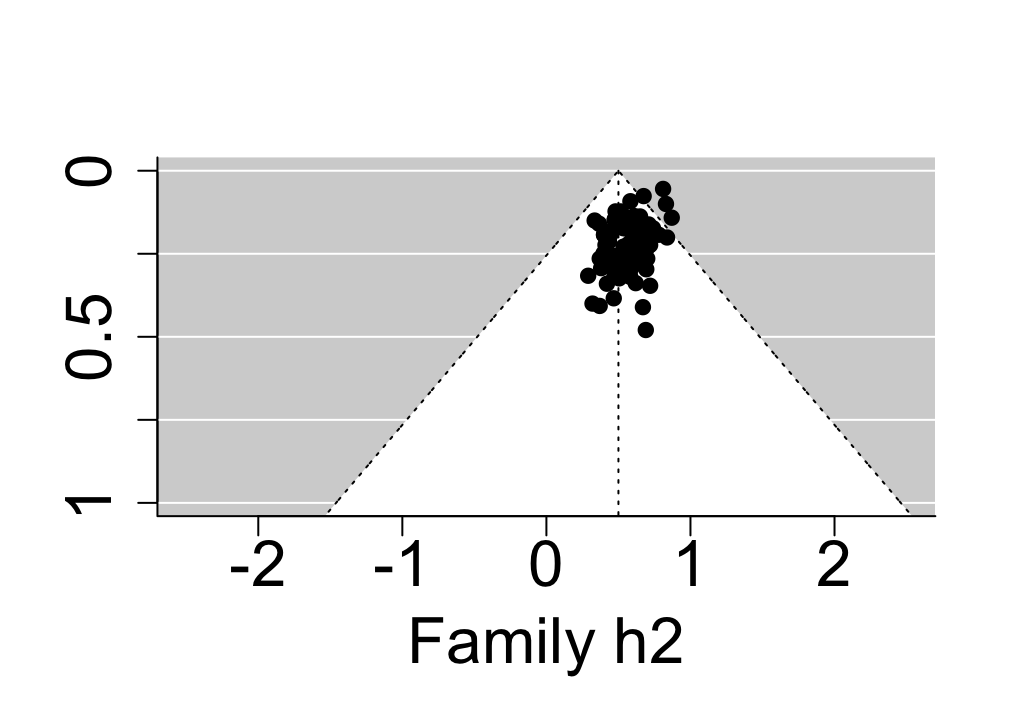 | 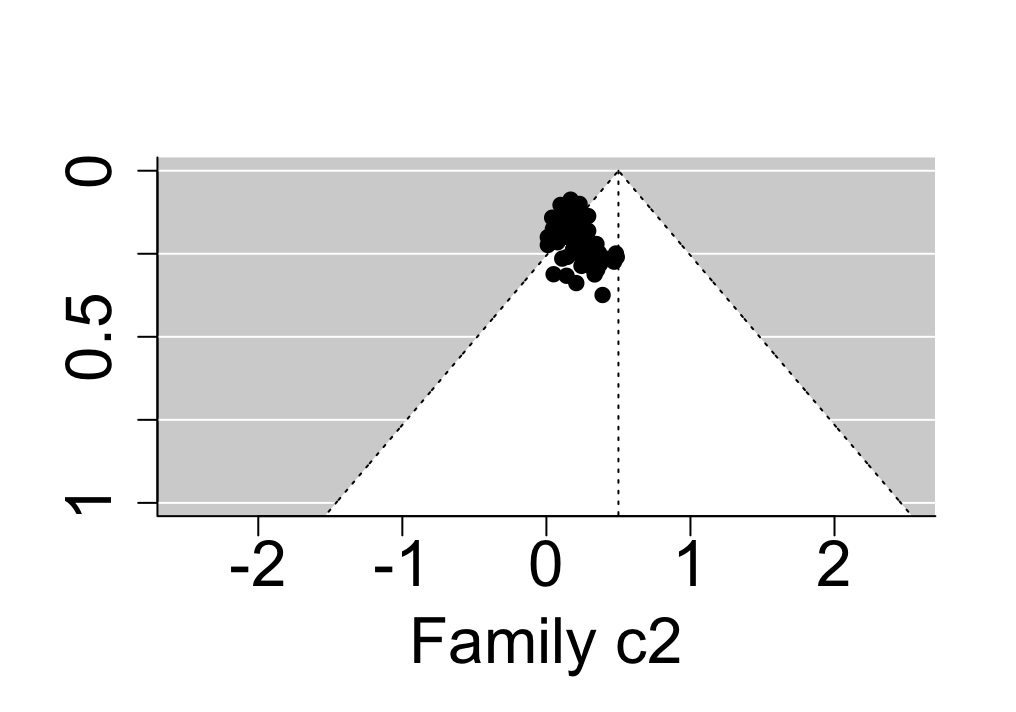 |
| 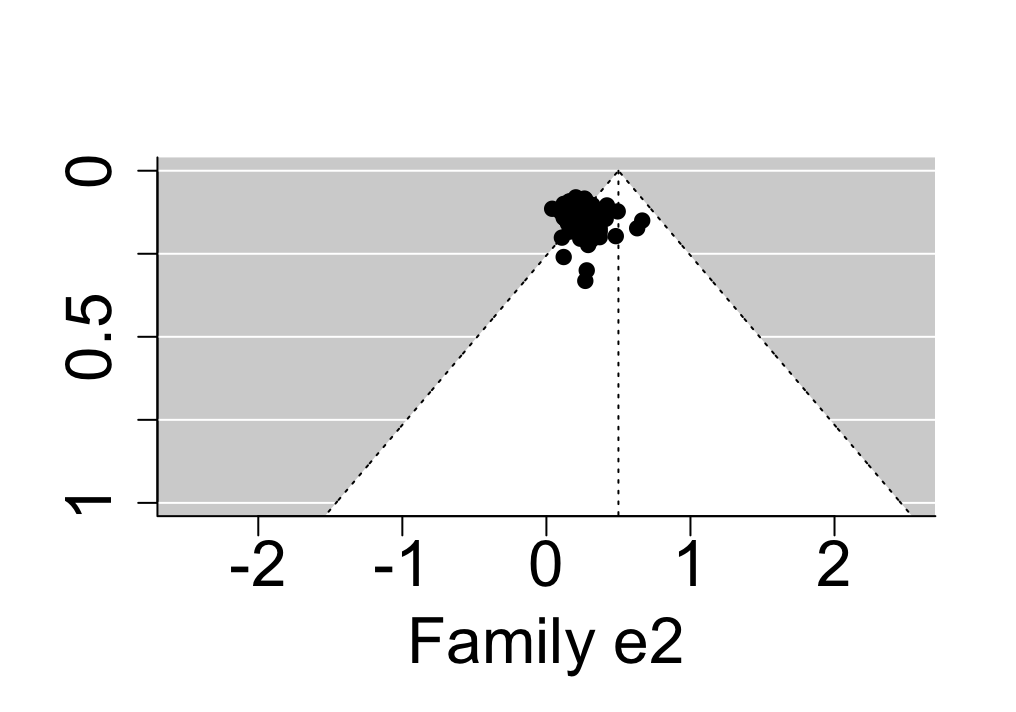 | 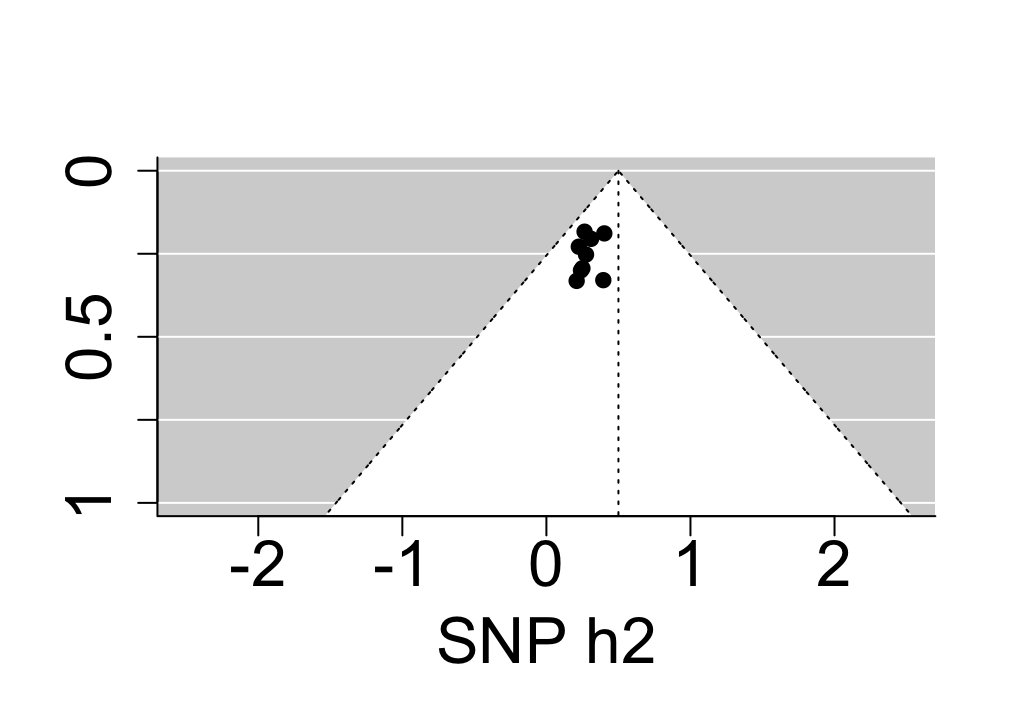 |

| **Supplementary Figure 11.** Funnel plots involving all studies addressing heritability and environmental influences on motor disorders. | |
| --- | --- |
| 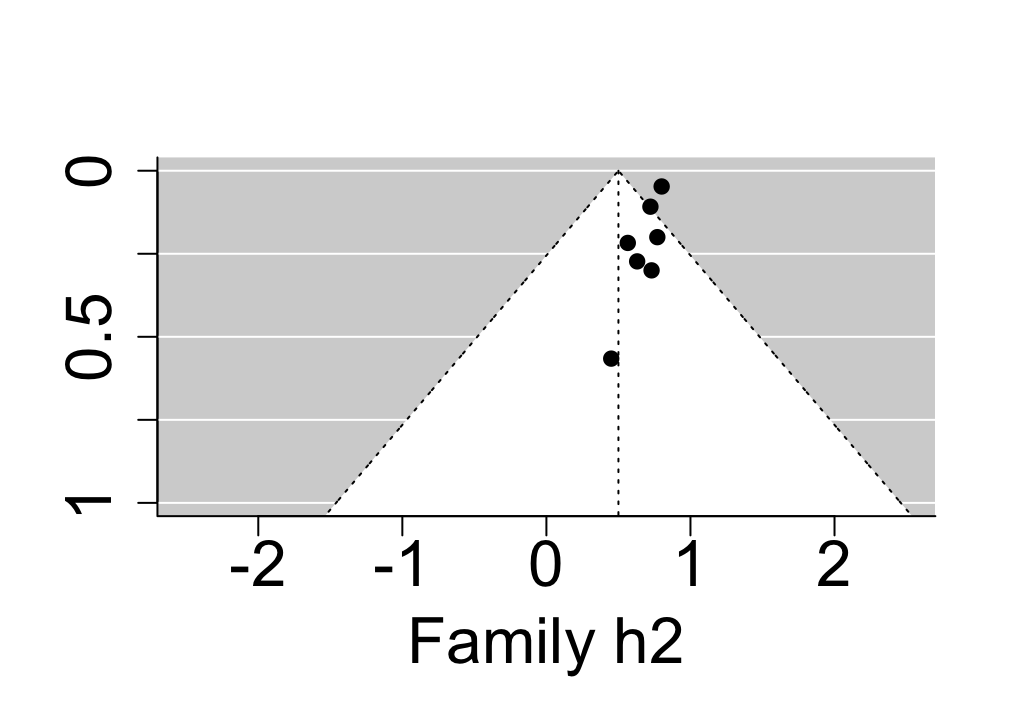 | 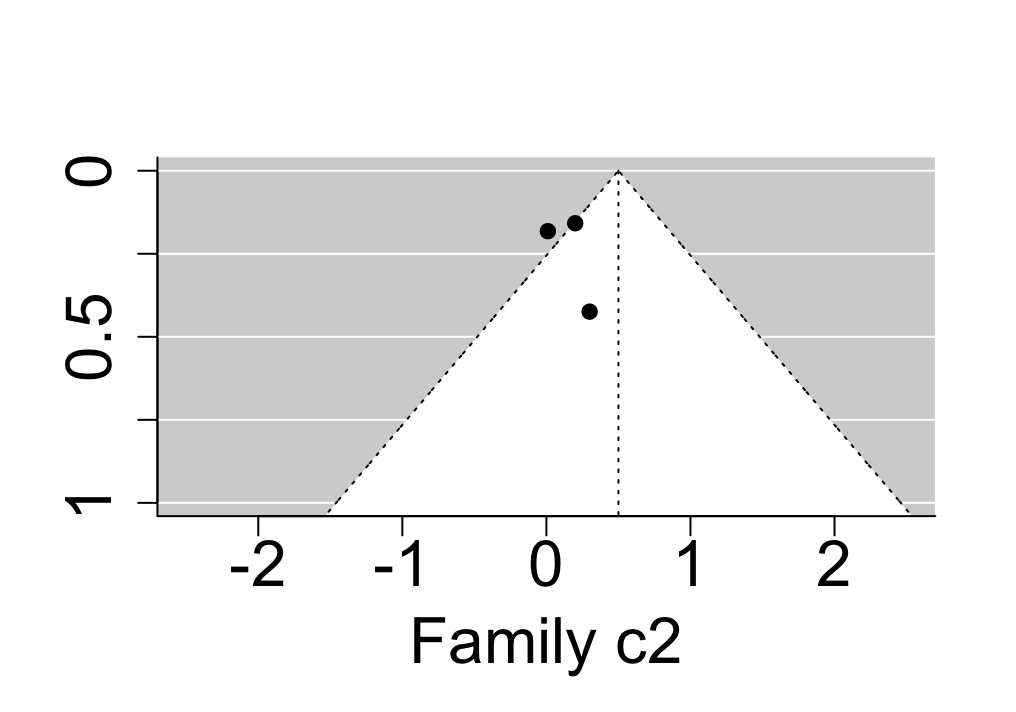 |
| 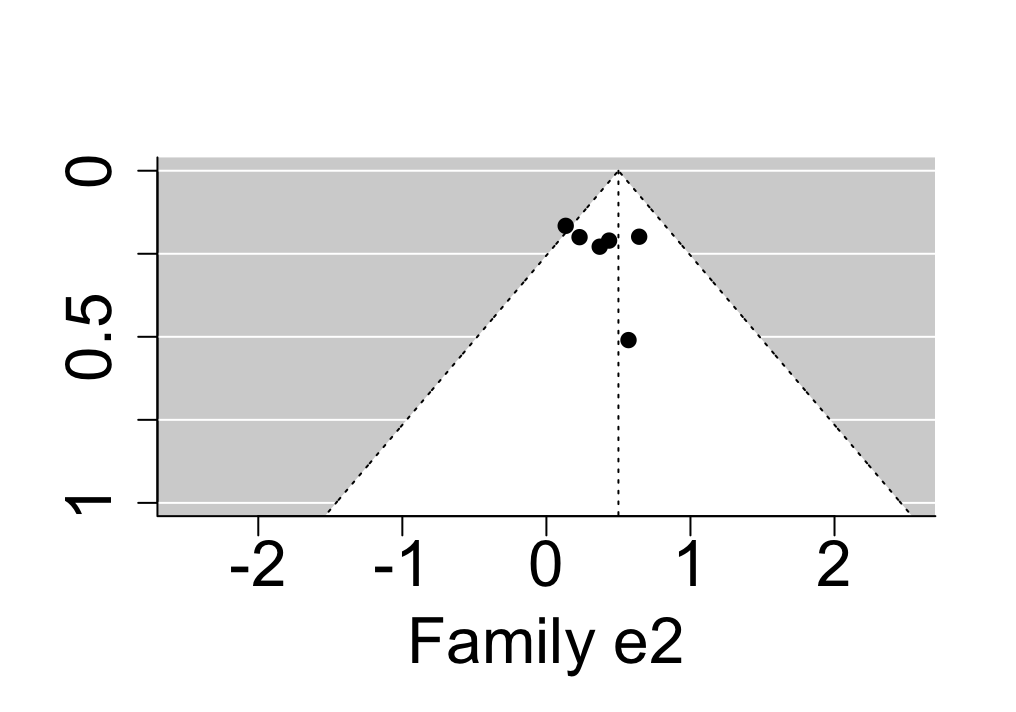 |  |

| **Supplementary Figure 12.** Funnel plots involving all studies addressing genetic and environmental overlap between neurodevelopmental disorders (NDDs). | |
| --- | --- |
| 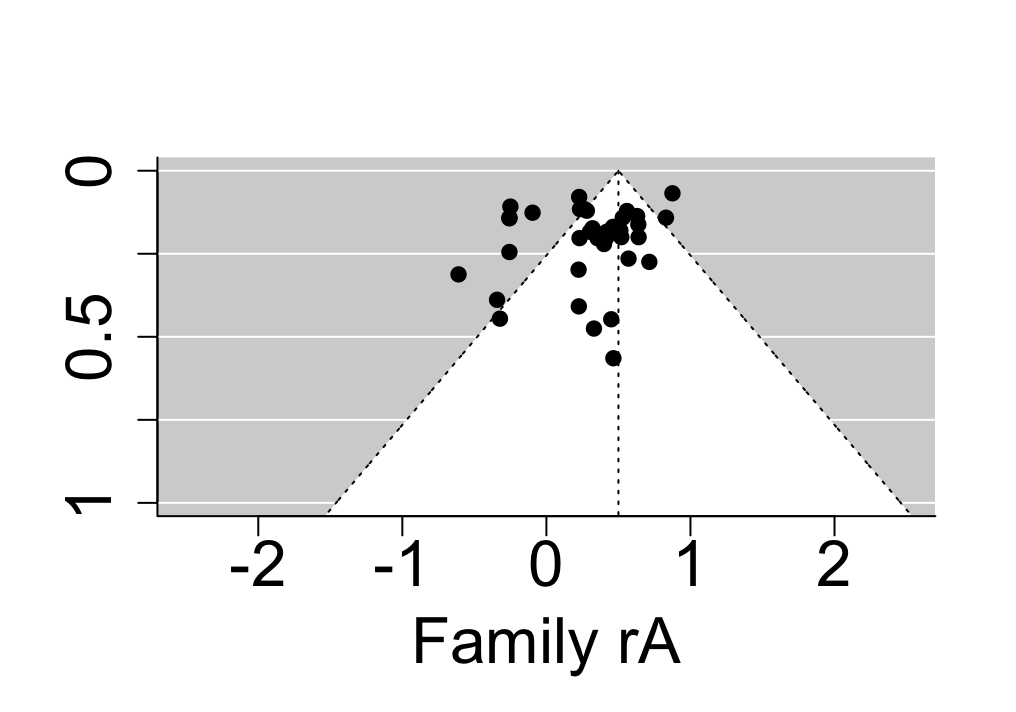 | 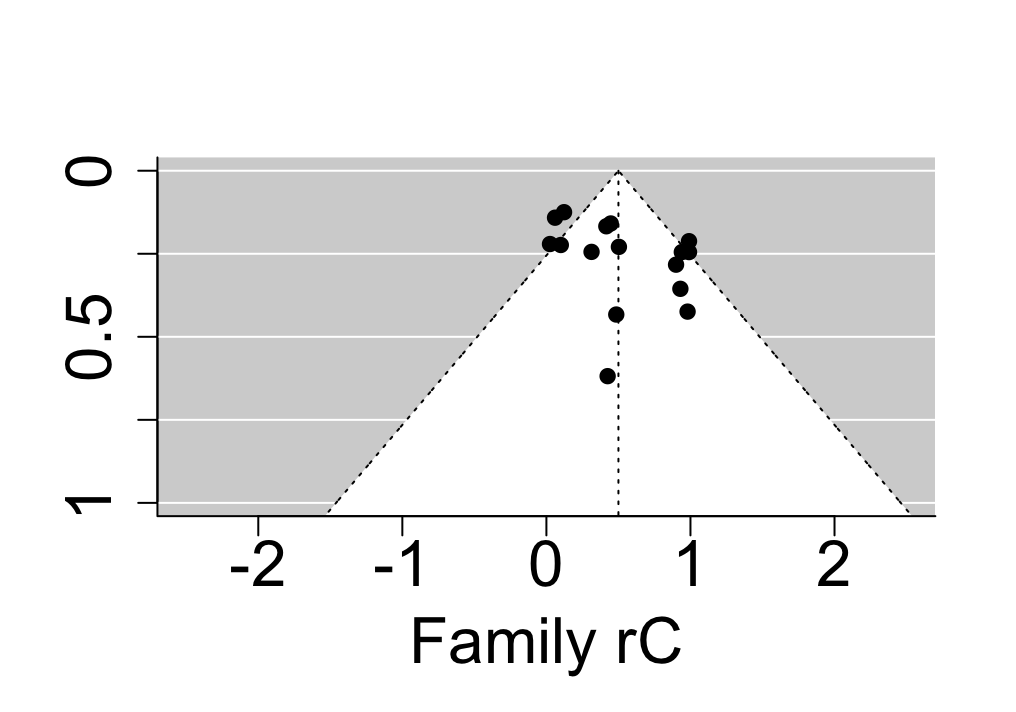 |
| 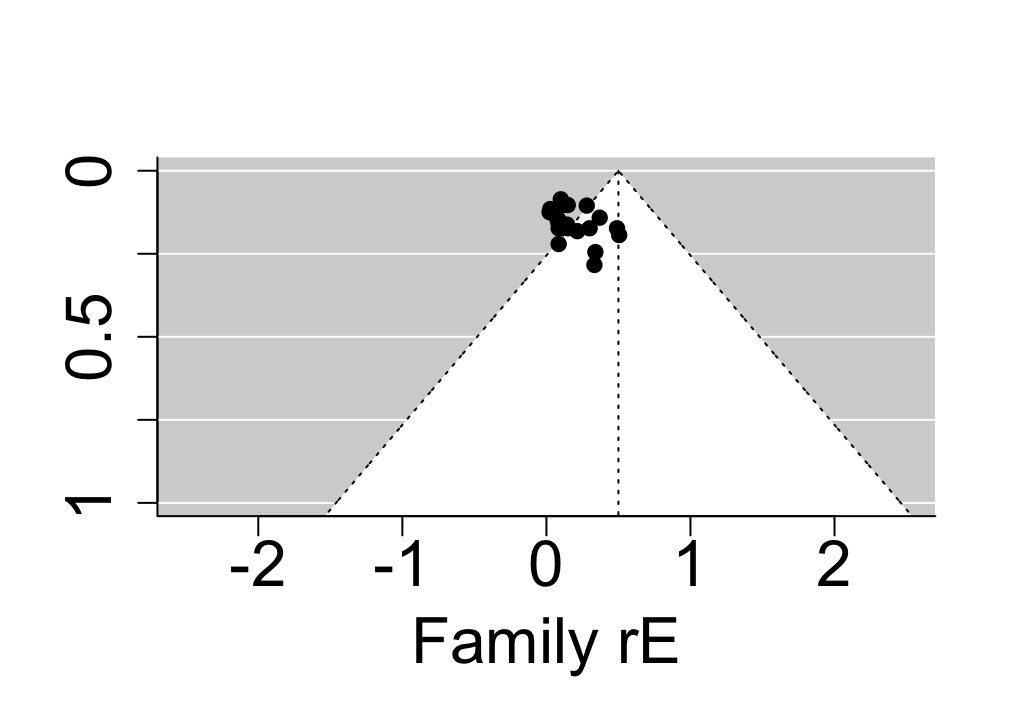 | 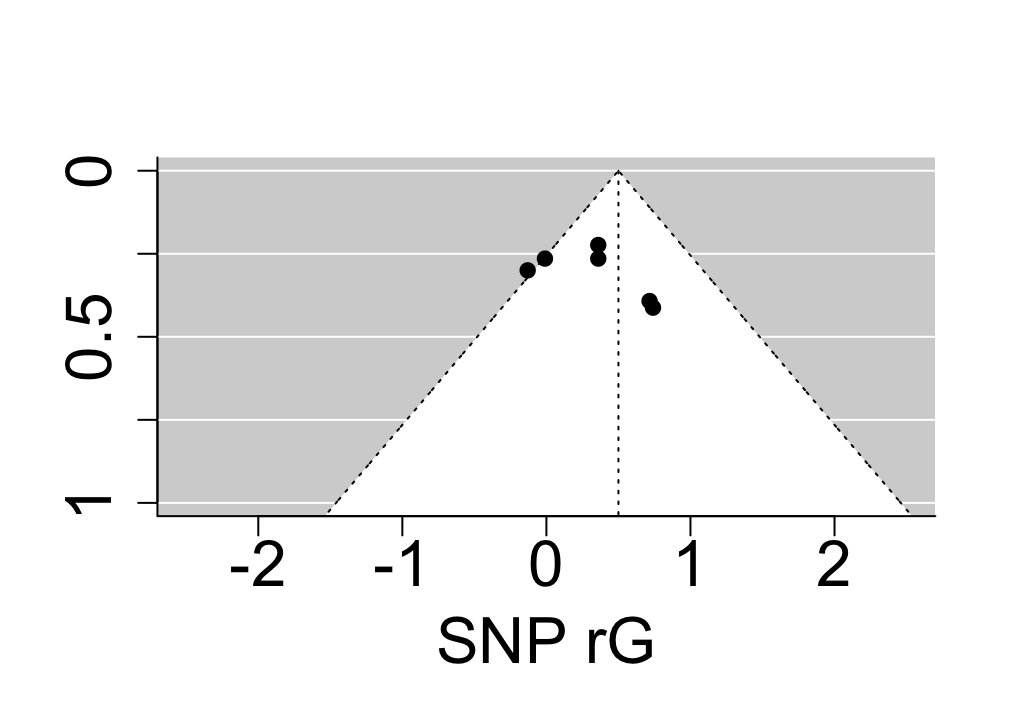 |

| **Supplementary Figure 13.** Funnel plots involving all studies addressing genetic and environmental overlap between ASD & ADHD. | |
| --- | --- |
| 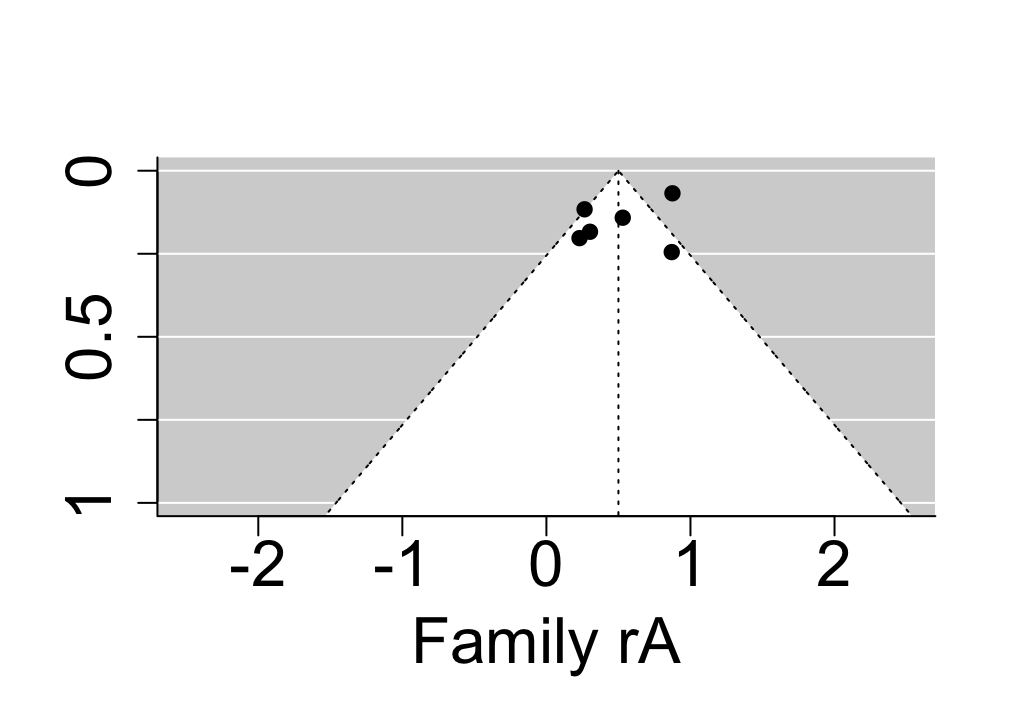 | 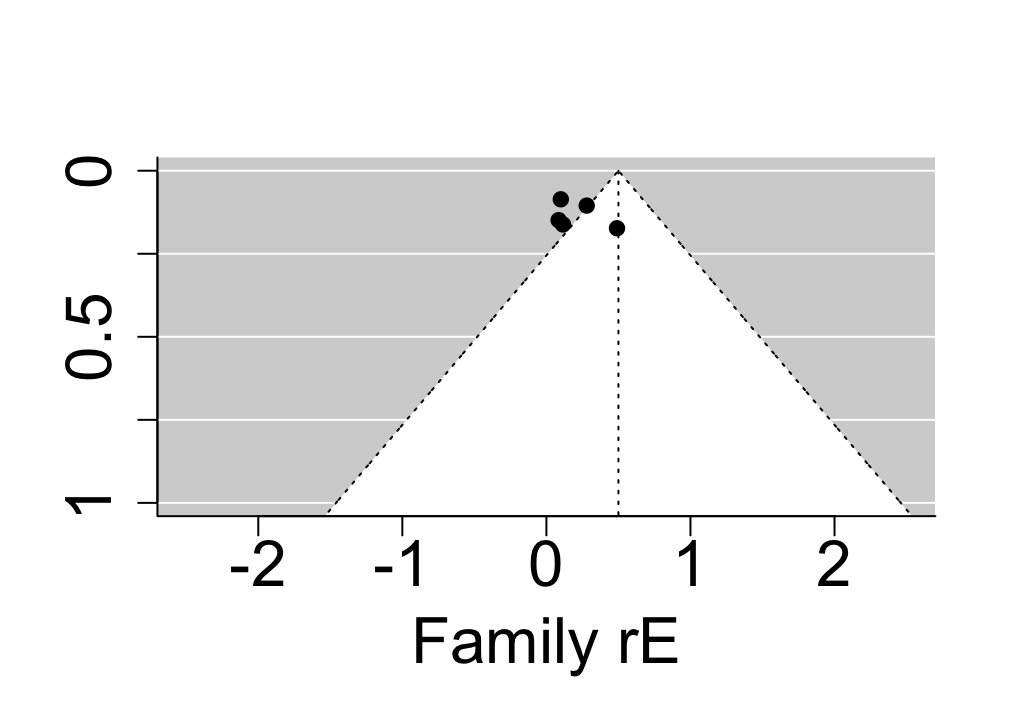 |
| 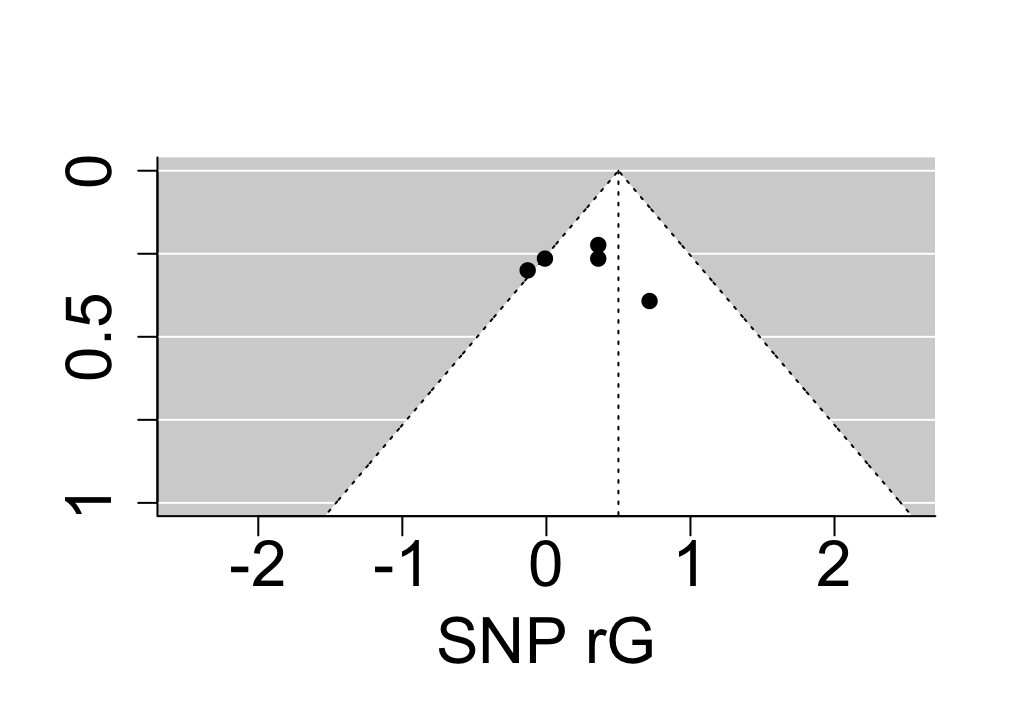 |  |

| **Supplementary Figure 14.** Funnel plots involving all studies addressing genetic and environmental overlap between ADHD & motor disorders. |
| --- |
| 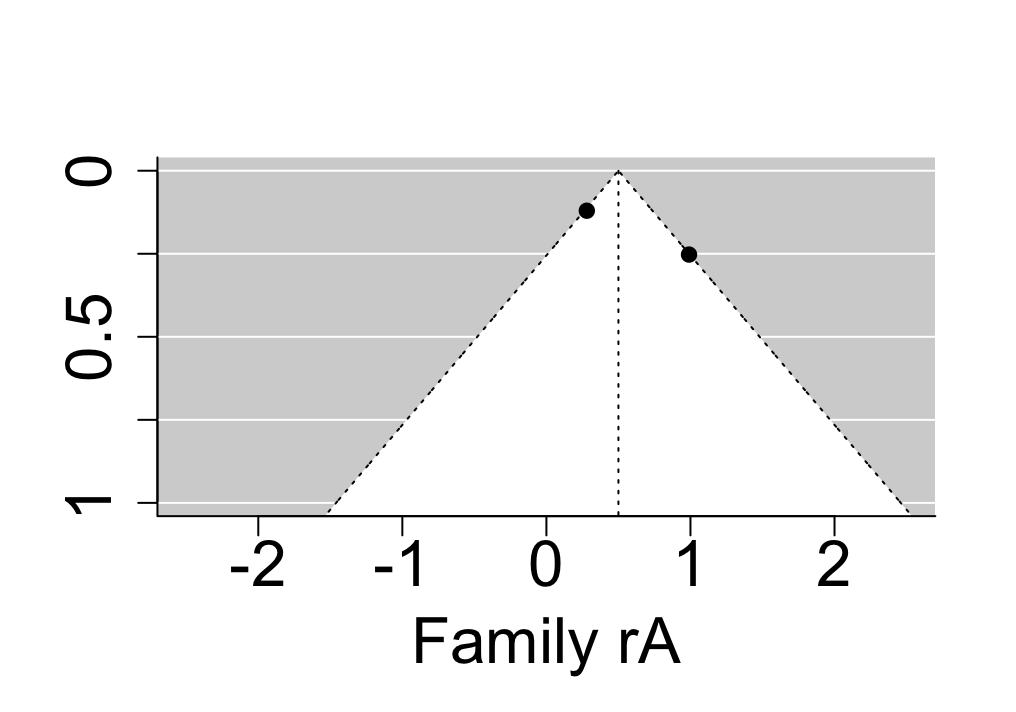 |

| **Supplementary Figure 15.** Funnel plots involving all studies addressing genetic and environmental overlap between ADHD & specific learning disorders. | |
| --- | --- |
| 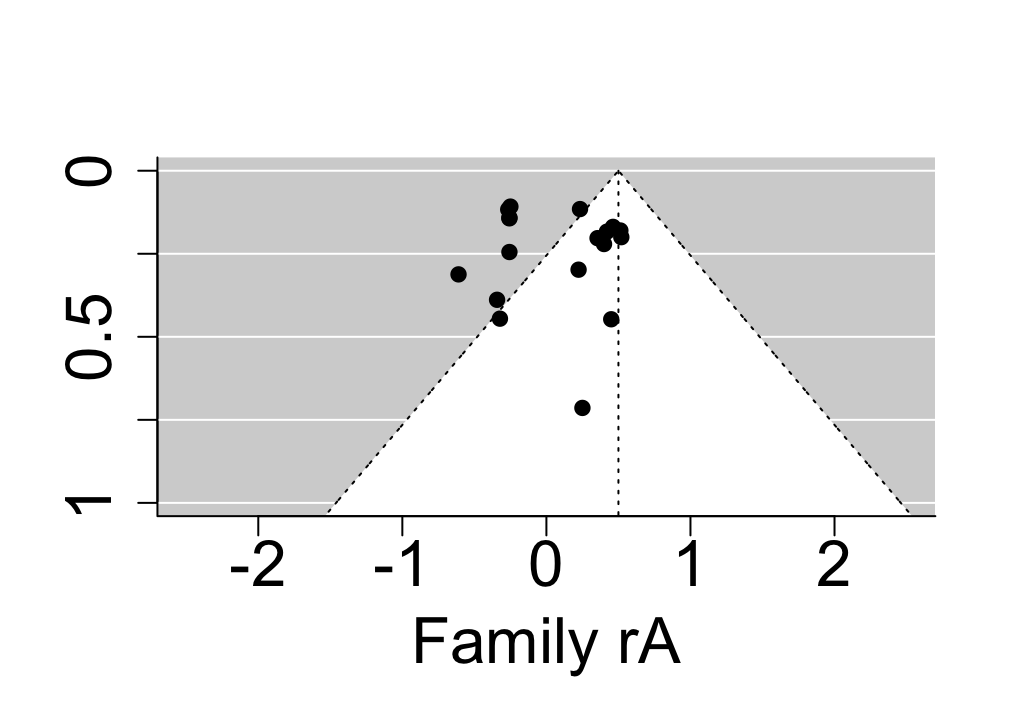 | 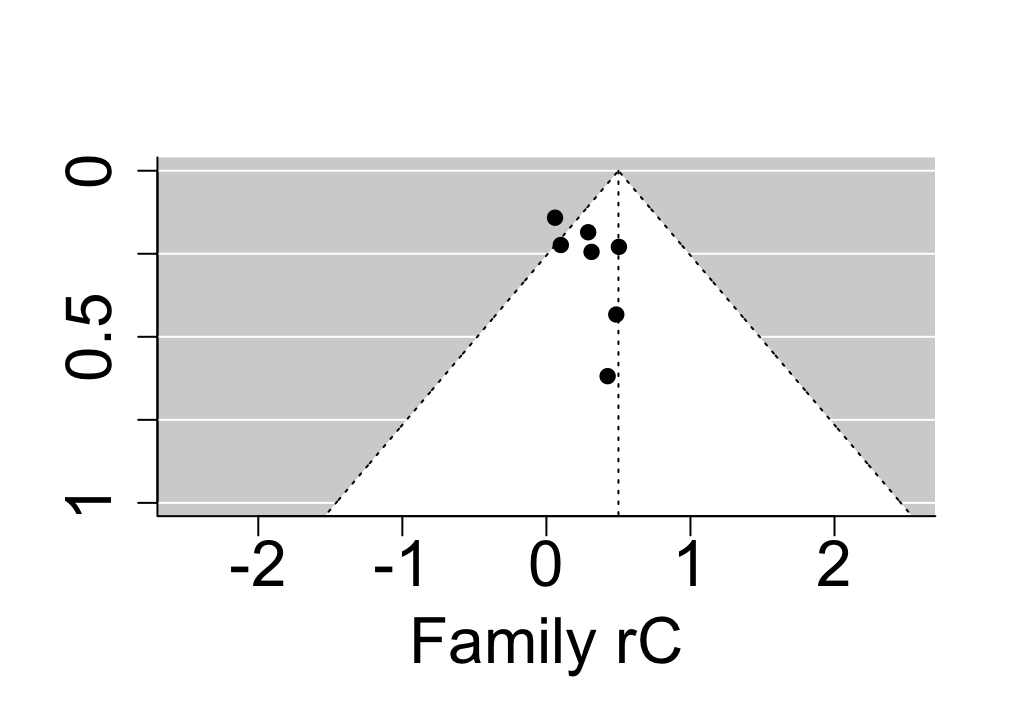 |
| 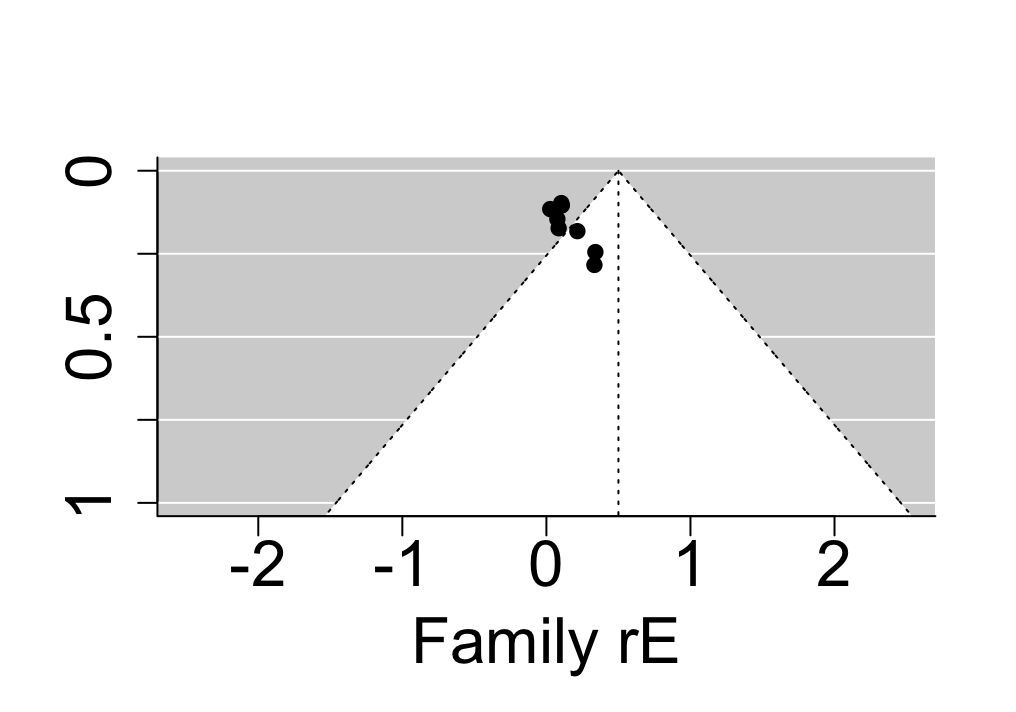 |  |

| **Supplementary Figure 16.** Funnel plots involving all studies addressing genetic and environmental overlap between communication disorders & motor disorders. |
| --- |
| 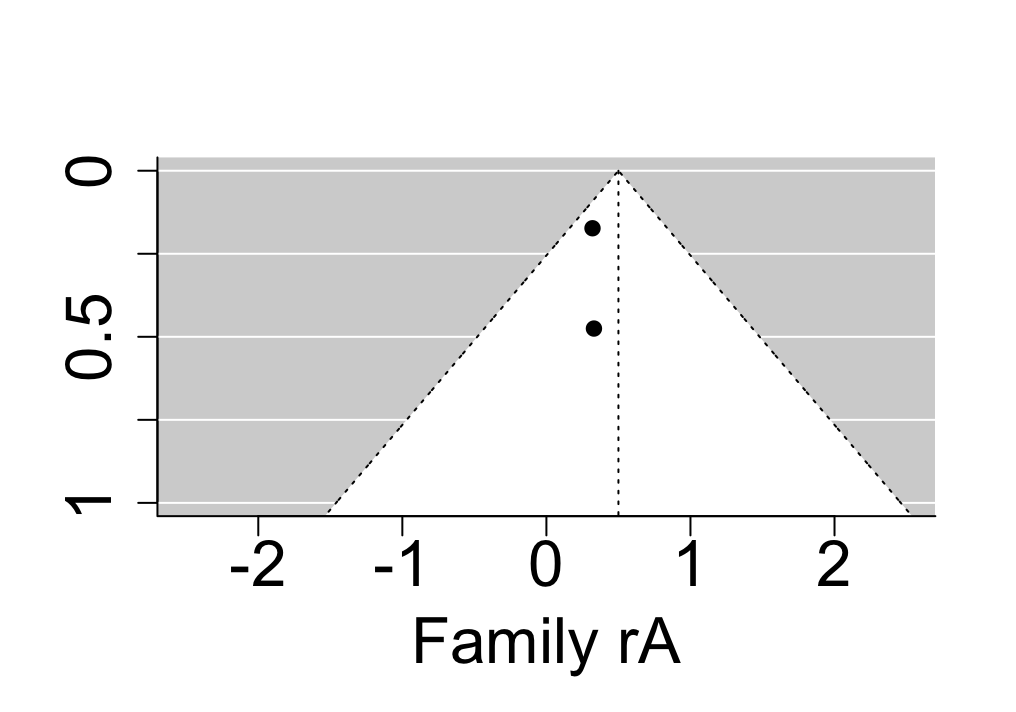 |

| **Supplementary Figure 17.** Funnel plots involving all studies addressing genetic and environmental overlap between communication disorders & specific learning disorders. |
| --- |
| 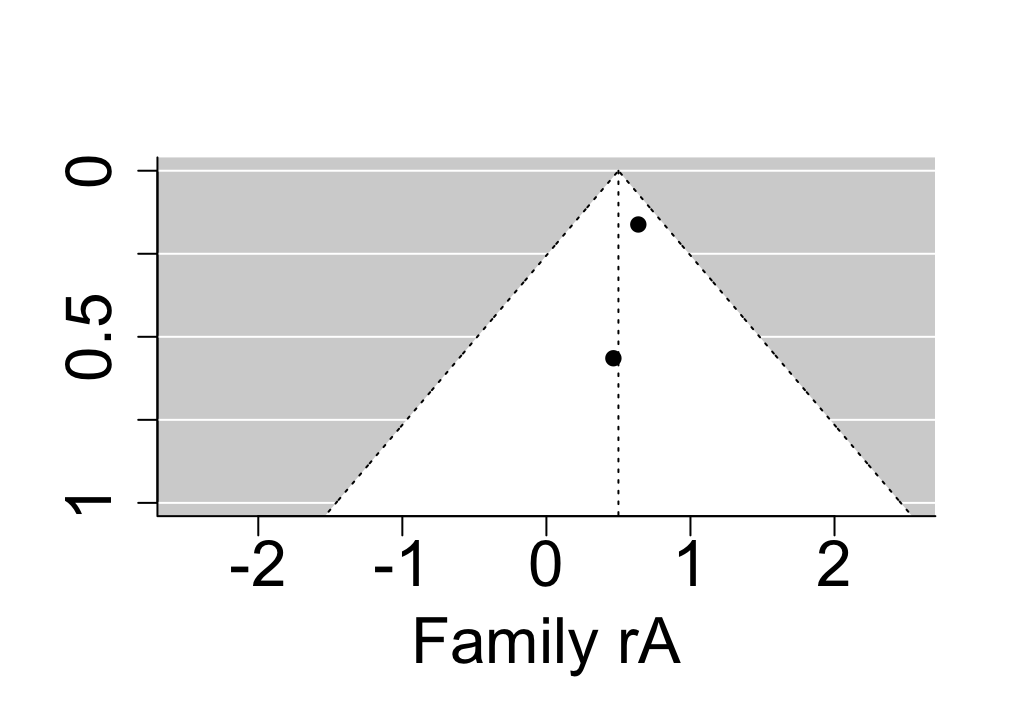 |

| **Supplementary Figure 18.** Funnel plots involving all studies addressing genetic and environmental overlap between neurodevelopmental disorders (NDDs) and disruptive, impulse control and conduct disorders (DICCs). | |
| --- | --- |
| 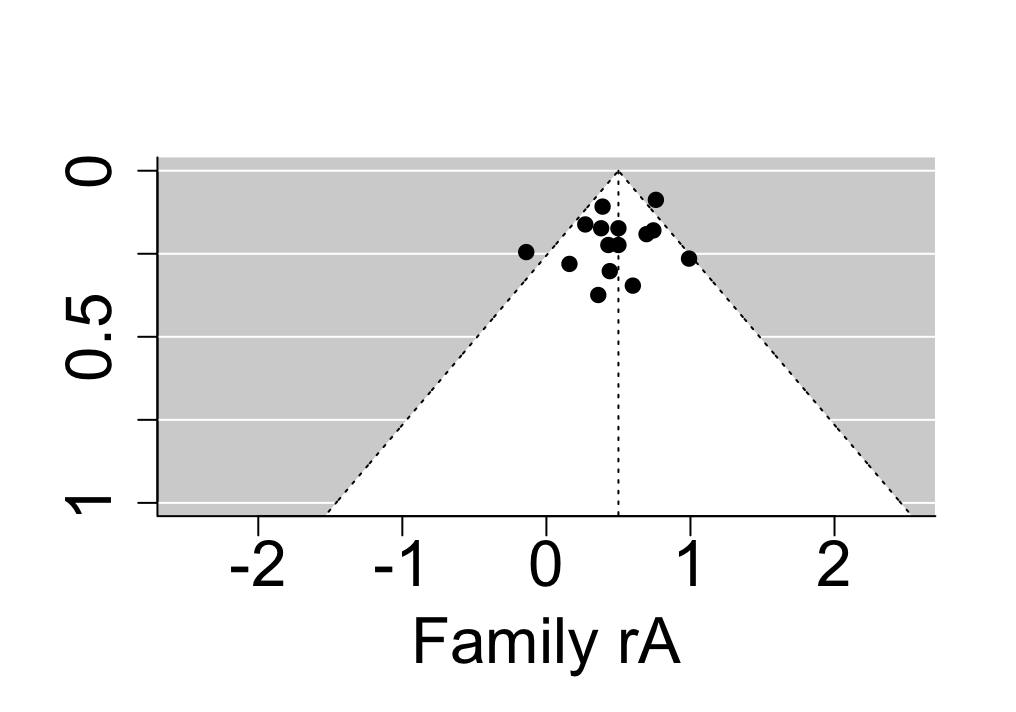 | 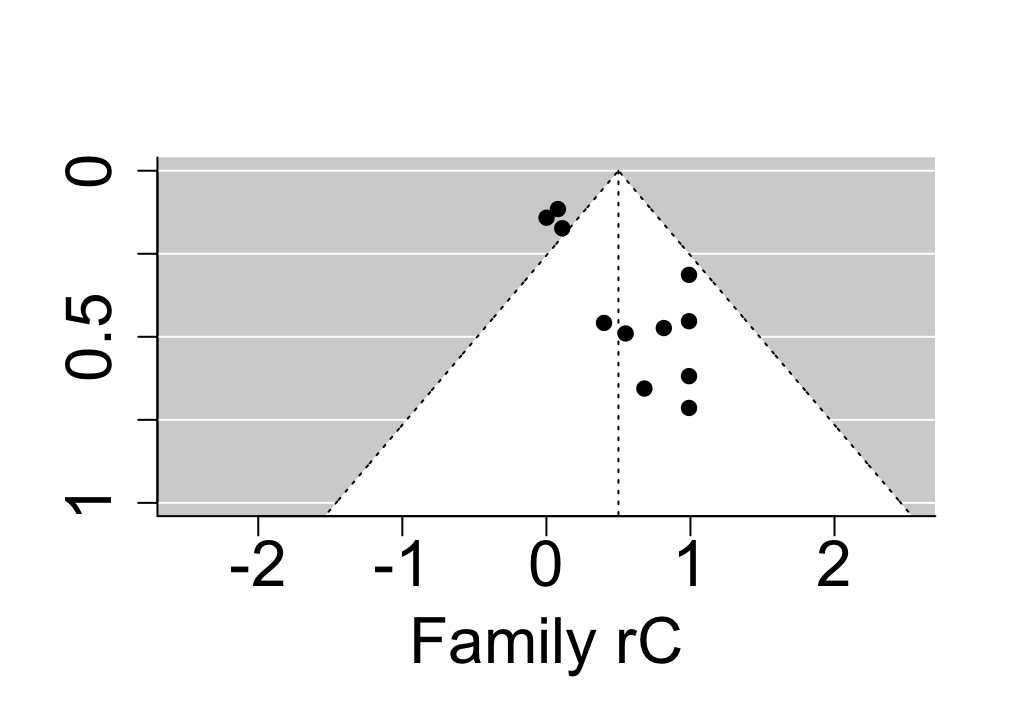 |
| 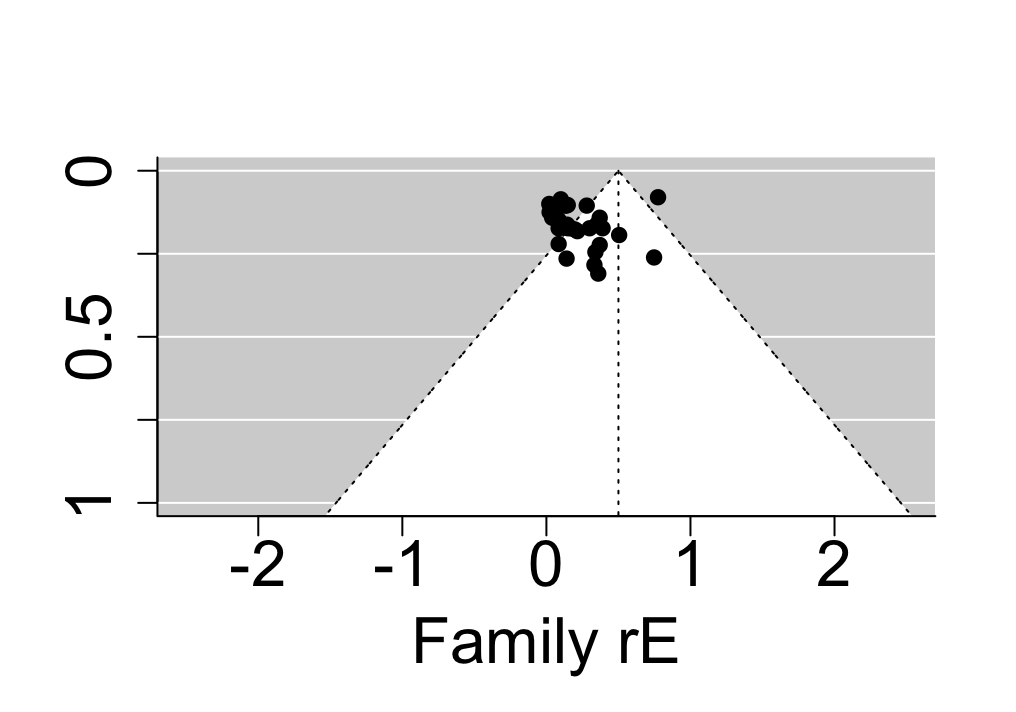 |  |

| **Supplementary Figure 19.** Funnel plots involving all studies addressing genetic and environmental overlap between ADHD & conduct disorder. | |
| --- | --- |
| 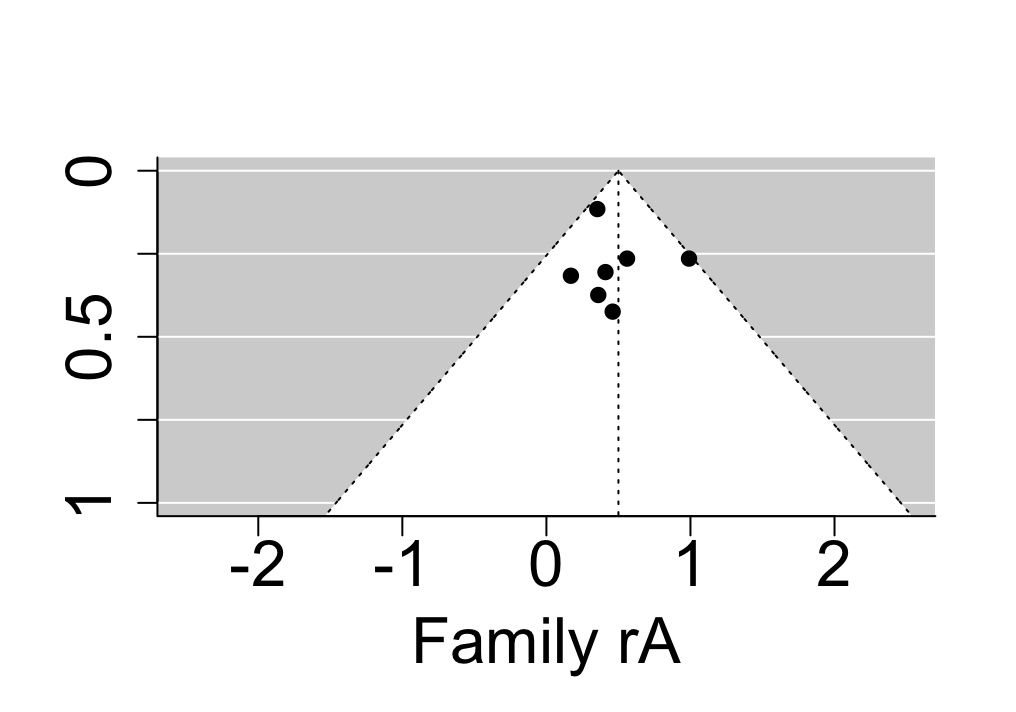 | 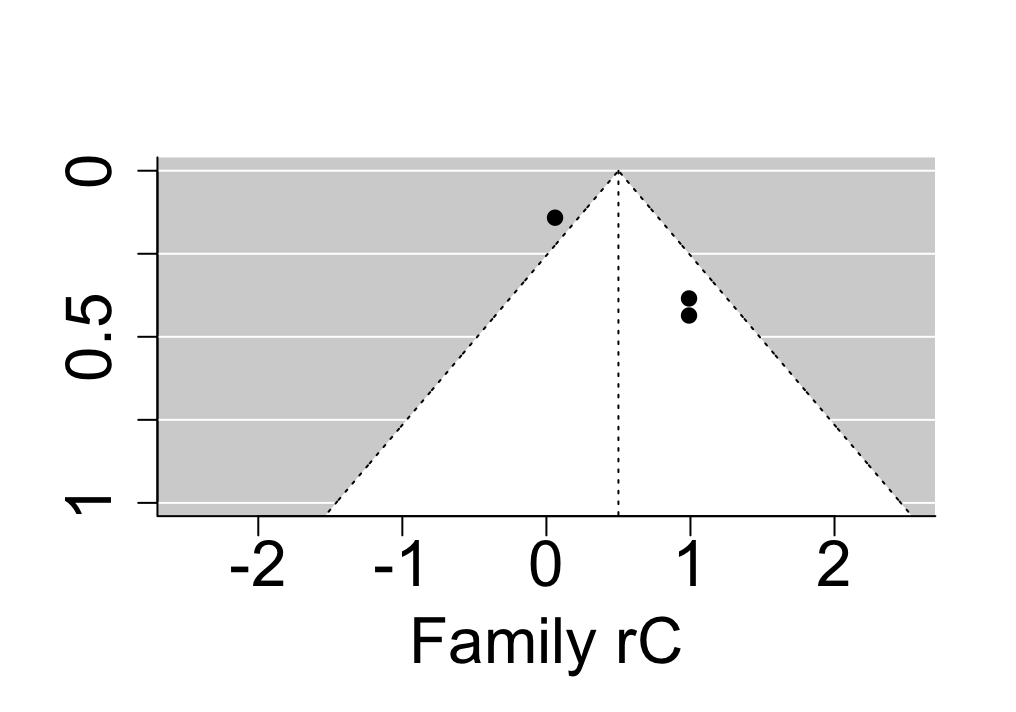 |
| 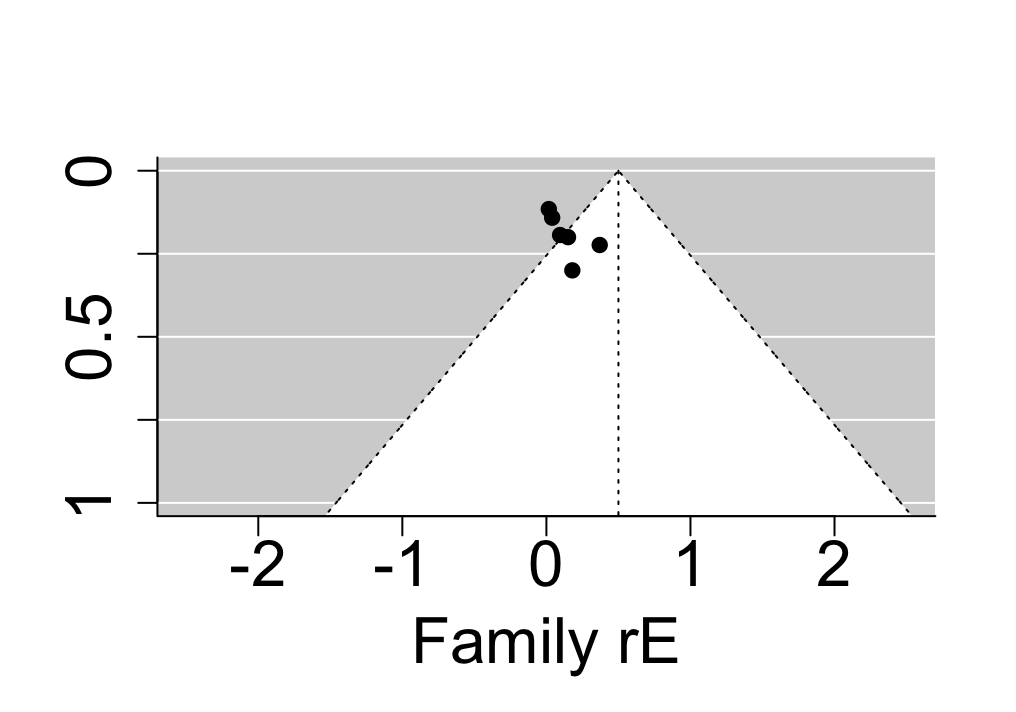 |  |

| **Supplementary Figure 20.** Funnel plots involving all studies addressing genetic and environmental overlap between ADHD & oppositional defiant disorder. | |
| --- | --- |
| 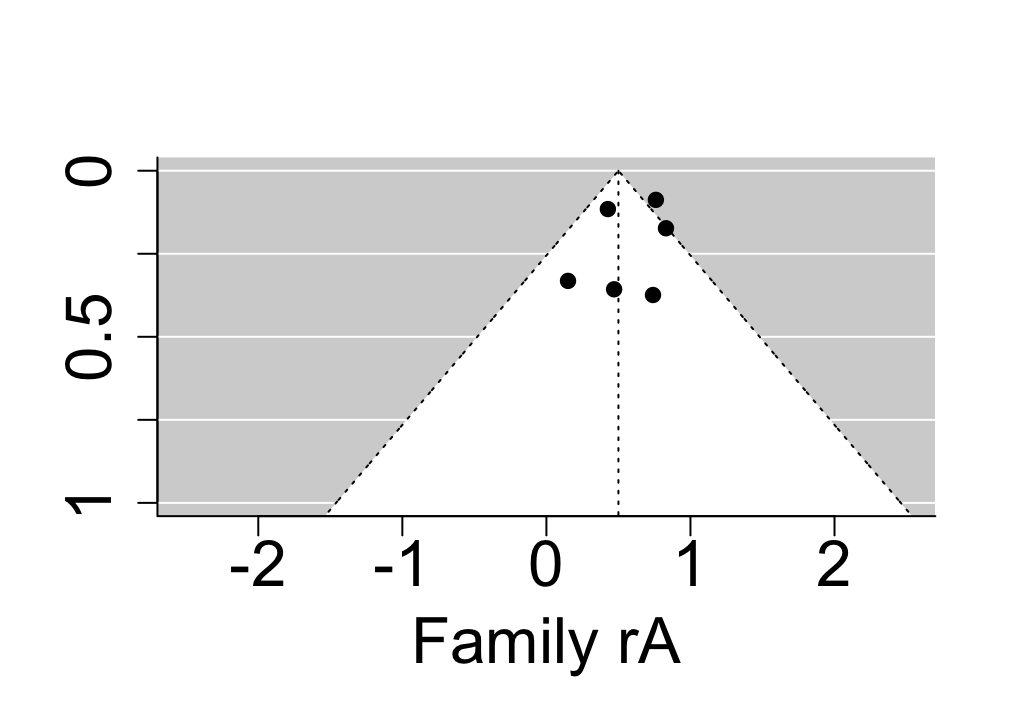 | 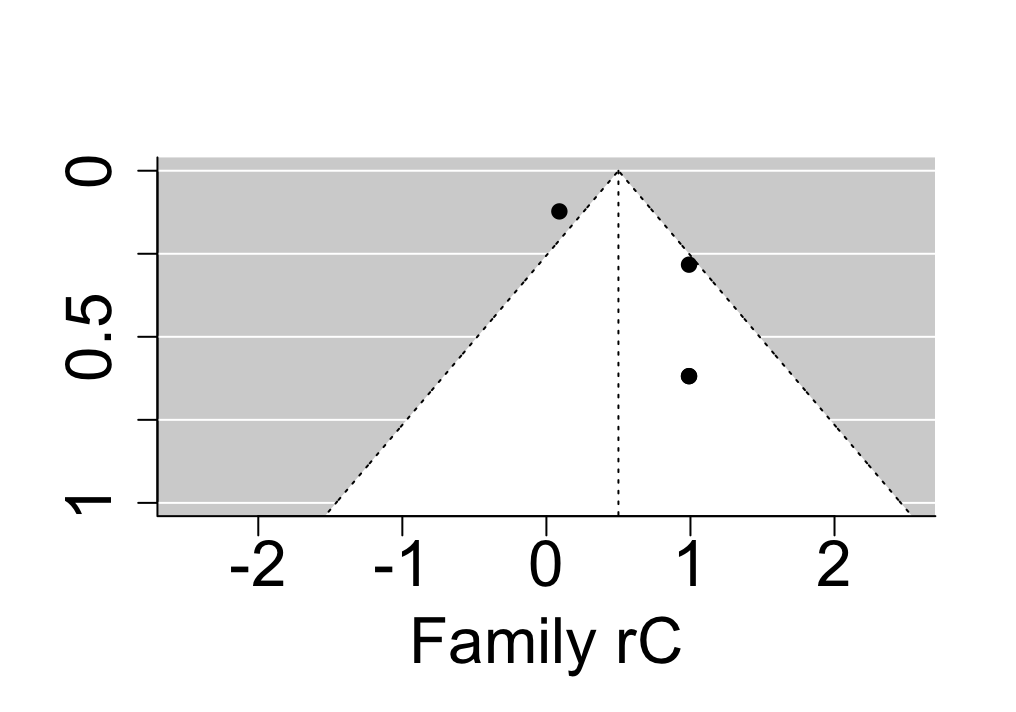 |
| 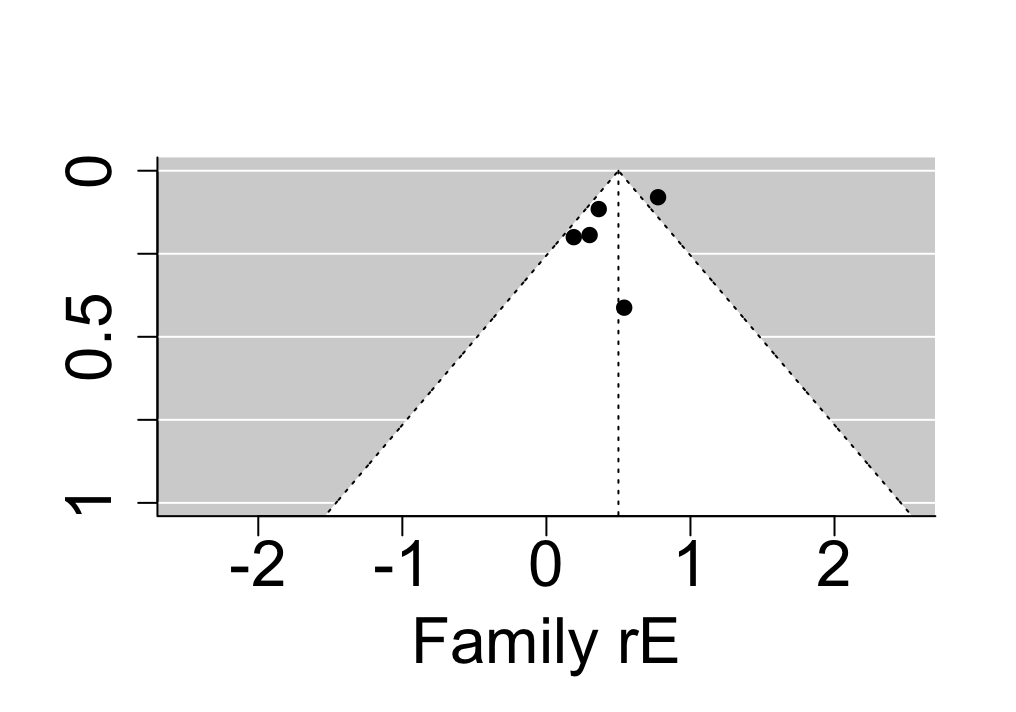 |  |

| **Supplementary Figure 21.** Funnel plots involving all studies addressing genetic and environmental overlap between ASD & conduct disorder. | |
| --- | --- |
| 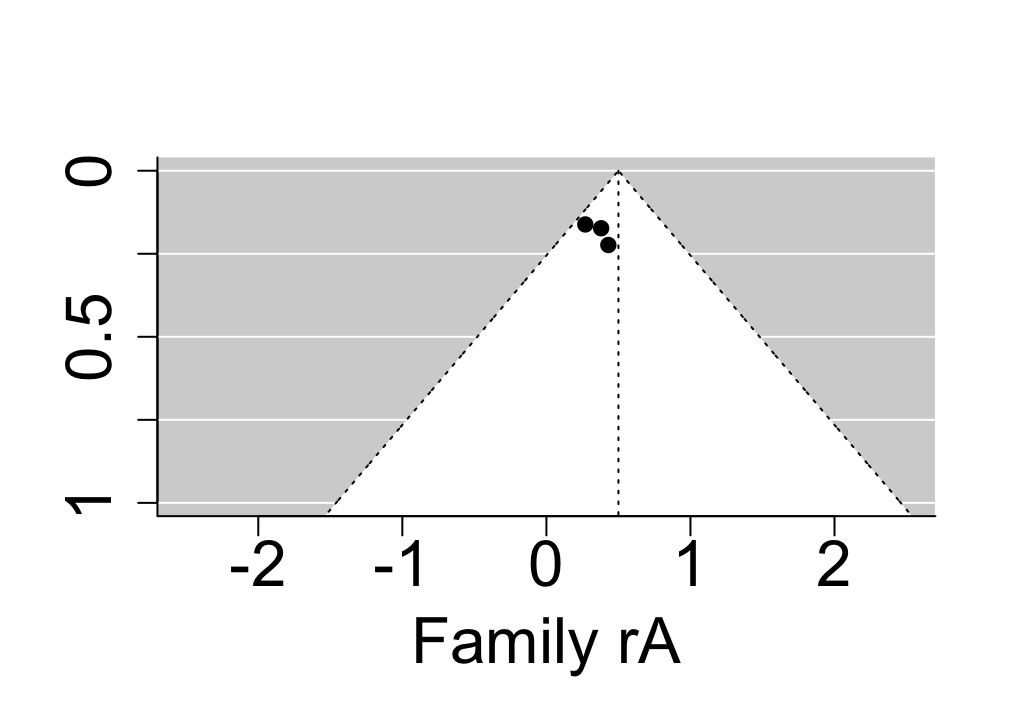 | 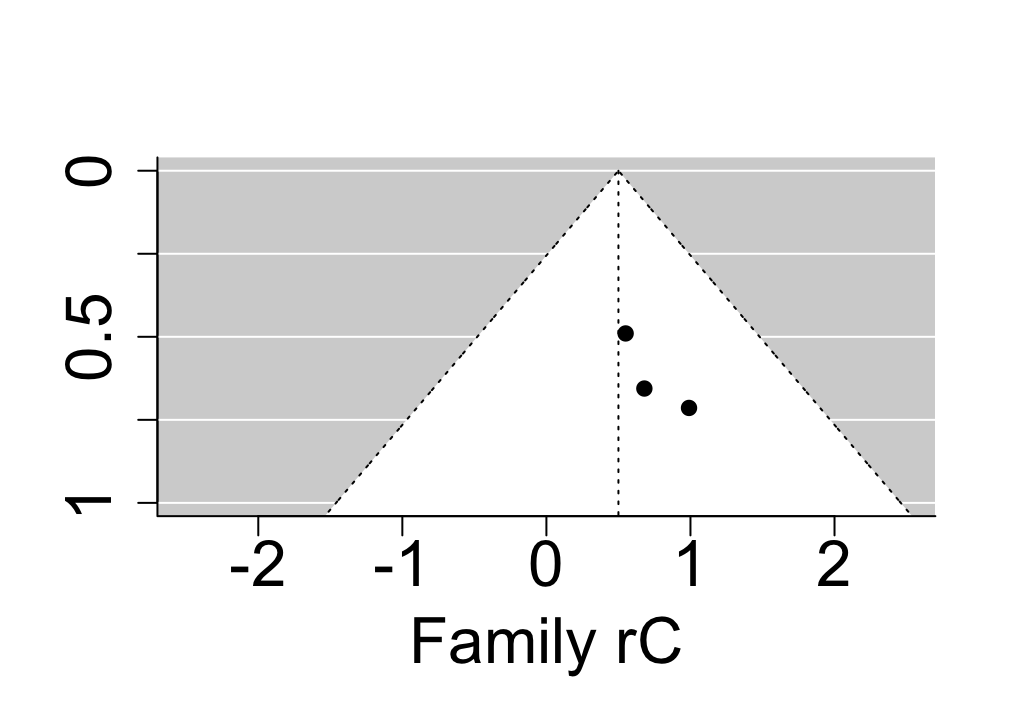 |
| 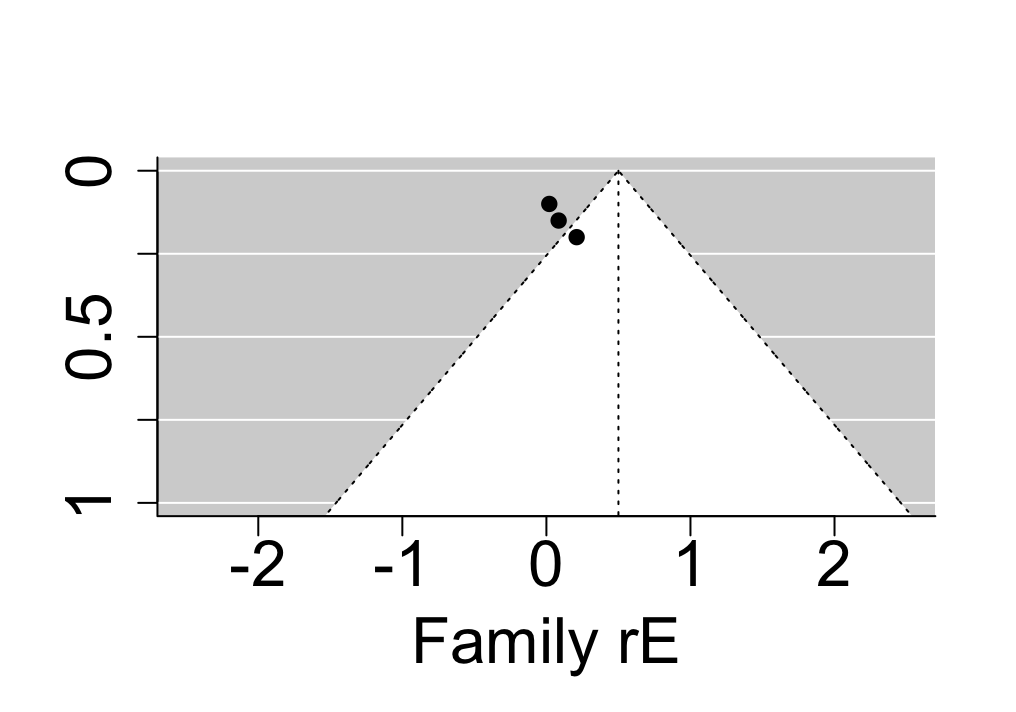 |  |

**Supplementary Figure 22**. Sources of variation in neurodevelopmental disorders (NDDs) (top panel), genetic and environmental overlap between NDDs (middle panel) and genetic and environmental overlap between NDDs and disruptive, impulse control and conduct disorders (DICCs) (bottom panel), stratified by measurement scales, i.e., categorical versus continuous measurement.

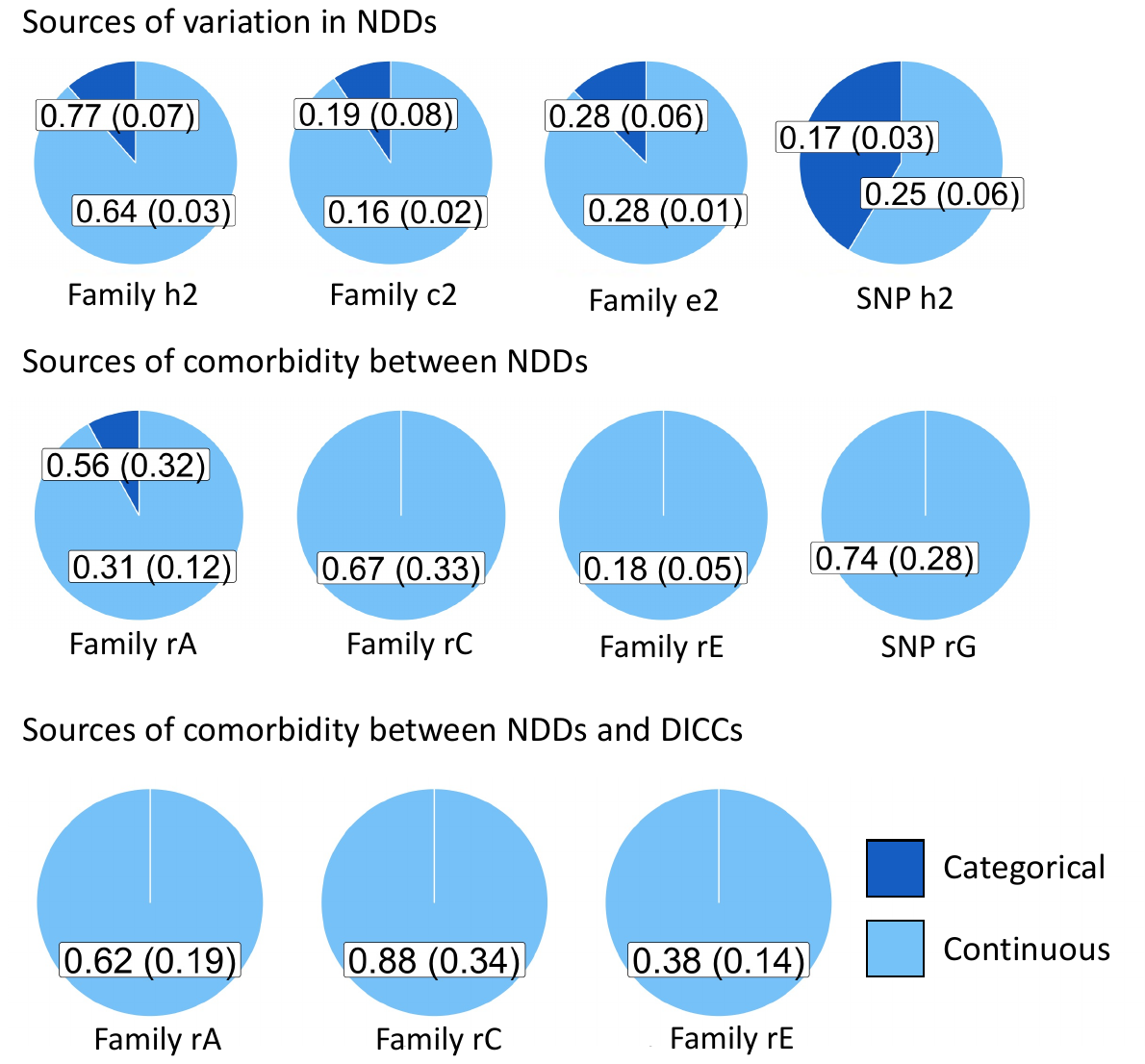

| **Supplementary Figure 23.** Diagram of searches and screening. Panel **A** shows study selection workflow of the primary search and Panel **B** shows workflow of the confirmatory search. | |
| --- | --- |
| **A** | **B** |
| 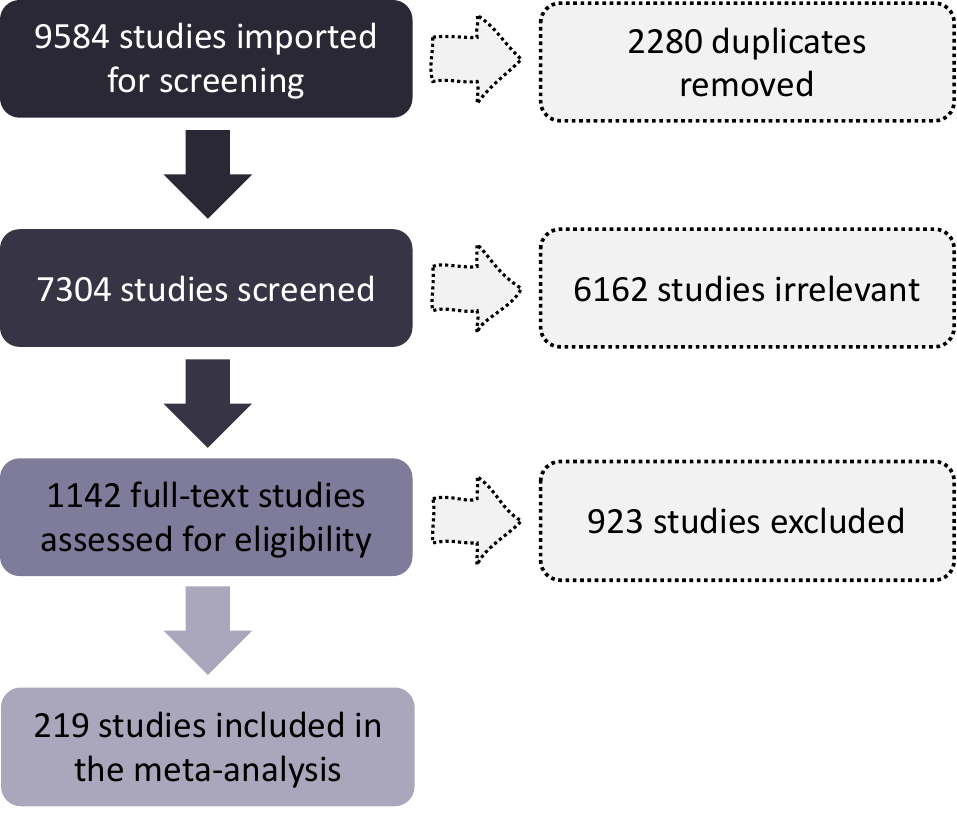 | 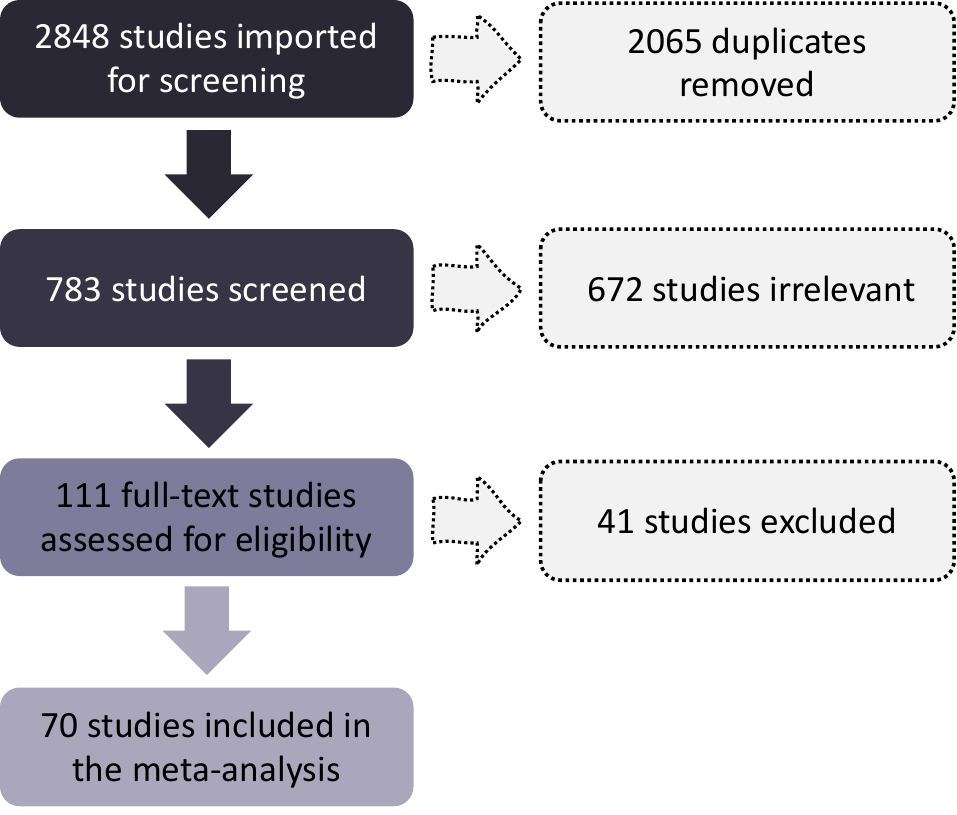 |

**Supplementary Figure 24**. Grand heritability and environmental influences across all neurodevelopmental disorders (NDDs) (panel **A**), grand genetic and environmental correlations across all NDDs (panel **B**) and grand genetic and environmental correlations across NDDs and disruptive, impulse control and conduct disorders (DICCs) (panel **C**) obtained using different aggregation techniques, i.e., aggregating by study, cohort, and country, using correlation thresholds of r= 0.3, r= 0.5 and r= 0.9.

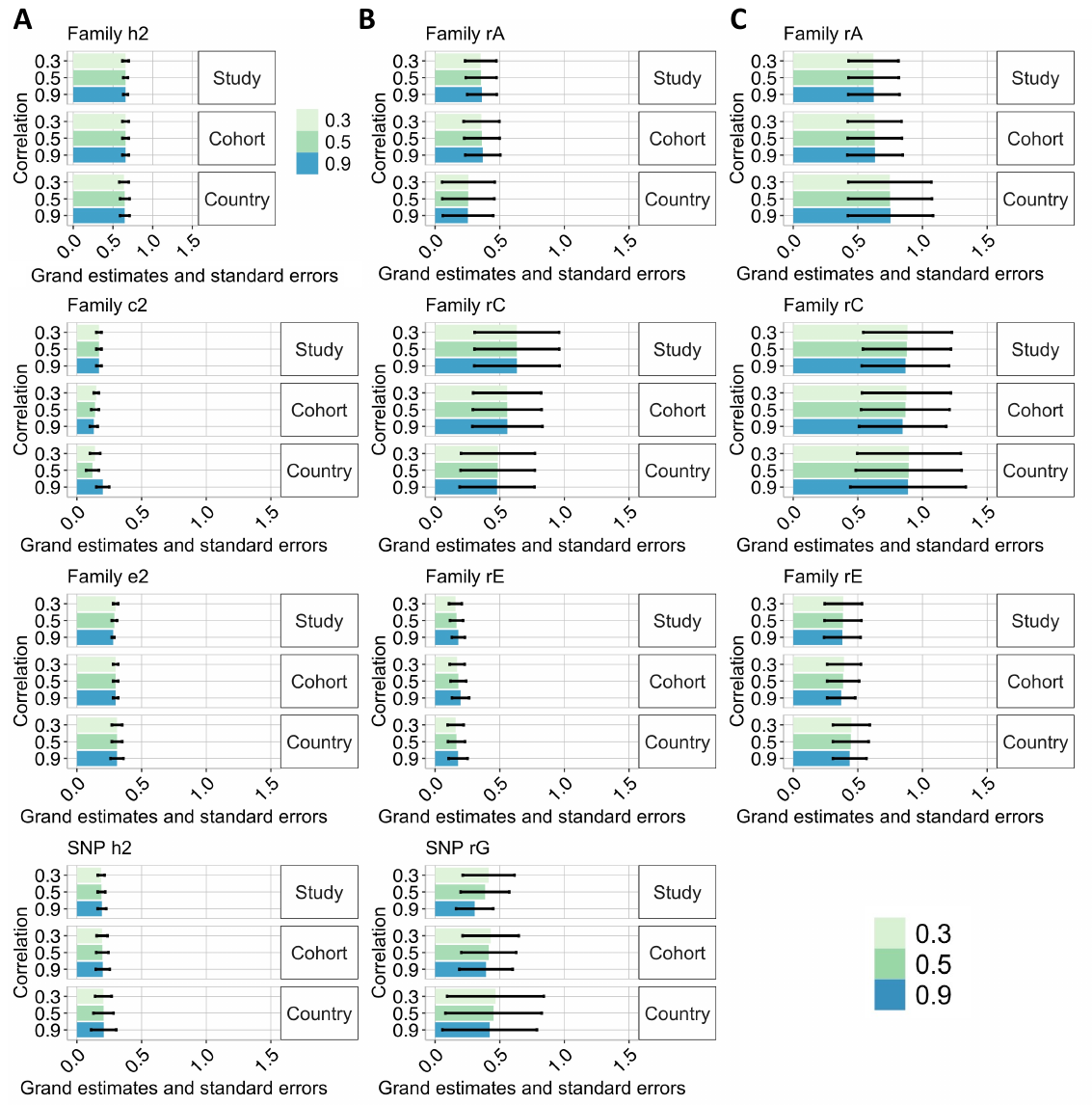

***Supplementary references***

1.	Lichtenstein, P., Carlström, E., Råstam, M., Gillberg, C. & Anckarsäter, H. The genetics of autism spectrum disorders and related neuropsychiatric disorders in childhood. *Am. J. Psychiatry* **167**, 1357–1363 (2010).

2.	McGue, M. & Bouchard, T. J. Adjustment of twin data for the effects of age and sex. *Behav. Genet.* **14**, 325–343 (1984).

3.	Abdellaoui, A. *et al.* Genetic correlates of social stratification in Great Britain. *Nat. Hum. Behav.* **3**, 1332–1342 (2019).

4.	Yang, J., Lee, S. H., Goddard, M. E. & Visscher, P. M. GCTA: A tool for genome-wide complex trait analysis. *Am. J. Hum. Genet.* **88**, 76–82 (2011).

5.	Bulik-Sullivan, B. K. *et al.* LD Score regression distinguishes confounding from polygenicity in genome-wide association studies. *Nat. Genet.* **47**, 291–295 (2015).

6.	Zeng, J. *et al.* Signatures of negative selection in the genetic architecture of human complex traits. *Nat. Genet.* **50**, 746–753 (2018).

7.	Kmet, L. M., Cook, L. S. & Lee, R. C. Standard quality assessment criteria for evaluating primary research papers from a variety of fields. (2004).

8.	Davis, L. K. *et al.* Partitioning the heritability of Tourette syndrome and obsessive compulsive disorder reveals differences in genetic architecture. *PLoS Genet.* **9**, e1003864 (2013).

9.	Lin, L. & Chu, H. Quantifying publication bias in meta-analysis. *Biometrics* **74**, 785–794 (2018).

10.	Du Rietz, E. *et al.* Overlap between attention-deficit hyperactivity disorder and neurodevelopmental, externalising and internalising disorders: separating unique from general psychopathology effects. *Br. J. Psychiatry* **218**, 35–42 (2021).

11.	Taylor, M. J. *et al.* Association of genetic risk factors for psychiatric disorders and traits of these disorders in a Swedish population twin sample. *JAMA Psychiatry* **76**, 280–289 (2019).

12.	Bishop, D. V. M. & Hayiou-Thomas, M. E. Heritability of specific language impairment depends on diagnostic criteria. *Genes Brain Behav.* **7**, 365–372 (2008).

13.	Cheesman, R. *et al.* Childhood behaviour problems show the greatest gap between DNA-based and twin heritability. *Transl. Psychiatry* **7**, 1–9 (2017).

14.	DeThorne, L. S. *et al.* Children’s history of speech-language difficulties: Genetic influences and associations with reading-related measures. (2006).

15.	Hayiou-Thomas, M. E., Dale, P. S. & Plomin, R. The etiology of variation in language skills changes with development: a longitudinal twin study of language from 2 to 12 years. *Dev. Sci.* **15**, 233–249 (2012).

16.	Hayiou-Thomas, M. E., Dale, P. S. & Plomin, R. Language impairment from 4 to 12 years: Prediction and etiology. *J. Speech Lang. Hear. Res.* **57**, 850–864 (2014).

17.	Hohnen, B. & Stevenson, J. The structure of genetic influences on general cognitive, language, phonological, and reading abilities. *Dev. Psychol.* **35**, 590 (1999).

18.	Tomblin, J. B. & Buckwalter, P. R. Heritability of poor language achievement among twins. *J. Speech Lang. Hear. Res.* **41**, 188–199 (1998).

19.	Trzaskowski, M., Dale, P. S. & Plomin, R. No genetic influence for childhood behavior problems from DNA analysis. *J. Am. Acad. Child Adolesc. Psychiatry* **52**, 1048-1056. e3 (2013).

20.	van Beijsterveldt, C. E. M., Felsenfeld, S. & Boomsma, D. I. Bivariate genetic analyses of stuttering and nonfluency in a large sample of 5-year-old twins. (2010).

21.	Bishop, D. V. Motor immaturity and specific speech and language impairment: Evidence for a common genetic basis. *Am. J. Med. Genet.* **114**, 56–63 (2002).

22.	Bishop, D. V. M. DeFries–Fulker analysis of twin data with skewed distributions: Cautions and recommendations from a study of children’s use of verb inflections. *Behav. Genet.* **35**, 479–490 (2005).

23.	Bishop, D. V., Adams, C. V. & Norbury, C. F. Distinct genetic influences on grammar and phonological short‐term memory deficits: evidence from 6‐year‐old twins. *Genes Brain Behav.* **5**, 158–169 (2006).

24.	Bishop, D. V., Laws, G., Adams, C. & Norbury, C. F. High heritability of speech and language impairments in 6-year-old twins demonstrated using parent and teacher report. *Behav. Genet.* **36**, 173–184 (2006).

25.	Bishop, D. V., North, T. & Donlan, C. Nonword repetition as a behavioural marker for inherited language impairment: Evidence from a twin study. *J. Child Psychol. Psychiatry* **37**, 391–403 (1996).

26.	Dale, P. S., Rice, M. L., Rimfeld, K. & Hayiou-Thomas, M. E. Grammar clinical marker yields substantial heritability for language impairments in 16-year-old twins. *J. Speech Lang. Hear. Res.* **61**, 66–78 (2018).

27.	Dionne, G. *et al.* Associations between sleep-wake consolidation and language development in early childhood: a longitudinal twin study. *Sleep* **34**, 987–995 (2011).

28.	Dworzynski, K., Remington, A., Rijsdijk, F., Howell, P. & Plomin, R. Genetic etiology in cases of recovered and persistent stuttering in an unselected, longitudinal sample of young twins. (2007).

29.	Hoekstra, R. A., Bartels, M., Van Leeuwen, M. & Boomsma, D. I. Genetic architecture of verbal abilities in children and adolescents. *Dev. Sci.* **12**, 1041–1053 (2009).

30.	Mimeau, C. *et al.* The genetic and environmental etiology of the association between vocabulary and syntax in first grade. *Lang. Learn. Dev.* **14**, 149–166 (2018).

31.	Price, T. S., Dale, P. S. & Plomin, R. A longitudinal genetic analysis of low verbal and nonverbal cognitive abilities in early childhood. *Twin Res. Hum. Genet.* **7**, 139–148 (2004).

32.	Tosto, M. G. *et al.* The genetic architecture of oral language, reading fluency, and reading comprehension: A twin study from 7 to 16 years. *Dev. Psychol.* **53**, 1115 (2017).

33.	Trzaskowski, M. *et al.* DNA evidence for strong genome-wide pleiotropy of cognitive and learning abilities. *Behav. Genet.* **43**, 267–273 (2013).

34.	Viding, E. *et al.* Genetic and environmental influence on language impairment in 4‐year‐old same‐sex and opposite‐sex twins. *J. Child Psychol. Psychiatry* **45**, 315–325 (2004).

35.	Bailey, A. *et al.* Autism as a strongly genetic disorder: evidence from a British twin study. *Psychol. Med.* **25**, 63–77 (1995).

36.	Deng, W. *et al.* The relationship among genetic heritability, environmental effects, and autism spectrum disorders: 37 pairs of ascertained twin study. *J. Child Neurol.* **30**, 1794–1799 (2015).

37.	Dworzynski, K. *et al.* Developmental path between language and autistic-like impairments: A twin study. *Infant Child Dev. Int. J. Res. Pract.* **17**, 121–136 (2008).

38.	Dworzynski, K., Happe, F., Bolton, P. & Ronald, A. Relationship Between Symptom Domains in Autism Spectrum Disorders: A Population Based Twin Study. *J. Autism Dev. Disord.* **39**, 1197–1210 (2009).

39.	Frazier, T. W. *et al.* A twin study of heritable and shared environmental contributions to autism. *J. Autism Dev. Disord.* **44**, 2013–2025 (2014).

40.	Hallett, V., Ronald, A. & Happé, F. Investigating the association between autistic-like and internalizing traits in a community-based twin sample. *J. Am. Acad. Child Adolesc. Psychiatry* **48**, 618–627 (2009).

41.	Hoekstra, R., Bartels, M., Verweij, C. & Boomsma, D. Heritability of Autistic Traits in the General Population. *Arch. Pediatr. Adolesc. Med.* **161**, 372–377 (2007).

42.	Jones, A. P. *et al.* Phenotypic and aetiological associations between psychopathic tendencies, autistic traits, and emotion attribution. *Crim. Justice Behav.* **36**, 1198–1212 (2009).

43.	Lundstrom, S. *et al.* Autism Spectrum Disorders and Autisticlike Traits Similar Etiology in the Extreme End and the Normal Variation. *Arch. Gen. Psychiatry* **69**, 46–52 (2012).

44.	Pinto, R., Rijsdijk, F., Ronald, A., Asherson, P. & Kuntsi, J. The genetic overlap of attention-deficit/hyperactivity disorder and autistic-like traits: an investigation of individual symptom scales and cognitive markers. *J. Abnorm. Child Psychol.* **44**, 335–345 (2016).

45.	Polderman, T. J., Posthuma, D., De Sonneville, L. M., Verhulst, F. C. & Boomsma, D. I. Genetic analyses of teacher ratings of problem behavior in 5-year-old twins. *Twin Res. Hum. Genet.* **9**, 122–130 (2006).

46.	Robinson, E. *et al.* Evidence That Autistic Traits Show the Same Etiology in the General Population and at the Quantitative Extremes (5%, 2.5%, and 1%). *Arch. Gen. Psychiatry* **68**, 1113–1121 (2011).

47.	Robinson, E. B. *et al.* A multivariate twin study of autistic traits in 12-year-olds: testing the fractionable autism triad hypothesis. *Behav Genet* **42**, 245–255 (2012).

48.	Ronald, A. *et al.* Genetic Heterogeneity Between the Three Components of the Autism Spectrum: A Twin Study. *J. Am. Acad. Child Adolesc. Psychiatry* **45**, 691–699 (2006).

49.	Ronald, A., Happe, F., Price, T., Baron-Cohen, S. & Plomin, R. Phenotypic and Genetic Overlap Between Autistic Traits at the Extremes of the General Population. *J. Am. Acad. Child Adolesc. Psychiatry* **45**, 1206–1214 (2006).

50.	Ronald, A., Larsson, H., Anckarsäter, H. & Lichtenstein, P. Symptoms of autism and ADHD: a Swedish twin study examining their overlap. *J. Abnorm. Psychol.* **123**, 440 (2014).

51.	Ronald, A., Simonoff, E., Kuntsi, J., Asherson, P. & Plomin, R. Evidence for overlapping genetic influences on autistic and ADHD behaviours in a community twin sample. *J. Child Psychol. Psychiatry* **49**, 535–542 (2008).

52.	Scherff, A. *et al.* What causes internalising traits and autistic traits to co-occur in adolescence? A community-based twin study. *J. Abnorm. Child Psychol.* **42**, 601–610 (2014).

53.	Scourfield, J., Martin, N., Eley, T. C. & McGuffin, P. The Genetic Relationship Between Social Cognition and Conduct Problems. *Behav Genet* **34**, 377–383 (2004).

54.	Taylor, M. J. *et al.* Examining the association between autistic traits and atypical sensory reactivity: A twin study. *J. Am. Acad. Child Adolesc. Psychiatry* **57**, 96–102 (2018).

55.	Taylor, M. J. *et al.* Etiology of Autism Spectrum Disorders and Autistic Traits Over Time. *JAMA Psychiatry* **77**, 936–943 (2020).

56.	Taylor, M. J., Charman, T. & Ronald, A. Where are the strongest associations between autistic traits and traits of ADHD? Evidence from a community-based twin study. *Eur. Child Adolesc. Psychiatry* **24**, 1129–1138 (2015).

57.	Tick, B. *et al.* Autism Spectrum Disorders and other mental health problems: Exploring etiological overlaps and phenotypic causal associations. *J. Am. Acad. Child Adolesc. Psychiatry* **55**, 106-113. e4 (2016).

58.	Towers, H. *et al.* Genetic and environmental influences on teacher ratings of the Child Behavior Checklist. *Int. J. Behav. Dev.* **24**, 373–381 (2000).

59.	Yip, B. H. K. *et al.* Heritable variation, with little or no maternal effect, accounts for recurrence risk to autism spectrum disorder in Sweden. *Biol. Psychiatry* **83**, 589–597 (2018).

60.	Hallmayer, J. *et al.* Genetic heritability and shared environmental factors among twin pairs with autism. *Arch. Gen. Psychiatry* **68**, 1095–1102 (2011).

61.	Lundstrom, S. *et al.* Autistic-like traits and their association with mental health problems in two nationwide twin cohorts of children and adults. *Psychol Med* **41**, 2423–2433 (2011).

62.	Taniai, H., Nishiyama, T., Miyachi, T., Imaeda, M. & Sumi, S. Genetic influences on the broad spectrum of autism: Study of proband-ascertained twins. *Am. J. Med. Genet. B Neuropsychiatr. Genet.* **147B**, 844–849 (2008).

63.	Lundstrom, S. *et al.* Trajectories leading to autism spectrum disorders are affected by paternal age: findings from two nationally representative twin studies. *J. Child Psychol. Psychiatry* **51**, 850–856 (2010).

64.	Colvert, E. *et al.* Heritability of autism spectrum disorder in a UK population-based twin sample. *JAMA Psychiatry* **72**, 415–423 (2015).

65.	Ronald, A., Happé, F. & Plomin, R. The genetic relationship between individual differences in social and nonsocial behaviours characteristic of autism. *Dev. Sci.* **8**, 444–458 (2005).

66.	Boomsma, D. I., Van Beijsterveldt, T. C., Odintsova, V. V., Neale, M. C. & Dolan, C. V. Genetically Informed Regression Analysis: Application to Aggression Prediction by Inattention and Hyperactivity in Children and Adults. *Behav. Genet.* 1–14 (2020).

67.	Brikell, I. *et al.* Relative immaturity in childhood and attention-deficit/hyperactivity disorder symptoms from childhood to early adulthood: exploring genetic and environmental overlap across development. *J. Am. Acad. Child Adolesc. Psychiatry* **55**, 886–895 (2016).

68.	Brooker, R. J. *et al.* Attentional Control Explains Covariation Between Symptoms of Attention-Deficit/Hyperactivity Disorder and Anxiety During Adolescence. *J. Res. Adolesc.* **30**, 126–141 (2020).

69.	Burt, S. A., Krueger, R. F., McGue, M. & Iacono, W. G. Sources of covariation among attention-deficit/hyperactivity disorder, oppositional defiant disorder, and conduct disorder: the importance of shared environment. *J. Abnorm. Psychol.* **110**, 516 (2001).

70.	Burt, S. A., Larsson, H., Lichtenstein, P. & Klump, K. L. Additional evidence against shared environmental contributions to attention-deficit/hyperactivity problems. *Behav. Genet.* **42**, 711–721 (2012).

71.	Chang, Z., Lichtenstein, P. & Larsson, H. The effects of childhood ADHD symptoms on early-onset substance use: A Swedish twin study. *J. Abnorm. Child Psychol.* **40**, 425–435 (2012).

72.	Chang, Z., Lichtenstein, P., Asherson, P. J. & Larsson, H. Developmental twin study of attention problems: high heritabilities throughout development. *JAMA Psychiatry* **70**, 311–318 (2013).

73.	Chen, T.-J. *et al.* Genetic and environmental influences on the relationship between ADHD symptoms and internalizing problems: A Chinese twin study. *Am. J. Med. Genet. Part B Neuropsychiatr. Genet. Off. Publ. Int. Soc. Psychiatr. Genet.* **171**, 931–7 (2016).

74.	Cheung, C. H., Fazier-Wood, A. C., Asherson, P., Rijsdijk, F. & Kuntsi, J. Shared cognitive impairments and aetiology in ADHD symptoms and reading difficulties. *PloS One* **9**, e98590 (2014).

75.	Coolidge, F. L., Thede, L. L. & Young, S. E. Heritability and the comorbidity of attention deficit hyperactivity disorder with behavioral disorders and executive function deficits: A preliminary investigation. *Dev. Neuropsychol.* **17**, 273–287 (2000).

76.	Curran, S. *et al.* CHIP: Defining a dimension of the vulnerability to attention deficit hyperactivity disorder (ADHD) using sibling and individual data of children in a community‐based sample. *Am. J. Med. Genet. B Neuropsychiatr. Genet.* **119**, 86–97 (2003).

77.	de Zeeuw, E. L., van Beijsterveldt, C. E., Lubke, G. H., Glasner, T. J. & Boomsma, D. I. Childhood ODD and ADHD behavior: The effect of classroom sharing, gender, teacher gender and their interactions. *Behav. Genet.* **45**, 394–408 (2015).

78.	Derks, E. *et al.* Genetic and Environmental Influences on the Relation Between Attention Problems and Attention Deficit Hyperactivity Disorder. *Behav Genet* **38**, 11–23 (2008).

79.	Derks, E. M., Dolan, C. V., Hudziak, J. J., Neale, M. C. & Boomsma, D. I. Assessment and etiology of attention deficit hyperactivity disorder and oppositional defiant disorder in boys and girls. *Behav. Genet.* **37**, 559–566 (2007).

80.	Derks, E. M., Hudziak, J. J., Van Beijsterveldt, C. E. M., Dolan, C. V. & Boomsma, D. I. Genetic analyses of maternal and teacher ratings on attention problems in 7-year-old Dutch twins. *Behav. Genet.* **36**, 833–844 (2006).

81.	Dick, D., Viken, R., Kaprio, J., Pulkkinen, L. & Rose, R. Understanding the Covariation Among Childhood Externalizing Symptoms: Genetic and Environmental Influences on Conduct Disorder, Attention Deficit Hyperactivity Disorder, and Oppositional Defiant Disorder Symptoms. *J Abnorm Child Psychol* **33**, 219–229 (2005).

82.	Dolan, C. V., De Zeeuw, E. L., Zayats, T., Van Beijsterveldt, C. E. M. & Boomsma, D. I. The (broad-sense) genetic correlations among four measures of inattention and hyperactivity in 12 year olds. *Behav. Genet.* **50**, 273–288 (2020).

83.	Ebejer, J. L. *et al.* Genetic and environmental influences on inattention, hyperactivity-impulsivity, and reading: Kindergarten to grade 2. *Sci. Stud. Read.* **14**, 293–316 (2010).

84.	Ebejer, J. L. *et al.* Contrast effects and sex influence maternal and self-report dimensional measures of Attention-Deficit Hyperactivity Disorder. *Behav. Genet.* **45**, 35–50 (2015).

85.	Edelbrock, C., Rende, R., Plomin, R. & Thompson, L. A. A twin study of competence and problem behavior in childhood and early adolescence. *J. Child Psychol. Psychiatry* **36**, 775–785 (1995).

86.	Gould, K. L., Coventry, W. L., Olson, R. K. & Byrne, B. Gene-environment interactions in ADHD: the roles of SES and chaos. *J. Abnorm. Child Psychol.* **46**, 251–263 (2018).

87.	Greven, C. U., Asherson, P., Rijsdijk, F. V. & Plomin, R. A longitudinal twin study on the association between inattentive and hyperactive-impulsive ADHD symptoms. *J. Abnorm. Child Psychol.* **39**, 623–632 (2011).

88.	Greven, C. U., Harlaar, N., Dale, P. S. & Plomin, R. Genetic overlap between ADHD symptoms and reading is largely driven by inattentiveness rather than hyperactivity-impulsivity. *J. Can. Acad. Child Adolesc. Psychiatry* **20**, 6 (2011).

89.	Greven, C. U., Kovas, Y., Willcutt, E. G., Petrill, S. A. & Plomin, R. Evidence for shared genetic risk between ADHD symptoms and reduced mathematics ability: a twin study. *J. Child Psychol. Psychiatry* **55**, 39–48 (2014).

90.	Greven, C., Rijsdijk, F., Asherson, P. & Plomin, R. A longitudinal twin study on the association between ADHD symptoms and reading. *J Child Psychol Psychiatry Allied Discipl* **53**, 234–242 (2012).

91.	Greven, C., Rijsdijk, F. & Plomin, R. A Twin Study of ADHD Symptoms in Early Adolescence: Hyperactivity-impulsivity and Inattentiveness Show Substantial Genetic Overlap but Also Genetic Specificity. *J Abnorm Child Psychol* **39**, 265–275 (2011).

92.	Hay, D. A., Bennett, K. S., Levy, F., Sergeant, J. & Swanson, J. A twin study of attention-deficit/hyperactivity disorder dimensions rated by the strengths and weaknesses of ADHD-symptoms and normal-behavior (SWAN) scale. *Biol. Psychiatry* **61**, 700–705 (2007).

93.	Heutink, P., Verhuls, F. C. & Boomsma, D. I. A longitudinal twin study on IQ, executive functioning, and attention problems during childhood and early adolescence. *Acta Neurol Belg* **106**, 191–207 (2006).

94.	Hudziak, J. J., Derks, E. M., Althoff, R. R., Rettew, D. C. & Boomsma, D. I. The Genetic and Environmental Contributions to Attention Deficit Hyperactivity Disorder as Measured by the Conners’ Rating Scales—Revised. *Am. J. Psychiatry* **162**, 1614–1620 (2005).

95.	Hur, Y. M. Increasing Phenotypic and Genetic variations in Hyperactivity/Inattention Problems from Age 3 to 13 Years: A Cross-Sectional Twin Study. *Twin Res. Hum. Genet.* **17**, 545–552 (2014).

96.	Jaffee, S. R., Hanscombe, K. B., Haworth, C. M., Davis, O. S. & Plomin, R. Chaotic homes and children’s disruptive behavior: A longitudinal cross-lagged twin study. *Psychol. Sci.* **23**, 643–650 (2012).

97.	Johnson, W., McGue, M. & Iacono, W. Disruptive Behavior and School Grades: Genetic and Environmental Relations in 11-Year-Olds. *J Educ Psychol* **97**, 391–405 (2005).

98.	Kan, K.-J. *et al.* Genetic and environmental stability in attention problems across the lifespan: evidence from the Netherlands twin register. *J. Am. Acad. Child Adolesc. Psychiatry* **52**, 12–25 (2013).

99.	Kan, K.-J., van Beijsterveldt, C. E., Bartels, M. & Boomsma, D. I. Assessing genetic influences on behavior: informant and context dependency as illustrated by the analysis of attention problems. *Behav. Genet.* **44**, 326–336 (2014).

100.	Kuja-Halkola, R., Lichtenstein, P., D’Onofrio, B. & Larsson, H. Codevelopment of ADHD and externalizing behavior from childhood to adulthood. *J Child Psychol Psychiatry Allied Discipl* **56**, 640–647 (2015).

101.	Kuntsi, J. & Stevenson, J. Psychological Mechanisms in Hyperactivity: II The Role of Genetic Factors. *J. Child Psychol. Psychiatry* **42**, 211–219 (2001).

102.	Kuntsi, J. *et al.* The Separation of ADHD Inattention and Hyperactivity-Impulsivity Symptoms: Pathways from Genetic Effects to Cognitive Impairments and Symptoms. *J Abnorm Child Psychol* **42**, 127–136 (2014).

103.	Kuntsi, J., Gayán, J. & Stevenson, J. Parents’ and teachers’ ratings of problem behaviours in children: Genetic and contrast effects. *Twin Res. Hum. Genet.* **3**, 251–258 (2000).

104.	Kuntsi, J., Rijsdijk, F., Ronald, A., Asherson, P. & Plomin, R. Genetic influences on the stability of attention-deficit/hyperactivity disorder symptoms from early to middle childhood. *Biol. Psychiatry* **57**, 647–654 (2005).

105.	Larsson, H., Anckarsater, H., Råstam, M., Chang, Z. & Lichtenstein, P. Childhood attention‐deficit hyperactivity disorder as an extreme of a continuous trait: A quantitative genetic study of 8,500 twin pairs. *J. Child Psychol. Psychiatry* **53**, 73–80 (2012).

106.	Larsson, H., Dilshad, R., Lichtenstein, P. & Barker, E. Developmental trajectories of DSM-IV symptoms of attention-deficit/hyperactivity disorder: genetic effects, family risk and associated psychopathology. *J Child Psychol Psychiatry Allied Discipl* **52**, 954–963 (2011).

107.	Lemery-Chalfant, K., Doelger, L. & Goldsmith, H. H. Genetic relations between effortful and attentional control and symptoms of psychopathology in middle childhood. *Infant Child Dev.* **17**, 365–385 (2008).

108.	Levy, F., Hay, D. A., McStephen, M., Wood, C. & Waldman, I. Attention-Deficit Hyperactivity Disorder: A Category or a Continuum? Genetic Analysis of a Large-Scale Twin Study. *J. Am. Acad. Child Adolesc. Psychiatry* **36**, 737–744 (1997).

109.	Lewis, G. J. & Plomin, R. Heritable influences on behavioural problems from early childhood to mid-adolescence: evidence for genetic stability and innovation. *Psychol. Med.* **45**, 2171–2179 (2015).

110.	Lewis, G., Haworth, C. & Plomin, R. Identical genetic influences underpin behavior problems in adolescence and basic traits of personality. *J Child Psychol Psychiatry Allied Discipl* **55**, 865–875 (2014).

111.	Lifford, K. J., Harold, G. T. & Thapar, A. Parent–child hostility and child ADHD symptoms: A genetically sensitive and longitudinal analysis. *J. Child Psychol. Psychiatry* **50**, 1468–1476 (2009).

112.	Little, C. W., Hart, S. A., Schatschneider, C. & Taylor, J. Examining associations among ADHD, homework behavior, and reading comprehension: A twin study. *J. Learn. Disabil.* **49**, 410–423 (2016).

113.	LoParo, D. & Waldman, I. Twins’ rearing environment similarity and childhood externalizing disorders: A test of the equal environments assumption. *Behav. Genet.* **44**, 606–613 (2014).

114.	Martin, N. C., Piek, J. P. & Hay, D. DCD and ADHD: a genetic study of their shared aetiology. *Hum. Mov. Sci.* **25**, 110–124 (2006).

115.	McLoughlin, G., Ronald, A., Kuntsi, J., Asherson, P. & Plomin, R. Genetic Support for the Dual Nature of Attention Deficit Hyperactivity Disorder: Substantial Genetic Overlap Between the Inattentive and Hyperactive–impulsive Components. *J Abnorm Child Psychol* **35**, 999–1008 (2007).

116.	Merwood, A. *et al.* Different heritabilities but shared etiological influences for parent, teacher and self-ratings of ADHD symptoms: an adolescent twin study. *Psychol. Med.* **43**, (2013).

117.	Michelini, G., Eley, T. C., Gregory, A. M. & McAdams, T. A. Aetiological overlap between anxiety and attention deficit hyperactivity symptom dimensions in adolescence. *J. Child Psychol. Psychiatry* **56**, 423–431 (2015).

118.	Mikolajewski, A. J., Allan, N. P., Hart, S. A., Lonigan, C. J. & Taylor, J. Negative affect shares genetic and environmental influences with symptoms of childhood internalizing and externalizing disorders. *J. Abnorm. Child Psychol.* **41**, 411–423 (2013).

119.	Molenaar, D., Middeldorp, C., van Beijsterveldt, T. & Boomsma, D. I. Analysis of behavioral and emotional problems in children highlights the role of genotype× environment interaction. *Child Dev.* **86**, 1999–2016 (2015).

120.	Moruzzi, S., Rijsdijk, F. & Battaglia, M. A twin study of the relationships among inattention, hyperactivity/impulsivity and sluggish cognitive tempo problems. *J. Abnorm. Child Psychol.* **42**, 63–75 (2014).

121.	Nikolas, M. A., Klump, K. L. & Burt, S. A. Parental involvement moderates etiological influences on attention deficit hyperactivity disorder behaviors in child twins. *Child Dev.* **86**, 224–240 (2015).

122.	Niv, S., Tuvblad, C., Raine, A., Wang, P. & Baker, L. A. Heritability and longitudinal stability of impulsivity in adolescence. *Behav Genet* **42**, 378–392 (2012).

123.	Paloyelis, Y., Rijsdijk, F., Wood, A. C., Asherson, P. & Kuntsi, J. The genetic association between ADHD symptoms and reading difficulties: the role of inattentiveness and IQ. *J. Abnorm. Child Psychol.* **38**, 1083–1095 (2010).

124.	Peng, C.-Z. *et al.* Familial influences on the full range of variability in attention and activity levels during adolescence: A longitudinal twin study. *Dev. Psychopathol.* **28**, 517 (2016).

125.	Pingault, J. B., Rijsdijk, F., Zheng, Y., Plomin, R. & Viding, E. Developmentally dynamic genome: Evidence of genetic influences on increases and decreases in conduct problems from early childhood to adolescence. *Sci. Rep.* **5**, 9 (2015).

126.	Plourde, V. *et al.* Phenotypic and genetic associations between reading comprehension, decoding skills, and ADHD dimensions: evidence from two population-based studies. *J. Child Psychol. Psychiatry* **56**, 1074–1082 (2015).

127.	Plourde, V., Boivin, M., Brendgen, M., Vitaro, F. & Dionne, G. Phenotypic and genetic associations between reading and attention-deficit/hyperactivity disorder dimensions in adolescence. *Dev. Psychopathol.* **29**, 1215–1226 (2017).

128.	Polderman, T. J. *et al.* A genetic study on attention problems and academic skills: results of a longitudinal study in twins. *J. Can. Acad. Child Adolesc. PsychiatryJournal Académie Can. Psychiatr. Enfant Adolesc.* (2011).

129.	Polderman, T. J., van Dongen, J. & Boomsma, D. I. The relation between ADHD symptoms and fine motor control: a genetic study. *Child Neuropsychol.* **17**, 138–150 (2011).

130.	Price, T. *et al.* Continuity and Change in Preschool ADHD Symptoms: Longitudinal Genetic Analysis with Contrast Effects. *Behav Genet* **35**, 121–132 (2005).

131.	Quinn, P. D. *et al.* Childhood attention‐deficit/hyperactivity disorder symptoms and the development of adolescent alcohol problems: A prospective, population‐based study of Swedish twins. *Am. J. Med. Genet. B Neuropsychiatr. Genet.* **171**, 958–970 (2016).

132.	Rosenberg, J., Pennington, B. F., Willcutt, E. G. & Olson, R. K. Gene by environment interactions influencing reading disability and the inattentive symptom dimension of attention deficit/hyperactivity disorder. *J. Child Psychol. Psychiatry* **53**, 243–251 (2012).

133.	Rydell, M., Taylor, M. & Larsson, H. Genetic and environmental contributions to the association between ADHD and affective problems in early childhood-A Swedish population-based twin study. *Am J Med Genet Part B Neuropsychiatr Genet* **174**, 538–546 (2017).

134.	Saudino, K. J. & Plomin, R. Why are hyperactivity and academic achievement related? *Child Dev.* **78**, 972–986 (2007).

135.	Saunders, M. C. *et al.* The associations between callous-unemotional traits and symptoms of conduct problems, hyperactivity and emotional problems: A study of adolescent twins screened for neurodevelopmental problems. *J. Abnorm. Child Psychol.* **47**, 447–457 (2019).

136.	Siebelink, N. M. *et al.* Genetic and environmental aetiologies of associations between dispositional mindfulness and ADHD traits: a population-based twin study. *Eur. Child Adolesc. Psychiatry* **28**, 1241–1251 (2019).

137.	Simonoff, E. *et al.* Genetic influences on childhood hyperactivity: Contrast effects imply parental rating bias, not sibling interaction. *Psychol. Med.* **28**, 825–837 (1998).

138.	Stern, A. *et al.* Associations between ADHD and emotional problems from childhood to young adulthood: a longitudinal genetically sensitive study. *J Child Psychol Psychiatry Allied Discipl* **61**, 1234–1242 (2020).

139.	Stevenson, J. Evidence for a genetic etiology in hyperactivity in children. *Behav Genet* **22**, 337–344 (1992).

140.	Stevenson, J., Pennington, B. F., Gilger, J. W., DeFries, J. C. & Gillis, J. J. Hyperactivity and Spelling Disability: Testing for Shared Genetic Aetiology. *J. Child Psychol. Psychiatry* **34**, 1137–1152 (1993).

141.	Taylor, J., Allan, N., Mikolajewski, A. & Hart, S. Common genetic and nonshared environmental factors contribute to the association between socioemotional dispositions and the externalizing factor in children. *J Child Psychol Psychiatry Allied Discipl* **54**, 67–76 (2013).

142.	Thapar, A., Hervas, A. & McGuffin, P. Childhood hyperactivity scores are highly heritable and show sibling competition effects: Twin study evidence. *Behav Genet* **25**, 537–544 (1995).

143.	Tuvblad, C., Zheng, M., Raine, A. & Baker, L. A Common Genetic Factor Explains the Covariation Among ADHD ODD and CD Symptoms in 9-10 Year Old Boys and Girls. *J Abnorm Child Psychol* **37**, 153–167 (2009).

144.	Tye, C. *et al.* Shared genetic influences on ADHD symptoms and very low-frequency EEG activity: a twin study. *J Child Psychol Psychiatry Allied Discipl* **53**, 706–715 (2012).

145.	Vendlinski, M. K. *et al.* Relative influence of genetics and shared environment on child mental health symptoms depends on comorbidity. *PloS One* **9**, e103080 (2014).

146.	Waszczuk, M. A., Zavos, H. M. & Eley, T. C. Why do depression, conduct, and hyperactivity symptoms co-occur across adolescence? The role of stable and dynamic genetic and environmental influences. *Eur. Child Adolesc. Psychiatry* 1–13 (2020).

147.	Willcutt, E. G. *et al.* Preschool twin study of the relation between attention-deficit/hyperactivity disorder and prereading skills. *Read. Writ.* **20**, 103–125 (2007).

148.	Willcutt, E. G. *et al.* Etiology and neuropsychology of comorbidity between RD and ADHD: The case for multiple-deficit models. *Cortex* **46**, 1345–1361 (2010).

149.	Wood, A. C., Rijsdijk, F., Asherson, P. & Kuntsi, J. Hyperactive-impulsive symptom scores and oppositional behaviours reflect alternate manifestations of a single liability. *Behav. Genet.* **39**, 447–460 (2009).

150.	Wood, A. C., Rijsdijk, F., Asherson, P. & Kuntsi, J. Inferring causation from cross-sectional data: examination of the causal relationship between hyperactivity–impulsivity and novelty seeking. *Front. Genet.* **2**, 6 (2011).

151.	Wood, A. C., Kuntsi, J., Asherson, P. & Saudino, K. J. Actigraph data are, reliable, with functional reliability increasing with aggregation. *Behav. Res. Methods* **40**, 873–878 (2008).

152.	Zheng, Y., Pingault, J.-B., Unger, J. B. & Rijsdijk, F. Genetic and environmental influences on attention-deficit/hyperactivity disorder symptoms in Chinese adolescents: a longitudinal twin study. *Eur. Child Adolesc. Psychiatry* **29**, 205–216 (2020).

153.	Zumberge, A., Baker, L. A. & Manis, F. R. Focus on words: a twin study of reading and inattention. *Behav. Genet.* **37**, 284–293 (2007).

154.	Burt, S. A., McGUE, M., Krueger, R. F. & Iacono, W. G. Sources of covariation among the child-externalizing disorders: informant effects and the shared environment. *Psychol. Med.* **35**, 1133 (2005).

155.	Chen, Q. *et al.* Shared familial risk factors between attention-deficit/hyperactivity disorder and overweight/obesity - a population-based familial coaggregation study in Sweden. *J Child Psychol Psychiatry Allied Discipl* **58**, 711–718 (2017).

156.	Crosbie, J. *et al.* Response inhibition and ADHD traits: correlates and heritability in a community sample. *J. Abnorm. Child Psychol.* **41**, 497–507 (2013).

157.	Eilertsen, E. M. *et al.* Development of ADHD symptoms in preschool children: Genetic and environmental contributions. *Dev. Psychopathol.* (2018).

158.	Fedko, I. O. *et al.* Heritability of behavioral problems in 7-year olds based on shared and unique aspects of parental views. *Behav. Genet.* **47**, 152–163 (2017).

159.	Haberstick, B. C. *et al.* Genetic and environmental contributions to retrospectively reported DSM-IV childhood attention deficit hyperactivity disorder. *Psychol. Med.* **38**, 1057–1066 (2008).

160.	Martin, N. C., Levy, F., Pieka, J. & Hay, D. A. A genetic study of attention deficit hyperactivity disorder, conduct disorder, oppositional defiant disorder and reading disability: Aetiological overlaps and implications. *Int. J. Disabil. Dev. Educ.* **53**, 21–34 (2006).

161.	Merwood, A., Asherson, P. & Larsson, H. Genetic associations between the ADHD symptom dimensions and Cloninger’s temperament dimensions in adult twins. *Eur. Neuropsychopharmacol.* **23**, 416–425 (2013).

162.	Mogensen, N., Larsson, H., Lundholm, C. & Almqvist, C. Association between childhood asthma and ADHD symptoms in adolescence–a prospective population‐based twin study. *Allergy* **66**, 1224–1230 (2011).

163.	Nadder, T., Silberg, J., Eaves, L., Maes, H. & Meyer, J. Genetic Effects on ADHD Symptomatology in 7- to 13-Year-Old Twins: Results from a Telephone Survey. *Behav Genet* **28**, 83–99 (1998).

164.	Rhee, S. H., Waldman, I. D., Hay, D. A. & Levy, F. Sex differences in genetic and environmental influences on DSM–III–R attention-deficit/hyperactivity disorder. *J. Abnorm. Psychol.* **108**, 24 (1999).

165.	Rimfeld, K. *et al.* Genetic correlates of psychological responses to the COVID-19 crisis in young adult twins in Great Britain. *Behav. Genet.* **51**, 110–124 (2021).

166.	Singh, A. & Waldman, I. The Etiology of Associations Between Negative Emotionality and Childhood Externalizing Disorders. *J. Abnorm. Psychol.* **119**, 376–388 (2010).

167.	Willcutt, E., Pennington, B. & DeFries, J. Etiology of Inattention and Hyperactivity/Impulsivity in a Community Sample of Twins with Learning Difficulties. *J Abnorm Child Psychol* **28**, 149–159 (2000).

168.	Willcutt, E. G., Pennington, B. F., Olson, R. K. & DeFries, J. C. Understanding comorbidity: A twin study of reading disability and attention‐deficit/hyperactivity disorder. *Am. J. Med. Genet. B Neuropsychiatr. Genet.* **144**, 709–714 (2007).

169.	Merwood, A. *et al.* Genetic associations between the symptoms of attention-deficit/hyperactivity disorder and emotional lability in child and adolescent twins. *J. Am. Acad. Child Adolesc. Psychiatry* **53**, 209-220. e4 (2014).

170.	Thapar, A., Harrington, R., Ross, K. & McGuffin, P. Does the Definition of ADHD Affect Heritability? *J. Am. Acad. Child Adolesc. Psychiatry* **39**, 1528–1536 (2000).

171.	Ehringer, M. A., Rhee, S. H., Young, S., Corley, R. & Hewitt, J. K. Genetic and environmental contributions to common psychopathologies of childhood and adolescence: a study of twins and their siblings. *J. Abnorm. Child Psychol.* **34**, 1–17 (2006).

172.	Smith, A. K. *et al.* The role of attention-deficit/hyperactivity disorder in the association between verbal ability and conduct disorder. *Front. Psychiatry* **2**, 3 (2011).

173.	Thapar, A., Harrington, R. & McGuffin, P. Examining the comorbidity of ADHD-related behaviours and conduct problems using a twin study design. *Br J Psychiatry* **179**, 224–229 (2001).

174.	Martin, N., Scourfield, J. & McGuffin, P. Observer effects and heritability of childhood attention-deficit hyperactivity disorder symptoms. *Br. J. Psychiatry* **180**, 260–265 (2002).

175.	Alarcón, M., DeFries, J. C., Light, J. G. & Pennington, B. F. A twin study of mathematics disability. *J. Learn. Disabil.* **30**, 617–623 (1997).

176.	Bishop, D. V. M. Genetic influences on language impairment and literacy problems in children: Same or different? *J. Child Psychol. Psychiatry* **42**, 189–198 (2001).

177.	Davis, C. J. *et al.* Etiology of reading difficulties and rapid naming: the Colorado Twin Study of Reading Disability. *Behav. Genet.* **31**, 625–635 (2001).

178.	Davis, O. S. P. *et al.* Generalist genes and the Internet generation: etiology of learning abilities by web testing at age 10. *Genes Brain Behav.* **7**, 455–462 (2008).

179.	Davis, O. S. *et al.* The correlation between reading and mathematics ability at age twelve has a substantial genetic component. *Nat. Commun.* **5**, 1–6 (2014).

180.	DeFries, J. C. & Alarcón, M. Genetics of specific reading disability. *Ment. Retard. Dev. Disabil. Res. Rev.* **2**, 39–47 (1996).

181.	DeFries, J. C., Knopik, V. S. & Wadsworth, S. J. Colorado twin study of reading disability. *Read. Atten. Disord. Neurobiol. Correl.* 17–41 (1999).

182.	Erbeli, F., Hart, S. A., Wagner, R. K. & Taylor, J. Examining the etiology of reading disability as conceptualized by the hybrid model. *Sci. Stud. Read.* **22**, 167–180 (2018).

183.	Erbeli, F., Hart, S. A. & Taylor, J. Genetic and environmental influences on achievement outcomes based on family history of learning disabilities status. *J. Learn. Disabil.* **52**, 135–145 (2019).

184.	Gayan, J. & Olson, R. K. Genetic and environmental influences on orthographic and phonological skills in children with reading disabilities. *Dev. Neuropsychol.* **20**, 483–507 (2001).

185.	Harlaar, N., Kovas, Y., Dale, P. S., Petrill, S. A. & Plomin, R. Mathematics is differentially related to reading comprehension and word decoding: Evidence from a genetically sensitive design. *J. Educ. Psychol.* **104**, 622 (2012).

186.	Harlaar, N., Trzaskowski, M., Dale, P. S. & Plomin, R. Word reading fluency: Role of genome‐wide single‐nucleotide polymorphisms in developmental stability and correlations with print exposure. *Child Dev.* **85**, 1190–1205 (2014).

187.	Hart, S. A., Petrill, S. A., Thompson, L. A. & Plomin, R. The ABCs of math: A genetic analysis of mathematics and its links with reading ability and general cognitive ability. *J. Educ. Psychol.* **101**, 388 (2009).

188.	Hensler, B., Schatschneider, C., Taylor, J. & Wagner, R. Behavioral Genetic Approach to the Study of Dyslexia. *J Dev Behav Pediatr* **31**, 525–532 (2010).

189.	Kovas, Y. *et al.* The genetic and environmental origins of learning abilities and disabilities in the early school years. *Monogr. Soc. Res. Child Dev.* i–156 (2007).

190.	Marlow, A. *et al.* Investigation of Quantitative Measures Related to Reading Disability in a Large Sample of Sib-Pairs from the UK. *Behav Genet* **31**, 219–230 (2001).

191.	Newsome, J., Boisvert, D. & Wright, J. P. Genetic and environmental influences on the co-occurrence of early academic achievement and externalizing behavior. *J. Crim. Justice* **42**, 45–53 (2014).

192.	Olson, R. K., Gillis, J. J., Rack, J. P., DeFries, J. C. & Fulker, D. W. Confirmatory factor analysis of word recognition and process measures in the Colorado Reading Project. *Read. Writ.* **3**, 235–248 (1991).

193.	Petrill, S. A. *et al.* Longitudinal genetic analysis of early reading: the Western Reserve reading project. *Read. Writ.* **20**, 127–146 (2007).

194.	Samuelsson, S. *et al.* Genetic and environmental influences on prereading skills and early reading and spelling development in the United States, Australia, and Scandinavia. *Read. Writ.* **20**, 51–75 (2007).

195.	Tosto, M. G. *et al.* Why do we differ in number sense? Evidence from a genetically sensitive investigation. *Intelligence* **43**, 35–46 (2014).

196.	Wadsworth, S. J., DeFries, J. C., Willcutt, E. G., Pennington, B. F. & Olson, R. K. The Colorado longitudinal twin study of reading difficulties and ADHD: Etiologies of comorbidity and stability. *Twin Res. Hum. Genet.* **18**, 755–761 (2015).

197.	Wadsworth, S. J., DeFries, J. C., Willcutt, E. G., Pennington, B. F. & Olson, R. K. Genetic etiologies of comorbidity and stability for reading difficulties and ADHD: A replication study. *Twin Res. Hum. Genet.* **19**, 647–651 (2016).

198.	Wadsworth, S. J., Olson, R. K. & DeFries, J. C. Differential genetic etiology of reading difficulties as a function of IQ: an update. *Behav. Genet.* **40**, 751–758 (2010).

199.	Wadsworth, S. J., Olson, R. K., Pennington, B. F. & DeFries, J. C. Differential genetic etiology of reading disability as a function of IQ. *J. Learn. Disabil.* **33**, 192–199 (2000).

200.	Willcutt, E. G. *et al.* Understanding comorbidity between specific learning disabilities. *New Dir. Child Adolesc. Dev.* **2019**, 91–109 (2019).

201.	Willcutt, E. G., Pennington, B. F. & DeFries, J. C. Twin study of the etiology of comorbidity between reading disability and attention‐deficit/hyperactivity disorder. *Am. J. Med. Genet.* **96**, 293–301 (2000).

202.	Astrom, R. L., Wadsworth, S. J., Olson, R. K., Willcutt, E. G. & DeFries, J. C. DeFries–Fulker analysis of longitudinal reading performance data from twin pairs ascertained for reading difficulties and from their nontwin siblings. *Behav. Genet.* **41**, 660–667 (2011).

203.	Betjemann, R. S. *et al.* Genetic covariation between brain volumes and IQ, reading performance, and processing speed. *Behav. Genet.* **40**, 135–145 (2010).

204.	Bishop, D. V., Adams, C. V. & Norbury, C. F. Using nonword repetition to distinguish genetic and environmental influences on early literacy development: A study of 6‐year‐old twins. *Am. J. Med. Genet. B Neuropsychiatr. Genet.* **129**, 94–96 (2004).

205.	Castles, A., Datta, H., Gayan, J. & Olson, R. K. Varieties of developmental reading disorder: Genetic and environmental influences. *J. Exp. Child Psychol.* **72**, 73–94 (1999).

206.	Christopher, M. E. *et al.* The genetic and environmental etiologies of individual differences in early reading growth in Australia, the United States, and Scandinavia. *J. Exp. Child Psychol.* **115**, 453–467 (2013).

207.	Daucourt, M. C., Haughbrook, R., Van Bergen, E. & Hart, S. A. The association of parent-reported executive functioning, reading, and math is explained by nature, not nurture. *Dev. Psychol.* (2020).

208.	DeFries, J. C., Fulker, D. W. & LaBuda, M. C. Evidence for a genetic aetiology in reading disability of twins. *Nature* **329**, 537–539 (1987).

209.	Erbeli, F., Hart, S. A. & Taylor, J. Longitudinal associations among reading‐related skills and reading comprehension: A twin study. *Child Dev.* **89**, e480–e493 (2018).

210.	Friend, A. *et al.* Heritability of high reading ability and its interaction with parental education. *Behav. Genet.* **39**, 427–436 (2009).

211.	Friend, A., DeFries, J. C., Wadsworth, S. J. & Olson, R. K. Genetic and environmental influences on word recognition and spelling deficits as a function of age. *Behav. Genet.* **37**, 477–486 (2007).

212.	Garon-Carrier, G. *et al.* Persistent genetic and family-wide environmental contributions to early number knowledge and later achievement in mathematics. *Psychol. Sci.* **28**, 1707–1718 (2017).

213.	Gayán, J. & Olson, R. K. Genetic and environmental influences on individual differences in printed word recognition. *J. Exp. Child Psychol.* **84**, 97–123 (2003).

214.	Gillis, J. J., DeFries, J. C. & Fulker, D. W. Confirmatory factor analysis of reading and mathematics performance: A twin study. *Acta Genet. Medicae Gemellol. Twin Res.* **41**, 287–300 (1992).

215.	Grasby, K. L. & Coventry, W. L. Longitudinal stability and growth in literacy and numeracy in Australian school students. *Behav. Genet.* **46**, 649–664 (2016).

216.	Harlaar, N., Dale, P. S. & Plomin, R. Reading exposure: A (largely) environmental risk factor with environmentally‐mediated effects on reading performance in the primary school years. *J. Child Psychol. Psychiatry* **48**, 1192–1199 (2007).

217.	Hart, S. A. *et al.* Exploring how nature and nurture affect the development of reading: an analysis of the Florida Twin Project on reading. *Dev. Psychol.* **49**, 1971 (2013).

218.	Hawke, J. L., Stallings, M. C., Wadsworth, S. J. & DeFries, J. C. DeFries–Fulker and Pearson–Aitken model-fitting analyses of reading performance data from selected and unselected twin pairs. *Behav. Genet.* **38**, 101–107 (2008).

219.	Knopik, V. S. *et al.* Differential genetic etiology of reading component processes as a function of IQ. *Behav. Genet.* **32**, 181–198 (2002).

220.	Knopik, V. S., Alarcón, M. & DeFries, J. C. Comorbidity of mathematics and reading deficits: Evidence for a genetic etiology. *Behav. Genet.* **27**, 447–453 (1997).

221.	Kovas, Y. *et al.* Literacy and numeracy are more heritable than intelligence in primary school. *Psychol. Sci.* **24**, 2048–2056 (2013).

222.	Kovas, Y. *et al.* Overlap and specificity of genetic and environmental influences on mathematics and reading disability in 10‐year‐old twins. *J. Child Psychol. Psychiatry* **48**, 914–922 (2007).

223.	Lazaroo, N. K. *et al.* Genetic structure of IQ, phonemic decoding skill, and academic achievement. *Front. Genet.* **10**, 195 (2019).

224.	Logan, J. A. *et al.* Reading development in young children: Genetic and environmental influences. *Child Dev.* **84**, 2131–2144 (2013).

225.	Malanchini, M. *et al.* Reading self-perceived ability, enjoyment and achievement: A genetically informative study of their reciprocal links over time. *Dev. Psychol.* **53**, 698 (2017).

226.	Malanchini, M. *et al.* Genetic factors underlie the association between anxiety, attitudes and performance in mathematics. *Transl. Psychiatry* **10**, 1–11 (2020).

227.	Malanchini, M., Engelhardt, L. E., Grotzinger, A. D., Harden, K. P. & Tucker-Drob, E. M. “Same but different”: Associations between multiple aspects of self-regulation, cognition, and academic abilities. *J. Pers. Soc. Psychol.* **117**, 1164 (2019).

228.	Oliver, B. R., Dale, P. S. & Plomin, R. Writing and reading skills as assessed by teachers in 7-year olds: A behavioral genetic approach. *Cogn. Dev.* **22**, 77–95 (2007).

229.	Petrill, S. A. *et al.* Genetic and environmental influences on the growth of early reading skills. *J. Child Psychol. Psychiatry* **51**, 660–667 (2010).

230.	Rimfeld, K. *et al.* The stability of educational achievement across school years is largely explained by genetic factors. *NPJ Sci. Learn.* **3**, 1–10 (2018).

231.	Rimfeld, K. *et al.* Teacher assessments during compulsory education are as reliable, stable and heritable as standardized test scores. *J. Child Psychol. Psychiatry* **60**, 1278–1288 (2019).

232.	Rimfeld, K., Ayorech, Z., Dale, P. S., Kovas, Y. & Plomin, R. Genetics affects choice of academic subjects as well as achievement. *Sci. Rep.* **6**, 1–9 (2016).

233.	Rimfeld, K., Kovas, Y., Dale, P. S. & Plomin, R. Pleiotropy across academic subjects at the end of compulsory education. *Sci. Rep.* **5**, 1–12 (2015).

234.	Shakeshaft, N. G. *et al.* Strong genetic influence on a UK nationwide test of educational achievement at the end of compulsory education at age 16. *PloS One* **8**, e80341 (2013).

235.	Swagerman, S. *et al.* Genetic transmission of reading ability. *Brain Lang* **172**, 3–8 (2017).

236.	Taylor, J. & Schatschneider, C. Genetic influence on literacy constructs in kindergarten and first grade: Evidence from a diverse twin sample. *Behav. Genet.* **40**, 591–602 (2010).

237.	Taylor, J., Erbeli, F., Hart, S. A. & Johnson, W. Early classroom reading gains moderate shared environmental influences on reading comprehension in adolescence. *J. Child Psychol. Psychiatry* **61**, 689–698 (2020).

238.	Tosto, M. G. *et al.* The nature of the association between number line and mathematical performance: An international twin study. *Br. J. Educ. Psychol.* **89**, 787–803 (2019).

239.	Tosto, M. G., Malykh, S., Voronin, I., Plomin, R. & Kovas, Y. The etiology of individual differences in maths beyond IQ: insights from 12-year old twins. *Procedia-Soc. Behav. Sci.* **86**, 429–434 (2013).

240.	Wadsworth, S. J., Olson, R. K., Willcutt, E. G. & DeFries, J. C. Multiple regression analysis of reading performance data from twin pairs with reading difficulties and nontwin siblings: The augmented model. *Twin Res. Hum. Genet.* **15**, 116–119 (2012).

241.	Wong, S. W., Chow, B. W.-Y., Ho, C. S.-H., Waye, M. M. & Bishop, D. V. Genetic and environmental overlap between Chinese and English reading-related skills in Chinese children. *Dev. Psychol.* **50**, 2539 (2014).

242.	Keenan, J. M., Betjemann, R. S., Wadsworth, S. J., DeFries, J. C. & Olson, R. K. Genetic and environmental influences on reading and listening comprehension. *J. Res. Read.* **29**, 75–91 (2006).

243.	Mataix-Cols, D. *et al.* Familial Risks of Tourette Syndrome and Chronic Tic Disorders: A Population-Based Cohort Study. *JAMA Psychiatry* **72**, 787 (2015).

244.	Fliers, E. *et al.* ADHD and poor motor performance from a family genetic perspective. *J. Am. Acad. Child Adolesc. Psychiatry* **48**, 25–34 (2009).

245.	Taylor, M. J. *et al.* Developmental associations between traits of autism spectrum disorder and attention-deficit/hyperactivity disorder: A genetically-informative, longitudinal twin study. *Psychol. Med.* **43**, 1735–1746 (2013).

246.	Light, J. G., Pennington, B. F., Gilger, J. W. & DeFries, J. C. Reading disability and hyperactivity disorder: Evidence for a common genetic etiology. *Dev. Neuropsychol.* **11**, 323–335 (1995).

247.	Ooki, S. Genetic and environmental influences on stuttering and tics in Japanese twin children. *Twin Res. Hum. Genet. Off. J. Int. Soc. Twin Stud.* **8**, 69–75 (2005).

248.	Hur, Y.-M. Genetic and environmental etiology of the relationship between childhood hyperactivity/inattention and conduct problems in a South Korean twin sample. *Twin Res. Hum. Genet.* **18**, 290–297 (2015).

249.	O’Nions, E. *et al.* Examining the Genetic and Environmental Associations between Autistic Social and Communication Deficits and Psychopathic Callous-Unemotional Traits. *PloS One* **10**, 12 (2015).

250.	Spinath, F. M., Price, T. S., Dale, P. S. & Plomin, R. The genetic and environmental origins of language disability and ability. *Child Dev.* **75**, 445–454 (2004).

251.	Taylor, M. J. *et al.* Language and traits of autism spectrum conditions: Evidence of limited phenotypic and etiological overlap. *Am. J. Med. Genet. B Neuropsychiatr. Genet.* **165**, 587–595 (2014).

252.	Constantino, J. N. & Todd, R. D. Autistic traits in the general population: a twin study. *Arch. Gen. Psychiatry* **60**, 524–530 (2003).

253.	Hallett, V., Ronald, A., Rijsdijk, F. & Happé, F. Disentangling the associations between autistic-like and internalizing traits: a community based twin study. *J. Abnorm. Child Psychol.* **40**, 815–827 (2012).

254.	Hoekstra, R., Happe, F., Baron-Cohen, S. & Ronald, A. Limited genetic covariance between autistic traits and intelligence: Findings from a longitudinal twin study+. *Am J Med Genet Part B Neuropsychiatr Genet* **153B**, 994–1007 (2010).

255.	Holmboe, K. *et al.* Strong genetic influences on the stability of autistic traits in childhood. *J. Am. Acad. Child Adolesc. Psychiatry* **53**, 221–230 (2014).

256.	Taylor, M. J., Gillberg, C., Lichtenstein, P. & Lundstrom, S. Etiological influences on the stability of autistic traits from childhood to early adulthood: evidence from a twin study. *Mol. Autism* **8**, 5 (2017).

257.	Mazefsky, C. A., Goin-Kochel, R. P., Riley, B. P. & Maes, H. H. Genetic and environmental influences on symptom domains in twins and siblings with autism. *Res. Autism Spectr. Disord.* **2**, 320–331 (2008).

258.	Cole, J., Ball, H., Martin, N., Scourfield, J. & McGuffin, P. Genetic Overlap Between Measures of Hyperactivity/Inattention and Mood in Children and Adolescents. *J. Am. Acad. Child Adolesc. Psychiatry* **48**, 1094–1101 (2009).

259.	Constantino, J., Hudziak, J. & Todd, R. Deficits in Reciprocal Social Behavior in Male Twins: Evidence for a Genetically Independent Domain of Psychopathology. *J. Am. Acad. Child Adolesc. Psychiatry* **42**, 458–467 (2003).

260.	Eaves, L. J. *et al.* Genetics and developmental psychopathology .2. The main effects of genes and environment on behavioral problems in the Virginia twin study of adolescent behavioral development. *J. Child Psychol. Psychiatry* **38**, 965–980 (1997).

261.	Eaves, L. *et al.* Genetic and Environmental Causes of Covariation in Interview Assessments of Disruptive Behavior in Child and Adolescent Twins. *Behav Genet* **30**, 321–334 (2000).

262.	Gregory, A. M., Eley, T. C., O’Connor, T. G. & Plomin, R. Etiologies of associations between childhood sleep and behavioral problems in a large twin sample. *J. Am. Acad. Child Adolesc. Psychiatry* **43**, 744–751 (2004).

263.	Hudziak, J., Rudiger, L., Neale, M., Heath, A. & Todd, R. A Twin Study of Inattentive, Aggressive, and Anxious/Depressed Behaviors. *J. Am. Acad. Child Adolesc. Psychiatry* **39**, 469–476 (2000).

264.	Kuo, P.-H., Lin, C. C., Yang, H.-J., Soong, W.-T. & Chen, W. J. A twin study of competence and behavioral/emotional problems among adolescents in Taiwan. *Behav. Genet.* **34**, 63–74 (2004).

265.	Larsson, H., Lichtenstein, P. & Larsson, J.-O. Genetic contributions to the development of ADHD subtypes from childhood to adolescence. *J. Am. Acad. Child Adolesc. Psychiatry* **45**, 973–981 (2006).

266.	Van Beijsterveldt, C. E. M., Verhulst, F. C., Molenaar, P. C. M. & Boomsma, D. I. The genetic basis of problem behavior in 5-year-old Dutch twin pairs. *Behav. Genet.* **34**, 229–242 (2004).

267.	Vierikko, E., Pulkkinen, L., Kaprio, J. & Rose, R. J. Genetic and environmental influences on the relationship between aggression and hyperactivity-impulsivity as rated by teachers and parents. *Twin Res. Hum. Genet.* **7**, 261–274 (2004).

268.	de Zeeuw, E. L., van Beijsterveldt, C. E. M., Ehli, E. A., de Geus, E. J. C. & Boomsma, D. I. Attention Deficit Hyperactivity Disorder Symptoms and Low Educational Achievement: Evidence Supporting A Causal Hypothesis. *Behav. Genet.* **47**, 278–289 (2017).

269.	Do, E. K. *et al.* The role of genetic and environmental influences on the association between childhood ADHD symptoms and BMI. *Int. J. Obes.* **43**, 33–42 (2019).

270.	Larsson, J.-O., Larsson, H. & Lichtenstein, P. Genetic and environmental contributions to stability and change of ADHD symptoms between 8 and 13 years of age: a longitudinal twin study. *J. Am. Acad. Child Adolesc. Psychiatry* **43**, 1267–1275 (2004).

271.	Nadder, T. S., Rutter, M., Silberg, J. L., Maes, H. H. & Eaves, L. J. Genetic effects on the variation and covariation of attention deficit-hyperactivity disorder (ADHD) and oppositional-defiant disorder/conduct disorder (ODD/CD) symptomatologies across informant and occasion of measurement. *Psychol. Med.* **32**, 39–53 (2002).

272.	Rietveld, M. J., Hudziak, J. J., Bartels, M., Van Beijsterveldt, C. E. M. & Boomsma, D. I. Heritability of attention problems in children: longitudinal results from a study of twins, age 3 to 12. *J. Child Psychol. Psychiatry* **45**, 577–588 (2004).

273.	Saudino, K., Ronald, A. & Plomin, R. The Etiology of Behavior Problems in 7-Year-Old Twins: Substantial Genetic Influence and Negligible Shared Environmental Influence for Parent Ratings and Ratings by Same and Different Teachers. *J Abnorm Child Psychol* **33**, 113–130 (2005).

274.	Silberg, J. *et al.* Genetic and environmental influences on the covariation between hyperactivity and conduct disturbance in juvenile twins. *J. Child Psychol. Psychiatry* **37**, 803–816 (1996).

275.	Sherman, D. K., Iacono, W. G. & McGue, M. K. Attention-Deficit Hyperactivity Disorder Dimensions: A Twin Study of Inattention and Impulsivity-Hyperactivity. *J. Am. Acad. Child Adolesc. Psychiatry* **36**, 745–753 (1997).

276.	Alarcón, M., DeFries, J. C. & Fulker, D. W. Etiology of individual differences in reading performance: A test of sex limitation. *Behav. Genet.* **25**, 17–23 (1995).

277.	Bates, T. C. *et al.* Behaviour genetic analyses of reading and spelling: A component processes approach. *Aust. J. Psychol.* **56**, 115–126 (2004).

278.	Harlaar, N., Spinath, F. M., Dale, P. S. & Plomin, R. Genetic influences on early word recognition abilities and disabilities: A study of 7‐year‐old twins. *J. Child Psychol. Psychiatry* **46**, 373–384 (2005).

279.	Reynolds, C. A. *et al.* The genetics of children’s oral reading performance. *J. Child Psychol. Psychiatry* **37**, 425–434 (1996).

280.	Knopik, V., Heath, A., Bucholz, K., Madden, P. & Waldron, M. Genetic and environmental influences on externalizing behavior and alcohol problems in adolescence: A female twin study. *Pharmacol Biochem Behav* **93**, 313–321 (2009).

281.	Neuman, R. J. *et al.* Latent class analysis of ADHD and comorbid symptoms in a population sample of adolescent female twins. *J. Child Psychol. Psychiatry* **42**, 933–942 (2001).

282.	Verhoef, E., Shapland, C. Y., Fisher, S. E., Dale, P. S. & St Pourcain, B. The developmental origins of genetic factors influencing language and literacy: Associations with early‐childhood vocabulary. *J. Child Psychol. Psychiatry* (2020).

283.	Gandal, M. J. *et al.* Shared molecular neuropathology across major psychiatric disorders parallels polygenic overlap. *Science* **359**, 693–697 (2018).

284.	Grove, J. *et al.* Identification of common genetic risk variants for autism spectrum disorder. *Nat. Genet.* **51**, 431–444 (2019).

285.	Hill, W. D. *et al.* Age-Dependent Pleiotropy Between General Cognitive Function and Major Psychiatric Disorders. *Biol. Psychiatry* **80**, 266–273 (2016).

286.	Lee, S. H. *et al.* Genetic relationship between five psychiatric disorders estimated from genome-wide SNPs. *Nat. Genet.* **45**, 984 (2013).

287.	Serdarevic, F. *et al.* Polygenic risk scores for developmental disorders, neuromotor functioning during infancy, and autistic traits in childhood. *Biol. Psychiatry* **87**, 132–138 (2020).

288.	Solberg, B. S. *et al.* Patterns of psychiatric comorbidity and genetic correlations provide new insights into differences between attention-deficit/hyperactivity disorder and autism spectrum disorder. *Biol. Psychiatry* **86**, 587–598 (2019).

289.	St Pourcain, B. *et al.* Variability in the common genetic architecture of social-communication spectrum phenotypes during childhood and adolescence. *Mol. Autism* **5**, 1–12 (2014).

290.	St Pourcain, B. *et al.* Developmental changes within the genetic architecture of social communication behavior: a multivariate study of genetic variance in unrelated individuals. *Biol. Psychiatry* **83**, 598–606 (2018).

291.	St Pourcain, B. *et al.* ASD and schizophrenia show distinct developmental profiles in common genetic overlap with population-based social communication difficulties. *Mol. Psychiatry* **23**, 263–270 (2018).

292.	Stergiakouli, E. *et al.* Shared genetic influences between dimensional ASD and ADHD symptoms during child and adolescent development. *Mol. Autism* **8**, 1–13 (2017).

293.	Warrier, V. & Baron-Cohen, S. Genetic contribution to ‘theory of mind’in adolescence. *Sci. Rep.* **8**, 1–9 (2018).

294.	Autism Spectrum Disorders Working Group of The Psychiatric Genomics, C. Meta-analysis of GWAS of over 16,000 individuals with autism spectrum disorder highlights a novel locus at 10q24.32 and a significant overlap with schizophrenia. *Mol. Autism* **8**, 21 (2017).

295.	Pettersson, E. *et al.* Genetic influences on eight psychiatric disorders based on family data of 4 408 646 full and half-siblings, and genetic data of 333 748 cases and controls. *Psychol. Med.* **49**, 1166–1173 (2019).

296.	Artigas, M. S. *et al.* Attention-deficit/hyperactivity disorder and lifetime cannabis use: genetic overlap and causality. *Mol. Psychiatry* **25**, 2493–2503 (2020).

297.	Demontis, D. *et al.* Discovery of the first genome-wide significant risk loci for attention deficit/hyperactivity disorder. *Nat. Genet.* **51**, 63–75 (2019).

298.	Martin, J. *et al.* A genetic investigation of sex bias in the prevalence of attention-deficit/hyperactivity disorder. *Biol. Psychiatry* **83**, 1044–1053 (2018).

299.	Micalizzi, L. *et al.* Single nucleotide polymorphism heritability and differential patterns of genetic overlap between inattention and four neurocognitive factors in youth. *Dev. Psychopathol.* **33**, 76–86 (2021).

300.	Middeldorp, C. M. *et al.* A genome-wide association meta-analysis of attention-deficit/hyperactivity disorder symptoms in population-based pediatric cohorts. *J. Am. Acad. Child Adolesc. Psychiatry* **55**, 896-905. e6 (2016).

301.	Pappa, I. *et al.* Single nucleotide polymorphism heritability of behavior problems in childhood: genome-wide complex trait analysis. *J. Am. Acad. Child Adolesc. Psychiatry* **54**, 737–744 (2015).

302.	Rovira, P. *et al.* Shared genetic background between children and adults with attention deficit/hyperactivity disorder. *Neuropsychopharmacology* 1–10 (2020).

303.	Gialluisi, A. *et al.* Genome-wide association study reveals new insights into the heritability and genetic correlates of developmental dyslexia. *Mol. Psychiatry* 1–14 (2020).

304.	Martin, J. *et al.* Examining sex-differentiated genetic effects across neuropsychiatric and behavioral traits. *Biol. Psychiatry* (2021).
